# Meaning of clinical calculator results: a cross-sectional analysis of the MDCalc catalogue

**DOI:** 10.64898/2026.09.08.26362556

**Authors:** Shuhan He, Joshua W. Joseph, Pedram Safari, Natasha Akomeh, Allison Goff, Pawel Jan Slusarz, Amal Mohamed, Spencer Lord, Joshua N. Goldstein, Ali S. Raja, Christopher Kabrhel, Brian W. Locke, Cory Rohlfsen, David M. Liebovitz

**Affiliations:** Division of AI, Department of Emergency Medicine, Massachusetts General Hospital, Mass General Brigham, Boston, Massachusetts, USA; Master of Science in Healthcare Data Analytics, Department of Health Sciences, School of Health and Rehabilitation Sciences, Mass General Brigham University of Health Professions, Boston, Massachusetts, USA; Department of Surgery, Massachusetts General Hospital, Mass General Brigham, Boston, Massachusetts, USA; Department of Emergency Medicine, Massachusetts General Hospital, Mass General Brigham, Boston, Massachusetts, USA; Division of Respiratory, Critical Care, and Occupational Pulmonary Medicine, Department of Internal Medicine, Spencer Fox Eccles School of Medicine at the University of Utah, Salt Lake City, Utah, USA; Department of Pulmonary and Critical Care, Intermountain Medical Center, Intermountain Health, Murray, Utah, USA; Division of General Internal Medicine, Department of Internal Medicine, College of Medicine, University of Nebraska Medical Center, Omaha, Nebraska, USA; Departments of Medicine and Preventive Medicine, Feinberg School of Medicine, Northwestern University, Chicago, Illinois, USA; Department of Emergency Medicine, Harvard Medical School, Boston, Massachusetts, USA

## Abstract

**Objectives:** To quantify complementary meanings of binary clinical calculator results: information beyond the observed outcome frequency, post-result risk, information direction, and precision.

**Design:** Catalogue-based cross-sectional analysis of published aggregate performance data, with a reproducible row-level audit.

**Setting:** The 847-calculator MDCalc catalogue and a fixed collection of publicly available reports assembled without a systematic literature search.

**Participants:** Evaluations comparing a binary calculator result with a binary clinical outcome using a complete or reconstructable 2 x 2 table.

**Main outcome measures:** Percentage reduction in uncertainty and corresponding information gain in bits; risk after positive and negative classifications; the proportion of information from each classification; and whether the upper 95% confidence bound for post-negative risk supported specified thresholds.

**Results:** The analysis included 482 evaluations of 407 calculators. The median study size was 422 and the median observed outcome frequency was 17.6%. Results reduced uncertainty by a median 14.9% (interquartile range 6.2%-30.5%; 95% confidence interval 12.1% to 17.2%), corresponding to 0.093 bits. In 329 evaluations (68.3%), one classification supplied more than 60% of average information. PERC reduced uncertainty by 4.3% on average while its negative classification lowered observed risk from 7.6% to 1.0%. Two HEART thresholds in the same cohort shifted the positive-classification information share from 39.1% to 79.8%. Post-negative risk was below 2% in 157 evaluations by point estimate, but the upper 95% confidence bound was below 2% in only 67; 90 of 157 (57%) did not support the apparent threshold.

**Conclusions:** Clinical calculator results have no single quantitative meaning. Conventional performance, information added beyond the observed outcome frequency, post-result risk, classification-specific information, and precision provide complementary interpretations. Whether acting on a result improves care remains a separate question.

**Key messages:** *What is already known on this topic:*

- Clinical calculators are conventionally described using sensitivity, specificity, predictive values, likelihood ratios, discrimination, and calibration, but these measures answer different questions about a result.

*What this study adds:*

- Across 482 evaluations, calculator results removed a median 14.9% of starting uncertainty; one classification supplied more than 60% of average information in 329 evaluations.
- Of 157 evaluations with a post-negative point estimate below 2%, only 67 had an upper 95% confidence bound below 2%.

*How this study might affect research, practice or policy:*

- Reports of clinical calculators could pair conventional accuracy with starting risk, post-result risks and intervals, and, when average learning matters, uncertainty reduction and its direction; none alone establishes clinical benefit.

## Introduction

Clinical calculators combine patient information to support decisions about diagnosis, prognosis, testing, treatment, or discharge.[1–3] Their performance is usually described using sensitivity, specificity, predictive values, discrimination, and calibration. These measures are familiar, but they answer different questions about what a result means.

One question is how much was learned beyond the outcome risk already present before classification. Published information-theory studies have quantified this change for diagnostic tools and individual clinical features.[4–10] A second question is where observed risk ended after a particular positive or negative result. A third is how strongly the evaluation supports that post-result risk, because a reassuring point estimate from a small group can remain imprecise. Whether the result should change care is further downstream and depends on an accepted action threshold and the consequences of acting.

We applied these complementary views to binary evaluations linked to the MDCalc catalogue. We asked how much information results added beyond the observed outcome frequency, where risk ended after each classification, which classification supplied the information, and whether the study estimated post-result risk precisely enough to support specified thresholds.

## Methods

### Study design and data source

We performed a cross-sectional secondary analysis of the MDCalc catalogue and published aggregate performance data. A pre-existing collection assembled from publicly available reports was fixed with the 847-calculator catalogue on 11 August 2026.[11] Each analysis represented one calculator evaluated in one study for a specified outcome and threshold. We followed applicable Strengthening the Reporting of Observational Studies in Epidemiology items.[12]

### Eligibility and selection

An evaluation was eligible if it provided, or allowed reconstruction of, a 2 x 2 table comparing a binary calculator result with a binary clinical outcome. Counts could be reconstructed from event totals, sample size, sensitivity, specificity, and prevalence. We retained separate thresholds, patient groups, and outcomes because each represents a different calculator use.

We excluded continuous values, stages, doses, or risk estimates without a binary threshold. The workbook contained 494 eligible study-level evaluations: 487 linked to a catalogue calculator and seven described four other tools or strategies. Removing one superseded reconstruction and four exact duplicate tables left 482 primary evaluations representing 407 unique catalogue calculators. Calculators could contribute multiple evaluations. Absence from the analytic set meant only that no primary binary evaluation was in the assembled workbook.

We matched names to catalogue identifiers using standardised names and documented aliases. For repeated identical counts, we retained the record with the clearest source information. The seven off- catalogue evaluations entered sensitivity analysis only. Full matching and duplicate rules are in the supplement.

### Data extraction and verification

The collection drew on MDCalc evidence pages, studies cited by MDCalc, PubMed abstracts, and available primary or validation reports. Complete tables were available or reconstructed for all 482 evaluations: 331 had directly reported counts, 64 were reconstructed from sensitivity and specificity, 19 from other metrics, and 68 were legacy records without a recorded data source. A large language model extracted analytic data for 157 evaluations (32.6%), returning either the four cells or the metrics needed to reconstruct them. The same prompt selected the study and threshold for those rows and, if no threshold was recommended, selected the highest Youden’s index. Because Youden’s index correlates closely with our measures, we report information yield by extraction route rather than treating route differences as corrected estimates. Searches were neither prespecified nor recorded as a systematic review, so conclusions are limited to this fixed collection.

For each evaluation, we recorded source, threshold, sample size, clinical use, specialty, disease area, confidence label, and counts. Automated checks identified missing or impossible counts, sample-size discrepancies, and calculation errors. We recalculated every measure from the four cells. These checks did not independently verify every row against its source. Flagged discrepancies and selected examples were checked against available reports; one corrected source count and its one-participant denominator discrepancy are documented in the supplement.[13]

The source workbook did not systematically record participant sex or gender, so results could not be disaggregated by sex or gender.

### Information measures

For a binary outcome, uncertainty is greatest when either outcome is equally likely and smaller when one is already likely. We quantified what the calculator result added beyond each table’s observed outcome frequency as the reduction in binary Shannon entropy. Information gained was starting entropy minus the weighted entropy remaining after positive and negative results. We report this mainly as the percentage of starting uncertainty removed; the value in bits is the formal information-theory measure. Each estimate describes one table at its observed frequency and does not indicate whether a result changes a decision.

We also decomposed average information into positive- and negative-result contributions. Equations, conventions, and derivations are in the supplement.

### Conventional performance measures

We recalculated sensitivity, specificity, predictive values, likelihood ratios, diagnostic odds ratio, and Youden’s J from the same tables.[14] Mathematical comparisons with information gain are in the supplement.

Because plug-in mutual information estimates can be upwardly biased in small samples, we applied the Miller-Madow correction in sensitivity analysis.[15] Corrected values were constrained to be non-negative; observed-table estimates remained primary.

### Clinical calculator examples

We prespecified seven evaluations for explanation: three familiar rule-out decisions, two pneumonia scores in one cohort, and two HEART thresholds in one cohort. They illustrated contrasts in the frozen data, not calculator rankings or best-evidence claims. For PERC, we compared risk after low clinical suspicion and a negative rule with the validation target and development-study testing threshold.[16,17]

Constructed comparisons held sensitivity and specificity constant while prevalence changed, or held prevalence and Youden’s J constant while sensitivity and specificity changed. Exact values and cautions are in tables S6-S8.

### Whether a negative result is supported by its study

For each evaluation, we calculated risk after a negative classification (one minus negative predictive value) and after a positive classification (positive predictive value), with Wilson 95% intervals. No observed missed cases does not establish a low miss rate; the negative-group denominator determines how low a rate the study can support.

At thresholds of 0.5%, 1%, 2%, 5%, 10%, and 20%, we asked whether the **upper** bound of post-negative risk fell below the threshold. The analogous lower bound of post-positive risk gave a rule-in statement. These describe what an evaluation can support, not recommended thresholds or benefits from acting.

### Statistical analysis

Continuous measures were summarised with medians and interquartile ranges. Spearman rank correlations were primary to describe monotonic associations with less sensitivity to extreme values; Pearson correlations were secondary. We examined associations with sensitivity, specificity, Youden’s J, outcome prevalence, starting entropy, and reported sample size. Confidence intervals for medians, proportions, and correlations were estimated by resampling calculators 2000 times while keeping all evaluations of the same calculator together. This accounted for the 34 calculators that contributed more than one evaluation. We emphasised estimates and 95% confidence intervals rather than P values.

Sensitivity analyses are listed in the supplement and covered off-catalogue inclusions, one evaluation per calculator, and restriction by count type, evidence confidence, and reconstruction status. We labelled sampling design from the recorded source description as cohort or consecutive, case-control or two-gate, or unclear. To avoid inferring design from incomplete citations, only explicit wording determined a label; we repeated the headline analysis after excluding records explicitly described as case-control or two-gate. Fixed-prevalence standardisations and constructed contrasts are reported in the supplement.

Clinical-purpose analyses included diagnosis, prognosis, rule-out, and treatment; labels could overlap. Analyses used Python 3.12 with pandas, NumPy, SciPy, and Matplotlib. Additional statistical details are provided in the supplement; the frozen script, data dictionary, row-level audit, and machine-readable outputs are provided in the reproducibility archive.

### Use of artificial intelligence

Anthropic Claude Sonnet 4 (claude-sonnet-4-20250514) extracted or reconstructed the 2 x 2 cells and supported study, threshold, and confidence-label selection for 157 evaluations, as described above. OpenAI Codex supported code editing, deterministic figure production, literature organisation, and language editing. Models did not perform scientific calculations. Deterministic code regenerated every number, table, and data-derived figure from frozen inputs, using seed 20260811; assertions stopped the build if core inputs or results changed. Human authors reviewed sources, decisions, citations, figures, interpretation, and wording and remain responsible for the work.

### Patient and public involvement

Patients and members of the public were not involved in setting the research question, designing or conducting the study, interpreting the results, or planning dissemination because this was a secondary analysis of publicly available aggregate calculator evidence.

### Ethics

The evidence collection was assembled and frozen on 11 August 2026, and initial analysis of the publicly available aggregate data preceded the written determination. On 14 August 2026, Mass General Brigham Human Research Affairs determined that the work covered by REDCap 4737 did not constitute human subjects research under 45 CFR 46 and did not require institutional review board approval. Subsequent validation, analysis, and manuscript preparation proceeded on that basis. No human participants or identifiable private information were involved.

## Results

### Analytic coverage

The primary analysis contained 482 evaluations representing 407 unique calculators, or 48.1% of the 847- calculator catalogue (fig S1). Among included calculators, 373 contributed one evaluation and 34 contributed more than one; one calculator contributed 15. Coverage was highest for the clearest binary clinical use: 43 of 44 catalogue calculators labelled for rule-out use (97.7%) were represented. Including the seven off-catalogue evaluations produced 489 evaluations representing 411 calculators or strategies.

Automated checks found complete analytic 2×2 tables and no impossible negative counts. All information calculations passed prespecified consistency checks. Direct count-based source evidence was available for 331 evaluations (68.7%), and 187 (38.8%) carried a high or moderate-high confidence label. A study citation was recorded for 417 (86.5%) and a threshold description for 411 (85.3%). These checks verified the calculations but did not independently verify every source or make the evidence collection a systematic review.

### Characteristics of included evaluations

The median study included 422 participants (interquartile range 169-1190; range 15-4,830,941), and the median outcome frequency was 17.6% (7.0-40.6%). A calculator could have several purposes: 283 evaluations included prognosis, 200 diagnosis, 56 rule-out use, and 36 treatment. Median sensitivity was 0.824, median specificity 0.781, and median Youden’s J 0.448 (table 1).

**Table 1.** Characteristics of 482 calculator-study-outcome-threshold evaluations.

| Measure | Median (interquartile range) | Range |
| --- | --- | --- |
| Reported sample size | 422 (169-1190) | 15-4,830,941 |
| Sensitivity* | 0.824 (0.652-0.931) | 0-1.000 |
| Specificity | 0.781 (0.561-0.905) | 0.024-1.000 |
| Observed outcome frequency | 0.176 (0.070-0.406) | 0.006-0.962 |
| Youden's J | 0.448 (0.251-0.638) | -0.015 to 1.000 |
| Reduction in uncertainty, %; corresponding information gain, bits | 14.9 (6.2-30.5); 0.093 (0.026-0.210) | <0.1-100.0; <0.001-0.763 |
| Starting uncertainty, bits | 0.649 (0.349-0.918) | 0.055-1.000 |

\*A sensitivity of 0 occurred in two evaluations with outcome events and no true-positive classifications.

### How much information did calculator results add?

The median result reduced uncertainty by 14.9% (interquartile range 6.2-30.5%; 95% confidence interval 12.1% to 17.2%). About one in five evaluations reduced uncertainty by less than 5%, whereas one in 10 reduced it by at least half. In detail, 299 of 482 evaluations (62.0%; 55.8% to 67.8%) reduced uncertainty by at least 10%, 150 (31.1%; 26.1% to 36.3%) by at least 25%, and 48 (10.0%; 7.2% to 13.1%) by at least half. The corresponding median information gain was 0.093 bits (0.026-0.210; 0.065 to 0.108) (fig 2B).

Percentage uncertainty reduction was closely correlated with Youden’s J (Spearman’s rho 0.971; 95% confidence interval 0.964 to 0.976). It should therefore not be interpreted as an independent universal ranking measure. Its distinct interpretation is the proportion of uncertainty removed beyond the observed outcome frequency, and the information framework also permits decomposition by classification and comparison across observed frequencies (table S3 and fig S2).

### Where did risk end after individual results?

Sensitivity describes the proportion of people with the outcome who have a positive result, and specificity the proportion without it who have a negative result. Reduction in uncertainty asks how much was learned on average across both results at their observed frequencies. Post-result risk asks where observed risk ended after one result. Figure 1 shows these quantities together for seven prespecified examples drawn from the PERC,[17] Canadian CT Head Rule,[18] Glasgow-Blatchford,[19] CRB-65/CURSI,[13,20] and HEART[21] source studies; exact values and source cautions are in table S8.

**Fig 1.**
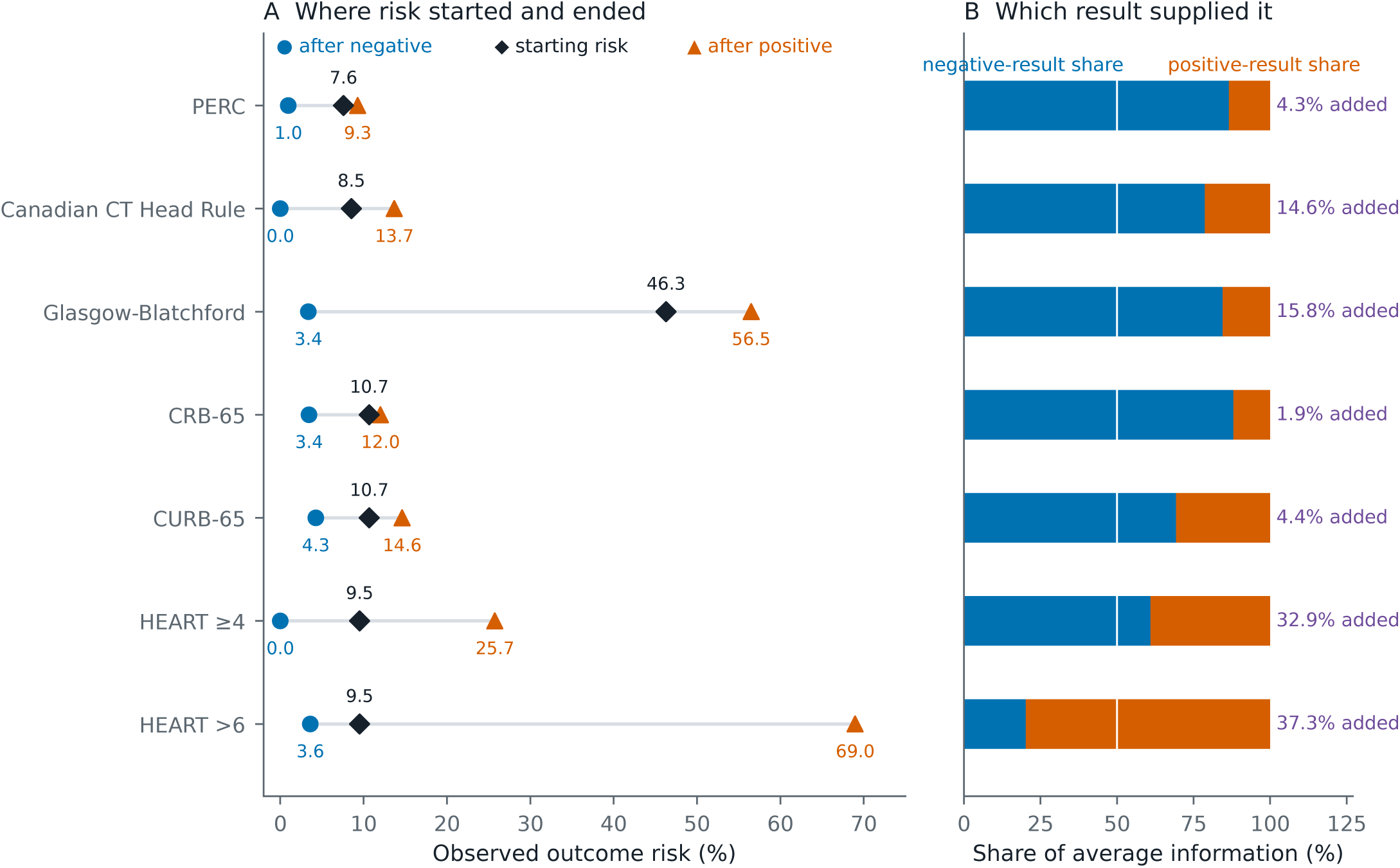
Different quantities describe different meanings of the same calculator result. Panel A shows the starting outcome risk and the risks observed after negative and positive classifications for seven prespecified examples. Panel B shows how much of the average information came from each classification; the text at right gives the percentage of starting uncertainty removed across both classifications. PERC illustrates modest average information with a low post-negative risk. The two HEART rows use different thresholds in the same 641 patients and reverse which classification supplies most information. Values describe these calculator-study-outcome-threshold evaluations, not calculator performance in every setting. Exact values, outcomes, and source cautions are in table S8.

PERC makes the distinction concrete. Among 8138 patients with suspected pulmonary embolism, low clinical suspicion combined with the PERC classification was 97.4% sensitive and 21.9% specific for venous thromboembolism or death within 45 days. The validation study prespecified a 2.0% target for that composite endpoint. A negative classification occurred in 1666 patients (20.5%) and lowered observed risk from 7.6% to 1.0%, below the target.[17] A positive classification occurred in 6472 patients and raised risk to 9.3%. Across both results, PERC reduced uncertainty by 4.3%; the negative classification supplied 86.6% of that information. High sensitivity, modest average information, and an observed post-negative risk below a prespecified target can therefore coexist. The separate 1.8% value was a pulmonary embolism testing threshold estimated during development.[16]

### Which classification supplied the information?

The median positive-classification share was 51.5% (interquartile range 34.7-67.2%). In 329 of 482 evaluations (68.3%), one classification supplied more than 60% of average information: 156 were negative dominant and 173 positive dominant. This descriptive proportion depends on the 40/60 boundary and is not itself a distinctive property of clinical calculators. Among 56 rule-out evaluations, negative classifications supplied a median 71.3% (95% confidence interval 65.6% to 79.0%) of average information.

Threshold changed this pattern within the same calculator and cohort. At HEART ≥4, negative classifications supplied 60.9% of the information; at HEART >6, positive classifications supplied 79.8%. In the same pneumonia cohort, CURB-65 traded some sensitivity for specificity and reduced uncertainty by 4.41% rather than 1.90% for CRB-65. These are properties of the evaluated threshold, population, and outcome rather than context-free properties of the calculator.

### How precisely were post-result risks supported?

Risk after a negative classification fell below 2% in 157 of 482 evaluations on the point estimate. The Wilson 95% upper bound also fell below 2% in 67. In the remaining 90 evaluations, 57% of those that appeared to reach the threshold, the data did not support a 2% claim (fig 3).

**Fig 2.**
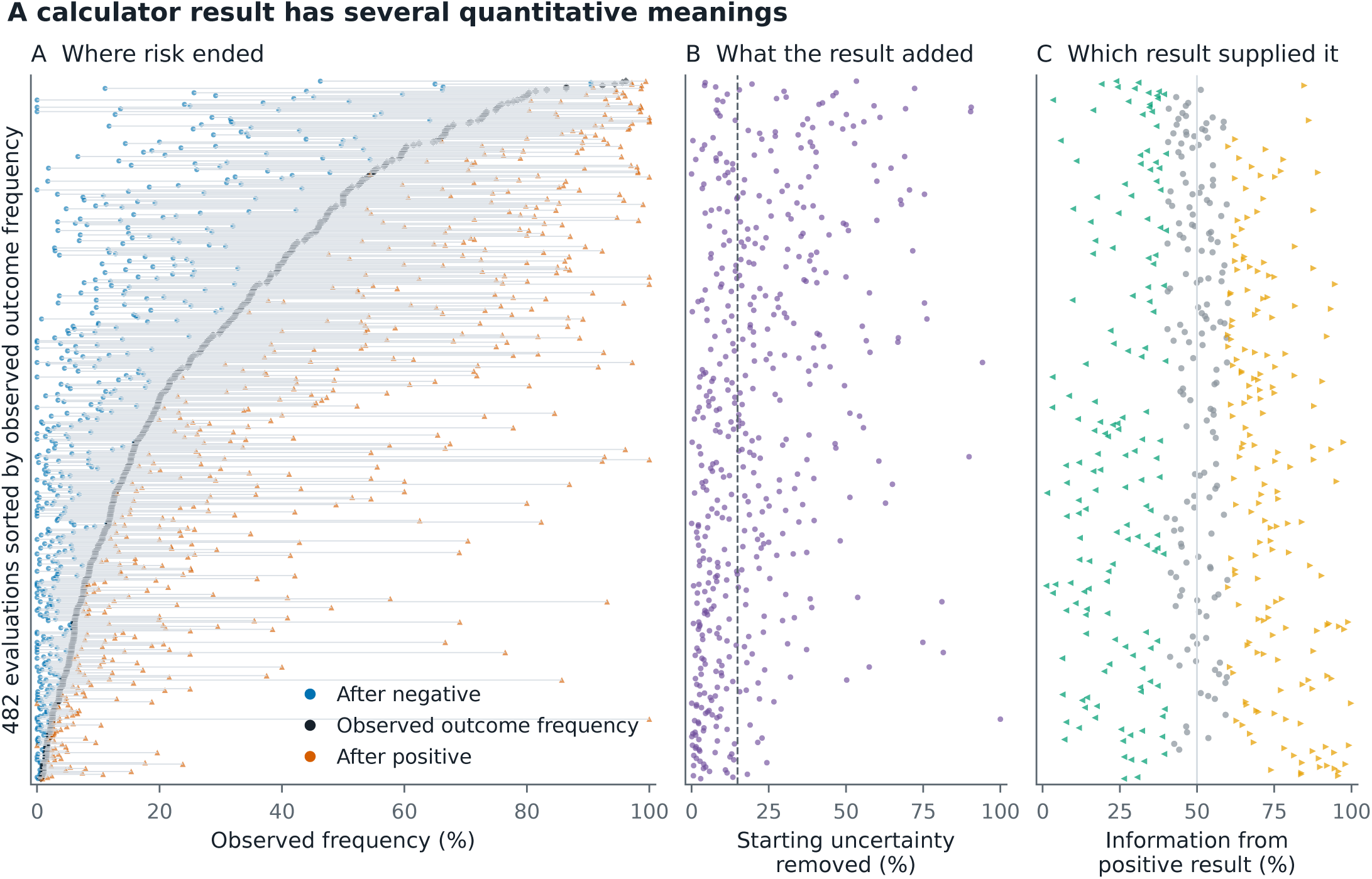
A calculator result has several quantitative meanings across the catalogue. All 482 calculator-study- outcome-threshold evaluations are aligned and sorted only by observed outcome frequency. Panel A shows the frequencies observed after negative and positive classifications around the starting frequency. Panel B shows the percentage of starting uncertainty removed; the dashed line marks the median of 14.9%. Panel C shows the positive-classification share of average information, with left-pointing, circular, and right-pointing markers denoting negative-dominant, balanced, and positive-dominant evaluations under the descriptive 40/60 boundaries. The aligned panels show why post-result frequency, average information, and information direction cannot be collapsed into one quality score.

**Fig 3.**
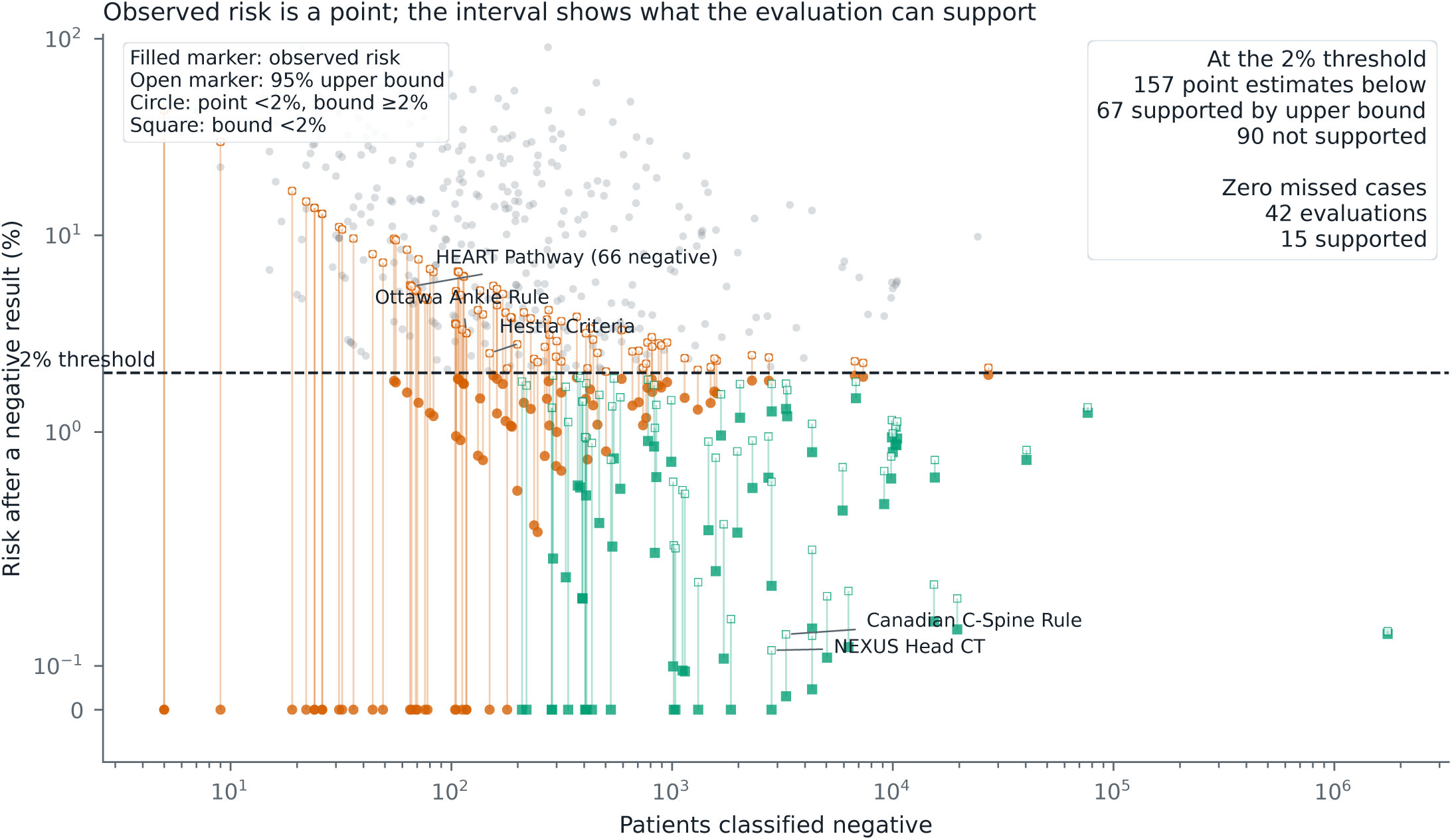
Observed post-result risk and the precision with which it was estimated. Each filled marker is the observed risk after a negative classification and each open marker its Wilson 95% upper bound; the vertical segment joins the two. Circles identify evaluations with a point estimate below 2% whose upper bound remained at or above 2%, and squares identify evaluations whose upper bound was below 2%. Of 157 point estimates below 2%, 67 were supported by the upper bound and 90 were not. Among 42 evaluations with no missed cases, 15 supported the threshold. Labels illustrate the effect of the negative-group denominator and are not a ranking. Each observation is one calculator-study-outcome-threshold evaluation.

The difference was clearest where no missed cases were observed. Forty-two evaluations recorded no false negatives; in 15, the 95% upper bound was also below 2%. Among the remaining 27, the Ottawa Ankle Rule evaluation recorded no missed fractures among 117 negative results, consistent with a true risk up to 3.18%; the Hestia criteria recorded none among 149, consistent with up to 2.51%; and the HEART Pathway recorded none among 66, consistent with up to 5.50%. Complete evaluation-level support estimates are in the reproducibility archive.

These statements apply to the included evaluation, not to the calculator as a whole. A calculator represented by one study here may have a larger validation literature elsewhere, and the thresholds span plausible values rather than recommendations for specific decisions.

### Why observed outcome frequency matters

Information gain depended strongly on the observed outcome frequency. When an outcome was already very rare in the analytic table, there was less uncertainty for the calculator to reduce. Information gain was associated with observed outcome frequency (Spearman’s rho 0.673; 95% confidence interval 0.593 to 0.738) and the uncertainty present before the result (rho 0.686; 0.611 to 0.748) (table S3). Every evaluation lay at or below the amount of uncertainty present before the result.

Information gain was also strongly associated with Youden’s J (rho 0.855; 95% confidence interval 0.820 to 0.880). Even when sensitivity and specificity were unchanged, the amount of new information still depended on the outcome frequency used in the calculation.

### Robustness of the findings

The main findings changed little across sensitivity analyses. Median reduction in uncertainty differed by evidence route rather than being uniform: 14.7% among the 331 evaluations with direct count-based source evidence, 20.3% among 64 reconstructed from reported sensitivity and specificity, 22.1% among 19 reconstructed from other reported metrics, and 7.7% among 68 legacy records without a recorded data source, against 14.9% overall. It was 16.6% among the 157 model-extracted evaluations and 13.0% among the remaining 325. Because these strata differ in opposite directions, the overall median should be read as their composite rather than as evidence of stability. Sensitivity analyses including the seven off- catalogue evaluations, restricting to one evaluation per calculator, and restricting by count type, evidence confidence, or reconstruction status kept the median between 13.0% and 15.7% (table S4). Source descriptions explicitly identified 21 cohort or consecutive evaluations and two case-control or two-gate evaluations; design remained unclear for 459. Excluding the two explicitly outcome-selected evaluations left 480 evaluations with a median reduction of 14.8% and median information gain of 0.092 bits. A small-sample correction changed the median only slightly, from 14.9% (0.093 bits) to 14.4% (0.092 bits).

## Discussion

Calculator results had several related but distinct quantitative interpretations, and no single measure captured all of them. Across the 482 evaluations, uncertainty reduction varied from less than 0.1% to 100.0%; one classification supplied more than 60% of average information in 329; clinically consequential post-result risk could coexist with modest average information; and accounting for precision reduced the 157 apparent 2% rule-outs to 67 supported evaluations.

These measures answer different questions rather than competing for one score. Sensitivity and specificity describe classification conditional on outcome status. Likelihood ratios describe how one result changes odds. Post-result risk shows where observed risk ended for patients with that result. Reduction in Shannon entropy describes average learning across both results beyond the observed outcome frequency, and its decomposition shows which result supplied that information. Precision describes how strongly the evaluation supports the post-result estimate. Clinical usefulness asks whether acting at an accepted threshold does more good than harm; this study did not test management, outcomes, or net benefit, and decision-curve analysis is one way to study that downstream question.[22]

The close correlation between percentage uncertainty reduction and Youden’s J (rho 0.971) matters. It argues against treating entropy reduction as a superior universal ranking measure. The information framework nevertheless retains a distinct interpretation as learning beyond the observed outcome frequency, depends on that frequency, and separates the average contributions of positive and negative classifications. Those features make it useful for explaining what a result added, not for grading calculators.

PERC and HEART show why the distinctions matter. The PERC evaluation combined high sensitivity and a post-negative risk below its prespecified target with only 4.3% average uncertainty reduction, because most patients received a positive classification that changed risk little. The HEART evaluations show that moving the threshold of one score in the same cohort can shift the information contribution from predominantly negative to predominantly positive. The quantitative meaning belongs to the evaluated threshold, population, outcome, and time horizon.

In a defined clinical population, starting risk also changes how much new information a calculator can provide. When an outcome is rare, a high negative predictive value largely reflects that most patients do not have the outcome. Lowering starting risk from 40% to 4% in the constructed example raised negative predictive value from 90.0% to 99.3% while uncertainty reduction fell from 20.7% to 12.6% (table S7). Previous diagnostic-test studies report the same dependence.[6,7,9,10,23]

The rule-out analysis adds the precision layer. A point estimate says where risk was observed to end; its denominator and interval say how low a risk the evaluation can support. Evaluations reporting no missed cases were especially vulnerable to overinterpretation because zero failures among a small negative group remains compatible with a clinically important miss rate. In this collection, 90 of 157 evaluations with a post-negative point estimate below 2% had an upper bound at or above 2%. This finding does not label those calculators unsafe or incompletely validated; it shows what the included evaluation can establish.

Strengths include the catalogue scale, recalculation from frozen complete or reconstructed 2×2 cells, internal calculation checks, separate analysis of positive and negative classifications, and resampling that kept repeated evaluations of the same calculator together. The principal limitation is that the evidence collection was fixed rather than systematic, so estimates describe the included evaluations rather than each calculator’s complete validation literature. Some counts were reconstructed from rounded summaries, the model-derived and legacy rows do not have a complete independent extraction audit trail, not every source was checked twice, and design was unclear for 459 evaluations. The source data did not systematically record sex or gender. Shared cohorts across different calculators were not consistently identifiable. Estimates at the observed outcome frequency should not be transported to a target population when sampling design is unclear or outcome selected. The threshold analysis used a spanning set rather than endorsed thresholds and measured statistical support, not benefit from acting. These concerns accord with published methodological reviews of clinical prediction-rule evaluation.[24,25]

For a defined population, outcome, threshold, and time horizon, calculator reports could state the starting risk, observed risk after each classification with its denominator and interval, and, when average learning is of interest, the percentage uncertainty removed and the classification supplying it. Future prospective studies should test whether presenting these quantities changes testing, treatment, or disposition and whether those changes improve outcomes.

## Conclusions

A clinical calculator result has no single quantitative meaning. Conventional performance describes classification, uncertainty reduction describes what was learned beyond the observed outcome frequency, post-result risk describes where risk ended, classification-specific contributions show which result supplied the information, and precision determines how strongly the risk is supported. Whether acting on the result improves care remains a separate question.

## Supporting information

Supplementary methods and results

Reproducibility archive: code, data and analysis outputs

## Data Availability

The submission includes a reproducibility archive containing the frozen analysis script, source workbook, cleaned primary and expanded datasets, row-level audit, data dictionary, generated tables, and figures. The versioned repository is available at https://github.com/EMAI-Research/mdcalc-entropy. Original analysis code is released under the MIT License. First-party derived data, generated tables and figures, and documentation that the authors have authority to license are released under the Creative Commons Attribution 4.0 International licence (CC BY 4.0). Third-party source material remains subject to its original terms and is not relicensed.

## Article information

### Funding

This work was supported by the National Academy of Medicine under Agreement No. 2026A008797. The National Academy of Medicine had no role in study design; data collection, analysis, or interpretation; preparation, review, or approval of the manuscript; or the decision to submit. The authors were independent from the funder. Shuhan He, as guarantor, had full access to the data and had final responsibility for the decision to submit.

### Competing interests

S.H. reports employment by Mass General Physician Organization and Mass General Brigham University of Health Professions; consulting fees from Bayesian Science; an unpaid volunteer role at Health Tech Without Borders; board membership at ConductScience Foundation; and being the founder of ConductScience. J.N.G. reports consulting fees from AstraZeneca, CSL Behring, Takeda, Octapharma, Wellumio, Cayuga, and PurpleAI, unrelated to this work. All other authors declare no competing interests.

### Contributorship

Conceptualization: S.H., J.W.J., J.N.G., A.S.R., C.K., B.W.L., and D.M.L. Methodology: S.H., J.W.J., P.S., S.L., J.N.G., A.S.R., C.K., B.W.L., and D.M.L. Software: S.H. and P.S. Validation: S.H., J.W.J., P.S., A.G., P.J.S., A.M., S.L., and J.N.G. Formal analysis: S.H. and P.S. Investigation: S.H., J.W.J., P.S., A.G., P.J.S., A.M., and S.L. Data curation: S.H., P.S., A.G., P.J.S., and A.M. Visualization: S.H. and P.S. Writing, original draft: S.H. Writing, review and editing: all authors. Supervision: S.H., J.N.G., and A.S.R. Project administration: S.H. Funding acquisition: S.H. Shuhan He is the manuscript guarantor. The corresponding author attests that all listed authors meet authorship criteria and that no others meeting the criteria have been omitted. Anthropic Claude Sonnet 4 and OpenAI Codex were used for the research and manuscript tasks described in Methods; neither was an author, and the human authors reviewed and remain responsible for all submitted content.

### Transparency declaration

The manuscript’s guarantor affirms that this manuscript is an honest, accurate, and transparent account of the study being reported; that no important aspects of the study have been omitted; and that any differences from the study as planned have been explained.

### Dissemination plans

Because this study used aggregate published evidence and enrolled no participants, there are no individual results to return. We will share the findings with clinicians, clinical calculator developers, researchers, and interested members of the public through publication and a plain language summary accompanying the public code and data release.

## Notes

### Author Declarations

Mass General Brigham Human Research Affairs determined on 14 August 2026 that this secondary analysis of published aggregate data does not constitute human subjects research (REDCap 4737); IRB approval and participant consent were not required. Only publicly available aggregate data were used; no individual patient data, identifiable information or restricted datasets were used. The No response to the public-data question follows the portal instruction to answer No when an institutional determination exists.

