## Supplementary methods and results for "Meaning of clinical calculator results: a cross-sectional analysis of the MDCalc catalogue"

Preprint supplement version: 0.1.0

**Data freeze:** 11 August 2026

### Supplementary methods

#### S1. Catalogue assembly, eligibility, mapping, and evidence provenance

The analysis used the complete set of 847 unique MDCalc catalogue identifiers in the frozen source workbook as its calculator-level sampling frame. The evidence collection supplied 494 candidate study-level binary evaluations. Of these, 487 linked to catalogue calculators; removing one superseded reconstruction and four exact duplicates left 482 primary evaluations representing 407 unique catalogue calculators. The 847 calculators and 494 candidate evaluations are different units, not consecutive attrition denominators. This was a catalogue-based cross-sectional analysis of a fixed evidence collection, not a systematic review of every validation study for every calculator. A calculator absent from the 482 evaluations was not judged invalid; it lacked a primary binary evaluation in the assembled evidence collection.

Evaluations were linked to catalogue identifiers by exact standardised names and a documented alias table. Among the 482 primary evaluations, 381 matched exactly, 100 used a documented alias, and one used a contained-name match. Seven eligible evaluations representing four tools or strategies lacked catalogue identifiers and were retained only in the expanded sensitivity analysis. Exact duplicates had the same calculator identity and true-positive, false-positive, false-negative, and true-negative cells; distinct populations, outcomes, and thresholds were retained. Direct 2×2 extraction and reported event counts took priority over legacy records, with metadata completeness resolving remaining ties.

Evidence was assigned to four mutually exclusive and exhaustive routes: direct count-based source evidence, reconstruction from reported sensitivity and specificity, reconstruction from other reported metrics, or legacy records without a recorded data source. All measures were recalculated from the final four cells. A study citation was recorded for 417 evaluations, a threshold for 411, and a high or moderate-high confidence label for 187. Rounded published metrics can correspond to more than one integer 2×2 table. The 19 metric-reconstructed records therefore retained the frozen cells, while analyses restricted to 331 records with direct count-based source evidence and 480 whole-number evaluations tested sensitivity to reconstruction. Source-report comparison was performed for flagged discrepancies and selected clinical examples rather than duplicate verification of every row.

Sampling design was not systematically recorded. Explicit source-description terms identified 21 cohort or consecutive evaluations, two case-control or two-gate evaluations, and 459 evaluations with unclear design. Excluding the two known outcome-selected evaluations did not materially change the headline estimate (table S4). One source count was corrected: the PRIEST record listed TP=4, but TP=44 reproduced the reported sensitivity of 83.0%, specificity of 69.4%, and a total of 177 participants. The source article reported 178 participants, leaving a one-participant denominator discrepancy.[13] All measures were recalculated from the four cells, so legacy spreadsheet formula errors did not propagate into this analysis.

Fig S1 Selection of binary evaluations and representation of the MDCalc catalogue. Panel A follows study-level evaluations from the assembled workbook to the 482 primary evaluations and then to the 407 represented calculators; the annotation gives the exact five exclusions. Panel B shows representation of the 847-calculator catalogue and near-complete representation of calculators labelled for rule-out use. The counts 847 and 494 describe different units-catalogue calculators and study-level evaluations-and are not successive stages of attrition.

#### Table S1. Evidence provenance and audit coverage

| Attribute | Evaluations, No. (%) |
|---|---|
| Direct count-based source evidence | 331 (68.7) |
| Reconstructed from reported sensitivity and specificity | 64 (13.3) |
| Reconstructed from other reported metrics | 19 (3.9) |
| Legacy records without a recorded data source | 68 (14.1) |
| Study citation recorded | 417 (86.5) |
| Threshold recorded | 411 (85.3) |
| High or moderate-high confidence | 187 (38.8) |
| Whole-number 2×2 cells | 480 (99.6) |
| Computational consistency checks passed | 482 (100.0) |

Percentages may not total 100 because of rounding. The row-level audit and full data dictionary are provided in the reproducibility archive.

#### Table S2. Excluded, superseded, duplicate, and off-catalogue evaluations

| Record or tool | Status | Reason |
|---|---|---|
| PLASMIC Score for TTP, source row 246 | Excluded | Superseded scaled-count extraction |
| Rapid Arterial oCclusion Evaluation Scale for Stroke, source row 256 | Excluded | Exact duplicate 2×2 evaluation |
| Edinburgh Postnatal Depression Scale, source row 425 | Excluded | Exact duplicate 2×2 evaluation |
| GAD-7, source row 448 | Excluded | Exact duplicate 2×2 evaluation |
| HARK, source row 494 | Excluded | Exact duplicate 2×2 evaluation |
| PedSRC Blunt Abdominal Trauma Rule, two evaluations | Expanded cohort only | No MDCalc catalogue identifier |
| Pediatric mental-health action screen, two evaluations | Expanded cohort only | No MDCalc catalogue identifier |
| CARE chest-pain rule, two evaluations | Expanded cohort only | No MDCalc catalogue identifier |
| Serial troponin strategy, one evaluation | Expanded cohort only | No MDCalc catalogue identifier |

### S2. Information-theory and statistical calculations

The main manuscript defines binary Shannon entropy, conditional entropy, information gain, and proportional reduction in uncertainty. Mutual information is also the expected improvement in logarithmic score after replacing one cohort-wide outcome probability with the risks observed after positive and negative results.

Proportional reduction is the fraction of intercept-only log loss, or null deviance, explained by those result-specific risks. These mathematical equivalences do not make the measure one of clinical utility or net benefit.

Let sensitivity be $Se$, specificity be $Sp$, outcome prevalence be $p$, and $q = P(T+) = pSe + (1-p)(1-Sp)$. With $h(\cdot)$ denoting binary entropy,

$$I(Y;T) = h(q) - p\,h(Se) - (1-p)\,h(1-Sp).$$

Youden's J is $Se + Sp - 1$. It retains only the sum of sensitivity and specificity, whereas mutual information also depends on prevalence and the separate sensitivity and specificity terms. The positive and negative result contributions were calculated as probability-weighted Kullback-Leibler divergences:

$$I_+ = P(T+)D_{KL}\{P(Y \mid T+) \parallel P(Y)\}, \qquad I_- = P(T-)D_{KL}\{P(Y \mid T-) \parallel P(Y)\}.$$

The contributions are non-negative and sum to total information. A result can contribute more because it is common, because it moves outcome risk farther from the starting risk, or both. The positive-result share was $I_+/(I_+ + I_-)$, and the negative share was its complement. Shares above 60% or below 40% were used descriptively to label positive-result and negative-result dominance; they were not clinical thresholds.

Confidence intervals used 2000 calculator-level bootstrap samples. All evaluations belonging to a selected calculator travelled together, the random-number seed was 20260811, and the 2.5th and 97.5th percentiles defined the 95% interval. This preserved dependence among repeated thresholds, outcomes, or populations for one calculator, although shared cohorts across different calculators could not be identified consistently. The one-evaluation-per-calculator analysis prioritised direct count-based source evidence, reported sensitivity and specificity, metric-reconstructed evidence, and legacy records without a recorded data source in that order, then retained the largest sample within evidence class.

Miller-Madow corrections were applied separately to the entropy of the binary outcome, the binary classification, and the joint 2×2 table using the observed number of non-empty categories. Corrected mutual information was corrected outcome entropy plus corrected classification entropy minus corrected joint entropy, constrained to be non-negative. The corrected percentage used corrected starting uncertainty as its denominator. Fixed-prevalence analyses applied each evaluation's sensitivity and specificity at outcome prevalences of 5%, 10%, 25%, and 50%; these are mathematical standardisations rather than transport estimates for new populations.

The seven examples in the main manuscript were selected to clarify interpretation, not to rank calculators. The complete source ledger, thresholds, four-cell tables, MDCalc URLs, and derived measures remain in the reproducibility archive. For PERC, the 2.0% validation target for venous thromboembolism or death within 45 days and the 1.8% development testing threshold for pulmonary embolism were treated as different endpoints.[16,17] In the validation cohort, starting risk was 7.58%, the combined low-clinical-suspicion and PERC-negative classification had a negative likelihood ratio of 0.118, and observed 45-day composite risk after a negative result was 1.0%. This comparison does not establish clinical net benefit.

Each information estimate applies to one included 2×2 table at its observed outcome frequency. When the source population was outcome selected or sampling design was unclear, the estimate should not be read as expected information in an unselected target population. Fixed-prevalence calculations isolate one mathematical consequence of changing starting risk, but they do not establish transportability. Clinical usefulness remains a separate question that depends on post-result risk, an accepted decision threshold, and the consequences of acting.

### Supplementary results and robustness

#### Table S3. Key correlation results

| Outcome | Predictor | Method | Estimate | Calculator-clustered 95% CI |
|---|---|---|---|---|
| Reduction in uncertainty, % | Youden's J | Spearman | 0.971 | 0.964-0.976 |
| Information gain, bits | Youden's J | Spearman | 0.855 | 0.820-0.880 |
| Information gain, bits | Observed outcome frequency | Spearman | 0.673 | 0.593-0.738 |
| Information gain, bits | Starting uncertainty | Spearman | 0.686 | 0.611-0.748 |

The complete prespecified Spearman and secondary Pearson correlation matrix is provided in the reproducibility archive.

#### Table S4. Primary robustness and sensitivity analyses

| Analysis | Evaluations | Estimate | Calculator-clustered 95% CI |
|---|---|---|---|
| Primary median reduction | 482 | 14.9% (0.093 bits) | 12.1-17.2% (0.065-0.108 bits) |
| Direct count-based source evidence | 331 | 14.7% (0.091 bits) | - |
| Whole-number cells | 480 | 14.8% (0.092 bits) | - |
| High or moderate-high confidence | 187 | 13.0% (0.077 bits) | - |
| One evaluation per calculator | 407 | 15.7% (0.100 bits) | - |
| Expanded off-catalogue cohort | 489 | 14.7% (0.092 bits) | - |
| Excluding explicit case-control or two-gate records | 480 | 14.8% (0.092 bits) | - |
| Miller-Madow-corrected median reduction | 482 | 14.4% (0.092 bits) | - |
| Model-assisted extraction | 157 | 16.6% | - |
| Other extraction routes | 325 | 13.0% | - |
| Median negative-result information share among rule-out evaluations | 56 | 71.3% | 65.6-79.0% |

Rows labelled median or proportion report descriptive estimates. A dash indicates that the analysis was included to show the range of point estimates rather than add another interval.

#### Table S5. Fixed-prevalence information estimates

| Fixed prevalence | Median information gain, bits (IQR) | Median uncertainty removed, % (IQR) | Spearman rho with observed reduction |
|---|---|---|---|
| 5% | 0.036 (0.015-0.072) | 12.5 (5.2-25.1) | 0.983 |
| 10% | 0.067 (0.029-0.135) | 14.2 (6.1-28.7) | 0.985 |
| 25% | 0.135 (0.059-0.267) | 16.7 (7.3-32.9) | 0.985 |
| 50% | 0.178 (0.078-0.342) | 17.8 (7.8-34.2) | 0.979 |

The relative ordering of evaluation-level estimates changed little across fixed prevalences, but the amount of available starting uncertainty changed. This is not a calculator-quality ranking. These calculations hold sensitivity and specificity constant and do not assume that performance would remain constant in a different clinical population.

**Table S6. Same starting prevalence and Youden's J, different uncertainty reduction**

| Accuracy profile* | Sensitivity; specificity; J | Risk after positive; negative result | Starting uncertainty removed | Information from positive; negative result |
|---|---|---|---|---|
| **Rule-out emphasis** | 95.0%; 55.0%; 0.500 | 19.0%; 1.0% | 16.6%; 0.0779 bits | Positive 33.9%; **negative 66.1%** |
| **Balanced accuracy** | 75.0%; 75.0%; 0.500 | 25.0%; 3.6% | 14.9%; 0.0700 bits | Positive 57.1%; negative 42.9% |
| **Rule-in emphasis** | 55.0%; 95.0%; 0.500 | 55.0%; 5.0% | 23.9%; 0.112 bits | **Positive 80.6%**; negative 19.4% |

*Constructed populations of 10,000 people with 10% outcome prevalence. Youden's J is 0.500 in all three examples; redistributing sensitivity and specificity changes both total uncertainty reduction and whether positive or negative results contribute most of the information.

Fig S2 Relationship between Youden's J and information yield. Panel A shows absolute information gain by Youden's J, coloured by observed outcome frequency; panel B shows proportional uncertainty removed. Percentage reduction tracked Youden's J closely, whereas absolute gain also depended on the uncertainty available at the observed outcome frequency.

**Table S7. Same test performance at different starting risks**

| Population | Sensitivity; specificity | PPV; NPV | LR+; LR-; J | Reduction in uncertainty | Information from positive vs negative result | Clinical meaning |
|---|---|---|---|---|---|---|
| **40% prevalence***; 400 outcomes per 1000 | 90.0%; 60.0% | 60.0%; 90.0% | 2.2; 0.17; 0.500 | 20.7%; 0.201 bits | Positive 35.0%; **negative 65.0%** | When the outcome is relatively common, there is more uncertainty for the test to reduce. |
| **4% prevalence***; 40 outcomes per 1000 | 90.0%; 60.0% | 8.6%; 99.3% | 2.2; 0.17; 0.500 | 12.6%; 0.0306 bits | Positive 41.1%; negative 58.9% | Negative predictive value rises to 99.3%, even though the test adds much less new information. |

*Constructed examples using the same sensitivity and specificity in populations of 1000 people. PPV is positive predictive value; NPV is negative predictive value. The two rows hold sensitivity, specificity, likelihood ratios, and Youden's J constant so that the effect of starting prevalence is visible.

When starting risk fell from 40% to 4%, negative predictive value increased from 90.0% to 99.3%, but the calculator reduced less uncertainty (20.7% versus 12.6%). A very high negative predictive value in a low-risk population therefore does not necessarily mean that the calculator adds much new information.

The share of information assigned to one classification depends on the descriptive boundary. Result dominance occurred in 83.8% of evaluations at a 55% boundary, 68.3% at 60%, and 40.5% at 70%. A comparable proportion arose under simulated sensitivity and specificity distributions, so 68.3% should not be interpreted as an intrinsic defining property of clinical calculators.

### Supplementary clinical examples

#### Table S8. Exact values for the seven explanatory clinical examples

| Clinical decision and evaluation | Cohort; observed outcome frequency | Sensitivity; specificity | Risk after positive; after negative | Uncertainty removed; information from positive vs negative | Interpretation in this evaluation |
|---|---|---|---|---|---|
| **Suspected pulmonary embolism - PERC**[17]. Low clinical suspicion (<15%) and PERC negative; venous thromboembolism or death within 45 days | n=8138; risk 7.6% | 97.4%; 21.9% | 9.3%; 1.0% | 4.29% removed; positive 13.4%, negative 86.6% | The combined negative result crossed the prespecified 2.0% validation target despite modest average uncertainty removal. |
| **Minor head injury - Canadian CT Head Rule**[18]. Positive criteria; clinically important brain injury | n=2707; risk 8.5% | 100.0%; 41.1% | 13.7%; 0.0% | 14.6% removed; positive 21.3%, negative 78.7% | The evaluated cohort had no observed clinically important brain injury after a negative classification. |
| **Upper gastrointestinal bleeding - Glasgow-Blatchford**[19]. Score >1; intervention or death within 30 days | n=2932; risk 46.3% | 98.6%; 34.6% | 56.5%; 3.4% | 15.8% removed; positive 15.5%, negative 84.5% | Most information came from the negative result even though the outcome was common. |
| **Community acquired pneumonia - CRB-65**[20]. Score ≥1; 30-day mortality | n=553; risk 10.7% | 94.9%; 17.0% | 12.0%; 3.4% | 1.90% removed; positive 12.0%, negative 88.0% | Few patients had a negative result, so average uncertainty removal was small despite high sensitivity. |
| **Community acquired pneumonia - CURB-65**[20]. Score ≥2; 30-day mortality | n=553; risk 10.7% | 84.7%; 40.9% | 14.6%; 4.3% | 4.41% removed; positive 30.7%, negative 69.3% | In the same patients, this threshold removed more uncertainty than CRB-65 while negative results still supplied most information. |
| **Chest pain - HEART, rule-out threshold**[21]. HEART ≥4; six-week major adverse cardiac events | n=641; risk 9.5% | 100.0%; 69.7% | 25.7%; 0.0% | 32.9% removed; positive 39.1%, negative 60.9% | At this threshold, the negative result supplied most information. |
| **Chest pain - HEART, rule-in threshold**[21]. HEART >6; six-week major adverse cardiac events | n=641; risk 9.5% | 65.6%; 96.9% | 69.0%; 3.6% | 37.3% removed; positive 79.8%, negative 20.2% | At the higher threshold, the pattern reversed and the positive result supplied most information. |

Observed outcome frequency is $(TP + FN)/N$ in the analytic table; in these explicitly prospective or consecutive cohorts it can be read as starting risk for the evaluated population. Risk after a positive result is positive predictive value; risk after a negative result is one minus negative predictive value. Positive and negative contributions sum to 100%. These rows were selected to explain contrasts already present in the frozen collection, not to rank calculators or assert that each is the best available validation. The PERC endpoint was the broader composite of venous thromboembolism or death within 45 days; the 1.8% development threshold was estimated for pulmonary embolism testing. The HEART cohort size, event frequency, and rule-out performance were checked against the cited prospective study.[21]

### Reproducibility

The separately submitted reproducibility archive contains the frozen analysis code; shareable source and cleaned data; row-level audit; data dictionary; complete machine-readable tables and figures; the full clinical-example ledger; unsorted evaluation-level post-negative and post-positive support estimates; exploratory study-size, specialty, dominance-boundary, purpose-coverage, and sensitivity outputs removed from this PDF; and a machine-readable manifest with checksums. The archive README records the analysis version, data freeze, software environment, file map, regeneration commands, and frozen analysis identifier bmj-v0.14.0. Exploratory outputs retained there are not primary manuscript findings. The open repository mirrors the submitted analysis but does not replace the BMJ-hosted archive.
