## Supplementary material for "Meaning of clinical calculator results: a cross-sectional analysis of the MDCalc catalogue": Reproducibility archive: code, data and analysis outputs: analyze.py

```
"""Build the MDCalc manuscript dataset, tables, figures, and audit trail."""

from __future__ import annotations

import hashlib
import json
import re
import sys
from pathlib import Path

import matplotlib

matplotlib.use("Agg")
import matplotlib.pyplot as plt
import numpy as np
import pandas as pd
from scipy import stats


HERE = Path(__file__).resolve().parent
SOURCE = HERE / "source" / "mdcalc_entropy_source_v0.2.0.xlsx"
EXPECTED_SHA256 = "02348f2e95039be35943e135d27b12010ba382d589d03623a497baef35dbc514"
BMJ_STYLE = HERE / "bmj.mplstyle"
INK, MUTED, GRID, LIGHT, WHITE = "#17212B", "#58636D", "#D9E0E5", "#F5F7F9", "#FFFFFF"
COLOURS = ("#0072B2", "#009E73", "#CC79A7", "#E69F00")
GRAYS = ("#111111", "#3D3D3D", "#696969", "#969696")
DOUBLE_COLUMN_MM, MM_PER_INCH, FIGURE_DPI = 180.0, 25.4, 600
plt.rcParams["svg.hashsalt"] = "mdcalc-entropy-bmj-v0.14.0"


ALIASES: list[tuple[str, int | None, str]] = [
    (r"absolute lymphocyte count", 2203, "Absolute Lymphocyte Count (ALC)"),
    (r"ascvd 2013 risk", 3398, "ASCVD 2013 Risk Calculator"),
    (r"geneva risk score for vte prophylaxis", 10073, "Geneva VTE Prophylaxis Risk Score"),
    (r"hfa icos.*multi targeted kinase", 10646, "HFA-ICOS CML Cardio-Oncology Risk"),
    (r"homa ir", 3120, "HOMA-IR"),
    (r"improvedd", 10350, "IMPROVEDD VTE Risk Score"),
    (r"isth s[sc]c bleeding", 10580, "ISTH Bleeding Assessment Tool"),
    (r"nirudak", 10593, "NIRUDAK Score"),
    (r"pediatric nexus ii", 10078, "Pediatric NEXUS II"),
    (r"pedsrc", None, "PedSRC Blunt Abdominal Trauma Rule"),
    (r"rcvs2", 10347, "RCVS2 Score"),
    (r"combined meld.*meld 3", 10437, "Combined MELD"),
    (r"qids sr16", 1845, "Quick Inventory of Depressive Symptomatology"),
    (r"solitary pulmonary nodule", 4057, "Mayo SPN Malignancy Risk Score"),
    (r"s t o n e nephrolithometry", 2044, "STONE Nephrolithometry Score"),
    (r"sick control one fat food", 10524, "SCOFF Questionnaire"),
    (r"edinburgh postnatal depression", 10466, "Edinburgh Postnatal Depression Scale"),
    (r"egsys", 3948, "EGSYS Syncope Score"),
    (r"el ganzouri", 10149, "El-Ganzouri Risk Index"),
    (r"emergency department assessment of chest pain", 1858, "EDACS"),
    (r"eutos score", 3906, "EUTOS Score"),
    (r"fatty liver index", 10001, "Fatty Liver Index"),
    (r"fibrosis 4", 2200, "FIB-4 Index"),
    (r"focused assessment with sonography", 4037, "FAST"),
    (r"gad 7", 1727, "GAD-7"),
    (r"galad model", 10094, "GALAD Model"),
    (r"hacor", 10460, "HACOR Score"),
    (r"harmless acute pancreatitis|using haps score", 3286, "Harmless Acute Pancreatitis Score"),
    (r"has bled|derivation bleed score", 807, "HAS-BLED"),
    (r"sudden cardiac death risk.*hypertrophic", 10481, "HCM Risk-SCD"),
    (r"cappelli.*pediatric mental health", None, "Pediatric mental-health action screen"),
    (r"moumneh.*care rule", None, "CARE chest-pain rule"),
    (r"moumneh.*heart score", 1752, "HEART Score"),
    (r"moumneh.*timi score", 111, "TIMI Risk Score for UA/NSTEMI"),
    (r"moumneh.*grace score", 1099, "GRACE ACS Calculator"),
    (r"heart pathway", 3975, "HEART Pathway"),
    (r"serial troponin", None, "Serial troponin strategy"),
    (r"hemorr2hages", 1785, "HEMORR2HAGES Score"),
    (r"hints three step|hints for stroke", 10184, "HINTS"),
    (r"hit testing using hep score", 1789, "HEP Score"),
    (r"he macs", 10165, "HE-MACS"),
    (r"hark to identify", 10420, "HARK"),
    (r"hscore", 10089, "HScore"),
    (r"interchest", 10225, "INTERCHEST"),
]


MAIN_ATLAS_ROWS = [
    {
        "source_index": 241,
        "block": "Stopping safely",
        "label": "Suspected pulmonary embolism — PERC",
        "context": "Low clinical suspicion (<15%) and PERC negative; venous thromboembolism or death within 45 days",
        "reading": "The combined negative result lowered risk to 1.0%, below the prespecified 2.0% validation target, despite modest average information.",
        "reference": 17,
    },
    {
        "source_index": 65,
        "block": "Stopping safely",
        "label": "Minor head injury — Canadian CT Head Rule",
        "context": "CCHR positive criteria; clinically important brain injury",
        "reading": "The evaluated cohort had no observed clinically important brain injury after a negative classification.",
        "reference": 18,
    },
    {
        "source_index": 145,
        "block": "Stopping safely",
        "label": "Upper gastrointestinal bleeding — Glasgow-Blatchford score",
        "context": "GBS >1; hospital intervention or death within 30 days",
        "reading": "Negative-result dominance also appears when the outcome is common, not only when it is rare.",
        "reference": 19,
    },
    {
        "source_index": 97,
        "block": "Same pneumonia cohort",
        "label": "Community acquired pneumonia — CRB-65",
        "context": "CRB-65 ≥1; 30-day mortality",
        "reading": "High sensitivity with very low specificity leaves little average information because few patients test negative.",
        "reference": 20,
    },
    {
        "source_index": 101,
        "block": "Same pneumonia cohort",
        "label": "Community acquired pneumonia — CURB-65",
        "context": "CURB-65 ≥2; 30-day mortality",
        "reading": "In the same cohort, added specificity more than doubles information while negative results remain dominant.",
        "reference": 20,
    },
    {
        "source_index": 473,
        "block": "Same HEART score, different thresholds",
        "label": "Chest pain — HEART rule-out threshold",
        "context": "HEART ≥4; six-week major adverse cardiac events",
        "reading": "At the rule-out threshold, the negative result supplies most of the information.",
        "reference": 21,
    },
    {
        "source_index": 474,
        "block": "Same HEART score, different thresholds",
        "label": "Chest pain — HEART rule-in threshold",
        "context": "HEART >6; six-week major adverse cardiac events",
        "reading": "Moving the threshold reverses the pattern: the positive result now carries most of the information.",
        "reference": 21,
    },
]


SUPPLEMENTAL_ATLAS_ROWS = [
    {
        "source_index": 75,
        "block": "Positive, balanced, and rare-outcome patterns",
        "label": "Pharyngitis — modified Centor/McIsaac score",
        "context": "Score ≥2; group A streptococcal pharyngitis",
        "reading": "The positive result dominates, a different information pattern from the rule-out tools.",
    },
    {
        "source_index": 3,
        "block": "Positive, balanced, and rare-outcome patterns",
        "label": "COVID-19 — 4C mortality score",
        "context": "Score >9; in-hospital mortality",
        "reading": "Positive and negative results contribute almost equally, showing that some scores genuinely inform both ways.",
    },
    {
        "source_index": 456,
        "block": "Positive, balanced, and rare-outcome patterns",
        "label": "Atrial fibrillation — HAS-BLED",
        "context": "Score ≥3; one-year major bleeding",
        "reading": "A 98.7% NPV coexists with 0.003 bits per patient because major bleeding was rare before scoring.",
    },
    {
        "source_index": 18,
        "block": "Additional rule-out and early-disposition tools",
        "label": "Chest pain — ADAPT protocol",
        "context": "ADAPT positive; major adverse cardiac events",
        "reading": "Near-perfect sensitivity and modest specificity concentrate most information in the negative result.",
    },
    {
        "source_index": 46,
        "block": "Additional rule-out and early-disposition tools",
        "label": "Child with cerebrospinal fluid pleocytosis — Bacterial Meningitis Score",
        "context": "Score >0; bacterial meningitis",
        "reading": "The very-low-risk negative result carries most information despite a low event frequency.",
    },
    {
        "source_index": 164,
        "block": "Additional rule-out and early-disposition tools",
        "label": "Acute vestibular syndrome — HINTS",
        "context": "Any dangerous HINTS sign; central stroke",
        "reading": "High sensitivity and specificity remove most available uncertainty, with the negative examination contributing more.",
    },
    {
        "source_index": 220,
        "block": "Additional rule-out and early-disposition tools",
        "label": "Blunt trauma — NEXUS cervical spine criteria",
        "context": "Any NEXUS criterion; clinically important cervical spine injury",
        "reading": "Near-perfect sensitivity coexists with little average yield because few patients satisfy every low-risk criterion.",
    },
    {
        "source_index": 230,
        "block": "Additional rule-out and early-disposition tools",
        "label": "Ankle injury — Ottawa Ankle Rule",
        "context": "Any ankle-zone criterion; fracture",
        "reading": "Perfect sensitivity here is clinically directional, while low specificity limits how many avoid imaging.",
    },
    {
        "source_index": 232,
        "block": "Additional rule-out and early-disposition tools",
        "label": "Acute headache — Ottawa SAH Rule",
        "context": "Any Ottawa SAH criterion; subarachnoid haemorrhage",
        "reading": "Perfect sensitivity here removes modest average uncertainty because informative negative results are uncommon.",
    },
    {
        "source_index": 234,
        "block": "Additional rule-out and early-disposition tools",
        "label": "Paediatric head injury — PECARN",
        "context": "Any age-specific predictor; clinically important traumatic brain injury",
        "reading": "With injury uncommon, a perfect-sensitivity rule still removes 13% of available uncertainty.",
    },
    {
        "source_index": 151,
        "block": "Positive-result and balanced tools",
        "label": "Non-invasive ventilation — HACOR",
        "context": "HACOR >5 at one hour; non-invasive ventilation failure",
        "reading": "A high-specificity threshold makes ventilation failure a positive-result-dominant pattern.",
    },
    {
        "source_index": 156,
        "block": "Positive-result and balanced tools",
        "label": "Chest pain — HEART score validation",
        "context": "HEART ≥4; major adverse cardiac events",
        "reading": "A low-risk HEART threshold again concentrates information in the negative result.",
    },
    {
        "source_index": 211,
        "block": "Positive-result and balanced tools",
        "label": "Left bundle branch block — modified Sgarbossa criteria",
        "context": "Any modified criterion; acute coronary occlusion",
        "reading": "The positive ECG pattern carries three quarters of the information, matching a rule-in task.",
    },
    {
        "source_index": 251,
        "block": "Positive-result and balanced tools",
        "label": "Pulmonary embolism — PESI",
        "context": "PESI >85; short-term mortality",
        "reading": "The negative result supplies most information for identifying low-risk pulmonary embolism.",
    },
    {
        "source_index": 252,
        "block": "Positive-result and balanced tools",
        "label": "Suspected infection — qSOFA",
        "context": "qSOFA ≥2; in-hospital mortality",
        "reading": "Positive results contribute most information, consistent with escalation rather than exclusion.",
    },
    {
        "source_index": 253,
        "block": "Positive-result and balanced tools",
        "label": "COVID-19 — quick COVID-19 Severity Index",
        "context": "qCSI >3; respiratory decompensation within 24 hours",
        "reading": "Positive and negative results contribute nearly evenly to early deterioration prediction.",
    },
    {
        "source_index": 399,
        "block": "Positive-result and balanced tools",
        "label": "Flank pain — STONE score",
        "context": "STONE 10–13; uncomplicated ureteral stone",
        "reading": "Positive results carry most information, but neither result alone determines whether imaging is needed.",
    },
]


SOURCE_CITATION_OVERRIDES = {
    241: "Kline JA, Courtney DM, Kabrhel C, et al. Prospective multicenter evaluation of the pulmonary embolism rule-out criteria. J Thromb Haemost. 2008;6(5):772-780. doi:10.1111/j.1538-7836.2008.02944.x",
    65: "Stiell IG, Clement CM, Rowe BH, et al. Comparison of the Canadian CT Head Rule and the New Orleans Criteria in patients with minor head injury. JAMA. 2005;294(12):1511-1518. doi:10.1001/jama.294.12.1511",
    145: "Stanley AJ, Laine L, Dalton HR, et al. Comparison of risk scoring systems for patients presenting with upper gastrointestinal bleeding: international multicentre prospective study. BMJ. 2017;356:i6432. doi:10.1136/bmj.i6432",
    97: "Nüllmann H, Pflug MA, Wesemann T, et al. External validation of the CURSI criteria in adults hospitalised for community-acquired pneumonia. BMC Infect Dis. 2014;14:39. doi:10.1186/1471-2334-14-39",
    101: "Nüllmann H, Pflug MA, Wesemann T, et al. External validation of the CURSI criteria in adults hospitalised for community-acquired pneumonia. BMC Infect Dis. 2014;14:39. doi:10.1186/1471-2334-14-39",
    473: "Moumneh T, Richard-Jourjon V, Friou E, et al. Reliability of the CARE rule and the HEART score to rule out an acute coronary syndrome in non-traumatic chest pain patients. Intern Emerg Med. 2018;13(7):1111-1119. doi:10.1007/s11739-018-1803-4",
    474: "Moumneh T, Richard-Jourjon V, Friou E, et al. Reliability of the CARE rule and the HEART score to rule out an acute coronary syndrome in non-traumatic chest pain patients. Intern Emerg Med. 2018;13(7):1111-1119. doi:10.1007/s11739-018-1803-4",
    75: "McIsaac WJ, Kellner JD, Aufricht P, Vanjaka A, Low DE. Empirical validation of guidelines for the management of pharyngitis in children and adults. JAMA. 2004;291(13):1587-1595. doi:10.1001/jama.291.13.1587",
    3: "Aletreby WT, Mumtaz SA, Shahzad SA, et al. External validation of 4C ISARIC Mortality Score in critically ill COVID-19 patients from Saudi Arabia. Saudi J Med Med Sci. 2022;10(1):19-24. doi:10.4103/sjmms.sjmms_480_21",
    456: "Pisters R, Lane DA, Nieuwlaat R, de Vos CB, Crijns HJGM, Lip GYH. A novel user-friendly score (HAS-BLED) to assess 1-year risk of major bleeding in patients with atrial fibrillation. Chest. 2010;138(5):1093-1100. doi:10.1378/chest.10-0134",
}


def normalize(value: object) -> str:
    text = str(value).lower().replace("₂", "2").replace("–", "-").replace("—", "-")
    text = re.sub(r"/none\s*$", "", text)
    text = re.sub(r"[^a-z0-9]+", " ", text)
    return " ".join(text.split())


def binary_entropy(values: pd.Series | np.ndarray) -> np.ndarray:
    p = np.asarray(values, dtype=float)
    out = np.zeros_like(p)
    mask = (p > 0) & (p < 1)
    out[mask] = -(p[mask] * np.log2(p[mask]) + (1 - p[mask]) * np.log2(1 - p[mask]))
    return out


def bernoulli_kl_divergence(
    posterior: pd.Series | np.ndarray, prior: pd.Series | np.ndarray
) -> np.ndarray:
    q = np.asarray(posterior, dtype=float)
    p = np.asarray(prior, dtype=float)
    with np.errstate(divide="ignore", invalid="ignore"):
        event_term = np.where(q > 0, q * np.log2(q / p), 0.0)
        nonevent_term = np.where(q < 1, (1 - q) * np.log2((1 - q) / (1 - p)), 0.0)
    return event_term + nonevent_term


def safe_div(num: pd.Series, den: pd.Series) -> pd.Series:
    return num.div(den.where(den.ne(0)))


def add_metrics(df: pd.DataFrame) -> pd.DataFrame:
    out = df.copy()
    tp, fp, fn, tn = (out[c] for c in ["tp", "fp", "fn", "tn"])
    total, diseased, nondiseased = tp + fp + fn + tn, tp + fn, fp + tn
    positive, negative = tp + fp, fn + tn
    out["count_total"] = total
    out["sensitivity"] = safe_div(tp, diseased)
    out["specificity"] = safe_div(tn, nondiseased)
    out["prevalence"] = safe_div(diseased, total)
    out["ppv"] = safe_div(tp, positive)
    out["npv"] = safe_div(tn, negative)
    out["youden_j"] = out["sensitivity"] + out["specificity"] - 1
    out["lr_positive"] = out["sensitivity"] / (1 - out["specificity"])
    out["lr_negative"] = (1 - out["sensitivity"]) / out["specificity"]
    out["diagnostic_odds_ratio"] = out["lr_positive"] / out["lr_negative"]
    out["parent_entropy_bits"] = binary_entropy(out["prevalence"])
    out["positive_fraction"] = safe_div(positive, total)
    out["negative_fraction"] = safe_div(negative, total)
    out["positive_entropy_bits"] = binary_entropy(safe_div(tp, positive).fillna(0))
    out["negative_entropy_bits"] = binary_entropy(safe_div(fn, negative).fillna(0))
    out["conditional_entropy_bits"] = (
        out["positive_fraction"].fillna(0) * out["positive_entropy_bits"]
        + out["negative_fraction"].fillna(0) * out["negative_entropy_bits"]
    )
    out["entropy_reduction_bits"] = out["parent_entropy_bits"] - out["conditional_entropy_bits"]
    out["entropy_removed_percent"] = 100 * safe_div(
        out["entropy_reduction_bits"], out["parent_entropy_bits"]
    )
    observed_test_categories = pd.concat([positive, negative], axis=1).gt(0).sum(axis=1)
    observed_outcome_categories = pd.concat([diseased, nondiseased], axis=1).gt(0).sum(axis=1)
    observed_joint_categories = out[["tp", "fp", "fn", "tn"]].gt(0).sum(axis=1)
    entropy_bias_unit = 1 / (2 * total * np.log(2))
    out["parent_entropy_mm_bits"] = out["parent_entropy_bits"] + (
        observed_outcome_categories - 1
    ) * entropy_bias_unit
    out["entropy_reduction_mm_bits"] = np.maximum(
        0,
        out["entropy_reduction_bits"]
        + (
            observed_test_categories
            + observed_outcome_categories
            - observed_joint_categories
            - 1
        )
        * entropy_bias_unit,
    )
    out["entropy_removed_mm_percent"] = 100 * safe_div(
        out["entropy_reduction_mm_bits"], out["parent_entropy_mm_bits"]
    )
    positive_event_probability = safe_div(tp, positive).fillna(out["prevalence"])
    negative_event_probability = safe_div(fn, negative).fillna(out["prevalence"])
    out["positive_information_bits"] = out["positive_fraction"].fillna(0) * bernoulli_kl_divergence(
        positive_event_probability, out["prevalence"]
    )
    out["negative_information_bits"] = out["negative_fraction"].fillna(0) * bernoulli_kl_divergence(
        negative_event_probability, out["prevalence"]
    )
    out["positive_information_share_percent"] = 100 * safe_div(
        out["positive_information_bits"], out["entropy_reduction_bits"]
    )
    out["negative_information_share_percent"] = 100 - out["positive_information_share_percent"]
    out["information_contribution_pattern"] = np.select(
        [
            out["positive_information_share_percent"].gt(60),
            out["positive_information_share_percent"].lt(40),
        ],
        ["Positive-result dominant", "Negative-result dominant"],
        default="Balanced",
    )
    return out


def confidence_group(value: object) -> str:
    text = normalize(value)
    if not text or text == "nan":
        return "Not reported"
    if "high" in text and "moderate" in text:
        return "Moderate-high"
    if "high" in text:
        return "High"
    if "moderate" in text or "medium" in text:
        return "Moderate"
    if "low" in text:
        return "Low"
    return "Other"


def evidence_group(value: object) -> str:
    text = normalize(value)
    if not text or text == "nan":
        return "Legacy/no recorded source"
    if (
        "metric reconstructed" in text
        or "rounded metrics" in text
        or "literature" in text
        or "meta analysis" in text
    ):
        return "Metric-reconstructed"
    if "sens spec" in text:
        return "Reported sensitivity/specificity"
    if (
        "event counts" in text
        or "validation counts" in text
        or "2x2" in text
        or text == "v1 raw"
    ):
        return "Count-based"
    return "Other"


def sampling_design_group(value: object) -> str:
    """Classify only study designs stated explicitly in the recorded source description."""
    text = normalize(value)
    if re.search(r"\b(case control|two gate)\b", text):
        return "Case-control/two-gate"
    if re.search(r"\b(cohort|consecutive)\b", text):
        return "Cohort/consecutive"
    return "Unclear"


def match_catalog(label: str, catalog: pd.DataFrame) -> tuple[float | None, str, str]:
    query = normalize(label)
    for pattern, calc_id, canonical in ALIASES:
        if re.search(pattern, query):
            return (float(calc_id) if calc_id is not None else None, canonical, "Manual alias")

    exact = catalog[catalog["match_name"].eq(query)]
    if len(exact) == 1:
        row = exact.iloc[0]
        return float(row["catalog_id"]), row["catalog_name"], "Exact"

    contained = catalog[
        catalog["match_name"].map(lambda name: len(name) >= 12 and (name in query or query in name))
    ]
    if len(contained):
        row = contained.assign(length=contained["match_name"].str.len()).sort_values("length").iloc[-1]
        return float(row["catalog_id"]), row["catalog_name"], "Contained name"
    return None, label, "Unmatched"


def sha256(path: Path) -> str:
    digest = hashlib.sha256()
    with path.open("rb") as handle:
        for block in iter(lambda: handle.read(1024 * 1024), b""):
            digest.update(block)
    return digest.hexdigest()


def cluster_bootstrap_correlation(
    frame: pd.DataFrame, x: str, y: str, method: str, iterations: int = 2000
) -> tuple[float, float]:
    clean = frame[["calculator_key", x, y]].replace([np.inf, -np.inf], np.nan).dropna()
    groups = [group[[x, y]].to_numpy() for _, group in clean.groupby("calculator_key")]
    rng = np.random.default_rng(20260811)
    estimates = np.empty(iterations)
    for index in range(iterations):
        sampled = rng.integers(0, len(groups), len(groups))
        boot = np.vstack([groups[group_index] for group_index in sampled])
        if method == "spearman":
            boot = np.column_stack([stats.rankdata(boot[:, 0]), stats.rankdata(boot[:, 1])])
        estimates[index] = np.corrcoef(boot[:, 0], boot[:, 1])[0, 1]
    return tuple(np.nanpercentile(estimates, [2.5, 97.5]))


def partial_spearman(x: np.ndarray, y: np.ndarray, control: np.ndarray) -> float:
    ranked = np.column_stack(
        [stats.rankdata(values) for values in (x, y, control)]
    )
    design = np.column_stack([np.ones(len(ranked)), ranked[:, 2]])
    x_residual = ranked[:, 0] - design @ np.linalg.lstsq(
        design, ranked[:, 0], rcond=None
    )[0]
    y_residual = ranked[:, 1] - design @ np.linalg.lstsq(
        design, ranked[:, 1], rcond=None
    )[0]
    return float(np.corrcoef(x_residual, y_residual)[0, 1])


def cluster_bootstrap_partial_spearman(
    frame: pd.DataFrame,
    x: str,
    y: str,
    control: str,
    iterations: int = 2000,
) -> tuple[float, float]:
    clean = frame[["calculator_key", x, y, control]].replace(
        [np.inf, -np.inf], np.nan
    ).dropna()
    groups = [
        group[[x, y, control]].to_numpy()
        for _, group in clean.groupby("calculator_key")
    ]
    rng = np.random.default_rng(20260811)
    estimates = np.empty(iterations)
    for index in range(iterations):
        sampled = rng.integers(0, len(groups), len(groups))
        boot = np.vstack([groups[group_index] for group_index in sampled])
        estimates[index] = partial_spearman(boot[:, 0], boot[:, 1], boot[:, 2])
    return tuple(np.nanpercentile(estimates, [2.5, 97.5]))


def describe(series: pd.Series) -> dict[str, float | int]:
    clean = series.replace([np.inf, -np.inf], np.nan).dropna()
    return {
        "n": int(len(clean)),
        "mean": float(clean.mean()),
        "sd": float(clean.std()),
        "median": float(clean.median()),
        "q1": float(clean.quantile(0.25)),
        "q3": float(clean.quantile(0.75)),
        "min": float(clean.min()),
        "max": float(clean.max()),
    }


def correlation_table(frame: pd.DataFrame) -> pd.DataFrame:
    predictors = [
        "sensitivity",
        "specificity",
        "youden_j",
        "prevalence",
        "parent_entropy_bits",
        "reported_sample_size",
    ]
    outcomes = ["entropy_reduction_bits", "entropy_removed_percent"]
    rows = []
    for outcome in outcomes:
        for predictor in predictors:
            clean = frame[[predictor, outcome]].replace([np.inf, -np.inf], np.nan).dropna()
            for method in ["spearman", "pearson"]:
                result = (
                    stats.spearmanr(clean[predictor], clean[outcome])
                    if method == "spearman"
                    else stats.pearsonr(clean[predictor], clean[outcome])
                )
                low, high = cluster_bootstrap_correlation(frame, predictor, outcome, method)
                rows.append(
                    {
                        "outcome": outcome,
                        "predictor": predictor,
                        "method": method,
                        "n": len(clean),
                        "estimate": result.statistic,
                        "ci_low": low,
                        "ci_high": high,
                    }
                )
    return pd.DataFrame(rows)


def weighted_median(values: np.ndarray, weights: np.ndarray) -> float:
    order = np.argsort(values)
    ordered_values = values[order]
    cumulative = np.cumsum(weights[order])
    total = int(cumulative[-1])
    lower_rank, upper_rank = (total - 1) // 2, total // 2
    lower = ordered_values[np.searchsorted(cumulative, lower_rank, side="right")]
    upper = ordered_values[np.searchsorted(cumulative, upper_rank, side="right")]
    return float((lower + upper) / 2)


def cluster_bootstrap_headline(frame: pd.DataFrame, iterations: int = 2000) -> pd.DataFrame:
    clean = frame[
        ["calculator_key", "entropy_reduction_bits", "entropy_removed_percent"]
    ].dropna()
    cluster_codes, clusters = pd.factorize(clean["calculator_key"], sort=False)
    bits = clean["entropy_reduction_bits"].to_numpy()
    percent = clean["entropy_removed_percent"].to_numpy()
    rng = np.random.default_rng(20260811)
    estimates = np.empty((iterations, 5))
    for index in range(iterations):
        sampled = rng.integers(0, len(clusters), len(clusters))
        cluster_weights = np.bincount(sampled, minlength=len(clusters))
        row_weights = cluster_weights[cluster_codes]
        estimates[index] = [
            weighted_median(bits, row_weights),
            weighted_median(percent, row_weights),
            100 * np.average(percent >= 10, weights=row_weights),
            100 * np.average(percent >= 25, weights=row_weights),
            100 * np.average(percent >= 50, weights=row_weights),
        ]
    definitions = [
        ("Median entropy reduction, bits", float(np.median(bits))),
        ("Median proportional uncertainty removed, %", float(np.median(percent))),
        ("Evaluations removing at least 10%, %", float(100 * np.mean(percent >= 10))),
        ("Evaluations removing at least 25%, %", float(100 * np.mean(percent >= 25))),
        ("Evaluations removing at least 50%, %", float(100 * np.mean(percent >= 50))),
    ]
    rows = []
    for column, (measure, point) in enumerate(definitions):
        low, high = np.percentile(estimates[:, column], [2.5, 97.5])
        rows.append({"measure": measure, "estimate": point, "ci_low": low, "ci_high": high})
    return pd.DataFrame(rows)


def cluster_bootstrap_median(
    frame: pd.DataFrame, column: str, iterations: int = 2000
) -> tuple[float, float]:
    clean = frame[["calculator_key", column]].replace([np.inf, -np.inf], np.nan).dropna()
    cluster_codes, clusters = pd.factorize(clean["calculator_key"], sort=False)
    values = clean[column].to_numpy()
    rng = np.random.default_rng(20260811)
    estimates = np.empty(iterations)
    for index in range(iterations):
        sampled = rng.integers(0, len(clusters), len(clusters))
        cluster_weights = np.bincount(sampled, minlength=len(clusters))
        estimates[index] = weighted_median(values, cluster_weights[cluster_codes])
    return tuple(np.percentile(estimates, [2.5, 97.5]))


def purpose_mask(frame: pd.DataFrame, purpose: str) -> pd.Series:
    return frame["catalog_purpose"].fillna("").str.contains(purpose, case=False, regex=False)


def specialty_mask(frame: pd.DataFrame, specialty: str) -> pd.Series:
    return frame["catalog_specialty"].fillna("").map(
        lambda value: specialty in {label.strip() for label in value.split(",")}
    )


def purpose_sample_size_table(primary: pd.DataFrame) -> pd.DataFrame:
    rows = []
    for purpose in ["Diagnosis", "Prognosis", "Rule Out", "Treatment"]:
        data = primary[purpose_mask(primary, purpose)]
        estimate = stats.spearmanr(
            data["reported_sample_size"], data["entropy_removed_percent"]
        ).statistic
        low, high = cluster_bootstrap_correlation(
            data, "reported_sample_size", "entropy_removed_percent", "spearman"
        )
        rows.append(
            {
                "purpose": purpose,
                "evaluations": len(data),
                "calculators": data["calculator_key"].nunique(),
                "rho_sample_size_percent": estimate,
                "ci_low": low,
                "ci_high": high,
            }
        )
    return pd.DataFrame(rows)


def information_at_prevalence(frame: pd.DataFrame, prevalence: float) -> pd.DataFrame:
    sensitivity = frame["sensitivity"].to_numpy()
    specificity = frame["specificity"].to_numpy()
    tp = sensitivity * prevalence
    fn = (1 - sensitivity) * prevalence
    tn = specificity * (1 - prevalence)
    fp = (1 - specificity) * (1 - prevalence)
    positive, negative = tp + fp, fn + tn
    positive_event_probability = np.divide(
        tp, positive, out=np.zeros_like(tp), where=positive > 0
    )
    negative_event_probability = np.divide(
        fn, negative, out=np.zeros_like(fn), where=negative > 0
    )
    baseline = float(binary_entropy(np.array([prevalence]))[0])
    conditional = (
        positive * binary_entropy(positive_event_probability)
        + negative * binary_entropy(negative_event_probability)
    )
    bits = baseline - conditional
    return pd.DataFrame(
        {
            "entropy_reduction_bits": bits,
            "entropy_removed_percent": 100 * bits / baseline,
        },
        index=frame.index,
    )


def prevalence_standardization_table(primary: pd.DataFrame) -> pd.DataFrame:
    rows = []
    for prevalence in [0.05, 0.10, 0.25, 0.50]:
        standardized = information_at_prevalence(primary, prevalence)
        rows.append(
            {
                "fixed_prevalence": prevalence,
                "median_bits": standardized["entropy_reduction_bits"].median(),
                "q1_bits": standardized["entropy_reduction_bits"].quantile(0.25),
                "q3_bits": standardized["entropy_reduction_bits"].quantile(0.75),
                "median_percent": standardized["entropy_removed_percent"].median(),
                "q1_percent": standardized["entropy_removed_percent"].quantile(0.25),
                "q3_percent": standardized["entropy_removed_percent"].quantile(0.75),
                "rho_observed_vs_standardized_percent": stats.spearmanr(
                    primary["entropy_removed_percent"],
                    standardized["entropy_removed_percent"],
                ).statistic,
            }
        )
    return pd.DataFrame(rows)


def specialty_comparison_table(
    primary: pd.DataFrame, minimum_evaluations: int = 50
) -> pd.DataFrame:
    specialties = sorted(
        {
            label.strip()
            for value in primary["catalog_specialty"].dropna()
            for label in value.split(",")
            if label.strip()
        }
    )
    rows = []
    for specialty in specialties:
        data = primary[specialty_mask(primary, specialty)].copy()
        if len(data) < minimum_evaluations:
            continue
        data["fixed_10_percent"] = information_at_prevalence(data, 0.10)[
            "entropy_removed_percent"
        ]
        observed_low, observed_high = cluster_bootstrap_median(
            data, "entropy_removed_percent"
        )
        fixed_low, fixed_high = cluster_bootstrap_median(data, "fixed_10_percent")
        rho = stats.spearmanr(
            data["reported_sample_size"], data["entropy_removed_percent"]
        ).statistic
        rho_low, rho_high = cluster_bootstrap_correlation(
            data, "reported_sample_size", "entropy_removed_percent", "spearman"
        )
        rows.append(
            {
                "specialty": specialty,
                "evaluations": len(data),
                "calculators": data["calculator_key"].nunique(),
                "median_prevalence_percent": 100 * data["prevalence"].median(),
                "median_removed_percent": data["entropy_removed_percent"].median(),
                "removed_ci_low": observed_low,
                "removed_ci_high": observed_high,
                "median_fixed_10_percent": data["fixed_10_percent"].median(),
                "fixed_10_ci_low": fixed_low,
                "fixed_10_ci_high": fixed_high,
                "median_positive_share_percent": data[
                    "positive_information_share_percent"
                ].median(),
                "rho_sample_size_percent": rho,
                "rho_ci_low": rho_low,
                "rho_ci_high": rho_high,
            }
        )
    return pd.DataFrame(rows).sort_values("median_removed_percent", ascending=False)


def catalog_representation_table(catalog: pd.DataFrame, primary: pd.DataFrame) -> pd.DataFrame:
    represented = set(primary["catalog_id"].dropna().astype(int))
    rows = []
    for purpose in ["Diagnosis", "Prognosis", "Rule Out", "Treatment"]:
        eligible = catalog[purpose_mask(catalog, purpose)]
        represented_count = int(eligible["catalog_id"].astype(int).isin(represented).sum())
        rows.append(
            {
                "purpose": purpose,
                "catalog_calculators": len(eligible),
                "represented_calculators": represented_count,
                "representation_percent": 100 * represented_count / len(eligible),
                "primary_evaluations": int(purpose_mask(primary, purpose).sum()),
            }
        )
    return pd.DataFrame(rows)


def pick_one_per_calculator(frame: pd.DataFrame) -> pd.DataFrame:
    priority = {
        "Count-based": 0,
        "Reported sensitivity/specificity": 1,
        "Metric-reconstructed": 2,
        "Legacy/no recorded source": 3,
    }
    ranked = frame.assign(
        evidence_priority=frame["evidence_group"].map(priority).fillna(9),
        reported_n_sort=frame["reported_sample_size"].fillna(-1),
    ).sort_values(["calculator_key", "evidence_priority", "reported_n_sort"], ascending=[True, True, False])
    return ranked.drop_duplicates("calculator_key", keep="first")


def sensitivity_table(primary: pd.DataFrame, expanded: pd.DataFrame) -> pd.DataFrame:
    scenarios = {
        "Primary analysis": primary,
        "Including 7 other tools or strategies": expanded,
        "One evaluation per calculator": pick_one_per_calculator(primary),
        "Whole-number counts only": primary[primary["integer_counts"]],
        "High or moderate-high confidence": primary[
            primary["confidence_group"].isin(["High", "Moderate-high"])
        ],
        "Evaluations based on reported counts": primary[primary["evidence_group"].eq("Count-based")],
        "Excluding explicitly case-control/two-gate sources": primary[
            ~primary["sampling_design"].eq("Case-control/two-gate")
        ],
    }
    rows = []
    for name, data in scenarios.items():
        rho = stats.spearmanr(data["youden_j"], data["entropy_reduction_bits"]).statistic
        rows.append(
            {
                "analysis": name,
                "evaluations": len(data),
                "calculators": data["calculator_key"].nunique(),
                "median_bits": data["entropy_reduction_bits"].median(),
                "q1_bits": data["entropy_reduction_bits"].quantile(0.25),
                "q3_bits": data["entropy_reduction_bits"].quantile(0.75),
                "median_percent": data["entropy_removed_percent"].median(),
                "q1_percent": data["entropy_removed_percent"].quantile(0.25),
                "q3_percent": data["entropy_removed_percent"].quantile(0.75),
                "rho_youden_bits": rho,
            }
        )
    return pd.DataFrame(rows)


def sampling_design_table(primary: pd.DataFrame) -> pd.DataFrame:
    rows = []
    for label in ["Cohort/consecutive", "Case-control/two-gate", "Unclear"]:
        data = primary[primary["sampling_design"].eq(label)]
        rows.append(
            {
                "sampling_design": label,
                "evaluations": len(data),
                "calculators": data["calculator_key"].nunique(),
                "median_bits": data["entropy_reduction_bits"].median(),
                "median_percent": data["entropy_removed_percent"].median(),
            }
        )
    return pd.DataFrame(rows)


def evidence_audit_table(primary: pd.DataFrame) -> pd.DataFrame:
    total = len(primary)
    present = lambda column: primary[column].fillna("").astype(str).str.strip().ne("")
    reported_n = primary["reported_sample_size"].notna()
    contribution_matches = np.isclose(
        primary["positive_information_bits"] + primary["negative_information_bits"],
        primary["entropy_reduction_bits"],
        atol=1e-10,
    )
    rows = [
        ("Count-based source evidence", primary["evidence_group"].eq("Count-based"), total),
        (
            "High or moderate-high confidence label",
            primary["confidence_group"].isin(["High", "Moderate-high"]),
            total,
        ),
        ("Study citation recorded", present("study"), total),
        ("Threshold description recorded", present("cutoff"), total),
        (
            "Complete, non-negative 2x2 cells",
            primary[["tp", "fp", "fn", "tn"]].notna().all(axis=1)
            & primary[["tp", "fp", "fn", "tn"]].ge(0).all(axis=1),
            total,
        ),
        (
            "Reported sample size within one of 2x2 total",
            (primary["reported_sample_size"] - primary["count_total"]).abs().le(1)
            & reported_n,
            int(reported_n.sum()),
        ),
        (
            "Finite information estimates",
            np.isfinite(primary["entropy_reduction_bits"])
            & np.isfinite(primary["entropy_removed_percent"]),
            total,
        ),
        ("Positive and negative contributions reproduce total information", contribution_matches, total),
    ]
    return pd.DataFrame(
        [
            {
                "audit_item": label,
                "evaluations_meeting": int(np.asarray(mask).sum()),
                "evaluations_assessed": assessed,
                "percent": 100 * np.asarray(mask).sum() / assessed,
            }
            for label, mask, assessed in rows
        ]
    )


def sample_size_table(primary: pd.DataFrame) -> pd.DataFrame:
    quartiles = primary.assign(
        sample_size_group=pd.qcut(
            primary["reported_sample_size"],
            4,
            labels=["Q1 (smallest)", "Q2", "Q3", "Q4 (largest)"],
        )
    )
    rows = []
    for label, data in quartiles.groupby("sample_size_group", observed=True):
        rows.append(
            {
                "analysis": f"Sample-size quartile {label}",
                "evaluations": len(data),
                "calculators": data["calculator_key"].nunique(),
                "minimum_n": data["reported_sample_size"].min(),
                "maximum_n": data["reported_sample_size"].max(),
                "median_n": data["reported_sample_size"].median(),
                "median_bits": data["entropy_reduction_bits"].median(),
                "median_percent": data["entropy_removed_percent"].median(),
            }
        )
    for minimum in [100, 200, 500, 1000, 5000]:
        data = primary[primary["reported_sample_size"].ge(minimum)]
        rows.append(
            {
                "analysis": f"Reported sample size >= {minimum}",
                "evaluations": len(data),
                "calculators": data["calculator_key"].nunique(),
                "minimum_n": data["reported_sample_size"].min(),
                "maximum_n": data["reported_sample_size"].max(),
                "median_n": data["reported_sample_size"].median(),
                "median_bits": data["entropy_reduction_bits"].median(),
                "median_percent": data["entropy_removed_percent"].median(),
            }
        )
    return pd.DataFrame(rows)


def study_size_bias_robustness_table(primary: pd.DataFrame) -> pd.DataFrame:
    data = primary.copy()
    data["fixed_10_percent"] = information_at_prevalence(data, 0.10)[
        "entropy_removed_percent"
    ]
    rows = []

    def add_correlation(label: str, outcome: str, subset: pd.DataFrame = data) -> None:
        estimate = stats.spearmanr(
            subset["reported_sample_size"], subset[outcome]
        ).statistic
        low, high = cluster_bootstrap_correlation(
            subset, "reported_sample_size", outcome, "spearman"
        )
        rows.append(
            {
                "analysis": label,
                "evaluations": len(subset),
                "estimate": estimate,
                "ci_low": low,
                "ci_high": high,
            }
        )

    add_correlation("Observed information gain, bits", "entropy_reduction_bits")
    add_correlation(
        "Miller–Madow corrected information gain, bits",
        "entropy_reduction_mm_bits",
    )
    add_correlation(
        "Observed proportional uncertainty removed",
        "entropy_removed_percent",
    )
    add_correlation(
        "Miller–Madow corrected proportional uncertainty removed",
        "entropy_removed_mm_percent",
    )

    estimate = partial_spearman(
        data["reported_sample_size"].to_numpy(),
        data["entropy_removed_mm_percent"].to_numpy(),
        data["parent_entropy_mm_bits"].to_numpy(),
    )
    low, high = cluster_bootstrap_partial_spearman(
        data,
        "reported_sample_size",
        "entropy_removed_mm_percent",
        "parent_entropy_mm_bits",
    )
    rows.append(
        {
            "analysis": "Corrected proportional uncertainty removed, adjusted for baseline uncertainty",
            "evaluations": len(data),
            "estimate": estimate,
            "ci_low": low,
            "ci_high": high,
        }
    )
    add_correlation(
        "Proportional uncertainty removed, standardised to 10% outcome prevalence",
        "fixed_10_percent",
    )

    data["prevalence_quartile"] = pd.qcut(
        data["prevalence"],
        4,
        labels=["Q1 lowest", "Q2", "Q3", "Q4 highest"],
    )
    for label, subset in data.groupby("prevalence_quartile", observed=True):
        add_correlation(
            f"Corrected proportional uncertainty removed within prevalence {label}",
            "entropy_removed_mm_percent",
            subset,
        )
    return pd.DataFrame(rows)


def atlas_numeric_rows(primary: pd.DataFrame, metadata: list[dict[str, object]]) -> pd.DataFrame:
    indexed = primary.set_index("source_index", drop=False)
    rows = []
    for item in metadata:
        source_index = int(item["source_index"])
        if source_index not in indexed.index:
            raise ValueError(f"Clinical atlas source index not found: {source_index}")
        source = indexed.loc[source_index]
        if isinstance(source, pd.DataFrame):
            raise ValueError(f"Clinical atlas source index is not unique: {source_index}")
        rows.append(
            {
                "block": item["block"],
                "row_type": "Observed",
                "source_index": source_index,
                "clinical_case": item["label"],
                "clinical_context": item["context"],
                "n": int(round(source["count_total"])),
                "starting_risk_percent": 100 * source["prevalence"],
                "tp": source["tp"],
                "fp": source["fp"],
                "fn": source["fn"],
                "tn": source["tn"],
                "sensitivity_percent": 100 * source["sensitivity"],
                "specificity_percent": 100 * source["specificity"],
                "risk_after_positive_percent": 100 * source["ppv"],
                "risk_after_negative_percent": 100 * (1 - source["npv"]),
                "lr_positive": source["lr_positive"],
                "lr_negative": source["lr_negative"],
                "youden_j": source["youden_j"],
                "information_gain_bits": source["entropy_reduction_bits"],
                "information_removed_percent": source["entropy_removed_percent"],
                "positive_information_share_percent": source[
                    "positive_information_share_percent"
                ],
                "negative_information_share_percent": source[
                    "negative_information_share_percent"
                ],
                "clinical_reading": item["reading"],
                "reference": item.get("reference", ""),
            }
        )
    return pd.DataFrame(rows)


def illustrative_comparison_rows() -> tuple[pd.DataFrame, pd.DataFrame]:
    counts = pd.DataFrame(
        [
            {"tp": 360, "fp": 240, "fn": 40, "tn": 360},
            {"tp": 36, "fp": 384, "fn": 4, "tn": 576},
            {"tp": 950, "fp": 4050, "fn": 50, "tn": 4950},
            {"tp": 750, "fp": 2250, "fn": 250, "tn": 6750},
            {"tp": 550, "fp": 450, "fn": 450, "tn": 8550},
        ],
        index=["Prevalence A", "Prevalence B", "J rule-out", "J balanced", "J rule-in"],
    )
    metrics = add_metrics(counts)
    metadata = [
        {
            "block": "Prevalence-shift experiment",
            "source_index": "Illustrative A",
            "label": "Same illustrative threshold: 40% prevalence",
            "context": "Per 1000; sensitivity 90% and specificity 60%",
            "reading": "This higher-prevalence population establishes the reference amount and direction of information.",
        },
        {
            "block": "Prevalence-shift experiment",
            "source_index": "Illustrative B",
            "label": "Same illustrative threshold: 4% prevalence",
            "context": "Per 1000; sensitivity 90% and specificity 60%",
            "reading": "Lower starting risk cuts information per patient by 84.8% despite unchanged sensitivity and specificity.",
        },
        {
            "block": "Same-prevalence, same-Youden experiment",
            "source_index": "J rule-out",
            "label": "Rule-out emphasis: 10% prevalence",
            "context": "Per 10,000; sensitivity 95% and specificity 55%",
            "reading": "The same J can concentrate information in negative results.",
        },
        {
            "block": "Same-prevalence, same-Youden experiment",
            "source_index": "J balanced",
            "label": "Balanced accuracy: 10% prevalence",
            "context": "Per 10,000; sensitivity 75% and specificity 75%",
            "reading": "Balancing sensitivity and specificity changes both the amount and direction of information.",
        },
        {
            "block": "Same-prevalence, same-Youden experiment",
            "source_index": "J rule-in",
            "label": "Rule-in emphasis: 10% prevalence",
            "context": "Per 10,000; sensitivity 55% and specificity 95%",
            "reading": "The same J can produce more information concentrated in positive results.",
        },
    ]
    rows = []
    for item, (_, source) in zip(metadata, metrics.iterrows()):
        rows.append(
            {
                "block": item["block"],
                "row_type": "Illustrative",
                "source_index": item["source_index"],
                "clinical_case": item["label"],
                "clinical_context": item["context"],
                "n": int(round(source["count_total"])),
                "starting_risk_percent": 100 * source["prevalence"],
                "tp": source["tp"],
                "fp": source["fp"],
                "fn": source["fn"],
                "tn": source["tn"],
                "sensitivity_percent": 100 * source["sensitivity"],
                "specificity_percent": 100 * source["specificity"],
                "risk_after_positive_percent": 100 * source["ppv"],
                "risk_after_negative_percent": 100 * (1 - source["npv"]),
                "lr_positive": source["lr_positive"],
                "lr_negative": source["lr_negative"],
                "youden_j": source["youden_j"],
                "information_gain_bits": source["entropy_reduction_bits"],
                "information_removed_percent": source["entropy_removed_percent"],
                "positive_information_share_percent": source[
                    "positive_information_share_percent"
                ],
                "negative_information_share_percent": source[
                    "negative_information_share_percent"
                ],
                "clinical_reading": item["reading"],
                "reference": "",
            }
        )
    result = pd.DataFrame(rows).set_index("source_index", drop=False)
    transport = result.loc[["Illustrative A", "Illustrative B"]].reset_index(drop=True)
    youden = result.loc[["J rule-out", "J balanced", "J rule-in"]].reset_index(drop=True)
    invariant = ["sensitivity", "specificity", "lr_positive", "lr_negative", "youden_j"]
    assert np.allclose(metrics.loc["Prevalence A", invariant], metrics.loc["Prevalence B", invariant])
    assert not np.isclose(
        metrics.loc["Prevalence A", "entropy_reduction_bits"],
        metrics.loc["Prevalence B", "entropy_reduction_bits"],
    )
    decline = 100 * (
        1
        - metrics.loc["Prevalence B", "entropy_reduction_bits"]
        / metrics.loc["Prevalence A", "entropy_reduction_bits"]
    )
    assert np.isclose(decline, 84.8, atol=0.1)
    assert np.allclose(metrics.loc[["J rule-out", "J balanced", "J rule-in"], "prevalence"], 0.1)
    assert np.allclose(metrics.loc[["J rule-out", "J balanced", "J rule-in"], "youden_j"], 0.5)
    assert metrics.loc[["J rule-out", "J balanced", "J rule-in"], "entropy_reduction_bits"].nunique() == 3
    assert list(
        metrics.loc[["J rule-out", "J balanced", "J rule-in"], "information_contribution_pattern"]
    ) == [
        "Negative-result dominant",
        "Balanced",
        "Positive-result dominant",
    ]
    return transport, youden


def format_atlas_lr(value: float) -> str:
    if np.isposinf(value):
        return "∞"
    if value == 0:
        return "0"
    return f"{value:.2g}"


def format_atlas_bits(value: float) -> str:
    return f"{value:.4f}" if value < 0.1 else f"{value:.3f}"


def atlas_display_table(frame: pd.DataFrame) -> pd.DataFrame:
    rows = []
    for _, row in frame.iterrows():
        citation = (
            f"[{int(row['reference'])}]"
            if row["reference"] != ""
            else "Constructed example"
            if row["row_type"] == "Illustrative"
            else "See source ledger"
        )
        positive_share = f"T+ {row['positive_information_share_percent']:.1f}%"
        negative_share = f"T− {row['negative_information_share_percent']:.1f}%"
        if row["positive_information_share_percent"] > 60:
            positive_share = f"**{positive_share}**"
        elif row["positive_information_share_percent"] < 40:
            negative_share = f"**{negative_share}**"
        removed = row["information_removed_percent"]
        removed_text = f"{removed:.2f}%" if removed < 10 else f"{removed:.1f}%"
        rows.append(
            {
                "Clinical decision and calculator": (
                    f"*{row['block']}*; **{row['clinical_case']}**; "
                    f"{row['row_type']}; {row['clinical_context']}"
                ),
                "Cohort and starting risk": (
                    f"n={row['n']:,}; Starting risk {row['starting_risk_percent']:.1f}%"
                ),
                "Accuracy (sensitivity; specificity; J; LR+; LR−)": (
                    f"{row['sensitivity_percent']:.1f}%; {row['specificity_percent']:.1f}%; "
                    f"{row['youden_j']:.3f}; {format_atlas_lr(row['lr_positive'])}; "
                    f"{format_atlas_lr(row['lr_negative'])}"
                ),
                "Risk after positive; negative": (
                    f"{row['risk_after_positive_percent']:.1f}% / "
                    f"{row['risk_after_negative_percent']:.1f}%"
                ),
                "Information yield": (
                    f"{format_atlas_bits(row['information_gain_bits'])} bits; "
                    f"{removed_text} uncertainty removed"
                ),
                "Dominant result and information share": f"{positive_share} / {negative_share}",
                "Why it matters clinically": row["clinical_reading"],
                "Source": citation,
            }
        )
    return pd.DataFrame(rows)


def clinical_atlas_source_ledger(
    primary: pd.DataFrame, metadata: list[dict[str, object]]
) -> pd.DataFrame:
    indexed = primary.set_index("source_index", drop=False)
    rows = []
    for item in metadata:
        source_index = int(item["source_index"])
        source = indexed.loc[source_index]
        rows.append(
            {
                "source_index": source_index,
                "clinical_case": item["label"],
                "study_citation": SOURCE_CITATION_OVERRIDES.get(source_index, source["study"]),
                "outcome_and_threshold": item["context"],
                "tp": source["tp"],
                "fp": source["fp"],
                "fn": source["fn"],
                "tn": source["tn"],
                "mdcalc_url": source["catalog_url"],
            }
        )
    return pd.DataFrame(rows)


def clinical_metric_atlas_tables(
    primary: pd.DataFrame,
) -> tuple[pd.DataFrame, pd.DataFrame, pd.DataFrame, pd.DataFrame, pd.DataFrame]:
    main_observed = atlas_numeric_rows(primary, MAIN_ATLAS_ROWS)
    prevalence_experiment, youden_experiment = illustrative_comparison_rows()
    supplemental_metadata = [*MAIN_ATLAS_ROWS, *SUPPLEMENTAL_ATLAS_ROWS]
    supplemental = atlas_numeric_rows(primary, supplemental_metadata)
    ledger = clinical_atlas_source_ledger(primary, supplemental_metadata)

    assert len(main_observed) == 7
    assert main_observed["row_type"].eq("Observed").all()
    assert len(prevalence_experiment) == 2
    assert prevalence_experiment["row_type"].eq("Illustrative").all()
    assert len(youden_experiment) == 3
    assert youden_experiment["row_type"].eq("Illustrative").all()
    assert len(supplemental) == 24
    assert supplemental["source_index"].is_unique
    assert len(ledger) == 24
    assert ledger["source_index"].is_unique
    assert ledger[["study_citation", "mdcalc_url"]].notna().all().all()
    assert np.allclose(
        supplemental["positive_information_share_percent"]
        + supplemental["negative_information_share_percent"],
        100,
    )
    pneumonia = main_observed[main_observed["source_index"].isin([97, 101])]
    heart = main_observed[main_observed["source_index"].isin([473, 474])]
    assert pneumonia["n"].nunique() == pneumonia["starting_risk_percent"].nunique() == 1
    assert heart["n"].nunique() == heart["starting_risk_percent"].nunique() == 1
    return main_observed, prevalence_experiment, youden_experiment, supplemental, ledger


def save_table_markdown(frame: pd.DataFrame, path: Path, digits: int = 3) -> None:
    rounded = frame.round(digits).fillna("")
    header = "| " + " | ".join(map(str, rounded.columns)) + " |"
    divider = "| " + " | ".join(["---"] * len(rounded.columns)) + " |"
    rows = ["| " + " | ".join(map(str, row)) + " |" for row in rounded.itertuples(index=False, name=None)]
    path.write_text("\n".join([header, divider, *rows, ""]), encoding="utf-8")


def configure_figure_style(grayscale: bool) -> tuple[str, ...]:
    plt.style.use("default")
    plt.style.use(str(BMJ_STYLE))
    text_colour = GRAYS[0] if grayscale else INK
    axis_colour = GRAYS[1] if grayscale else MUTED
    plt.rcParams.update(
        {
            "svg.hashsalt": "mdcalc-entropy-bmj-v0.14.0",
            "text.color": text_colour,
            "axes.labelcolor": text_colour,
            "axes.edgecolor": axis_colour,
            "xtick.color": axis_colour,
            "ytick.color": axis_colour,
        }
    )
    return GRAYS if grayscale else COLOURS


def figure_size(height_mm: float) -> tuple[float, float]:
    return DOUBLE_COLUMN_MM / MM_PER_INCH, height_mm / MM_PER_INCH


def save_figure(fig: plt.Figure, stem: Path) -> None:
    fig.savefig(
        stem.with_suffix(".png"),
        format="png",
        dpi=FIGURE_DPI,
        facecolor=WHITE,
        edgecolor="none",
        metadata={"Software": "MDCalc BMJ 0.14.0 deterministic figure builder"},
    )
    fig.savefig(
        stem.with_suffix(".pdf"),
        format="pdf",
        facecolor=WHITE,
        edgecolor="none",
        metadata={
            "Title": stem.stem,
            "Author": "MDCalc BMJ 0.14.0 deterministic figure builder",
            "Creator": "analysis/analyze.py",
            "CreationDate": None,
            "ModDate": None,
        },
    )
    svg = stem.with_suffix(".svg")
    fig.savefig(
        svg,
        format="svg",
        facecolor=WHITE,
        edgecolor="none",
        metadata={"Title": stem.stem, "Creator": "analysis/analyze.py", "Date": "2026-08-29"},
    )
    svg.write_text(
        "\n".join(line.rstrip() for line in svg.read_text(encoding="utf-8").splitlines()) + "\n",
        encoding="utf-8",
        newline="\n",
    )
    plt.close(fig)


def deterministic_offsets(count: int, width: float = 0.28) -> np.ndarray:
    """Return stable low-discrepancy offsets without a random-number generator."""

    return (((np.arange(count, dtype=float) * 0.6180339887498949) % 1.0) - 0.5) * width


def label_panel(ax: plt.Axes, label: str, title: str) -> None:
    ax.set_title(f"{label}  {title}", loc="left", fontsize=8.5, weight="semibold", pad=6)


def strip_interval(
    ax: plt.Axes,
    values: pd.Series | np.ndarray,
    position: float,
    colour: str,
    *,
    marker: str = "o",
    width: float = 0.30,
) -> None:
    plotted = np.sort(np.asarray(values, dtype=float))
    ax.scatter(
        position + deterministic_offsets(len(plotted), width),
        plotted,
        s=7,
        marker=marker,
        color=colour,
        alpha=0.22,
        edgecolors="none",
        zorder=2,
    )
    q1, median, q3 = np.quantile(plotted, [0.25, 0.50, 0.75])
    ax.vlines(position, plotted.min(), plotted.max(), color=MUTED, lw=0.7, zorder=3)
    ax.vlines(position, q1, q3, color=colour, lw=5.0, zorder=4)
    ax.scatter(position, median, s=30, marker="D", color=INK, edgecolor=WHITE, linewidth=0.5, zorder=5)


def make_figures(
    primary: pd.DataFrame,
    flow: dict[str, int],
    catalog_representation: pd.DataFrame,
    specialty_comparison: pd.DataFrame,
    figure_dir: Path,
) -> None:
    figure_dir.mkdir(parents=True, exist_ok=True)
    median_removed = float(primary["entropy_removed_percent"].median())
    values = np.sort(primary["entropy_removed_percent"].dropna().to_numpy())
    contribution_counts = primary["information_contribution_pattern"].value_counts()
    dominant_count = int(
        contribution_counts["Negative-result dominant"]
        + contribution_counts["Positive-result dominant"]
    )
    assert dominant_count == 329
    purposes = ["Diagnosis", "Prognosis", "Rule Out", "Treatment"]
    purpose_groups = [
        primary.loc[purpose_mask(primary, purpose), "positive_information_share_percent"]
        for purpose in purposes
    ]
    prevalence_labels = ["Q1 lowest", "Q2", "Q3", "Q4 highest"]
    prevalence_plot = primary.copy()
    prevalence_plot["prevalence_quartile"] = pd.qcut(
        prevalence_plot["prevalence"], 4, labels=prevalence_labels
    )
    prevalence_groups = [
        prevalence_plot.loc[
            prevalence_plot["prevalence_quartile"].eq(label), "entropy_reduction_bits"
        ]
        for label in prevalence_labels
    ]
    sample_labels = ["Q1 smallest", "Q2", "Q3", "Q4 largest"]
    sample_plot = primary.assign(
        sample_size_quartile=pd.qcut(
            primary["reported_sample_size"], 4, labels=sample_labels
        )
    )
    sample_groups = [
        sample_plot.loc[
            sample_plot["sample_size_quartile"].eq(label), "entropy_removed_percent"
        ]
        for label in sample_labels
    ]
    prevalence_scenarios = [0.05, 0.10, 0.25, 0.50]
    standardized = [information_at_prevalence(primary, prevalence) for prevalence in prevalence_scenarios]
    standardized_10 = standardized[1]["entropy_removed_percent"]

    def save_variants(stem: str, builder: object) -> None:
        for grayscale in (False, True):
            colours = configure_figure_style(grayscale)
            fig = builder(colours, grayscale)
            suffix = "_grayscale" if grayscale else ""
            save_figure(fig, figure_dir / f"{stem}{suffix}")

    def graphical_abstract(colours: tuple[str, ...], grayscale: bool) -> plt.Figure:
        fig, ax = plt.subplots(figsize=figure_size(88), constrained_layout=True)
        ax.set(xlim=(0, 1), ylim=(0, 1))
        ax.axis("off")
        fig.suptitle(
            "What does a clinical calculator result mean?",
            x=0.04,
            y=0.97,
            ha="left",
            fontsize=14,
            weight="semibold",
        )
        ax.vlines([0.31, 0.69], 0.18, 0.84, color=GRID, lw=0.8)

        ax.text(0.03, 0.80, "CATALOGUE EVIDENCE", fontsize=7.5, weight="semibold", color=MUTED)
        ax.text(0.03, 0.61, "482", fontsize=26, weight="bold", color=colours[0])
        ax.text(0.03, 0.49, "binary evaluations", fontsize=9)
        ax.text(0.03, 0.34, "407 calculators represented", fontsize=9)

        ax.text(0.35, 0.80, "HOW MUCH WAS ADDED?", fontsize=7.5, weight="semibold", color=MUTED)
        ax.hlines(0.54, 0.36, 0.65, color=GRID, lw=1.2)
        for value, label, y, dx, alignment, colour, marker in (
            (5.0, "21.2% below 5%", 0.64, -0.008, "right", colours[2], "^"),
            (median_removed, "14.9% median", 0.44, 0.000, "center", colours[3], "D"),
            (50.0, "10.0% at least 50%", 0.64, 0.008, "left", colours[1], "s"),
        ):
            x = 0.36 + 0.29 * value / 100
            ax.scatter(x, 0.54, s=36, marker=marker, color=colour, zorder=3)
            ax.text(x + dx, y, label, ha=alignment, va="center", fontsize=7.2)
        ax.text(0.50, 0.27, "Percentage of starting\nuncertainty removed", ha="center", fontsize=8.3)

        ax.text(0.73, 0.80, "WHICH CLASSIFICATION?", fontsize=7.5, weight="semibold", color=MUTED)
        ax.text(0.73, 0.62, "68.3%", fontsize=24, weight="bold", color=colours[0])
        ax.text(0.73, 0.50, "result dominant\n(329 of 482 evaluations)", fontsize=8.5, linespacing=1.2)
        ax.text(0.73, 0.31, "71.3%", fontsize=17, weight="semibold", color=colours[1])
        ax.text(0.73, 0.21, "median negative share among\nrule-out evaluations", fontsize=7.5, va="center")
        ax.text(0.03, 0.08, "Average information describes learning in an evaluated population; it is not clinical utility.", fontsize=8.5, color=MUTED)
        return fig

    def selection_flow(colours: tuple[str, ...], grayscale: bool) -> plt.Figure:
        fig, axes = plt.subplots(
            1,
            2,
            figsize=figure_size(96),
            gridspec_kw={"width_ratios": [1.35, 1]},
            constrained_layout=True,
        )
        ax = axes[0]
        label_panel(ax, "A", "Evidence selection")
        ax.set(xlim=(0, 1), ylim=(0, 1))
        ax.axis("off")
        stages = (
            (0.84, "494", "binary evaluation records"),
            (0.63, "487", "linked to the MDCalc catalogue"),
            (0.42, "482", "primary evaluations"),
            (0.17, "407", "unique catalogue calculators represented"),
        )
        ax.plot([0.16, 0.16], [0.17, 0.84], color=colours[0], lw=1.5)
        for y, count, label in stages:
            ax.scatter(0.16, y, s=42, color=colours[0], edgecolor=WHITE, linewidth=0.6, zorder=3)
            ax.text(0.23, y + 0.015, count, fontsize=12, weight="semibold", va="center")
            ax.text(0.23, y - 0.045, label, fontsize=8, va="center")
        ax.annotate(
            "7 evaluations of 4 off-catalogue\ntools or strategies; sensitivity analysis only",
            xy=(0.16, 0.70),
            xytext=(0.58, 0.72),
            arrowprops={"arrowstyle": "-", "color": MUTED, "lw": 0.8},
            fontsize=7.5,
            va="center",
        )
        ax.annotate(
            "5 removed: 1 superseded reconstruction\nand 4 duplicate 2×2 tables",
            xy=(0.16, 0.49),
            xytext=(0.58, 0.50),
            arrowprops={"arrowstyle": "-", "color": MUTED, "lw": 0.8},
            fontsize=7.5,
            va="center",
        )
        ax.text(0.23, 0.06, "Repeated studies, populations, outcomes, and thresholds were retained.", fontsize=7.5, color=MUTED)

        ax = axes[1]
        label_panel(ax, "B", "Catalogue coverage")
        represented = 100 * flow["mapped_calculators"] / flow["catalog_calculators"]
        ax.barh([0.72], [represented], color=colours[0], height=0.16)
        ax.barh([0.72], [100 - represented], left=[represented], color=GRID, height=0.16)
        ax.text(represented / 2, 0.72, "407 represented\n48.1%", ha="center", va="center", color=WHITE, fontsize=8, weight="semibold")
        ax.text(represented + (100 - represented) / 2, 0.72, "440 without an included\nbinary evaluation", ha="center", va="center", fontsize=7.5)
        rule_out = catalog_representation.loc[catalog_representation["purpose"].eq("Rule Out")].iloc[0]
        ax.hlines(0.30, 0, 100, color=GRID, lw=1.2)
        ax.scatter(rule_out["representation_percent"], 0.30, s=42, marker="s", color=colours[1], zorder=3)
        ax.text(0, 0.39, "Rule-out calculators", fontsize=8, weight="semibold")
        ax.text(
            rule_out["representation_percent"] - 1,
            0.20,
            f"{int(rule_out['represented_calculators'])} of {int(rule_out['catalog_calculators'])} ({rule_out['representation_percent']:.1f}%)",
            ha="right",
            fontsize=8,
            color=colours[1],
        )
        ax.set(xlim=(0, 100), ylim=(0, 1), xlabel="Catalogue calculators represented (%)")
        ax.set_yticks([])
        return fig

    def information_distribution(colours: tuple[str, ...], grayscale: bool) -> plt.Figure:
        fig, axes = plt.subplots(1, 2, figsize=figure_size(92), constrained_layout=True)
        axes[0].hist(
            primary["entropy_removed_percent"],
            bins=np.arange(0, 100.1, 5),
            color=GRAYS[3] if grayscale else "#9AA8B8",
            edgecolor=WHITE,
            alpha=0.95,
        )
        label_panel(axes[0], "A", "Wide variation across evaluations")
        axes[0].set(xlabel="Starting uncertainty removed (%)", ylabel="Evaluations", xlim=(0, 100))
        axes[1].plot(values, 100 * np.arange(1, len(values) + 1) / len(values), color=colours[0], lw=1.8)
        label_panel(axes[1], "B", "Cumulative distribution")
        axes[1].set(xlabel="Starting uncertainty removed (%)", ylabel="Evaluations at or below value (%)", xlim=(0, 100), ylim=(0, 101))
        for ax in axes:
            for value, colour, linestyle in ((5.0, colours[2], ":"), (median_removed, colours[3], "--"), (50.0, colours[1], "-.")):
                ax.axvline(value, color=colour, ls=linestyle, lw=1.2)
        axes[0].text(4.0, 101, "102 below 5%", ha="left", va="bottom", fontsize=7.5, color=colours[2])
        axes[0].text(median_removed + 1.2, 88, "Median 14.9%", rotation=90, va="top", fontsize=7.5, color=colours[3])
        axes[0].text(51.5, 58, "48 at least 50%", rotation=90, va="top", fontsize=7.5, color=colours[1])
        return fig

    def result_contributions(colours: tuple[str, ...], grayscale: bool) -> plt.Figure:
        fig, axes = plt.subplots(1, 2, figsize=figure_size(94), constrained_layout=True)
        axes[0].hist(
            primary["positive_information_share_percent"],
            bins=np.linspace(0, 100, 21),
            color=GRAYS[3] if grayscale else "#9AA8B8",
            edgecolor=WHITE,
        )
        axes[0].axvline(40, color=MUTED, ls="--", lw=0.9)
        axes[0].axvline(60, color=MUTED, ls="--", lw=0.9)
        label_panel(axes[0], "A", "Result dominance")
        axes[0].set(xlabel="Average information accounted for by positive classification (%)", ylabel="Evaluations", xlim=(0, 100))
        top = axes[0].get_ylim()[1]
        axes[0].text(20, top * 0.94, "Negative dominant\n156", ha="center", va="top", fontsize=7.4, color=colours[1])
        axes[0].text(50, top * 0.94, "Balanced\n153", ha="center", va="top", fontsize=7.4, color=MUTED)
        axes[0].text(80, top * 0.94, "Positive dominant\n173", ha="center", va="top", fontsize=7.4, color=colours[3])
        axes[0].text(50, top * 0.73, "329/482 (68.3%) result dominant", ha="center", fontsize=8, weight="semibold")

        label_panel(axes[1], "B", "Clinical purpose")
        for position, (purpose, data) in enumerate(zip(purposes, purpose_groups), start=1):
            colour = colours[1] if purpose == "Rule Out" else colours[0]
            marker = "s" if purpose == "Rule Out" else "o"
            strip_interval(axes[1], data, position, colour, marker=marker)
        axes[1].axhline(50, color=MUTED, ls="--", lw=0.8)
        axes[1].set_xticks(range(1, 5), [f"{purpose}\n(n={len(data)})" for purpose, data in zip(purposes, purpose_groups)])
        axes[1].set(xlabel="Clinical purpose (labels may overlap)", ylabel="Positive-classification information share (%)", ylim=(0, 100))
        rule_out_median = float(purpose_groups[2].median())
        axes[1].annotate(
            f"28.7% positive\n71.3% negative",
            xy=(3, rule_out_median),
            xytext=(3.38, 17),
            arrowprops={"arrowstyle": "-", "color": colours[1], "lw": 0.8},
            fontsize=7.5,
            color=colours[1],
        )
        return fig

    def starting_risk(colours: tuple[str, ...], grayscale: bool) -> plt.Figure:
        fig, axes = plt.subplots(1, 2, figsize=figure_size(94), constrained_layout=True)
        label_panel(axes[0], "A", "Starting risk")
        for position, group in enumerate(prevalence_groups, start=1):
            strip_interval(axes[0], group, position, colours[0])
            axes[0].text(position, float(group.median()) + 0.025, f"{group.median():.3f}", ha="center", fontsize=7.2, color=colours[0])
        axes[0].set_xticks(range(1, 5), ["Q1", "Q2", "Q3", "Q4"])
        axes[0].set(xlabel="Outcome-frequency quartile", ylabel="Information gained (bits)")
        cmap = "Greys" if grayscale else "viridis"
        scatter = axes[1].scatter(
            primary["parent_entropy_bits"],
            primary["entropy_reduction_bits"],
            c=primary["youden_j"],
            cmap=cmap,
            s=14,
            alpha=0.58,
            edgecolors="none",
        )
        label_panel(axes[1], "B", "Available uncertainty")
        axes[1].set(xlabel="Starting uncertainty (bits)", ylabel="Information gained (bits)")
        fig.colorbar(scatter, ax=axes[1], label="Youden's J", shrink=0.88)
        return fig

    def study_size(colours: tuple[str, ...], grayscale: bool) -> plt.Figure:
        fig, axes = plt.subplots(1, 2, figsize=figure_size(94), constrained_layout=True)
        label_panel(axes[0], "A", "Reduction was lower in larger reported studies")
        for position, group in enumerate(sample_groups, start=1):
            strip_interval(axes[0], group, position, colours[0])
            axes[0].text(position, float(group.median()) + 3.2, f"{group.median():.1f}%", ha="center", fontsize=7.3, color=colours[0])
        axes[0].set_xticks(range(1, 5), ["Q1", "Q2", "Q3", "Q4"])
        axes[0].set(xlabel="Reported study-size quartile", ylabel="Starting uncertainty removed (%)", ylim=(0, 104))
        axes[1].scatter(
            primary["reported_sample_size"],
            primary["entropy_removed_percent"],
            color=colours[0],
            s=13,
            alpha=0.28,
            edgecolors="none",
        )
        axes[1].set_xscale("log")
        label_panel(axes[1], "B", "Evaluation-level association")
        axes[1].set(xlabel="Reported sample size (log scale)", ylabel="Starting uncertainty removed (%)")
        rho = float(stats.spearmanr(primary["reported_sample_size"], primary["entropy_removed_percent"]).statistic)
        axes[1].text(0.03, 0.95, f"Spearman ρ = {rho:.3f}", transform=axes[1].transAxes, va="top", fontsize=7.5)
        outlier = primary.loc[primary["reported_sample_size"].idxmax()]
        axes[1].annotate(
            "4.83 million participants",
            (outlier["reported_sample_size"], outlier["entropy_removed_percent"]),
            xytext=(-100, 18),
            textcoords="offset points",
            arrowprops={"arrowstyle": "-", "color": colours[3], "lw": 0.8},
            color=colours[3],
            fontsize=7.3,
        )
        return fig

    def youden_relationship(colours: tuple[str, ...], grayscale: bool) -> plt.Figure:
        fig, axes = plt.subplots(1, 2, figsize=figure_size(94), constrained_layout=True)
        cmap = "Greys" if grayscale else "viridis"
        scatter = axes[0].scatter(
            primary["youden_j"],
            primary["entropy_reduction_bits"],
            c=100 * primary["prevalence"],
            cmap=cmap,
            s=14,
            alpha=0.58,
            edgecolors="none",
        )
        label_panel(axes[0], "A", "Information gain and Youden's J")
        axes[0].set(xlabel="Youden's J", ylabel="Information gained (bits)")
        fig.colorbar(scatter, ax=axes[0], label="Outcome frequency (%)", shrink=0.88)
        axes[1].scatter(
            primary["youden_j"],
            primary["entropy_removed_percent"],
            color=colours[0],
            s=14,
            alpha=0.34,
            edgecolors="none",
        )
        label_panel(axes[1], "B", "Percentage reduction and Youden's J")
        axes[1].set(xlabel="Youden's J", ylabel="Starting uncertainty removed (%)")
        return fig

    def prevalence_standardization(colours: tuple[str, ...], grayscale: bool) -> plt.Figure:
        fig, axes = plt.subplots(1, 2, figsize=figure_size(94), constrained_layout=True)
        label_panel(axes[0], "A", "Information yield at fixed outcome frequencies")
        for position, data in enumerate(standardized, start=1):
            strip_interval(axes[0], data["entropy_removed_percent"], position, colours[2], marker="^")
        axes[0].set_xticks(range(1, 5), ["5%", "10%", "25%", "50%"])
        axes[0].set(xlabel="Fixed outcome frequency", ylabel="Starting uncertainty removed (%)", ylim=(0, 104))
        axes[1].scatter(
            primary["entropy_removed_percent"],
            standardized_10,
            color=colours[0],
            s=13,
            alpha=0.30,
            edgecolors="none",
        )
        axes[1].plot([0, 100], [0, 100], color=MUTED, ls="--", lw=0.8)
        rho = float(stats.spearmanr(primary["entropy_removed_percent"], standardized_10).statistic)
        label_panel(axes[1], "B", "Observed versus standardised ranking")
        axes[1].set(xlabel="Observed uncertainty removed (%)", ylabel="Uncertainty removed at 10% frequency (%)", xlim=(0, 100), ylim=(0, 100))
        axes[1].text(0.04, 0.94, f"Spearman ρ = {rho:.3f}", transform=axes[1].transAxes, va="top", fontsize=7.5)
        return fig

    def catalogue_purpose(colours: tuple[str, ...], grayscale: bool) -> plt.Figure:
        fig, ax = plt.subplots(figsize=figure_size(84), constrained_layout=True)
        bars = ax.barh(
            catalog_representation["purpose"],
            catalog_representation["representation_percent"],
            color=colours[0],
            alpha=0.78,
        )
        for bar, row in zip(bars, catalog_representation.itertuples()):
            ax.text(
                min(bar.get_width() + 1.5, 99),
                bar.get_y() + bar.get_height() / 2,
                f"{row.represented_calculators}/{row.catalog_calculators} ({row.representation_percent:.1f}%)",
                va="center",
                fontsize=7.5,
            )
        ax.set(
            xlabel="Catalogue calculators represented by included binary evaluations (%)",
            ylabel="Nonexclusive clinical-purpose label",
            title="Binary-evaluation coverage differed by clinical purpose",
            xlim=(0, 106),
        )
        return fig

    def specialty_yield(colours: tuple[str, ...], grayscale: bool) -> plt.Figure:
        plotted = specialty_comparison.sort_values("median_removed_percent")
        positions = np.arange(len(plotted))
        fig, ax = plt.subplots(figsize=figure_size(132), constrained_layout=True)
        ax.errorbar(
            plotted["median_removed_percent"],
            positions + 0.12,
            xerr=np.vstack(
                [
                    plotted["median_removed_percent"] - plotted["removed_ci_low"],
                    plotted["removed_ci_high"] - plotted["median_removed_percent"],
                ]
            ),
            fmt="o",
            color=colours[0],
            capsize=2,
            label="Observed outcome frequency",
        )
        ax.errorbar(
            plotted["median_fixed_10_percent"],
            positions - 0.12,
            xerr=np.vstack(
                [
                    plotted["median_fixed_10_percent"] - plotted["fixed_10_ci_low"],
                    plotted["fixed_10_ci_high"] - plotted["median_fixed_10_percent"],
                ]
            ),
            fmt="s",
            color=colours[2],
            capsize=2,
            label="All outcomes set to 10%",
        )
        ax.set_yticks(positions, plotted["specialty"])
        ax.set(
            xlabel="Median starting uncertainty removed (%)",
            ylabel="Clinical specialty (labels may overlap)",
            title="Information yield across commonly represented specialties",
        )
        ax.legend(loc="upper left")
        ax.grid(axis="x", color=GRID, lw=0.5)
        return fig

    save_variants("graphical_abstract", graphical_abstract)
    save_variants("figure1_selection_flow", selection_flow)
    save_variants("figure2_information_yield_distribution", information_distribution)
    save_variants("figure3_result_information_contributions", result_contributions)
    save_variants("figure4_prevalence_information_yield", starting_risk)
    save_variants("figureS1_sample_size_information_yield", study_size)
    save_variants("figureS2_youden_information_relationship", youden_relationship)
    save_variants("figureS3_prevalence_standardization", prevalence_standardization)
    save_variants("figureS4_catalog_representation_by_purpose", catalogue_purpose)
    save_variants("figureS5_specialty_information_yield", specialty_yield)


def main(source: Path = SOURCE) -> None:
    if not source.exists():
        raise FileNotFoundError(source)
    source_hash = sha256(source)
    if source_hash != EXPECTED_SHA256:
        raise ValueError(f"Unexpected workbook SHA-256: {source_hash}")

    data_dir, table_dir, figure_dir = HERE / "data", HERE / "tables", HERE / "figures"
    for directory in [data_dir, table_dir, figure_dir]:
        directory.mkdir(parents=True, exist_ok=True)

    complete_raw = pd.read_excel(source, sheet_name="Complete (494)").dropna(how="all")
    blank_name_rows = complete_raw[complete_raw["Calculator"].isna()].copy()
    complete = complete_raw[complete_raw["Calculator"].notna()].copy().reset_index(names="source_index")
    complete["source_excel_row"] = complete["source_index"] + 2

    catalog_raw = pd.read_excel(source, sheet_name="Full Dataset").dropna(how="all")
    catalog = catalog_raw.rename(
        columns={
            "ID": "catalog_id",
            "Calculator": "catalog_name",
            "URL": "catalog_url",
            "Purpose": "catalog_purpose",
            "Specialty": "catalog_specialty",
            "Disease": "catalog_disease",
        }
    )[["catalog_id", "catalog_name", "catalog_url", "catalog_purpose", "catalog_specialty", "catalog_disease"]]
    catalog["match_name"] = catalog["catalog_name"].map(normalize)

    rename = {
        "Calculator": "source_label",
        "Specialty": "source_specialty",
        "Disease": "source_disease",
        "Sample Size": "reported_sample_size",
        "TP": "tp",
        "FP": "fp",
        "FN": "fn",
        "TN": "tn",
        "Data Source": "data_source",
        "Confidence": "confidence",
        "Study": "study",
        "Cutoff": "cutoff",  # Legacy source-workbook column; manuscript wording uses "threshold".
    }
    evaluations = complete.rename(columns=rename)
    for column in ["reported_sample_size", "tp", "fp", "fn", "tn"]:
        evaluations[column] = pd.to_numeric(evaluations[column], errors="coerce")
    if evaluations[["tp", "fp", "fn", "tn"]].isna().any().any():
        raise ValueError("Named complete rows must contain TP, FP, FN, and TN")

    evaluations["source_correction"] = ""
    priest = evaluations["source_label"].eq("PRIEST COVID-19 Clinical Severity Score")
    if priest.sum() != 1 or evaluations.loc[priest, "tp"].iloc[0] != 4:
        raise ValueError("Expected the single PRIEST source typo (TP=4)")
    evaluations.loc[priest, "tp"] = 44
    evaluations.loc[priest, "source_correction"] = (
        "TP corrected from 4 to 44 to match reported sensitivity 83%, workbook N=177, and the remaining "
        "2x2 cells; source full cohort N=178, leaving a one-participant unresolved discrepancy; "
        "Paraskevas et al. 2022, doi:10.2478/rjim-2022-0015"
    )

    perc = evaluations["source_label"].eq("PERC Rule for Pulmonary Embolism")
    if perc.sum() != 1:
        raise ValueError("Expected one PERC validation record")
    evaluations.loc[perc, "source_disease"] = "Venous thromboembolism or death within 45 days"
    evaluations.loc[perc, "cutoff"] = "Low clinical suspicion (<15%) and all eight PERC criteria negative"

    matches = [match_catalog(label, catalog) for label in evaluations["source_label"]]
    evaluations[["catalog_id", "canonical_name", "match_method"]] = pd.DataFrame(matches, index=evaluations.index)
    catalog_lookup = catalog.set_index("catalog_id")
    for target, source_column in [
        ("catalog_url", "catalog_url"),
        ("catalog_purpose", "catalog_purpose"),
        ("catalog_specialty", "catalog_specialty"),
        ("catalog_disease", "catalog_disease"),
    ]:
        evaluations[target] = evaluations["catalog_id"].map(catalog_lookup[source_column])
    evaluations["calculator_key"] = evaluations.apply(
        lambda row: f"mdcalc:{int(row.catalog_id)}" if pd.notna(row.catalog_id) else f"external:{normalize(row.canonical_name)}",
        axis=1,
    )
    evaluations["confidence_group"] = evaluations["confidence"].map(confidence_group)
    evaluations["evidence_group"] = evaluations["data_source"].map(evidence_group)
    evaluations["sampling_design"] = evaluations["study"].map(sampling_design_group)
    evaluations["integer_counts"] = evaluations[["tp", "fp", "fn", "tn"]].apply(
        lambda row: bool(np.all(np.isclose(row, np.round(row)))), axis=1
    )
    evaluations = add_metrics(evaluations)

    evaluations["included"] = True
    evaluations["exclusion_reason"] = ""
    superseded = evaluations["source_label"].eq("PLASMIC Score for TTP") & evaluations["data_source"].eq("v1_raw")
    evaluations.loc[superseded, ["included", "exclusion_reason"]] = [False, "Superseded scaled-count extraction"]

    duplicate_columns = ["calculator_key", "tp", "fp", "fn", "tn"]
    duplicate_rows = evaluations[evaluations.duplicated(duplicate_columns, keep=False)]
    source_priority = {
        "v2_llm_2x2_raw": 0,
        "v2_llm_event_counts": 1,
        "v2_reported_event_counts": 1,
        "v1_raw": 2,
    }
    for _, group in duplicate_rows.groupby(duplicate_columns, dropna=False):
        ranked = group.assign(
            priority=group["data_source"].map(source_priority).fillna(9),
            metadata=group[["source_specialty", "source_disease", "study", "cutoff"]].notna().sum(axis=1),
        ).sort_values(["priority", "metadata", "source_excel_row"], ascending=[True, False, False])
        evaluations.loc[ranked.index[1:], ["included", "exclusion_reason"]] = [False, "Duplicate 2x2 evaluation"]
    included = evaluations[evaluations["included"]].copy()
    primary = included[included["catalog_id"].notna()].copy()
    expanded = included.copy()

    evaluations["reported_n_delta"] = evaluations["reported_sample_size"] - evaluations["count_total"]
    evaluations["reported_n_discrepancy"] = evaluations["reported_n_delta"].abs().gt(1)
    evaluations["original_entropy_error"] = complete["Entropy Removal"].astype(str).str.startswith("#").values
    evaluations["original_lr_error"] = complete["LR+"].astype(str).str.startswith("#").values

    primary.to_csv(data_dir / "mdcalc_evaluations_primary.csv", index=False)
    expanded.to_csv(data_dir / "mdcalc_evaluations_expanded.csv", index=False)
    evaluations.to_csv(data_dir / "mdcalc_evaluation_audit.csv", index=False)
    catalog.to_csv(data_dir / "mdcalc_catalog.csv", index=False)

    flow = {
        "catalog_calculators": int(catalog["catalog_id"].nunique()),
        "complete_rows": int(len(complete)),
        "blank_formula_rows": int(len(blank_name_rows)),
        "mapped_rows": int(evaluations["catalog_id"].notna().sum()),
        "off_catalog_rows": int(evaluations["catalog_id"].isna().sum()),
        "off_catalog_calculators": int(
            evaluations.loc[evaluations["catalog_id"].isna(), "calculator_key"].nunique()
        ),
        "excluded_rows": int((~evaluations["included"]).sum()),
        "primary_evaluations": int(len(primary)),
        "expanded_evaluations": int(len(expanded)),
        "mapped_calculators": int(primary["catalog_id"].nunique()),
    }

    metric_names = [
        "reported_sample_size",
        "sensitivity",
        "specificity",
        "prevalence",
        "youden_j",
        "entropy_removed_percent",
        "parent_entropy_bits",
        "entropy_reduction_bits",
    ]
    descriptions = pd.DataFrame([{"measure": name, **describe(primary[name])} for name in metric_names])
    correlations = correlation_table(primary)
    sensitivities = sensitivity_table(primary, expanded)
    evidence_audit = evidence_audit_table(primary)
    sampling_design = sampling_design_table(primary)
    sample_sizes = sample_size_table(primary)
    headline_uncertainty = cluster_bootstrap_headline(primary)
    rule_out_information = primary.loc[
        purpose_mask(primary, "Rule Out"),
        ["calculator_key", "positive_information_share_percent"],
    ].copy()
    rule_out_information["negative_information_share_percent"] = (
        100 - rule_out_information["positive_information_share_percent"]
    )
    rule_out_ci = cluster_bootstrap_median(
        rule_out_information, "negative_information_share_percent"
    )
    headline_uncertainty = pd.concat(
        [
            headline_uncertainty,
            pd.DataFrame(
                [
                    {
                        "measure": "Median negative-result information share among rule-out evaluations, %",
                        "estimate": rule_out_information[
                            "negative_information_share_percent"
                        ].median(),
                        "ci_low": rule_out_ci[0],
                        "ci_high": rule_out_ci[1],
                    }
                ]
            ),
        ],
        ignore_index=True,
    )
    purpose_sample_sizes = purpose_sample_size_table(primary)
    prevalence_standardization = prevalence_standardization_table(primary)
    study_size_bias_robustness = study_size_bias_robustness_table(primary)
    specialty_comparison = specialty_comparison_table(primary)
    catalog_representation = catalog_representation_table(catalog, primary)
    clinical_atlas, prevalence_experiment, youden_experiment, supplemental_atlas, clinical_atlas_ledger = (
        clinical_metric_atlas_tables(primary)
    )
    descriptions.to_csv(table_dir / "table1_descriptive_statistics.csv", index=False)
    correlations.to_csv(table_dir / "table2_correlations.csv", index=False)
    sensitivities.to_csv(table_dir / "table3_sensitivity_analyses.csv", index=False)
    evidence_audit.to_csv(table_dir / "tableS2_evidence_audit.csv", index=False)
    sampling_design.to_csv(table_dir / "tableS14_sampling_design_sensitivity.csv", index=False)
    sample_sizes.to_csv(table_dir / "table4_sample_size_analyses.csv", index=False)
    headline_uncertainty.to_csv(table_dir / "tableS10_headline_cluster_bootstrap.csv", index=False)
    purpose_sample_sizes.to_csv(table_dir / "table5_purpose_sample_size_correlations.csv", index=False)
    prevalence_standardization.to_csv(table_dir / "tableS8_prevalence_standardization.csv", index=False)
    study_size_bias_robustness.to_csv(
        table_dir / "tableS13_study_size_bias_robustness.csv", index=False
    )
    catalog_representation.to_csv(table_dir / "tableS9_catalog_representation.csv", index=False)
    specialty_comparison.to_csv(table_dir / "tableS11_specialty_comparison.csv", index=False)
    clinical_atlas.to_csv(table_dir / "table2_clinical_metric_atlas.csv", index=False)
    prevalence_experiment.to_csv(
        table_dir / "table3_prevalence_shift_experiment.csv", index=False
    )
    youden_experiment.to_csv(table_dir / "table4_same_youden_experiment.csv", index=False)
    supplemental_atlas.to_csv(table_dir / "tableS12_clinical_metric_atlas.csv", index=False)
    clinical_atlas_ledger.to_csv(
        table_dir / "tableS12_clinical_metric_atlas_source_ledger.csv", index=False
    )
    save_table_markdown(descriptions, table_dir / "table1_descriptive_statistics.md")
    save_table_markdown(correlations, table_dir / "table2_correlations.md")
    save_table_markdown(sensitivities, table_dir / "table3_sensitivity_analyses.md")
    save_table_markdown(evidence_audit, table_dir / "tableS2_evidence_audit.md")
    save_table_markdown(
        sampling_design,
        table_dir / "tableS14_sampling_design_sensitivity.md",
    )
    save_table_markdown(sample_sizes, table_dir / "table4_sample_size_analyses.md")
    save_table_markdown(headline_uncertainty, table_dir / "tableS10_headline_cluster_bootstrap.md")
    save_table_markdown(purpose_sample_sizes, table_dir / "table5_purpose_sample_size_correlations.md")
    save_table_markdown(prevalence_standardization, table_dir / "tableS8_prevalence_standardization.md")
    save_table_markdown(
        study_size_bias_robustness,
        table_dir / "tableS13_study_size_bias_robustness.md",
    )
    save_table_markdown(catalog_representation, table_dir / "tableS9_catalog_representation.md")
    save_table_markdown(
        specialty_comparison,
        table_dir / "tableS11_specialty_comparison.md",
    )
    save_table_markdown(
        atlas_display_table(clinical_atlas),
        table_dir / "table2_clinical_metric_atlas.md",
    )
    save_table_markdown(
        atlas_display_table(prevalence_experiment),
        table_dir / "table3_prevalence_shift_experiment.md",
    )
    save_table_markdown(
        atlas_display_table(youden_experiment),
        table_dir / "table4_same_youden_experiment.md",
    )
    save_table_markdown(
        atlas_display_table(supplemental_atlas),
        table_dir / "tableS12_clinical_metric_atlas.md",
    )
    save_table_markdown(
        clinical_atlas_ledger,
        table_dir / "tableS12_clinical_metric_atlas_source_ledger.md",
    )
    shapiro_bits = stats.shapiro(primary["entropy_reduction_bits"])
    shapiro_percent = stats.shapiro(primary["entropy_removed_percent"])
    summary = {
        "version": "BMJ 0.14.0",
        "source_sha256": source_hash,
        "flow": flow,
        "descriptive": {name: describe(primary[name]) for name in metric_names},
        "thresholds": {
            "percent_ge_10": int(primary["entropy_removed_percent"].ge(10).sum()),
            "percent_ge_25": int(primary["entropy_removed_percent"].ge(25).sum()),
            "percent_ge_50": int(primary["entropy_removed_percent"].ge(50).sum()),
            "percent_lt_5": int(primary["entropy_removed_percent"].lt(5).sum()),
        },
        "assumptions": {
            "entropy_bits_shapiro_w": shapiro_bits.statistic,
            "entropy_bits_shapiro_p": shapiro_bits.pvalue,
            "entropy_percent_shapiro_w": shapiro_percent.statistic,
            "entropy_percent_shapiro_p": shapiro_percent.pvalue,
            "primary_correlation": "Spearman for monotonic associations with less sensitivity to extreme values; Pearson secondary",
        },
        "sample_size": {
            "rho_bits": stats.spearmanr(
                primary["reported_sample_size"], primary["entropy_reduction_bits"]
            ).statistic,
            "rho_percent": stats.spearmanr(
                primary["reported_sample_size"], primary["entropy_removed_percent"]
            ).statistic,
            "smallest_quartile_median_percent": sample_sizes.iloc[0]["median_percent"],
            "largest_quartile_median_percent": sample_sizes.iloc[3]["median_percent"],
            "median_mm_bits": float(primary["entropy_reduction_mm_bits"].median()),
            "median_mm_percent": float(primary["entropy_removed_mm_percent"].median()),
            "bias_robustness": study_size_bias_robustness.to_dict(orient="records"),
        },
        "information_contributions": {
            "median_positive_bits": float(primary["positive_information_bits"].median()),
            "median_negative_bits": float(primary["negative_information_bits"].median()),
            "median_positive_share_percent": float(
                primary["positive_information_share_percent"].median()
            ),
            "q1_positive_share_percent": float(
                primary["positive_information_share_percent"].quantile(0.25)
            ),
            "q3_positive_share_percent": float(
                primary["positive_information_share_percent"].quantile(0.75)
            ),
            "positive_result_dominant": int(
                primary["information_contribution_pattern"].eq("Positive-result dominant").sum()
            ),
            "balanced": int(primary["information_contribution_pattern"].eq("Balanced").sum()),
            "negative_result_dominant": int(
                primary["information_contribution_pattern"].eq("Negative-result dominant").sum()
            ),
            "rule_out_median_positive_share_percent": float(
                primary.loc[
                    purpose_mask(primary, "Rule Out"), "positive_information_share_percent"
                ].median()
            ),
            "rule_out_evaluations": int(len(rule_out_information)),
            "rule_out_median_negative_share_percent": float(
                rule_out_information["negative_information_share_percent"].median()
            ),
            "rule_out_median_negative_share_ci_low": float(rule_out_ci[0]),
            "rule_out_median_negative_share_ci_high": float(rule_out_ci[1]),
        },
        "headline_uncertainty": headline_uncertainty.to_dict(orient="records"),
        "purpose_sample_size": purpose_sample_sizes.to_dict(orient="records"),
        "prevalence_standardization": prevalence_standardization.to_dict(orient="records"),
        "sampling_design": sampling_design.to_dict(orient="records"),
        "catalog_representation": catalog_representation.to_dict(orient="records"),
        "specialty_comparison": specialty_comparison.to_dict(orient="records"),
        "clinical_atlas": {
            "main_rows": int(len(clinical_atlas)),
            "main_observed_rows": int(clinical_atlas["row_type"].eq("Observed").sum()),
            "prevalence_experiment_rows": int(len(prevalence_experiment)),
            "same_youden_experiment_rows": int(len(youden_experiment)),
            "supplemental_empirical_rows": int(len(supplemental_atlas)),
        },
        "audit": {
            "reported_n_discrepancies": int(evaluations["reported_n_discrepancy"].sum()),
            "source_count_corrections": int(evaluations["source_correction"].ne("").sum()),
            "original_entropy_formula_errors": int(evaluations["original_entropy_error"].sum()),
            "original_lr_formula_errors": int(evaluations["original_lr_error"].sum()),
            "unmatched_rows": int(evaluations["match_method"].eq("Unmatched").sum()),
            "missing_study_citations_primary": int(primary["study"].isna().sum()),
            "missing_thresholds_primary": int(primary["cutoff"].isna().sum()),
        },
    }
    (HERE / "results_summary.json").write_text(json.dumps(summary, indent=2), encoding="utf-8")
    (HERE / "results_summary.md").write_text(
        "\n".join(
            [
                "# MDCalc Entropy Analysis Results",
                "",
                f"Source workbook SHA-256: `{source_hash}`",
                "",
                f"The primary analysis contains {flow['primary_evaluations']} evaluations representing {flow['mapped_calculators']} MDCalc calculators from a catalogue of {flow['catalog_calculators']}.",
                f"Median entropy reduction was {summary['descriptive']['entropy_reduction_bits']['median']:.3f} bits (IQR {summary['descriptive']['entropy_reduction_bits']['q1']:.3f}-{summary['descriptive']['entropy_reduction_bits']['q3']:.3f}).",
                f"Median proportional uncertainty removed was {summary['descriptive']['entropy_removed_percent']['median']:.1f}% (IQR {summary['descriptive']['entropy_removed_percent']['q1']:.1f}%-{summary['descriptive']['entropy_removed_percent']['q3']:.1f}%).",
                f"The 95% confidence interval for median proportional uncertainty removed was {headline_uncertainty.iloc[1]['ci_low']:.1f}%-{headline_uncertainty.iloc[1]['ci_high']:.1f}%, estimated by resampling calculators and keeping repeated evaluations together.",
                f"One classification supplied more than 60% of average information in {summary['information_contributions']['positive_result_dominant'] + summary['information_contributions']['negative_result_dominant']} evaluations; {summary['information_contributions']['positive_result_dominant']} were positive-result dominant, {summary['information_contributions']['negative_result_dominant']} negative-result dominant, and {summary['information_contributions']['balanced']} balanced.",
                f"Among {summary['information_contributions']['rule_out_evaluations']} rule-out evaluations, the median negative-result information share was {summary['information_contributions']['rule_out_median_negative_share_percent']:.1f}% (95% confidence interval {summary['information_contributions']['rule_out_median_negative_share_ci_low']:.1f}%-{summary['information_contributions']['rule_out_median_negative_share_ci_high']:.1f}%).",
                f"Median proportional uncertainty removed declined from {summary['sample_size']['smallest_quartile_median_percent']:.1f}% in the smallest-study quartile to {summary['sample_size']['largest_quartile_median_percent']:.1f}% in the largest-study quartile.",
                f"After Miller–Madow correction, median information gain was {summary['sample_size']['median_mm_bits']:.3f} bits and median proportional uncertainty removed was {summary['sample_size']['median_mm_percent']:.1f}%.",
                f"The corrected study-size correlation was {study_size_bias_robustness.iloc[3]['estimate']:.3f}; after adjustment for baseline uncertainty it was {study_size_bias_robustness.iloc[4]['estimate']:.3f}, and at fixed 10% prevalence it was {study_size_bias_robustness.iloc[5]['estimate']:.3f}.",
                f"Across {len(specialty_comparison)} specialty labels represented by at least 50 evaluations, median proportional uncertainty removed ranged from {specialty_comparison['median_removed_percent'].min():.1f}% to {specialty_comparison['median_removed_percent'].max():.1f}%.",
                f"Excluding {int(primary['sampling_design'].eq('Case-control/two-gate').sum())} evaluations explicitly described as case-control or two-gate left a median information gain of {sensitivities.iloc[-1]['median_bits']:.3f} bits and {sensitivities.iloc[-1]['median_percent']:.1f}% of baseline uncertainty removed.",
                "",
                "One source count was corrected with a DOI-linked audit note; no included record retained a sample-size discrepancy greater than 1.",
                "Spearman correlations are primary for monotonic associations with less sensitivity to extreme values; Pearson correlations are secondary.",
                "Conventional P values are omitted because the study is descriptive; confidence intervals resample calculators and keep repeated evaluations together.",
                "",
            ]
        ),
        encoding="utf-8",
    )
    make_figures(primary, flow, catalog_representation, specialty_comparison, figure_dir)

    expected_figure_stems = (
        "graphical_abstract",
        "figure1_selection_flow",
        "figure2_information_yield_distribution",
        "figure3_result_information_contributions",
        "figure4_prevalence_information_yield",
        "figureS1_sample_size_information_yield",
        "figureS2_youden_information_relationship",
        "figureS3_prevalence_standardization",
        "figureS4_catalog_representation_by_purpose",
        "figureS5_specialty_information_yield",
    )
    assert all(
        (figure_dir / f"{stem}{suffix}.{extension}").is_file()
        for stem in expected_figure_stems
        for suffix in ("", "_grayscale")
        for extension in ("pdf", "svg", "png")
    )

    assert len(complete) == 494
    assert flow["catalog_calculators"] == 847
    assert flow["off_catalog_calculators"] == 4
    assert primary["evidence_group"].value_counts().to_dict() == {
        "Count-based": 331,
        "Legacy/no recorded source": 68,
        "Reported sensitivity/specificity": 64,
        "Metric-reconstructed": 19,
    }
    assert primary["confidence_group"].isin(["High", "Moderate-high"]).sum() == 187
    assert primary["entropy_reduction_bits"].notna().all()
    assert primary["entropy_removed_percent"].between(0, 100 + 1e-9).all()
    assert primary["entropy_reduction_bits"].ge(-1e-10).all()
    assert np.allclose(
        primary["positive_information_bits"] + primary["negative_information_bits"],
        primary["entropy_reduction_bits"],
        atol=1e-10,
    )
    assert primary["positive_information_share_percent"].between(0, 100).all()
    assert primary["information_contribution_pattern"].value_counts().sum() == len(primary)
    assert primary["information_contribution_pattern"].ne("Balanced").sum() == 329
    assert weighted_median(np.array([1.0, 2.0]), np.array([1, 1])) == 1.5
    assert weighted_median(np.array([1.0, 2.0]), np.array([2, 1])) == 1.0
    assert len(headline_uncertainty) == 6
    assert len(rule_out_information) == 56
    assert rule_out_ci[0] <= rule_out_information["negative_information_share_percent"].median() <= rule_out_ci[1]
    assert len(purpose_sample_sizes) == 4
    assert len(prevalence_standardization) == 4
    assert len(study_size_bias_robustness) == 10
    assert study_size_bias_robustness.iloc[:6]["estimate"].lt(0).all()
    assert len(catalog_representation) == 4
    assert len(specialty_comparison) == 14
    assert len(clinical_atlas) == 7
    assert len(evidence_audit) == 8
    assert sampling_design["evaluations"].sum() == len(primary)
    assert len(sensitivities) == 7
    assert len(prevalence_experiment) == 2
    assert len(youden_experiment) == 3
    assert len(supplemental_atlas) == 24
    assert len(clinical_atlas_ledger) == 24
    assert specialty_comparison["evaluations"].ge(50).all()
    assert specialty_comparison["specialty"].is_unique
    assert specialty_comparison["rho_sample_size_percent"].lt(0).all()
    print(json.dumps(summary, indent=2))


if __name__ == "__main__":
    main(Path(sys.argv[1]) if len(sys.argv) > 1 else SOURCE)
```
