## Supplementary material for "Meaning of clinical calculator results: a cross-sectional analysis of the MDCalc catalogue": Reproducibility archive: code, data and analysis outputs: build_result_support.py

```
"""Build evaluation-level post-result risk and precision outputs.

This script deliberately does not rank calculators or select a calculator's
most favourable outcome. Each row remains one
calculator-study-outcome-threshold evaluation, in source-index order. Wilson
95% intervals describe the sampling precision of the observed post-negative
and post-positive risks.

The frozen primary dataset does not contain a separate, uniformly populated
exact-outcome field. ``source_label`` and ``source_disease`` are retained as
context, but users must verify the exact outcome in the cited source before
applying an estimate clinically.

Outputs:
    ruleout_support_evaluations.csv
    rulein_support_evaluations.csv
    result_support_summary.json
"""

import json
from pathlib import Path

import numpy as np
import pandas as pd


HERE = Path(__file__).resolve().parent
DATA = HERE / "data" / "mdcalc_evaluations_primary.csv"
THRESHOLDS = [0.005, 0.01, 0.02, 0.05, 0.10, 0.20]
Z = 1.959963984540054


def wilson(k, n, z=Z):
    """Return point estimate and Wilson score interval."""
    k = np.asarray(k, dtype=float)
    n = np.asarray(n, dtype=float)
    with np.errstate(invalid="ignore", divide="ignore"):
        p = np.where(n > 0, k / n, np.nan)
        denominator = 1 + z**2 / n
        centre = (p + z**2 / (2 * n)) / denominator
        half_width = (z / denominator) * np.sqrt(
            p * (1 - p) / n + z**2 / (4 * n**2)
        )
    return p, np.clip(centre - half_width, 0, 1), np.clip(centre + half_width, 0, 1)


def lowest_cleared(upper_bound):
    """Lowest action threshold strictly above the Wilson upper bound."""
    return np.array(
        [min((t for t in THRESHOLDS if value < t), default=np.nan) for value in upper_bound],
        dtype=float,
    )


def highest_cleared(lower_bound):
    """Highest action threshold strictly below the Wilson lower bound."""
    return np.array(
        [max((t for t in THRESHOLDS if value > t), default=np.nan) for value in lower_bound],
        dtype=float,
    )


df = pd.read_csv(DATA, encoding="utf-8-sig")
for column in ["tp", "fp", "fn", "tn"]:
    df[column] = pd.to_numeric(df[column], errors="coerce")

df["N"] = df.tp + df.fp + df.fn + df.tn
df["n_negative"] = df.fn + df.tn
df["n_positive"] = df.tp + df.fp
df["negative_fraction"] = df.n_negative / df.N
df["positive_fraction"] = df.n_positive / df.N
df["observed_prevalence"] = (df.tp + df.fn) / df.N
df["post_negative_risk"], df["post_negative_risk_lo"], df["post_negative_risk_hi"] = wilson(
    df.fn, df.n_negative
)
df["post_positive_risk"], df["post_positive_risk_lo"], df["post_positive_risk_hi"] = wilson(
    df.tp, df.n_positive
)
df["lowest_ruleout_threshold_supported"] = lowest_cleared(df.post_negative_risk_hi.values)
df["highest_rulein_threshold_supported"] = highest_cleared(df.post_positive_risk_lo.values)

source = df.data_source.fillna("")
df["provenance"] = np.where(
    source.str.startswith("v2_llm"),
    "model-extracted",
    np.where(source == "", "no recorded source", "human-extracted"),
)
df["exact_outcome_note"] = (
    "Exact outcome is not separately encoded in every frozen row; verify against source."
)

identifiers = [
    "source_index",
    "source_excel_row",
    "calculator_key",
    "catalog_id",
    "canonical_name",
    "catalog_url",
    "source_label",
    "source_disease",
    "study",
    "cutoff",
    "catalog_purpose",
    "sampling_design",
    "data_source",
    "provenance",
    "confidence",
    "confidence_group",
    "evidence_group",
    "exact_outcome_note",
]

ruleout_columns = identifiers + [
    "N",
    "tp",
    "fp",
    "fn",
    "tn",
    "n_negative",
    "negative_fraction",
    "observed_prevalence",
    "post_negative_risk",
    "post_negative_risk_lo",
    "post_negative_risk_hi",
    "lowest_ruleout_threshold_supported",
]
rulein_columns = identifiers + [
    "N",
    "tp",
    "fp",
    "fn",
    "tn",
    "n_positive",
    "positive_fraction",
    "observed_prevalence",
    "post_positive_risk",
    "post_positive_risk_lo",
    "post_positive_risk_hi",
    "highest_rulein_threshold_supported",
]

# Source order is stable and is not a clinical or statistical ordering.
ordered = df.sort_values("source_index", kind="stable")
ordered[ruleout_columns].to_csv(
    HERE / "ruleout_support_evaluations.csv", index=False, float_format="%.10g"
)
ordered[rulein_columns].to_csv(
    HERE / "rulein_support_evaluations.csv", index=False, float_format="%.10g"
)

summary = {
    "manuscript_version": "BMJ 0.14.0",
    "unit_of_analysis": "calculator-study-outcome-threshold evaluation",
    "ordering": "source_index only; no clinical ranking is provided",
    "outcome_caveat": (
        "The exact outcome is not separately encoded in every frozen row; "
        "source_label and source_disease provide context, and the cited source must be verified."
    ),
    "n_evaluations": int(len(df)),
    "thresholds": THRESHOLDS,
    "n_point_estimate_under_2pct": int((df.post_negative_risk < 0.02).sum()),
    "n_upper_bound_under_2pct": int((df.post_negative_risk_hi < 0.02).sum()),
    "n_point_estimate_under_2pct_not_supported_by_upper_bound": int(
        ((df.post_negative_risk < 0.02) & (df.post_negative_risk_hi >= 0.02)).sum()
    ),
    "n_zero_observed_false_negatives": int((df.fn == 0).sum()),
    "n_zero_false_negatives_clearing_2pct": int(
        ((df.fn == 0) & (df.post_negative_risk_hi < 0.02)).sum()
    ),
    "provenance_counts": {str(k): int(v) for k, v in df.provenance.value_counts().items()},
}

assert summary["n_evaluations"] == 482
assert summary["n_point_estimate_under_2pct"] == 157
assert summary["n_upper_bound_under_2pct"] == 67
assert summary["n_point_estimate_under_2pct_not_supported_by_upper_bound"] == 90
assert summary["n_zero_observed_false_negatives"] == 42
assert summary["n_zero_false_negatives_clearing_2pct"] == 15

(HERE / "result_support_summary.json").write_text(
    json.dumps(summary, indent=2) + "\n", encoding="utf-8"
)
print(json.dumps(summary, indent=2))
```
