## Supplementary material for "Meaning of clinical calculator results: a cross-sectional analysis of the MDCalc catalogue": Reproducibility archive: code, data and analysis outputs: DATA_DICTIONARY.md

# Data dictionary

| Variable | Definition |
| --- | --- |
| `source_excel_row` | One-based source worksheet row including the header offset |
| `source_label` | Calculator or tool label in the complete evidence worksheet |
| `catalog_id` | Numeric MDCalc catalogue identifier; missing for tools outside the catalogue |
| `canonical_name` | Catalogue name or assigned identity for a tool outside the catalogue |
| `calculator_key` | Stable clustered-analysis identity |
| `study` | Source-study citation recorded in the workbook |
| `cutoff` | Evaluated binary threshold or classification rule |
| `reported_sample_size` | Sample size stated in the extracted record |
| `tp`, `fp`, `fn`, `tn` | Final 2×2 cells used by the analysis |
| `data_source` | Original workbook lineage label |
| `evidence_group` | Mutually exclusive route: count-based, reported sensitivity/specificity, metric-reconstructed, or legacy/no recorded source |
| `confidence_group` | Normalised extraction-confidence label |
| `sampling_design` | Cohort/consecutive, case-control/two-gate, or unclear, based only on explicit wording in the source description |
| `source_correction` | Analytic correction and source rationale, if any |
| `sensitivity` | TP/(TP+FN) |
| `specificity` | TN/(TN+FP) |
| `prevalence` | Observed outcome frequency, (TP+FN)/N, in the analytic table |
| `youden_j` | Sensitivity + specificity − 1 |
| `parent_entropy_bits` | Binary outcome entropy before classification |
| `conditional_entropy_bits` | Weighted binary outcome entropy after classification |
| `entropy_reduction_bits` | Parent entropy minus conditional entropy |
| `entropy_removed_percent` | 100 × entropy reduction / parent entropy |
| `positive_information_bits`, `negative_information_bits` | Probability-weighted information contributed by each classification |
| `positive_information_share_percent`, `negative_information_share_percent` | Percentage of total information contributed by each classification |
| `included` | Whether the record remains after duplicate and supersession rules |
| `exclusion_reason` | Deterministic reason for exclusion, if applicable |
