## Supplementary material for "Meaning of clinical calculator results: a cross-sectional analysis of the MDCalc catalogue": Reproducibility archive: code, data and analysis outputs: decision_relevance.py

```
"""Decision-relevance analyses retained for the MDCalc BMJ 0.14.0 package.

Computes, from the frozen 11 August 2026 primary dataset:
  A. Decision-relevance interval [r-, r+] per evaluation and its width.
  B. Whether decision width is redundant with Youden's J (the novelty test).
  C. Threshold-flip census: same score, same cohort, different cut point.
  D. Stratum-specific medians (replacing the cancellation argument).
  E. Cohort-level clustering for CIs, not calculator-level.
  F. Bound on the max-Youden extraction rule's effect on the headline.

Reads only. Writes JSON to stdout and the scratchpad.
"""

import json
import sys
from pathlib import Path

import numpy as np
import pandas as pd
from scipy import stats

HERE = Path(__file__).resolve().parent
DATA = HERE / "data" / "mdcalc_evaluations_primary.csv"
OUT = HERE

RNG = np.random.default_rng(20260830)
B = 2000

df = pd.read_csv(DATA, encoding="utf-8-sig")
res = {"n_rows": int(len(df))}


def med_iqr(x):
    x = np.asarray(pd.to_numeric(x, errors="coerce").dropna(), dtype=float)
    if x.size == 0:
        return None
    return {
        "n": int(x.size),
        "median": float(np.median(x)),
        "q1": float(np.percentile(x, 25)),
        "q3": float(np.percentile(x, 75)),
    }


# ---------------------------------------------------------------- cells
for c in ["tp", "fp", "fn", "tn"]:
    df[c] = pd.to_numeric(df[c], errors="coerce")

tp, fp, fn, tn = df.tp, df.fp, df.fn, df.tn
n = tp + fp + fn + tn
df["_n"] = n
df["_pos_n"] = tp + fp                      # people getting a positive result
df["_neg_n"] = fn + tn
df["_p_pos"] = df._pos_n / n
df["_p_neg"] = df._neg_n / n
# post-result risks
df["_r_pos"] = np.where(df._pos_n > 0, tp / df._pos_n.replace(0, np.nan), np.nan)   # PPV
df["_r_neg"] = np.where(df._neg_n > 0, fn / df._neg_n.replace(0, np.nan), np.nan)   # 1 - NPV
df["_prev"] = (tp + fn) / n
df["_sens"] = np.where((tp + fn) > 0, tp / (tp + fn).replace(0, np.nan), np.nan)
df["_spec"] = np.where((fp + tn) > 0, tn / (fp + tn).replace(0, np.nan), np.nan)
df["_J"] = df._sens + df._spec - 1

# A. decision-relevance interval
df["_dec_width"] = df._r_pos - df._r_neg     # threshold range where the result decides

res["decision_width"] = med_iqr(df._dec_width)
res["pct_removed"] = med_iqr(df["entropy_removed_percent"] if "entropy_removed_percent" in df else df["% Removed"])
res["youden"] = med_iqr(df._J)

# B. novelty test -- is decision width redundant with J?
def spearman(a, b):
    m = pd.notna(a) & pd.notna(b)
    if m.sum() < 3:
        return None
    r, p = stats.spearmanr(a[m], b[m])
    return {"rho": float(r), "n": int(m.sum())}


pct_col = "entropy_removed_percent" if "entropy_removed_percent" in df else "% Removed"
df["_pct"] = pd.to_numeric(df[pct_col], errors="coerce")

res["redundancy"] = {
    "pct_removed_vs_J": spearman(df._pct, df._J),
    "decision_width_vs_J": spearman(df._dec_width, df._J),
    "decision_width_vs_pct_removed": spearman(df._dec_width, df._pct),
    "decision_width_vs_prevalence": spearman(df._dec_width, df._prev),
    "pct_removed_vs_prevalence": spearman(df._pct, df._prev),
}

# positive-result information share -- use the paper's own column.
# NB: contributions can be negative (a result can move risk toward 50/50 and raise
# entropy), so a naive cpos/(cpos+cneg) ratio is not the share and must not be used.
df["_pos_share"] = pd.to_numeric(df["positive_information_share_percent"], errors="coerce")
res["redundancy"]["pos_share_vs_J"] = spearman(df._pos_share, df._J)

# C. threshold-flip census
key_cols = [c for c in ["calculator_key", "canonical_name", "catalog_id"] if c in df]
kc = key_cols[0]
df["_cohort"] = df[kc].astype(str) + "|" + df._n.astype("Int64").astype(str)

flips = []
for cohort, g in df.groupby("_cohort", dropna=True):
    g2 = g.dropna(subset=["_pos_share"])
    if len(g2) < 2:
        continue
    lo, hi = g2._pos_share.min(), g2._pos_share.max()
    dom = lambda v: "positive" if v > 60 else ("negative" if v < 40 else "balanced")
    doms = {dom(s) for s in g2._pos_share}
    flipped = ("positive" in doms and "negative" in doms)
    flips.append({
        "calculator": str(g2[kc].iloc[0]), "name": str(g2.get("canonical_name", pd.Series([""])).iloc[0])[:40], "cohort": str(cohort), "k": int(len(g2)),
        "pos_share_min": float(lo), "pos_share_max": float(hi),
        "span": float(hi - lo), "dominance_set": sorted(doms), "flipped": bool(flipped),
        "cutoffs": [str(x) for x in g2.get("cutoff", pd.Series(dtype=str)).tolist()],
    })
flips.sort(key=lambda d: -d["span"])
res["threshold_flip"] = {
    "n_calculator_cohort_groups_with_multiple_thresholds": len(flips),
    "n_flipped_full_reversal": sum(f["flipped"] for f in flips),
    "n_changed_dominance_class": sum(len(f["dominance_set"]) > 1 for f in flips),
    "top": flips[:15],
}

# C2. decisiveness across a spanning menu of plausible action thresholds
THRESHOLDS = [0.01, 0.02, 0.05, 0.10, 0.20, 0.50]
for t in THRESHOLDS:
    df[f"_dec_{t}"] = (df._r_neg < t) & (df._r_pos > t)
df["_n_dec"] = df[[f"_dec_{t}" for t in THRESHOLDS]].sum(axis=1)
res["decisiveness"] = {
    "thresholds": THRESHOLDS,
    "counts_by_n_thresholds_decisive": {str(k): int(v) for k, v in df._n_dec.value_counts().sort_index().items()},
    "n_never_decisive": int((df._n_dec == 0).sum()),
    "pct_never_decisive": float(100 * (df._n_dec == 0).mean()),
}
res["redundancy"]["J_vs_n_thresholds_decisive"] = spearman(df._J, df._n_dec)
res["redundancy"]["pct_removed_vs_n_thresholds_decisive"] = spearman(df._pct, df._n_dec)

# D. stratum medians
res["strata"] = {}
for col in ["data_source", "evidence_group", "confidence_group", "sampling_design"]:
    if col not in df:
        continue
    out = {}
    for k, g in df.groupby(df[col].fillna("(missing)")):
        out[str(k)] = {"pct_removed": med_iqr(g._pct), "decision_width": med_iqr(g._dec_width),
                       "n_median": med_iqr(g._n)}
    res["strata"][col] = out

# LLM vs non-LLM (the max-Youden rule applies to LLM rows)
df["_llm"] = df.data_source.fillna("").str.startswith("v2_llm")
res["llm_vs_not"] = {
    "llm": {"pct": med_iqr(df.loc[df._llm, "_pct"]), "J": med_iqr(df.loc[df._llm, "_J"]),
            "n": med_iqr(df.loc[df._llm, "_n"]), "rows": int(df._llm.sum())},
    "not_llm": {"pct": med_iqr(df.loc[~df._llm, "_pct"]), "J": med_iqr(df.loc[~df._llm, "_J"]),
                "n": med_iqr(df.loc[~df._llm, "_n"]), "rows": int((~df._llm).sum())},
}
# F. bound: headline recomputed excluding rows subject to the max-Youden rule
res["headline_sensitivity"] = {
    "all_482": med_iqr(df._pct),
    "excluding_llm_rows": med_iqr(df.loc[~df._llm, "_pct"]),
    "count_based_only": med_iqr(df.loc[df.get("evidence_group", pd.Series(dtype=str)).astype(str).str.contains("ount", na=False), "_pct"]),
}

# E. clustering: cohort vs calculator
def boot_median(values, groups, B=B):
    gser = pd.Series(groups).fillna("(na)").astype(str).values
    vals = pd.to_numeric(pd.Series(values), errors="coerce").values
    ok = ~pd.isna(vals)
    vals, gser = vals[ok], gser[ok]
    uniq = np.unique(gser)
    idx = {g: np.where(gser == g)[0] for g in uniq}
    meds = np.empty(B)
    for b in range(B):
        pick = RNG.choice(uniq, size=uniq.size, replace=True)
        sel = np.concatenate([idx[g] for g in pick])
        meds[b] = np.median(vals[sel])
    return {"median": float(np.median(vals)),
            "ci_low": float(np.percentile(meds, 2.5)),
            "ci_high": float(np.percentile(meds, 97.5)),
            "n_clusters": int(uniq.size)}


res["clustering"] = {
    "by_calculator": boot_median(df._pct, df[kc]),
    "by_cohort": boot_median(df._pct, df._cohort),
}
# how much sharing is there?
res["cohort_sharing"] = {
    "n_distinct_cohort_keys": int(df._cohort.nunique()),
    "n_rows_sharing_exact_sample_size": int(df._n.duplicated(keep=False).sum()),
}

# rule-out subgroup, and the fn=0 concern
ro = df[df.get("catalog_purpose", pd.Series(dtype=str)).astype(str).str.contains("rule", case=False, na=False)]
res["rule_out"] = {
    "n": int(len(ro)),
    "neg_share_median": (1 - ro._pos_share).median() if len(ro) else None,
    "n_with_fn_zero": int((ro.fn == 0).sum()) if len(ro) else 0,
}

out_path = OUT / "decision_relevance_results.json"
out_path.write_text(json.dumps(res, indent=2, default=str), encoding="utf-8")
print(json.dumps(res, indent=2, default=str)[:6000])
print(f"\n\nWROTE {out_path}")
```
