## Supplementary material for "Meaning of clinical calculator results: a cross-sectional analysis of the MDCalc catalogue": Reproducibility archive: code, data and analysis outputs: figure5_ruleout_support.py

```
"""Figure 5 - whether a negative classification is supported by its study.

Deterministic, matching the repository figure contract used by analyze.py:
same style sheet, same palette, same double-column width, colour and
grayscale variants, PNG/PDF/SVG, no timestamps in metadata.

Panel A: risk after a negative classification against the number of patients
classified negative, point estimate and 95% upper bound, with the 2% action
threshold marked. Evaluations sitting below the line on the point estimate but
above it on the upper bound are the ones the study cannot support.

Panel B: how many evaluations support each action threshold, point estimate
against upper bound.
"""

from pathlib import Path

import matplotlib
matplotlib.use("Agg")
import matplotlib.pyplot as plt
import numpy as np
import pandas as pd

HERE = Path(__file__).resolve().parent
DATA = HERE / "data" / "mdcalc_evaluations_primary.csv"
FIGDIR = HERE / "figures"
BMJ_STYLE = HERE / "bmj.mplstyle"

INK, MUTED, WHITE = "#17212B", "#58636D", "#FFFFFF"
COLOURS = ("#0072B2", "#009E73", "#CC79A7", "#E69F00")
GRAYS = ("#111111", "#3D3D3D", "#696969", "#969696")
MM_PER_INCH, DOUBLE_COLUMN_MM, FIGURE_DPI = 25.4, 180.0, 600
THRESHOLDS = [0.005, 0.01, 0.02, 0.05, 0.10, 0.20]
Z = 1.959963984540054


def wilson_upper(k, n, z=Z):
    with np.errstate(invalid="ignore", divide="ignore"):
        p = np.where(n > 0, k / n, np.nan)
        d = 1 + z**2 / n
        c = (p + z**2 / (2 * n)) / d
        h = (z / d) * np.sqrt(p * (1 - p) / n + z**2 / (4 * n**2))
    return p, np.clip(c + h, 0, 1)


def configure(grayscale):
    plt.style.use("default")
    plt.style.use(str(BMJ_STYLE))
    text_colour = GRAYS[0] if grayscale else INK
    axis_colour = GRAYS[1] if grayscale else MUTED
    plt.rcParams.update({
        "svg.hashsalt": "mdcalc-entropy-bmj-v0.14.0",
        "text.color": text_colour, "axes.labelcolor": text_colour,
        "axes.edgecolor": axis_colour, "xtick.color": axis_colour, "ytick.color": axis_colour,
    })
    return GRAYS if grayscale else COLOURS


def save(fig, stem):
    fig.savefig(stem.with_suffix(".png"), format="png", dpi=FIGURE_DPI,
                facecolor=WHITE, edgecolor="none",
                metadata={"Software": "MDCalc BMJ 0.14.0 deterministic figure builder"})
    fig.savefig(stem.with_suffix(".pdf"), format="pdf", facecolor=WHITE, edgecolor="none",
                metadata={"Title": stem.stem,
                          "Author": "MDCalc BMJ 0.14.0 deterministic figure builder",
                          "Creator": "analysis/figure5_ruleout_support.py",
                          "CreationDate": None, "ModDate": None})
    svg = stem.with_suffix(".svg")
    fig.savefig(svg, format="svg", facecolor=WHITE, edgecolor="none",
                metadata={"Title": stem.stem,
                          "Creator": "analysis/figure5_ruleout_support.py", "Date": "2026-08-30"})
    svg.write_text("\n".join(l.rstrip() for l in svg.read_text(encoding="utf-8").splitlines()) + "\n",
                   encoding="utf-8", newline="\n")
    plt.close(fig)


df = pd.read_csv(DATA, encoding="utf-8-sig")
for c in ["tp", "fp", "fn", "tn"]:
    df[c] = pd.to_numeric(df[c], errors="coerce")
n_neg = df.fn + df.tn
r_neg, r_hi = wilson_upper(df.fn.values, n_neg.values)
ok = np.isfinite(r_neg) & np.isfinite(r_hi) & (n_neg.values > 0)
r_neg, r_hi, n_neg = r_neg[ok], r_hi[ok], n_neg.values[ok]

supported = r_hi < 0.02
looks_only = (r_neg < 0.02) & ~supported


def build(colours, grayscale):
    fig, (ax1, ax2) = plt.subplots(
        1, 2, figsize=(DOUBLE_COLUMN_MM / MM_PER_INCH, 78.0 / MM_PER_INCH))

    faint = GRAYS[3] if grayscale else "#9AA8B8"
    ax1.scatter(n_neg[~(supported | looks_only)], 100 * r_neg[~(supported | looks_only)],
                s=7, c=faint, alpha=.55, linewidths=0, label="Above 2% either way")
    ax1.scatter(n_neg[looks_only], 100 * r_neg[looks_only], s=13, c=colours[2],
                linewidths=0, label="Appears to clear 2%, not supported")
    ax1.scatter(n_neg[supported], 100 * r_neg[supported], s=13, c=colours[1],
                linewidths=0, label="Supported at 2%")
    ax1.axhline(2.0, color=GRAYS[1] if grayscale else MUTED, lw=.8, ls="--")
    ax1.annotate("2% action threshold", (1.6, 2.3), fontsize=7,
                 color=GRAYS[1] if grayscale else MUTED)
    ax1.set_xscale("log")
    ax1.set_yscale("symlog", linthresh=1.0)
    ax1.set_ylim(bottom=-0.35)   # risk cannot be negative; suppress the symlog decade below 0
    ax1.set_xlabel("Patients classified negative")
    ax1.set_ylabel("Risk after a negative classification (%)")
    ax1.set_title("A  Small negative groups cannot support a low risk", loc="left")
    ax1.legend(loc="upper right", frameon=True, facecolor=WHITE,
               edgecolor="none", framealpha=.88)

    pt = [100 * np.mean(r_neg < t) for t in THRESHOLDS]
    ub = [100 * np.mean(r_hi < t) for t in THRESHOLDS]
    x = np.arange(len(THRESHOLDS))
    ax2.bar(x - .19, pt, .38, color=faint, label="Point estimate")
    ax2.bar(x + .19, ub, .38, color=colours[0], label="95% upper bound")
    for xi, (a, b) in enumerate(zip(pt, ub)):
        ax2.annotate(f"{a:.0f}", (xi - .19, a), ha="center", va="bottom", fontsize=6.5)
        ax2.annotate(f"{b:.0f}", (xi + .19, b), ha="center", va="bottom", fontsize=6.5)
    ax2.set_xticks(x)
    ax2.set_xticklabels([f"{100*t:g}%" for t in THRESHOLDS])
    ax2.set_xlabel("Action threshold")
    ax2.set_ylabel("Evaluations reaching it (%)")
    ax2.set_title("B  Requiring the bound roughly halves the count", loc="left")
    ax2.legend(loc="upper left", frameon=True, facecolor=WHITE,
               edgecolor="none", framealpha=.88)

    fig.tight_layout(pad=0.6)
    return fig


for gs in (False, True):
    cols = configure(gs)
    save(build(cols, gs), FIGDIR / f"figure5_ruleout_support{'_grayscale' if gs else ''}")

print(f"figure5 written to {FIGDIR}")
print(f"  supported at 2%: {supported.sum()}   appears-but-not-supported: {looks_only.sum()}"
      f"   point-estimate under 2%: {(r_neg < 0.02).sum()}")
```
