## Supplementary material for "Meaning of clinical calculator results: a cross-sectional analysis of the MDCalc catalogue": Reproducibility archive: code, data and analysis outputs: graphical_abstract.pdf

### What does a clinical calculator result mean?

#### CATALOGUE EVIDENCE

**482**

binary evaluations

407 calculators represented

#### HOW MUCH WAS ADDED?

21.2% below 5%

10.0% at least 50%

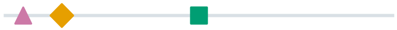

14.9% median

Percentage of starting uncertainty removed

#### WHICH CLASSIFICATION?

**68.3%**

result dominant
(329 of 482 evaluations)

**71.3%**

median negative share among rule-out evaluations

Average information describes learning in an evaluated population; it is not clinical utility.
