## Supplementary material for "Meaning of clinical calculator results: a cross-sectional analysis of the MDCalc catalogue": Reproducibility archive: code, data and analysis outputs: graphical_abstract_grayscale.pdf

### What does a clinical calculator result mean?

**CATALOGUE EVIDENCE**

# 482

binary evaluations

407 calculators represented

**HOW MUCH WAS ADDED?**

21.2% below 5%

10.0% at least 50%

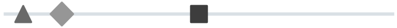

14.9% median

Percentage of starting uncertainty removed

**WHICH CLASSIFICATION?**

# 68.3%

result dominant
(329 of 482 evaluations)

## 71.3%

median negative share among rule-out evaluations

Average information describes learning in an evaluated population; it is not clinical utility.
