## Supplementary material for "Meaning of clinical calculator results: a cross-sectional analysis of the MDCalc catalogue": Reproducibility archive: code, data and analysis outputs: LICENSE.md

# Licensing

This repository uses a scoped licence split so that original code and first-party research outputs can be reused without purporting to relicense third-party material.

## Original analysis code: MIT License

Original software included in a tagged public release of this repository is licensed under the MIT License.

Copyright (c) 2026 MDCalc entropy manuscript authors

Permission is hereby granted, free of charge, to any person obtaining a copy of this software and associated documentation files (the "Software"), to deal in the Software without restriction, including without limitation the rights to use, copy, modify, merge, publish, distribute, sublicense, and/or sell copies of the Software, and to permit persons to whom the Software is furnished to do so, subject to the following conditions:

The above copyright notice and this permission notice shall be included in all copies or substantial portions of the Software.

THE SOFTWARE IS PROVIDED "AS IS", WITHOUT WARRANTY OF ANY KIND, EXPRESS OR IMPLIED, INCLUDING BUT NOT LIMITED TO THE WARRANTIES OF MERCHANTABILITY, FITNESS FOR A PARTICULAR PURPOSE AND NONINFRINGEMENT. IN NO EVENT SHALL THE AUTHORS OR COPYRIGHT HOLDERS BE LIABLE FOR ANY CLAIM, DAMAGES OR OTHER LIABILITY, WHETHER IN AN ACTION OF CONTRACT, TORT OR OTHERWISE, ARISING FROM, OUT OF OR IN CONNECTION WITH THE SOFTWARE OR THE USE OR OTHER DEALINGS IN THE SOFTWARE.

## First-party data and documentation: CC BY 4.0

First-party derived datasets, generated tables and figures, and documentation included in a tagged public release, where the authors have authority to grant a licence, are licensed under the Creative Commons Attribution 4.0 International licence: https://creativecommons.org/licenses/by/4.0/.

## Exclusions

Third-party source material, linked publications, institutional records, and any file marked restricted or excluded are not covered by these grants. Their original terms continue to apply. A public release must include only material that the authors are permitted to redistribute.
