## Supplementary material for "Meaning of clinical calculator results: a cross-sectional analysis of the MDCalc catalogue": Reproducibility archive: code, data and analysis outputs: LICENSING_STATUS.md

# Licensing status for BMJ Evidence-Based Medicine 0.1.4

The reproducibility archive is supplied for editorial and peer-review scrutiny. The versioned repository is available at https://github.com/EMAI-Research/mdcalc-entropy.

- Original analysis code: MIT License.
- First-party derived data, generated tables and figures, and documentation that the authors have authority to license: Creative Commons Attribution 4.0 International, https://creativecommons.org/licenses/by/4.0/.
- Third-party source material: governed by its original terms and not relicensed.

The scoped repository notice is `LICENSE.md`. A Zenodo DOI is not claimed in the manuscript because no stable record has yet been cited. If a Zenodo archive is created before acceptance, add only the actual minted DOI and release identifier.
