## Supplementary material for "Meaning of clinical calculator results: a cross-sectional analysis of the MDCalc catalogue": Reproducibility archive: code, data and analysis outputs: README.md

# Reproduce the BMJ Evidence-Based Medicine 0.1.4 analysis

This is the reproducibility archive for the BMJ Evidence-Based Medicine submission package 0.1.4. It retains the frozen analysis from BMJ 0.14.0 (data freeze 11 August 2026; historical release tag `bmj-v0.14.0`). This patch corrects submission metadata and these instructions; scientific inputs, code, and results are unchanged. The public repository is https://github.com/EMAI-Research/mdcalc-entropy. This ZIP is the reproducibility record supplied with the submission.

The scientific data and calculations remain frozen. `analyze.py` verifies the source-workbook hash, rebuilds the cleaned datasets, recalculates all reported measures, writes machine-readable tables and exploratory outputs, and stops if a consistency check fails. `redesign_figures.py` builds the publication figures without modifying data. `build_result_support.py` writes post-negative and post-positive risk estimates with Wilson 95% intervals for every evaluation.

## Environment

Use Python 3.12 with the pinned packages in `requirements.txt`: matplotlib 3.10.9, NumPy 2.0.2, openpyxl 3.1.5, pandas 3.0.3, and SciPy 1.17.1.

Extract the reproducibility ZIP into a new folder. Open PowerShell in that folder (the one containing analyze.py and requirements.txt), then run:

```powershell
python -m venv .venv
.\.venv\Scripts\python.exe -m pip install -r requirements.txt
.\.venv\Scripts\python.exe analyze.py
.\.venv\Scripts\python.exe build_result_support.py
.\.venv\Scripts\python.exe redesign_figures.py
```

These commands regenerate analysis outputs, support estimates and the archived figure set. Historical figure filenames retain their BMJ provenance. The archive does not contain the complete EBM manuscript, supplement or submission-package build sources; the nine submission files are supplied separately in the outer package.

MANIFEST.json records SHA-256 checksums and sizes for the supplied archive contents. Verify it before running the scripts, which regenerate output files.

## Frozen input and expected results

The publication workbook is `source/mdcalc_entropy_source_v0.2.0.xlsx`, SHA-256 `02348f2e95039be35943e135d27b12010ba382d589d03623a497baef35dbc514`. Bootstrap and figure-jitter random number generators use seed `20260811`.

The expected primary results are 482 evaluations representing 407 calculators; median reduction in starting uncertainty 14.9%, or 0.093 bits; 329 evaluations in which one classification supplies more than 60% of average information; and median negative-result information share 71.3% (95% confidence interval 65.6% to 79.0%) among 56 rule-out evaluations. Post-negative risk is below 2% in 157 point estimates but in only 67 Wilson upper bounds; 42 evaluations have no observed false negatives, of which 15 support a 2% threshold by their upper bound.

## File map

- `source/`: frozen source workbook and data dictionary.
- `data/`: primary, expanded, catalogue, and row-level audit datasets.
- `tables/`: full generated manuscript, sensitivity, correlation, specialty, clinical-example, and purpose-coverage tables. Outputs omitted from the lean PDF remain exploratory or supporting material, not primary findings.
- `figures/`: publication figures plus removed study-size, distribution, prevalence, dominance, and other supporting figures retained for auditability.
- `analyze.py`: deterministic reconstruction, analysis, assertions, tables, and legacy supporting figures.
- `redesign_figures.py`: final publication-figure generator.
- `decision_relevance.py`, `decision_relevance_results.json`, and `figure5_ruleout_support.py`: supporting decision-threshold analysis and figure code.
- `ruleout_support_evaluations.csv` and `rulein_support_evaluations.csv`: complete evaluation-level support estimates in source-index order; these files are not rankings.
- `result_support_summary.json`: headline precision counts used in the manuscript.
- `results_summary.json` and `results_summary.md`: machine-readable and concise analysis summaries, including archived exploratory results.

## Interpretation guard for support files

Each support-file row is one calculator-study-outcome-threshold evaluation. No calculator-level “best outcome” is selected and no ordinal quality rank is supplied. Comparisons across rows may involve different outcomes, definitions, thresholds, designs, and clinical settings. The frozen dataset does not contain a separate, uniformly populated exact-outcome field, so `source_label` and `source_disease` provide context and users must verify the exact outcome in the cited source before applying an estimate clinically. Zero observed events do not imply zero residual risk; the Wilson upper bound shows how much risk remains compatible with the evaluation's denominator.

The regenerated primary CSV may differ in non-numeric evidence-group labels from earlier releases because four rows were reassigned into exhaustive provenance groups and a legacy label was clarified. Numeric analytic values and headline results are unchanged.
