## Supplementary material for "Meaning of clinical calculator results: a cross-sectional analysis of the MDCalc catalogue": Reproducibility archive: code, data and analysis outputs: redesign_figures.py

```
"""Build the BMJ 0.14.0 figure prototypes and selected main figures.

All values come from the frozen 11 August 2026 primary CSV. The script changes
only presentation: it neither edits the dataset nor creates a new scientific
measure. Run from any directory with Python 3.12 and the pinned analysis
requirements installed.
"""

from pathlib import Path

import matplotlib

matplotlib.use("Agg")
import matplotlib.pyplot as plt
from matplotlib.lines import Line2D
import numpy as np
import pandas as pd

HERE = Path(__file__).resolve().parent
DATA = HERE / "data" / "mdcalc_evaluations_primary.csv"
STYLE = HERE / "bmj.mplstyle"
FIGURES = HERE / "figures"
PROTOTYPES = HERE / "prototypes"

MM_PER_INCH = 25.4
WIDTH_MM = 180.0
DPI = 600
Z = 1.959963984540054
WHITE = "#FFFFFF"
INK = "#17212B"
MUTED = "#5C6770"
GRID = "#D8DEE4"
NEG = "#0072B2"
POS = "#D55E00"
INFO = "#6F4C9B"
BALANCED = "#8A949C"
NEG_DOM = "#009E73"
POS_DOM = "#E69F00"

EXAMPLE_ROWS = {
    241: "PERC",
    65: "Canadian CT Head Rule",
    145: "Glasgow-Blatchford",
    97: "CRB-65",
    101: "CURB-65",
    473: "HEART ≥4",
    474: "HEART >6",
}


def load_data() -> pd.DataFrame:
    df = pd.read_csv(DATA, encoding="utf-8-sig")
    for column in ("tp", "fp", "fn", "tn"):
        df[column] = pd.to_numeric(df[column], errors="raise")
    df = df.assign(
        start_risk=100 * df["prevalence"],
        positive_risk=100 * df["tp"] / (df["tp"] + df["fp"]),
        negative_risk=100 * df["fn"] / (df["fn"] + df["tn"]),
        negative_n=df["fn"] + df["tn"],
    )
    p = df["fn"] / df["negative_n"]
    denominator = 1 + Z**2 / df["negative_n"]
    centre = (p + Z**2 / (2 * df["negative_n"])) / denominator
    half_width = Z / denominator * np.sqrt(
        p * (1 - p) / df["negative_n"] + Z**2 / (4 * df["negative_n"] ** 2)
    )
    df["negative_risk_upper"] = 100 * np.clip(centre + half_width, 0, 1)
    assert len(df) == 482
    assert df["calculator_key"].nunique() == 407
    assert set(EXAMPLE_ROWS).issubset(set(df["source_index"]))
    assert int((df["negative_risk"] < 2).sum()) == 157
    assert int((df["negative_risk_upper"] < 2).sum()) == 67
    return df


def configure(grayscale: bool = False) -> dict[str, str]:
    plt.style.use("default")
    plt.style.use(str(STYLE))
    palette = {
        "ink": "#111111" if grayscale else INK,
        "muted": "#555555" if grayscale else MUTED,
        "grid": "#D0D0D0" if grayscale else GRID,
        "negative": "#222222" if grayscale else NEG,
        "positive": "#777777" if grayscale else POS,
        "information": "#444444" if grayscale else INFO,
        "balanced": "#9A9A9A" if grayscale else BALANCED,
        "negative_dominant": "#333333" if grayscale else NEG_DOM,
        "positive_dominant": "#6D6D6D" if grayscale else POS_DOM,
    }
    plt.rcParams.update(
        {
            "svg.hashsalt": "mdcalc-entropy-bmj-v0.14.0",
            "text.color": palette["ink"],
            "axes.labelcolor": palette["ink"],
            "axes.edgecolor": palette["muted"],
            "xtick.color": palette["muted"],
            "ytick.color": palette["muted"],
        }
    )
    return palette


def figure_size(height_mm: float) -> tuple[float, float]:
    return WIDTH_MM / MM_PER_INCH, height_mm / MM_PER_INCH


def save_final(fig: plt.Figure, stem: str, grayscale: bool) -> None:
    suffix = "_grayscale" if grayscale else ""
    path = FIGURES / f"{stem}{suffix}"
    metadata = {"Title": path.name, "Creator": "analysis/redesign_figures.py"}
    fig.savefig(
        path.with_suffix(".png"),
        dpi=DPI,
        facecolor=WHITE,
        edgecolor="none",
        metadata={"Software": "MDCalc BMJ 0.14.0 deterministic figure builder"},
    )
    fig.savefig(
        path.with_suffix(".pdf"),
        facecolor=WHITE,
        edgecolor="none",
        metadata={**metadata, "CreationDate": None, "ModDate": None},
    )
    svg = path.with_suffix(".svg")
    fig.savefig(svg, facecolor=WHITE, edgecolor="none", metadata={**metadata, "Date": "2026-08-30"})
    svg.write_text(
        "\n".join(line.rstrip() for line in svg.read_text(encoding="utf-8").splitlines()) + "\n",
        encoding="utf-8",
        newline="\n",
    )
    plt.close(fig)


def save_prototype(fig: plt.Figure, stem: str) -> None:
    fig.savefig(PROTOTYPES / f"{stem}.png", dpi=250, facecolor=WHITE, edgecolor="none")
    fig.savefig(
        PROTOTYPES / f"{stem}.pdf",
        facecolor=WHITE,
        edgecolor="none",
        metadata={"Title": stem, "Creator": "analysis/redesign_figures.py", "CreationDate": None},
    )
    plt.close(fig)


def direction_colours(df: pd.DataFrame, palette: dict[str, str]) -> np.ndarray:
    share = df["positive_information_share_percent"].to_numpy()
    return np.where(
        share < 40,
        palette["negative_dominant"],
        np.where(share > 60, palette["positive_dominant"], palette["balanced"]),
    )


def prototype_risk_transition(df: pd.DataFrame) -> plt.Figure:
    palette = configure()
    ordered = df.sort_values(["start_risk", "source_index"]).reset_index(drop=True)
    y = np.arange(len(ordered))
    fig, ax = plt.subplots(figsize=figure_size(112), constrained_layout=True)
    ax.hlines(y, ordered["negative_risk"], ordered["positive_risk"], color=palette["grid"], lw=0.45)
    ax.scatter(ordered["negative_risk"], y, s=3, color=palette["negative"], alpha=0.65)
    ax.scatter(ordered["start_risk"], y, s=3, color=palette["ink"], alpha=0.7)
    ax.scatter(ordered["positive_risk"], y, s=3, color=palette["positive"], alpha=0.65)
    ax.set(xlim=(-1, 101), ylim=(-4, len(ordered) + 3), xlabel="Observed risk (%)", ylabel="482 evaluations sorted by starting risk")
    ax.set_yticks([])
    ax.set_title("Prototype 1 — risk-transition atlas", loc="left")
    return fig


def prototype_ranked_landscape(df: pd.DataFrame) -> plt.Figure:
    palette = configure()
    ordered = df.sort_values(["entropy_removed_percent", "source_index"]).reset_index(drop=True)
    y = np.arange(len(ordered))
    fig, (ax1, ax2) = plt.subplots(1, 2, figsize=figure_size(112), sharey=True, constrained_layout=True)
    ax1.scatter(ordered["entropy_removed_percent"], y, s=4, color=palette["information"])
    ax1.set(xlabel="Starting uncertainty removed (%)", ylabel="Ordered evaluations")
    ax1.set_yticks([])
    delta_negative = ordered["start_risk"] - ordered["negative_risk"]
    delta_positive = ordered["positive_risk"] - ordered["start_risk"]
    ax2.hlines(y, -delta_negative, delta_positive, color=palette["grid"], lw=0.45)
    ax2.scatter(-delta_negative, y, s=3, color=palette["negative"])
    ax2.scatter(delta_positive, y, s=3, color=palette["positive"])
    ax2.axvline(0, color=palette["muted"], lw=0.7)
    ax2.set(xlabel="Change from starting risk (percentage points)")
    fig.suptitle("Prototype 2 — ranked landscape (ordered by information, not quality)", x=0.01, ha="left")
    return fig


def prototype_small_multiples(df: pd.DataFrame) -> plt.Figure:
    palette = configure()
    quartiles = pd.qcut(df["start_risk"], 4, labels=["Q1 lowest", "Q2", "Q3", "Q4 highest"])
    fig, axes = plt.subplots(2, 2, figsize=figure_size(112), sharex=True, sharey=True, constrained_layout=True)
    for ax, label in zip(axes.flat, quartiles.cat.categories):
        group = df.loc[quartiles.eq(label)]
        ax.scatter(
            group["entropy_removed_percent"],
            group["positive_information_share_percent"],
            s=7,
            c=direction_colours(group, palette),
            alpha=0.55,
            linewidths=0,
        )
        ax.axhline(50, color=palette["grid"], lw=0.7)
        ax.set_title(f"{label} starting risk (n={len(group)})", loc="left")
    axes[1, 0].set(xlabel="Uncertainty removed (%)", ylabel="Positive information share (%)")
    axes[1, 1].set(xlabel="Uncertainty removed (%)")
    axes[0, 0].set_ylabel("Positive information share (%)")
    fig.suptitle("Prototype 3 — prevalence small multiples", x=0.01, ha="left")
    return fig


def prototype_bivariate(df: pd.DataFrame) -> plt.Figure:
    palette = configure()
    fig, ax = plt.subplots(figsize=figure_size(112), constrained_layout=True)
    points = ax.scatter(
        df["negative_risk"],
        df["positive_risk"],
        c=df["start_risk"],
        s=7 + 0.5 * df["entropy_removed_percent"],
        cmap="viridis",
        alpha=0.62,
        linewidths=0,
    )
    ax.plot([0, 100], [0, 100], color=palette["grid"], lw=0.8)
    ax.set(xlim=(-1, 101), ylim=(-1, 101), xlabel="Risk after negative result (%)", ylabel="Risk after positive result (%)")
    fig.colorbar(points, ax=ax, label="Starting risk (%)", pad=0.02)
    ax.set_title("Prototype 4 — bivariate post-result-risk scatter", loc="left")
    return fig


def prototype_tile_atlas(df: pd.DataFrame) -> plt.Figure:
    palette = configure()
    ordered = df.sort_values("source_index").reset_index(drop=True)
    side = 22
    values = np.full(side * side, np.nan)
    values[: len(ordered)] = ordered["entropy_removed_percent"]
    share = np.full(side * side, np.nan)
    share[: len(ordered)] = ordered["positive_information_share_percent"]
    fig, (ax1, ax2) = plt.subplots(1, 2, figsize=figure_size(105), constrained_layout=True)
    im1 = ax1.imshow(values.reshape(side, side), vmin=0, vmax=100, cmap="Blues", interpolation="nearest")
    im2 = ax2.imshow(share.reshape(side, side), vmin=0, vmax=100, cmap="coolwarm", interpolation="nearest")
    for ax, title in zip((ax1, ax2), ("Uncertainty removed", "Positive information share")):
        ax.set_title(title, loc="left")
        ax.set_xticks([])
        ax.set_yticks([])
        for spine in ax.spines.values():
            spine.set_visible(False)
    fig.colorbar(im1, ax=ax1, label="%", shrink=0.74)
    fig.colorbar(im2, ax=ax2, label="%", shrink=0.74)
    fig.suptitle("Prototype 5 — paired tile atlas (source-row order)", x=0.01, ha="left")
    return fig


def hybrid_axes(df: pd.DataFrame, palette: dict[str, str], title: str) -> plt.Figure:
    ordered = df.sort_values(["start_risk", "source_index"]).reset_index(drop=True)
    y = np.arange(len(ordered))
    fig, axes = plt.subplots(
        1,
        3,
        figsize=figure_size(116),
        sharey=True,
        gridspec_kw={"width_ratios": [1.75, 0.9, 0.9]},
        constrained_layout=True,
    )
    risk, information, direction = axes
    risk.hlines(y, ordered["negative_risk"], ordered["positive_risk"], color=palette["grid"], lw=0.42)
    risk.scatter(ordered["negative_risk"], y, s=3.2, marker="o", color=palette["negative"], alpha=0.62, linewidths=0)
    risk.scatter(ordered["start_risk"], y, s=3.8, marker="D", color=palette["ink"], alpha=0.60, linewidths=0)
    risk.scatter(ordered["positive_risk"], y, s=4.0, marker="^", color=palette["positive"], alpha=0.62, linewidths=0)
    risk.set(xlim=(-1, 101), xlabel="Observed frequency (%)", ylabel="482 evaluations sorted by observed outcome frequency")
    information.scatter(ordered["entropy_removed_percent"], y, s=4, color=palette["information"], alpha=0.65, linewidths=0)
    information.axvline(14.9, color=palette["muted"], ls="--", lw=0.7)
    information.set(xlim=(-2, 102), xlabel="Starting uncertainty\nremoved (%)")
    share = ordered["positive_information_share_percent"]
    for mask, colour, marker in (
        (share.lt(40), palette["negative_dominant"], "<"),
        (share.between(40, 60), palette["balanced"], "o"),
        (share.gt(60), palette["positive_dominant"], ">"),
    ):
        direction.scatter(share[mask], y[mask], s=5, color=colour, marker=marker, alpha=0.72, linewidths=0)
    direction.axvline(50, color=palette["grid"], lw=0.7)
    direction.set(xlim=(-2, 102), xlabel="Information from\npositive result (%)")
    for ax, panel in zip(axes, ("A  Where risk ended", "B  What the result added", "C  Which result supplied it")):
        ax.set_title(panel, loc="left")
        ax.set_ylim(-4, len(ordered) + 3)
        ax.set_yticks([])
    handles = [
        Line2D([], [], marker="o", color="none", markerfacecolor=palette["negative"], markeredgecolor="none", label="After negative"),
        Line2D([], [], marker="o", color="none", markerfacecolor=palette["ink"], markeredgecolor="none", label="Observed outcome frequency"),
        Line2D([], [], marker="o", color="none", markerfacecolor=palette["positive"], markeredgecolor="none", label="After positive"),
    ]
    risk.legend(handles=handles, loc="lower right", ncol=1)
    fig.suptitle(title, x=0.01, ha="left", fontsize=9.5, weight="semibold")
    return fig


def prototype_hybrid(df: pd.DataFrame) -> plt.Figure:
    return hybrid_axes(df, configure(), "Prototype 6 — coordinated hybrid atlas")


def figure1_examples(df: pd.DataFrame, palette: dict[str, str]) -> plt.Figure:
    examples = df[df["source_index"].isin(EXAMPLE_ROWS)].copy()
    examples["label"] = examples["source_index"].map(EXAMPLE_ROWS)
    examples["order"] = examples["source_index"].map({key: i for i, key in enumerate(EXAMPLE_ROWS)})
    examples = examples.sort_values("order")
    y = np.arange(len(examples))[::-1]
    fig, (risk, contribution) = plt.subplots(
        1,
        2,
        figsize=figure_size(112),
        gridspec_kw={"width_ratios": [1.65, 1]},
        constrained_layout=True,
    )
    risk.hlines(y, examples["negative_risk"], examples["positive_risk"], color=palette["grid"], lw=1.2)
    risk.scatter(examples["negative_risk"], y, s=28, color=palette["negative"], zorder=3)
    risk.scatter(examples["start_risk"], y, s=24, marker="D", color=palette["ink"], zorder=4)
    risk.scatter(examples["positive_risk"], y, s=30, marker="^", color=palette["positive"], zorder=3)
    for yi, row in zip(y, examples.itertuples()):
        risk.text(row.negative_risk, yi - 0.20, f"{row.negative_risk:.1f}", ha="center", va="top", fontsize=6.3, color=palette["negative"])
        risk.text(row.start_risk, yi + 0.18, f"{row.start_risk:.1f}", ha="center", va="bottom", fontsize=6.3, color=palette["ink"])
        risk.text(row.positive_risk, yi - 0.20, f"{row.positive_risk:.1f}", ha="center", va="top", fontsize=6.3, color=palette["positive"])
    risk.set_yticks(y, examples["label"])
    risk.set(xlim=(-2, 75), ylim=(-0.7, len(examples) - 0.3), xlabel="Observed outcome risk (%)")
    risk.set_title("A  Where risk started and ended", loc="left")
    risk.text(0.02, 0.97, "● after negative", transform=risk.transAxes, va="bottom", fontsize=6.8, color=palette["negative"])
    risk.text(0.36, 0.97, "◆ starting risk", transform=risk.transAxes, va="bottom", fontsize=6.8, color=palette["ink"])
    risk.text(0.66, 0.97, "▲ after positive", transform=risk.transAxes, va="bottom", fontsize=6.8, color=palette["positive"])
    negative_share = 100 - examples["positive_information_share_percent"]
    contribution.barh(y, negative_share, color=palette["negative"], height=0.48, label="Negative result")
    contribution.barh(
        y,
        examples["positive_information_share_percent"],
        left=negative_share,
        color=palette["positive"],
        height=0.48,
        label="Positive result",
    )
    for yi, removed in zip(y, examples["entropy_removed_percent"]):
        contribution.text(102, yi, f"{removed:.1f}% added", va="center", fontsize=6.7, color=palette["information"])
    contribution.axvline(50, color=WHITE, lw=0.7)
    contribution.set_yticks([])
    contribution.set(xlim=(0, 127), ylim=risk.get_ylim(), xlabel="Share of average information (%)")
    contribution.set_title("B  Which result supplied it", loc="left")
    contribution.text(0.15, 0.97, "negative-result share", transform=contribution.transAxes, ha="center", va="top", fontsize=6.8, color=palette["negative"])
    contribution.text(0.77, 0.97, "positive-result share", transform=contribution.transAxes, ha="center", va="top", fontsize=6.8, color=palette["positive"])
    return fig


def figure3_precision(df: pd.DataFrame, palette: dict[str, str]) -> plt.Figure:
    point_under = df["negative_risk"].lt(2)
    supported = df["negative_risk_upper"].lt(2)
    looks_only = point_under & ~supported
    fig, ax = plt.subplots(figsize=figure_size(105), constrained_layout=True)
    ax.scatter(
        df.loc[~point_under, "negative_n"],
        df.loc[~point_under, "negative_risk"],
        s=7,
        color=palette["balanced"],
        alpha=0.32,
        linewidths=0,
        label="Point estimate at least 2%",
    )
    for mask, colour, marker in (
        (looks_only, palette["positive"], "o"),
        (supported, palette["negative_dominant"], "s"),
    ):
        ax.vlines(
            df.loc[mask, "negative_n"],
            df.loc[mask, "negative_risk"],
            df.loc[mask, "negative_risk_upper"],
            color=colour,
            alpha=0.30,
            lw=0.65,
        )
        ax.scatter(
            df.loc[mask, "negative_n"],
            df.loc[mask, "negative_risk"],
            s=12,
            marker=marker,
            color=colour,
            alpha=0.78,
            linewidths=0,
        )
        ax.scatter(
            df.loc[mask, "negative_n"],
            df.loc[mask, "negative_risk_upper"],
            s=8,
            marker=marker,
            facecolors=WHITE,
            edgecolors=colour,
            linewidths=0.55,
            alpha=0.82,
        )
    ax.axhline(2, color=palette["ink"], ls="--", lw=0.9)
    ax.text(1.1, 2.15, "2% threshold", fontsize=7.2, color=palette["ink"])
    ax.set_xscale("log")
    ax.set_yscale("symlog", linthresh=0.5)
    ax.set(xlabel="Patients classified negative", ylabel="Risk after a negative result (%)", ylim=(-0.12, 105))
    ax.set_title("Observed risk is a point; the interval shows what the evaluation can support", loc="left")
    zero = df["fn"].eq(0)
    ax.text(
        0.985,
        0.98,
        "At the 2% threshold\n157 point estimates below\n67 supported by upper bound\n90 not supported\n\nZero missed cases\n42 evaluations\n15 supported",
        transform=ax.transAxes,
        ha="right",
        va="top",
        fontsize=7.7,
        bbox={"boxstyle": "round,pad=0.4", "facecolor": WHITE, "edgecolor": palette["grid"], "alpha": 0.94},
    )
    label_rows = {
        222: ("NEXUS Head CT", 5400, 0.13),
        64: ("Canadian C-Spine Rule", 7600, 0.19),
        230: ("Ottawa Ankle Rule", 45, 4.5),
        159: ("Hestia Criteria", 165, 3.2),
        479: ("HEART Pathway (66 negative)", 85, 7.2),
    }
    for source_index, (label, text_x, text_y) in label_rows.items():
        row = df.loc[df["source_index"].eq(source_index)].iloc[0]
        y_anchor = row["negative_risk_upper"]
        ax.annotate(
            label,
            xy=(row["negative_n"], y_anchor),
            xytext=(text_x, text_y),
            arrowprops={"arrowstyle": "-", "color": palette["muted"], "lw": 0.55},
            fontsize=6.6,
            color=palette["ink"],
        )
    assert int(zero.sum()) == 42
    assert int((zero & supported).sum()) == 15
    ax.text(
        0.02,
        0.98,
        "Filled marker: observed risk\nOpen marker: 95% upper bound\nCircle: point <2%, bound ≥2%\nSquare: bound <2%",
        transform=ax.transAxes,
        ha="left",
        va="top",
        fontsize=6.8,
        color=palette["ink"],
        bbox={"boxstyle": "round,pad=0.35", "facecolor": WHITE, "edgecolor": palette["grid"], "alpha": 0.9},
    )
    return fig


def main() -> None:
    FIGURES.mkdir(parents=True, exist_ok=True)
    PROTOTYPES.mkdir(parents=True, exist_ok=True)
    df = load_data()
    prototypes = (
        ("prototype01_risk_transition_atlas", prototype_risk_transition),
        ("prototype02_ranked_landscape", prototype_ranked_landscape),
        ("prototype03_small_multiple_strips", prototype_small_multiples),
        ("prototype04_bivariate_scatter", prototype_bivariate),
        ("prototype05_matrix_tile_atlas", prototype_tile_atlas),
        ("prototype06_hybrid_atlas", prototype_hybrid),
    )
    for stem, builder in prototypes:
        save_prototype(builder(df), stem)
    for grayscale in (False, True):
        palette = configure(grayscale)
        save_final(figure1_examples(df, palette), "figure1_clinical_examples", grayscale)
        save_final(
            hybrid_axes(df, palette, "A calculator result has several quantitative meanings"),
            "figure2_catalogue_atlas",
            grayscale,
        )
        save_final(figure3_precision(df, palette), "figure3_ruleout_precision", grayscale)
    print(f"Built 6 prototypes in {PROTOTYPES}")
    print(f"Built 3 main figures in colour and grayscale PNG/PDF/SVG in {FIGURES}")
    print("Checks: 482 evaluations; 407 calculators; 157 point estimates <2%; 67 supported")


if __name__ == "__main__":
    main()
```
