## Supplementary material for "Meaning of clinical calculator results: a cross-sectional analysis of the MDCalc catalogue": Reproducibility archive: code, data and analysis outputs: results_summary.md

# MDCalc Entropy Analysis Results

Source workbook SHA-256: `02348f2e95039be35943e135d27b12010ba382d589d03623a497baef35dbc514`

The primary analysis contains 482 evaluations representing 407 MDCalc calculators from a catalogue of 847.
Median entropy reduction was 0.093 bits (IQR 0.026-0.210).
Median proportional uncertainty removed was 14.9% (IQR 6.2%-30.5%).
The 95% confidence interval for median proportional uncertainty removed was 12.1%-17.2%, estimated by resampling calculators and keeping repeated evaluations together.
One classification supplied more than 60% of average information in 329 evaluations; 173 were positive-result dominant, 156 negative-result dominant, and 153 balanced.
Among 56 rule-out evaluations, the median negative-result information share was 71.3% (95% confidence interval 65.6%-79.0%).
Median proportional uncertainty removed declined from 25.4% in the smallest-study quartile to 6.2% in the largest-study quartile.
After Miller–Madow correction, median information gain was 0.092 bits and median proportional uncertainty removed was 14.4%.
The corrected study-size correlation was -0.438; after adjustment for baseline uncertainty it was -0.279, and at fixed 10% prevalence it was -0.397.
Across 14 specialty labels represented by at least 50 evaluations, median proportional uncertainty removed ranged from 8.6% to 18.9%.
Excluding 2 evaluations explicitly described as case-control or two-gate left a median information gain of 0.092 bits and 14.8% of baseline uncertainty removed.

One source count was corrected with a DOI-linked audit note; no included record retained a sample-size discrepancy greater than 1.
Spearman correlations are primary for monotonic associations with less sensitivity to extreme values; Pearson correlations are secondary.
Conventional P values are omitted because the study is descriptive; confidence intervals resample calculators and keep repeated evaluations together.
