## Supplementary material for "Meaning of clinical calculator results: a cross-sectional analysis of the MDCalc catalogue": Reproducibility archive: code, data and analysis outputs: table1_descriptive_statistics.md

| measure | n | mean | sd | median | q1 | q3 | min | max |
| --- | --- | --- | --- | --- | --- | --- | --- | --- |
| reported_sample_size | 482 | 12554.411 | 220198.621 | 422.0 | 169.25 | 1189.75 | 15.0 | 4830941.0 |
| sensitivity | 482 | 0.752 | 0.236 | 0.824 | 0.652 | 0.931 | 0.0 | 1.0 |
| specificity | 482 | 0.704 | 0.247 | 0.781 | 0.561 | 0.905 | 0.024 | 1.0 |
| prevalence | 482 | 0.258 | 0.228 | 0.176 | 0.07 | 0.406 | 0.006 | 0.962 |
| youden_j | 482 | 0.455 | 0.234 | 0.448 | 0.251 | 0.638 | -0.015 | 1.0 |
| entropy_removed_percent | 482 | 20.921 | 19.371 | 14.887 | 6.206 | 30.486 | 0.0 | 100.0 |
| parent_entropy_bits | 482 | 0.623 | 0.296 | 0.649 | 0.349 | 0.918 | 0.055 | 1.0 |
| entropy_reduction_bits | 482 | 0.15 | 0.165 | 0.093 | 0.026 | 0.21 | 0.0 | 0.763 |
