## Supplementary material for "Meaning of clinical calculator results: a cross-sectional analysis of the MDCalc catalogue": Reproducibility archive: code, data and analysis outputs: table2_clinical_metric_atlas.md

| Clinical decision and calculator | Cohort and starting risk | Accuracy (sensitivity; specificity; J; LR+; LR−) | Risk after positive; negative | Information yield | Dominant result and information share | Why it matters clinically | Source |
| --- | --- | --- | --- | --- | --- | --- | --- |
| *Stopping safely*; **Suspected pulmonary embolism — PERC**; Observed; Low clinical suspicion (<15%) and PERC negative; venous thromboembolism or death within 45 days | n=8,138; Starting risk 7.6% | 97.4%; 21.9%; 0.193; 1.2; 0.12 | 9.3% / 1.0% | 0.0166 bits; 4.29% uncertainty removed | T+ 13.4% / **T− 86.6%** | The combined negative result lowered risk to 1.0%, below the prespecified 2.0% validation target, despite modest average information. | [17] |
| *Stopping safely*; **Minor head injury — Canadian CT Head Rule**; Observed; CCHR positive criteria; clinically important brain injury | n=2,707; Starting risk 8.5% | 100.0%; 41.1%; 0.411; 1.7; 0 | 13.7% / 0.0% | 0.0615 bits; 14.6% uncertainty removed | T+ 21.3% / **T− 78.7%** | The evaluated cohort had no observed clinically important brain injury after a negative classification. | [18] |
| *Stopping safely*; **Upper gastrointestinal bleeding — Glasgow-Blatchford score**; Observed; GBS >1; hospital intervention or death within 30 days | n=2,932; Starting risk 46.3% | 98.6%; 34.6%; 0.332; 1.5; 0.04 | 56.5% / 3.4% | 0.157 bits; 15.8% uncertainty removed | T+ 15.5% / **T− 84.5%** | Negative-result dominance also appears when the outcome is common, not only when it is rare. | [19] |
| *Same pneumonia cohort*; **Community acquired pneumonia — CRB-65**; Observed; CRB-65 ≥1; 30-day mortality | n=553; Starting risk 10.7% | 94.9%; 17.0%; 0.119; 1.1; 0.3 | 12.0% / 3.4% | 0.0093 bits; 1.90% uncertainty removed | T+ 12.0% / **T− 88.0%** | High sensitivity with very low specificity leaves little average information because few patients test negative. | [20] |
| *Same pneumonia cohort*; **Community acquired pneumonia — CURB-65**; Observed; CURB-65 ≥2; 30-day mortality | n=553; Starting risk 10.7% | 84.7%; 40.9%; 0.256; 1.4; 0.37 | 14.6% / 4.3% | 0.0216 bits; 4.41% uncertainty removed | T+ 30.7% / **T− 69.3%** | In the same cohort, added specificity more than doubles information while negative results remain dominant. | [20] |
| *Same HEART score, different thresholds*; **Chest pain — HEART rule-out threshold**; Observed; HEART ≥4; six-week major adverse cardiac events | n=641; Starting risk 9.5% | 100.0%; 69.7%; 0.697; 3.3; 0 | 25.7% / 0.0% | 0.149 bits; 32.9% uncertainty removed | T+ 39.1% / **T− 60.9%** | At the rule-out threshold, the negative result supplies most of the information. | [21] |
| *Same HEART score, different thresholds*; **Chest pain — HEART rule-in threshold**; Observed; HEART >6; six-week major adverse cardiac events | n=641; Starting risk 9.5% | 65.6%; 96.9%; 0.625; 21; 0.36 | 69.0% / 3.6% | 0.169 bits; 37.3% uncertainty removed | **T+ 79.8%** / T− 20.2% | Moving the threshold reverses the pattern: the positive result now carries most of the information. | [21] |
