## Supplementary material for "Meaning of clinical calculator results: a cross-sectional analysis of the MDCalc catalogue": Reproducibility archive: code, data and analysis outputs: table2_correlations.md

| outcome | predictor | method | n | estimate | ci_low | ci_high |
| --- | --- | --- | --- | --- | --- | --- |
| entropy_reduction_bits | sensitivity | spearman | 482 | 0.226 | 0.09 | 0.369 |
| entropy_reduction_bits | sensitivity | pearson | 482 | 0.308 | 0.226 | 0.385 |
| entropy_reduction_bits | specificity | spearman | 482 | 0.441 | 0.246 | 0.589 |
| entropy_reduction_bits | specificity | pearson | 482 | 0.466 | 0.376 | 0.542 |
| entropy_reduction_bits | youden_j | spearman | 482 | 0.855 | 0.82 | 0.88 |
| entropy_reduction_bits | youden_j | pearson | 482 | 0.802 | 0.773 | 0.829 |
| entropy_reduction_bits | prevalence | spearman | 482 | 0.673 | 0.593 | 0.738 |
| entropy_reduction_bits | prevalence | pearson | 482 | 0.516 | 0.437 | 0.593 |
| entropy_reduction_bits | parent_entropy_bits | spearman | 482 | 0.686 | 0.611 | 0.748 |
| entropy_reduction_bits | parent_entropy_bits | pearson | 482 | 0.58 | 0.526 | 0.63 |
| entropy_reduction_bits | reported_sample_size | spearman | 482 | -0.583 | -0.658 | -0.495 |
| entropy_reduction_bits | reported_sample_size | pearson | 482 | -0.049 | -0.24 | -0.045 |
| entropy_removed_percent | sensitivity | spearman | 482 | 0.294 | 0.172 | 0.411 |
| entropy_removed_percent | sensitivity | pearson | 482 | 0.37 | 0.297 | 0.439 |
| entropy_removed_percent | specificity | spearman | 482 | 0.494 | 0.315 | 0.632 |
| entropy_removed_percent | specificity | pearson | 482 | 0.516 | 0.42 | 0.594 |
| entropy_removed_percent | youden_j | spearman | 482 | 0.971 | 0.964 | 0.976 |
| entropy_removed_percent | youden_j | pearson | 482 | 0.916 | 0.907 | 0.927 |
| entropy_removed_percent | prevalence | spearman | 482 | 0.417 | 0.31 | 0.508 |
| entropy_removed_percent | prevalence | pearson | 482 | 0.338 | 0.232 | 0.438 |
| entropy_removed_percent | parent_entropy_bits | spearman | 482 | 0.4 | 0.297 | 0.489 |
| entropy_removed_percent | parent_entropy_bits | pearson | 482 | 0.334 | 0.232 | 0.424 |
| entropy_removed_percent | reported_sample_size | spearman | 482 | -0.466 | -0.544 | -0.373 |
| entropy_removed_percent | reported_sample_size | pearson | 482 | -0.05 | -0.228 | -0.046 |
