## Supplementary material for "Meaning of clinical calculator results: a cross-sectional analysis of the MDCalc catalogue": Reproducibility archive: code, data and analysis outputs: table3_prevalence_shift_experiment.md

| Clinical decision and calculator | Cohort and starting risk | Accuracy (sensitivity; specificity; J; LR+; LR−) | Risk after positive; negative | Information yield | Dominant result and information share | Why it matters clinically | Source |
| --- | --- | --- | --- | --- | --- | --- | --- |
| *Prevalence-shift experiment*; **Same illustrative threshold: 40% prevalence**; Illustrative; Per 1000; sensitivity 90% and specificity 60% | n=1,000; Starting risk 40.0% | 90.0%; 60.0%; 0.500; 2.2; 0.17 | 60.0% / 10.0% | 0.201 bits; 20.7% uncertainty removed | T+ 35.0% / **T− 65.0%** | This higher-prevalence population establishes the reference amount and direction of information. | Constructed example |
| *Prevalence-shift experiment*; **Same illustrative threshold: 4% prevalence**; Illustrative; Per 1000; sensitivity 90% and specificity 60% | n=1,000; Starting risk 4.0% | 90.0%; 60.0%; 0.500; 2.2; 0.17 | 8.6% / 0.7% | 0.0306 bits; 12.6% uncertainty removed | T+ 41.1% / T− 58.9% | Lower starting risk cuts information per patient by 84.8% despite unchanged sensitivity and specificity. | Constructed example |
