## Supplementary material for "Meaning of clinical calculator results: a cross-sectional analysis of the MDCalc catalogue": Reproducibility archive: code, data and analysis outputs: table3_sensitivity_analyses.md

| analysis | evaluations | calculators | median_bits | q1_bits | q3_bits | median_percent | q1_percent | q3_percent | rho_youden_bits |
| --- | --- | --- | --- | --- | --- | --- | --- | --- | --- |
| Primary analysis | 482 | 407 | 0.093 | 0.026 | 0.21 | 14.887 | 6.206 | 30.486 | 0.855 |
| Including 7 other tools or strategies | 489 | 411 | 0.092 | 0.026 | 0.209 | 14.679 | 6.352 | 30.494 | 0.854 |
| One evaluation per calculator | 407 | 407 | 0.1 | 0.03 | 0.228 | 15.703 | 7.668 | 31.106 | 0.839 |
| Whole-number counts only | 480 | 406 | 0.092 | 0.026 | 0.209 | 14.757 | 6.201 | 30.47 | 0.854 |
| High or moderate-high confidence | 187 | 180 | 0.077 | 0.022 | 0.203 | 13.004 | 6.325 | 28.465 | 0.842 |
| Evaluations based on reported counts | 331 | 325 | 0.092 | 0.026 | 0.208 | 14.659 | 6.466 | 30.559 | 0.832 |
| Excluding explicitly case-control/two-gate sources | 480 | 405 | 0.092 | 0.026 | 0.208 | 14.757 | 6.201 | 30.47 | 0.854 |
