## Supplementary material for "Meaning of clinical calculator results: a cross-sectional analysis of the MDCalc catalogue": Reproducibility archive: code, data and analysis outputs: table4_same_youden_experiment.md

| Clinical decision and calculator | Cohort and starting risk | Accuracy (sensitivity; specificity; J; LR+; LR−) | Risk after positive; negative | Information yield | Dominant result and information share | Why it matters clinically | Source |
| --- | --- | --- | --- | --- | --- | --- | --- |
| *Same-prevalence, same-Youden experiment*; **Rule-out emphasis: 10% prevalence**; Illustrative; Per 10,000; sensitivity 95% and specificity 55% | n=10,000; Starting risk 10.0% | 95.0%; 55.0%; 0.500; 2.1; 0.091 | 19.0% / 1.0% | 0.0779 bits; 16.6% uncertainty removed | T+ 33.9% / **T− 66.1%** | The same J can concentrate information in negative results. | Constructed example |
| *Same-prevalence, same-Youden experiment*; **Balanced accuracy: 10% prevalence**; Illustrative; Per 10,000; sensitivity 75% and specificity 75% | n=10,000; Starting risk 10.0% | 75.0%; 75.0%; 0.500; 3; 0.33 | 25.0% / 3.6% | 0.0700 bits; 14.9% uncertainty removed | T+ 57.1% / T− 42.9% | Balancing sensitivity and specificity changes both the amount and direction of information. | Constructed example |
| *Same-prevalence, same-Youden experiment*; **Rule-in emphasis: 10% prevalence**; Illustrative; Per 10,000; sensitivity 55% and specificity 95% | n=10,000; Starting risk 10.0% | 55.0%; 95.0%; 0.500; 11; 0.47 | 55.0% / 5.0% | 0.112 bits; 23.9% uncertainty removed | **T+ 80.6%** / T− 19.4% | The same J can produce more information concentrated in positive results. | Constructed example |
