## Supplementary material for "Meaning of clinical calculator results: a cross-sectional analysis of the MDCalc catalogue": Reproducibility archive: code, data and analysis outputs: table4_sample_size_analyses.md

| analysis | evaluations | calculators | minimum_n | maximum_n | median_n | median_bits | median_percent |
| --- | --- | --- | --- | --- | --- | --- | --- |
| Sample-size quartile Q1 (smallest) | 121 | 115 | 15.0 | 169.0 | 98.0 | 0.203 | 25.394 |
| Sample-size quartile Q2 | 120 | 110 | 170.0 | 421.0 | 284.0 | 0.119 | 18.734 |
| Sample-size quartile Q3 | 120 | 103 | 423.0 | 1168.0 | 620.5 | 0.065 | 13.624 |
| Sample-size quartile Q4 (largest) | 121 | 97 | 1197.0 | 4830941.0 | 3776.0 | 0.017 | 6.205 |
| Reported sample size >= 100 | 418 | 348 | 100.0 | 4830941.0 | 503.0 | 0.072 | 13.002 |
| Reported sample size >= 200 | 347 | 284 | 200.0 | 4830941.0 | 664.0 | 0.058 | 11.62 |
| Reported sample size >= 500 | 211 | 172 | 502.0 | 4830941.0 | 1551.0 | 0.027 | 8.349 |
| Reported sample size >= 1000 | 138 | 110 | 1000.0 | 4830941.0 | 2674.5 | 0.017 | 6.592 |
| Reported sample size >= 5000 | 49 | 33 | 5099.0 | 4830941.0 | 10507.0 | 0.01 | 4.024 |
