## Supplementary material for "Meaning of clinical calculator results: a cross-sectional analysis of the MDCalc catalogue": Reproducibility archive: code, data and analysis outputs: table5_purpose_sample_size_correlations.md

| purpose | evaluations | calculators | rho_sample_size_percent | ci_low | ci_high |
| --- | --- | --- | --- | --- | --- |
| Diagnosis | 200 | 169 | -0.466 | -0.57 | -0.342 |
| Prognosis | 283 | 238 | -0.459 | -0.555 | -0.332 |
| Rule Out | 56 | 43 | -0.307 | -0.589 | -0.013 |
| Treatment | 36 | 33 | -0.584 | -0.764 | -0.316 |
