## Supplementary material for "Meaning of clinical calculator results: a cross-sectional analysis of the MDCalc catalogue": Reproducibility archive: code, data and analysis outputs: tableS2_evidence_audit.md

| audit_item | evaluations_meeting | evaluations_assessed | percent |
| --- | --- | --- | --- |
| Count-based source evidence | 331 | 482 | 68.672 |
| High or moderate-high confidence label | 187 | 482 | 38.797 |
| Study citation recorded | 417 | 482 | 86.515 |
| Threshold description recorded | 411 | 482 | 85.27 |
| Complete, non-negative 2x2 cells | 482 | 482 | 100.0 |
| Reported sample size within one of 2x2 total | 482 | 482 | 100.0 |
| Finite information estimates | 482 | 482 | 100.0 |
| Positive and negative contributions reproduce total information | 482 | 482 | 100.0 |
