## Supplementary material for "Meaning of clinical calculator results: a cross-sectional analysis of the MDCalc catalogue": Reproducibility archive: code, data and analysis outputs: tableS8_prevalence_standardization.md

| fixed_prevalence | median_bits | q1_bits | q3_bits | median_percent | q1_percent | q3_percent | rho_observed_vs_standardized_percent |
| --- | --- | --- | --- | --- | --- | --- | --- |
| 0.05 | 0.036 | 0.015 | 0.072 | 12.549 | 5.228 | 25.145 | 0.983 |
| 0.1 | 0.067 | 0.029 | 0.135 | 14.228 | 6.084 | 28.69 | 0.985 |
| 0.25 | 0.135 | 0.059 | 0.267 | 16.673 | 7.253 | 32.872 | 0.985 |
| 0.5 | 0.178 | 0.078 | 0.342 | 17.789 | 7.819 | 34.176 | 0.979 |
