## Supplementary material for "Meaning of clinical calculator results: a cross-sectional analysis of the MDCalc catalogue": Reproducibility archive: code, data and analysis outputs: tableS9_catalog_representation.md

| purpose | catalog_calculators | represented_calculators | representation_percent | primary_evaluations |
| --- | --- | --- | --- | --- |
| Diagnosis | 296 | 169 | 57.095 | 200 |
| Prognosis | 475 | 238 | 50.105 | 283 |
| Rule Out | 44 | 43 | 97.727 | 56 |
| Treatment | 85 | 33 | 38.824 | 36 |
