## Supplementary material for "Meaning of clinical calculator results: a cross-sectional analysis of the MDCalc catalogue": Reproducibility archive: code, data and analysis outputs: tableS10_headline_cluster_bootstrap.md

| measure | estimate | ci_low | ci_high |
| --- | --- | --- | --- |
| Median entropy reduction, bits | 0.093 | 0.065 | 0.108 |
| Median proportional uncertainty removed, % | 14.887 | 12.078 | 17.152 |
| Evaluations removing at least 10%, % | 62.033 | 55.846 | 67.804 |
| Evaluations removing at least 25%, % | 31.12 | 26.087 | 36.345 |
| Evaluations removing at least 50%, % | 9.959 | 7.202 | 13.111 |
| Median negative-result information share among rule-out evaluations, % | 71.299 | 65.602 | 79.05 |
