## Supplementary material for "Meaning of clinical calculator results: a cross-sectional analysis of the MDCalc catalogue": Reproducibility archive: code, data and analysis outputs: tableS11_specialty_comparison.md

| specialty | evaluations | calculators | median_prevalence_percent | median_removed_percent | removed_ci_low | removed_ci_high | median_fixed_10_percent | fixed_10_ci_low | fixed_10_ci_high | median_positive_share_percent | rho_sample_size_percent | rho_ci_low | rho_ci_high |
| --- | --- | --- | --- | --- | --- | --- | --- | --- | --- | --- | --- | --- | --- |
| Neurology | 70 | 62 | 18.306 | 18.874 | 13.39 | 22.828 | 16.39 | 13.785 | 21.757 | 43.449 | -0.543 | -0.703 | -0.345 |
| Primary Care | 124 | 115 | 21.497 | 17.075 | 13.964 | 23.022 | 18.106 | 13.237 | 22.901 | 52.774 | -0.399 | -0.533 | -0.249 |
| Surgery (General) | 73 | 69 | 16.898 | 16.727 | 11.514 | 20.22 | 15.708 | 11.424 | 17.743 | 47.839 | -0.525 | -0.676 | -0.315 |
| Family Practice | 173 | 156 | 21.831 | 16.129 | 13.385 | 20.925 | 15.599 | 12.335 | 21.051 | 55.661 | -0.451 | -0.56 | -0.328 |
| Gastroenterology | 61 | 54 | 25.581 | 15.703 | 11.901 | 18.862 | 14.542 | 10.418 | 19.067 | 54.135 | -0.348 | -0.534 | -0.097 |
| Infectious Disease | 51 | 47 | 25.0 | 15.591 | 12.775 | 21.915 | 15.326 | 10.915 | 21.404 | 49.357 | -0.476 | -0.678 | -0.192 |
| Internal Medicine | 320 | 277 | 19.535 | 14.984 | 12.124 | 18.968 | 14.451 | 11.523 | 16.938 | 52.841 | -0.435 | -0.525 | -0.329 |
| Hospitalist Medicine | 261 | 232 | 16.517 | 14.39 | 11.62 | 18.329 | 13.223 | 10.915 | 17.127 | 50.631 | -0.456 | -0.545 | -0.34 |
| Emergency Medicine | 298 | 242 | 13.202 | 12.901 | 9.597 | 15.591 | 12.341 | 9.878 | 15.826 | 48.192 | -0.531 | -0.609 | -0.434 |
| Critical Care | 224 | 183 | 12.605 | 11.645 | 8.286 | 16.341 | 11.745 | 8.664 | 15.654 | 52.607 | -0.511 | -0.604 | -0.385 |
| Hematology and Oncology | 75 | 66 | 18.043 | 11.565 | 8.172 | 17.115 | 10.311 | 7.595 | 15.997 | 50.906 | -0.454 | -0.629 | -0.24 |
| Pulmonology | 87 | 66 | 14.894 | 10.967 | 8.214 | 17.225 | 10.498 | 7.363 | 15.947 | 60.072 | -0.505 | -0.669 | -0.144 |
| Cardiology | 126 | 89 | 10.078 | 9.109 | 6.567 | 17.126 | 10.016 | 6.252 | 15.499 | 44.964 | -0.438 | -0.561 | -0.29 |
| Surgery (Trauma) | 54 | 36 | 6.11 | 8.578 | 5.793 | 16.341 | 10.434 | 6.285 | 19.732 | 61.758 | -0.74 | -0.838 | -0.564 |
