## Supplementary material for "Meaning of clinical calculator results: a cross-sectional analysis of the MDCalc catalogue": Reproducibility archive: code, data and analysis outputs: tableS12_clinical_metric_atlas.md

| Clinical decision and calculator | Cohort and starting risk | Accuracy (sensitivity; specificity; J; LR+; LR−) | Risk after positive; negative | Information yield | Dominant result and information share | Why it matters clinically | Source |
| --- | --- | --- | --- | --- | --- | --- | --- |
| *Stopping safely*; **Suspected pulmonary embolism — PERC**; Observed; Low clinical suspicion (<15%) and PERC negative; venous thromboembolism or death within 45 days | n=8,138; Starting risk 7.6% | 97.4%; 21.9%; 0.193; 1.2; 0.12 | 9.3% / 1.0% | 0.0166 bits; 4.29% uncertainty removed | T+ 13.4% / **T− 86.6%** | The combined negative result lowered risk to 1.0%, below the prespecified 2.0% validation target, despite modest average information. | [17] |
| *Stopping safely*; **Minor head injury — Canadian CT Head Rule**; Observed; CCHR positive criteria; clinically important brain injury | n=2,707; Starting risk 8.5% | 100.0%; 41.1%; 0.411; 1.7; 0 | 13.7% / 0.0% | 0.0615 bits; 14.6% uncertainty removed | T+ 21.3% / **T− 78.7%** | The evaluated cohort had no observed clinically important brain injury after a negative classification. | [18] |
| *Stopping safely*; **Upper gastrointestinal bleeding — Glasgow-Blatchford score**; Observed; GBS >1; hospital intervention or death within 30 days | n=2,932; Starting risk 46.3% | 98.6%; 34.6%; 0.332; 1.5; 0.04 | 56.5% / 3.4% | 0.157 bits; 15.8% uncertainty removed | T+ 15.5% / **T− 84.5%** | Negative-result dominance also appears when the outcome is common, not only when it is rare. | [19] |
| *Same pneumonia cohort*; **Community acquired pneumonia — CRB-65**; Observed; CRB-65 ≥1; 30-day mortality | n=553; Starting risk 10.7% | 94.9%; 17.0%; 0.119; 1.1; 0.3 | 12.0% / 3.4% | 0.0093 bits; 1.90% uncertainty removed | T+ 12.0% / **T− 88.0%** | High sensitivity with very low specificity leaves little average information because few patients test negative. | [20] |
| *Same pneumonia cohort*; **Community acquired pneumonia — CURB-65**; Observed; CURB-65 ≥2; 30-day mortality | n=553; Starting risk 10.7% | 84.7%; 40.9%; 0.256; 1.4; 0.37 | 14.6% / 4.3% | 0.0216 bits; 4.41% uncertainty removed | T+ 30.7% / **T− 69.3%** | In the same cohort, added specificity more than doubles information while negative results remain dominant. | [20] |
| *Same HEART score, different thresholds*; **Chest pain — HEART rule-out threshold**; Observed; HEART ≥4; six-week major adverse cardiac events | n=641; Starting risk 9.5% | 100.0%; 69.7%; 0.697; 3.3; 0 | 25.7% / 0.0% | 0.149 bits; 32.9% uncertainty removed | T+ 39.1% / **T− 60.9%** | At the rule-out threshold, the negative result supplies most of the information. | [21] |
| *Same HEART score, different thresholds*; **Chest pain — HEART rule-in threshold**; Observed; HEART >6; six-week major adverse cardiac events | n=641; Starting risk 9.5% | 65.6%; 96.9%; 0.625; 21; 0.36 | 69.0% / 3.6% | 0.169 bits; 37.3% uncertainty removed | **T+ 79.8%** / T− 20.2% | Moving the threshold reverses the pattern: the positive result now carries most of the information. | [21] |
| *Positive, balanced, and rare-outcome patterns*; **Pharyngitis — modified Centor/McIsaac score**; Observed; Score ≥2; group A streptococcal pharyngitis | n=621; Starting risk 16.1% | 85.0%; 92.1%; 0.771; 11; 0.16 | 67.5% / 3.0% | 0.296 bits; 46.5% uncertainty removed | **T+ 65.1%** / T− 34.9% | The positive result dominates, a different information pattern from the rule-out tools. | See source ledger |
| *Positive, balanced, and rare-outcome patterns*; **COVID-19 — 4C mortality score**; Observed; Score >9; in-hospital mortality | n=1,493; Starting risk 38.0% | 70.5%; 74.0%; 0.445; 2.7; 0.4 | 62.4% / 19.6% | 0.140 bits; 14.7% uncertainty removed | T+ 53.7% / T− 46.3% | Positive and negative results contribute almost equally, showing that some scores genuinely inform both ways. | See source ledger |
| *Positive, balanced, and rare-outcome patterns*; **Atrial fibrillation — HAS-BLED**; Observed; Score ≥3; one-year major bleeding | n=3,071; Starting risk 1.6% | 25.0%; 92.4%; 0.174; 3.3; 0.81 | 4.9% / 1.3% | 0.0031 bits; 2.66% uncertainty removed | **T+ 87.4%** / T− 12.6% | A 98.7% NPV coexists with 0.003 bits per patient because major bleeding was rare before scoring. | See source ledger |
| *Additional rule-out and early-disposition tools*; **Chest pain — ADAPT protocol**; Observed; ADAPT positive; major adverse cardiac events | n=1,975; Starting risk 15.3% | 99.7%; 23.4%; 0.230; 1.3; 0.014 | 19.0% / 0.3% | 0.0495 bits; 8.02% uncertainty removed | T+ 11.8% / **T− 88.2%** | Near-perfect sensitivity and modest specificity concentrate most information in the negative result. | See source ledger |
| *Additional rule-out and early-disposition tools*; **Child with cerebrospinal fluid pleocytosis — Bacterial Meningitis Score**; Observed; Score >0; bacterial meningitis | n=3,295; Starting risk 3.7% | 98.3%; 53.9%; 0.523; 2.1; 0.031 | 7.5% / 0.1% | 0.0354 bits; 15.6% uncertainty removed | T+ 31.8% / **T− 68.2%** | The very-low-risk negative result carries most information despite a low event frequency. | See source ledger |
| *Additional rule-out and early-disposition tools*; **Acute vestibular syndrome — HINTS**; Observed; Any dangerous HINTS sign; central stroke | n=101; Starting risk 75.2% | 100.0%; 96.0%; 0.960; 25; 0 | 98.7% / 0.0% | 0.731 bits; 90.6% uncertainty removed | T+ 34.5% / **T− 65.5%** | High sensitivity and specificity remove most available uncertainty, with the negative examination contributing more. | See source ledger |
| *Additional rule-out and early-disposition tools*; **Blunt trauma — NEXUS cervical spine criteria**; Observed; Any NEXUS criterion; clinically important cervical spine injury | n=34,069; Starting risk 2.4% | 99.0%; 12.9%; 0.120; 1.1; 0.076 | 2.7% / 0.2% | 0.0035 bits; 2.13% uncertainty removed | T+ 7.6% / **T− 92.4%** | Near-perfect sensitivity coexists with little average yield because few patients satisfy every low-risk criterion. | See source ledger |
| *Additional rule-out and early-disposition tools*; **Ankle injury — Ottawa Ankle Rule**; Observed; Any ankle-zone criterion; fracture | n=559; Starting risk 12.0% | 100.0%; 23.8%; 0.238; 1.3; 0 | 15.2% / 0.0% | 0.0436 bits; 8.25% uncertainty removed | T+ 11.6% / **T− 88.4%** | Perfect sensitivity here is clinically directional, while low specificity limits how many avoid imaging. | See source ledger |
| *Additional rule-out and early-disposition tools*; **Acute headache — Ottawa SAH Rule**; Observed; Any Ottawa SAH criterion; subarachnoid haemorrhage | n=1,999; Starting risk 6.5% | 100.0%; 15.3%; 0.153; 1.2; 0 | 7.6% / 0.0% | 0.0150 bits; 4.33% uncertainty removed | T+ 7.6% / **T− 92.4%** | Perfect sensitivity here removes modest average uncertainty because informative negative results are uncommon. | See source ledger |
| *Additional rule-out and early-disposition tools*; **Paediatric head injury — PECARN**; Observed; Any age-specific predictor; clinically important traumatic brain injury | n=2,428; Starting risk 0.8% | 100.0%; 54.5%; 0.545; 2.2; 0 | 1.7% / 0.0% | 0.0088 bits; 13.4% uncertainty removed | T+ 30.7% / **T− 69.3%** | With injury uncommon, a perfect-sensitivity rule still removes 13% of available uncertainty. | See source ledger |
| *Positive-result and balanced tools*; **Non-invasive ventilation — HACOR**; Observed; HACOR >5 at one hour; non-invasive ventilation failure | n=358; Starting risk 39.4% | 75.9%; 92.6%; 0.685; 10; 0.26 | 87.0% / 14.5% | 0.384 bits; 39.7% uncertainty removed | **T+ 63.1%** / T− 36.9% | A high-specificity threshold makes ventilation failure a positive-result-dominant pattern. | See source ledger |
| *Positive-result and balanced tools*; **Chest pain — HEART score validation**; Observed; HEART ≥4; major adverse cardiac events | n=2,440; Starting risk 17.7% | 96.5%; 42.6%; 0.391; 1.7; 0.081 | 26.6% / 1.7% | 0.0918 bits; 13.6% uncertainty removed | T+ 24.5% / **T− 75.5%** | A low-risk HEART threshold again concentrates information in the negative result. | See source ledger |
| *Positive-result and balanced tools*; **Left bundle branch block — modified Sgarbossa criteria**; Observed; Any modified criterion; acute coronary occlusion | n=294; Starting risk 15.3% | 80.0%; 98.8%; 0.788; 66; 0.2 | 92.3% / 3.5% | 0.374 bits; 60.7% uncertainty removed | **T+ 75.3%** / T− 24.7% | The positive ECG pattern carries three quarters of the information, matching a rule-in task. | See source ledger |
| *Positive-result and balanced tools*; **Pulmonary embolism — PESI**; Observed; PESI >85; short-term mortality | n=357; Starting risk 5.9% | 90.5%; 54.8%; 0.452; 2; 0.17 | 11.1% / 1.1% | 0.0370 bits; 11.5% uncertainty removed | T+ 37.1% / **T− 62.9%** | The negative result supplies most information for identifying low-risk pulmonary embolism. | See source ledger |
| *Positive-result and balanced tools*; **Suspected infection — qSOFA**; Observed; qSOFA ≥2; in-hospital mortality | n=879; Starting risk 8.4% | 70.3%; 79.4%; 0.496; 3.4; 0.37 | 23.9% / 3.3% | 0.0618 bits; 14.8% uncertainty removed | **T+ 62.4%** / T− 37.6% | Positive results contribute most information, consistent with escalation rather than exclusion. | See source ledger |
| *Positive-result and balanced tools*; **COVID-19 — quick COVID-19 Severity Index**; Observed; qCSI >3; respiratory decompensation within 24 hours | n=299; Starting risk 13.4% | 80.0%; 78.0%; 0.580; 3.6; 0.26 | 36.0% / 3.8% | 0.123 bits; 21.7% uncertainty removed | T+ 56.5% / T− 43.5% | Positive and negative results contribute nearly evenly to early deterioration prediction. | See source ledger |
| *Positive-result and balanced tools*; **Flank pain — STONE score**; Observed; STONE 10–13; uncomplicated ureteral stone | n=845; Starting risk 39.2% | 53.2%; 87.2%; 0.403; 4.1; 0.54 | 72.7% / 25.7% | 0.137 bits; 14.2% uncertainty removed | **T+ 69.7%** / T− 30.3% | Positive results carry most information, but neither result alone determines whether imaging is needed. | See source ledger |
