## Supplementary material for "Meaning of clinical calculator results: a cross-sectional analysis of the MDCalc catalogue": Reproducibility archive: code, data and analysis outputs: tableS12_clinical_metric_atlas_source_ledger.md

| source_index | clinical_case | study_citation | outcome_and_threshold | tp | fp | fn | tn | mdcalc_url |
| --- | --- | --- | --- | --- | --- | --- | --- | --- |
| 241 | Suspected pulmonary embolism — PERC | Kline JA, Courtney DM, Kabrhel C, et al. Prospective multicenter evaluation of the pulmonary embolism rule-out criteria. J Thromb Haemost. 2008;6(5):772-780. doi:10.1111/j.1538-7836.2008.02944.x | Low clinical suspicion (<15%) and PERC negative; venous thromboembolism or death within 45 days | 601.0 | 5871.0 | 16.0 | 1650.0 | https://www.mdcalc.com/calc/347/perc-rule-pulmonary-embolism |
| 65 | Minor head injury — Canadian CT Head Rule | Stiell IG, Clement CM, Rowe BH, et al. Comparison of the Canadian CT Head Rule and the New Orleans Criteria in patients with minor head injury. JAMA. 2005;294(12):1511-1518. doi:10.1001/jama.294.12.1511 | CCHR positive criteria; clinically important brain injury | 231.0 | 1458.0 | 0.0 | 1018.0 | https://www.mdcalc.com/calc/608/canadian-ct-head-injury-trauma-rule |
| 145 | Upper gastrointestinal bleeding — Glasgow-Blatchford score | Stanley AJ, Laine L, Dalton HR, et al. Comparison of risk scoring systems for patients presenting with upper gastrointestinal bleeding: international multicentre prospective study. BMJ. 2017;356:i6432. doi:10.1136/bmj.i6432 | GBS >1; hospital intervention or death within 30 days | 1338.0 | 1030.0 | 19.0 | 545.0 | https://www.mdcalc.com/calc/518/glasgow-blatchford-bleeding-score-gbs |
| 97 | Community acquired pneumonia — CRB-65 | Nüllmann H, Pflug MA, Wesemann T, et al. External validation of the CURSI criteria in adults hospitalised for community-acquired pneumonia. BMC Infect Dis. 2014;14:39. doi:10.1186/1471-2334-14-39 | CRB-65 ≥1; 30-day mortality | 56.0 | 410.0 | 3.0 | 84.0 | https://www.mdcalc.com/calc/10604/crb-65-score-for-pneumonia-severity-crb-65 |
| 101 | Community acquired pneumonia — CURB-65 | Nüllmann H, Pflug MA, Wesemann T, et al. External validation of the CURSI criteria in adults hospitalised for community-acquired pneumonia. BMC Infect Dis. 2014;14:39. doi:10.1186/1471-2334-14-39 | CURB-65 ≥2; 30-day mortality | 50.0 | 292.0 | 9.0 | 202.0 | https://www.mdcalc.com/calc/324/curb-65-score-pneumonia-severity |
| 473 | Chest pain — HEART rule-out threshold | Moumneh T, Richard-Jourjon V, Friou E, et al. Reliability of the CARE rule and the HEART score to rule out an acute coronary syndrome in non-traumatic chest pain patients. Intern Emerg Med. 2018;13(7):1111-1119. doi:10.1007/s11739-018-1803-4 | HEART ≥4; six-week major adverse cardiac events | 61.0 | 176.0 | 0.0 | 404.0 | https://www.mdcalc.com/calc/1752/heart-score-major-cardiac-events |
| 474 | Chest pain — HEART rule-in threshold | Moumneh T, Richard-Jourjon V, Friou E, et al. Reliability of the CARE rule and the HEART score to rule out an acute coronary syndrome in non-traumatic chest pain patients. Intern Emerg Med. 2018;13(7):1111-1119. doi:10.1007/s11739-018-1803-4 | HEART >6; six-week major adverse cardiac events | 40.0 | 18.0 | 21.0 | 562.0 | https://www.mdcalc.com/calc/1752/heart-score-major-cardiac-events |
| 75 | Pharyngitis — modified Centor/McIsaac score | McIsaac WJ, Kellner JD, Aufricht P, Vanjaka A, Low DE. Empirical validation of guidelines for the management of pharyngitis in children and adults. JAMA. 2004;291(13):1587-1595. doi:10.1001/jama.291.13.1587 | Score ≥2; group A streptococcal pharyngitis | 85.0 | 41.0 | 15.0 | 480.0 | https://www.mdcalc.com/calc/104/centor-score-modified-mcisaac-strep-pharyngitis |
| 3 | COVID-19 — 4C mortality score | Aletreby WT, Mumtaz SA, Shahzad SA, et al. External validation of 4C ISARIC Mortality Score in critically ill COVID-19 patients from Saudi Arabia. Saudi J Med Med Sci. 2022;10(1):19-24. doi:10.4103/sjmms.sjmms_480_21 | Score >9; in-hospital mortality | 400.0 | 241.0 | 167.0 | 685.0 | https://www.mdcalc.com/calc/10338/4c-mortality-score-covid-19 |
| 456 | Atrial fibrillation — HAS-BLED | Pisters R, Lane DA, Nieuwlaat R, de Vos CB, Crijns HJGM, Lip GYH. A novel user-friendly score (HAS-BLED) to assess 1-year risk of major bleeding in patients with atrial fibrillation. Chest. 2010;138(5):1093-1100. doi:10.1378/chest.10-0134 | Score ≥3; one-year major bleeding | 12.0 | 231.0 | 36.0 | 2792.0 | https://www.mdcalc.com/calc/807/has-bled-score-major-bleeding-risk |
| 18 | Chest pain — ADAPT protocol | Than et al., 2012, ADAPT Trial | ADAPT positive; major adverse cardiac events | 301.0 | 1282.0 | 1.0 | 391.0 | https://www.mdcalc.com/calc/1857/adapt-protocol-cardiac-event-risk |
| 46 | Child with cerebrospinal fluid pleocytosis — Bacterial Meningitis Score | Nigrovic LE et al. Clinical prediction rule for identifying children with cerebrospinal fluid pleocytosis at very low risk of bacterial meningitis. JAMA. 2007;297:52–60. Multicenter validation across 20 US academic emergency departments. | Score >0; bacterial meningitis | 119.0 | 1462.0 | 2.0 | 1712.0 | https://www.mdcalc.com/calc/3911/bacterial-meningitis-score-children |
| 164 | Acute vestibular syndrome — HINTS | Kattah et al., 2009 | Any dangerous HINTS sign; central stroke | 76.0 | 1.0 | 0.0 | 24.0 | https://www.mdcalc.com/calc/10184/hints-stroke-acute-vestibular-syndrome |
| 220 | Blunt trauma — NEXUS cervical spine criteria | Hoffman et al., NEJM 2000 | Any NEXUS criterion; clinically important cervical spine injury | 810.0 | 28950.0 | 8.0 | 4301.0 | https://www.mdcalc.com/calc/703/nexus-criteria-c-spine-imaging |
| 230 | Ankle injury — Ottawa Ankle Rule | Plint et al., Validation of the Ottawa Ankle Rules in Children With Ankle Injuries, Acad Emerg Med 1999 | Any ankle-zone criterion; fracture | 67.0 | 375.0 | 0.0 | 117.0 | https://www.mdcalc.com/calc/1670/ottawa-ankle-rule |
| 232 | Acute headache — Ottawa SAH Rule | Prospective validation study of Ottawa SAH Rule at 10 sites | Any Ottawa SAH criterion; subarachnoid haemorrhage | 130.0 | 1583.0 | 0.0 | 286.0 | https://www.mdcalc.com/calc/3875/ottawa-subarachnoid-hemorrhage-sah-rule-for-headache-evaluation |
| 234 | Paediatric head injury — PECARN | Schonfeld et al., PECARN Head Injury Clinical Prediction Rules Are Reliable in Practice, Arch Dis Child. 2014 | Any age-specific predictor; clinically important traumatic brain injury | 19.0 | 1096.0 | 0.0 | 1313.0 | https://www.mdcalc.com/calc/589/pecarn-pediatric-head-injury-trauma-algorithm |
| 151 | Non-invasive ventilation — HACOR | Duan et al., 2017—prospective validation cohort | HACOR >5 at one hour; non-invasive ventilation failure | 107.0 | 16.0 | 34.0 | 201.0 | https://www.mdcalc.com/calc/10460/hacor-score |
| 156 | Chest pain — HEART score validation | Backus et al., 2013 | HEART ≥4; major adverse cardiac events | 418.0 | 1152.0 | 15.0 | 855.0 | https://www.mdcalc.com/calc/1752/heart-score-major-cardiac-events |
| 211 | Left bundle branch block — modified Sgarbossa criteria | Meyers HP, Limkakeng AT Jr, Jaffa EJ, et al. Validation of the modified Sgarbossa criteria for acute coronary occlusion in the setting of left bundle branch block: a retrospective case-control study. Am Heart J. 2015;170(6):1255–1264. PMID: 26678648. | Any modified criterion; acute coronary occlusion | 36.0 | 3.0 | 9.0 | 246.0 | https://www.mdcalc.com/calc/10353/modified-sgarbossas-criteria-mi-left-bundle-branch-block |
| 251 | Pulmonary embolism — PESI | Donzé J, Le Gal G, Fine MJ, et al. Prospective validation of the Pulmonary Embolism Severity Index: a clinical prognostic model for pulmonary embolism. Thromb Haemost. 2008;100(5):943–948. PMID: 18989542. doi:10.1160/TH08-05-0285. | PESI >85; short-term mortality | 19.0 | 152.0 | 2.0 | 184.0 | https://www.mdcalc.com/calc/1304/pulmonary-embolism-severity-index-pesi |
| 252 | Suspected infection — qSOFA | Freund Y, Lemachatti N, Krastinova E, et al. Prognostic Accuracy of Sepsis-3 Criteria for In-Hospital Mortality Among Patients With Suspected Infection Presenting to the Emergency Department. JAMA. 2017;317(3):301–308. PMID: 28114554. | qSOFA ≥2; in-hospital mortality | 52.0 | 166.0 | 22.0 | 639.0 | https://www.mdcalc.com/calc/2654/qsofa-quick-sofa-score-sepsis |
| 253 | COVID-19 — quick COVID-19 Severity Index | Haimovich AD, Ravindra NG, Stoytchev S, et al. Development and Validation of the Quick COVID-19 Severity Index: A Prognostic Tool for Early Clinical Decompensation. Ann Emerg Med. 2020;76(4):442–453. doi:10.1016/j.annemergmed.2020.07.022. | qCSI >3; respiratory decompensation within 24 hours | 32.0 | 57.0 | 8.0 | 202.0 | https://www.mdcalc.com/calc/10304/quick-covid-19-severity-index-qcsi |
| 399 | Flank pain — STONE score | Wang et al., External Validation of the STONE Score, a Clinical Prediction Rule for Ureteral Stone: An Observational Multi-institutional Study | STONE 10–13; uncomplicated ureteral stone | 176.0 | 66.0 | 155.0 | 448.0 | https://www.mdcalc.com/calc/10110/stone-score-uncomplicated-ureteral-stone |
