## Supplementary material for "Meaning of clinical calculator results: a cross-sectional analysis of the MDCalc catalogue": Reproducibility archive: code, data and analysis outputs: tableS13_study_size_bias_robustness.md

| analysis | evaluations | estimate | ci_low | ci_high |
| --- | --- | --- | --- | --- |
| Observed information gain, bits | 482 | -0.583 | -0.658 | -0.495 |
| Miller–Madow corrected information gain, bits | 482 | -0.563 | -0.639 | -0.471 |
| Observed proportional uncertainty removed | 482 | -0.466 | -0.544 | -0.373 |
| Miller–Madow corrected proportional uncertainty removed | 482 | -0.438 | -0.518 | -0.342 |
| Corrected proportional uncertainty removed, adjusted for baseline uncertainty | 482 | -0.279 | -0.363 | -0.18 |
| Proportional uncertainty removed, standardised to 10% outcome prevalence | 482 | -0.397 | -0.477 | -0.302 |
| Corrected proportional uncertainty removed within prevalence Q1 lowest | 121 | -0.358 | -0.528 | -0.165 |
| Corrected proportional uncertainty removed within prevalence Q2 | 120 | -0.327 | -0.48 | -0.162 |
| Corrected proportional uncertainty removed within prevalence Q3 | 120 | -0.352 | -0.505 | -0.183 |
| Corrected proportional uncertainty removed within prevalence Q4 highest | 121 | -0.002 | -0.17 | 0.178 |
