## Supplementary material for "Meaning of clinical calculator results: a cross-sectional analysis of the MDCalc catalogue": Reproducibility archive: code, data and analysis outputs: tableS14_sampling_design_sensitivity.md

| sampling_design | evaluations | calculators | median_bits | median_percent |
| --- | --- | --- | --- | --- |
| Cohort/consecutive | 21 | 21 | 0.037 | 11.323 |
| Case-control/two-gate | 2 | 2 | 0.313 | 42.895 |
| Unclear | 459 | 387 | 0.094 | 15.133 |
