## Supplementary figures and images for "Meaning of clinical calculator results: a cross-sectional analysis of the MDCalc catalogue"

### figure1_clinical_examples.pdf

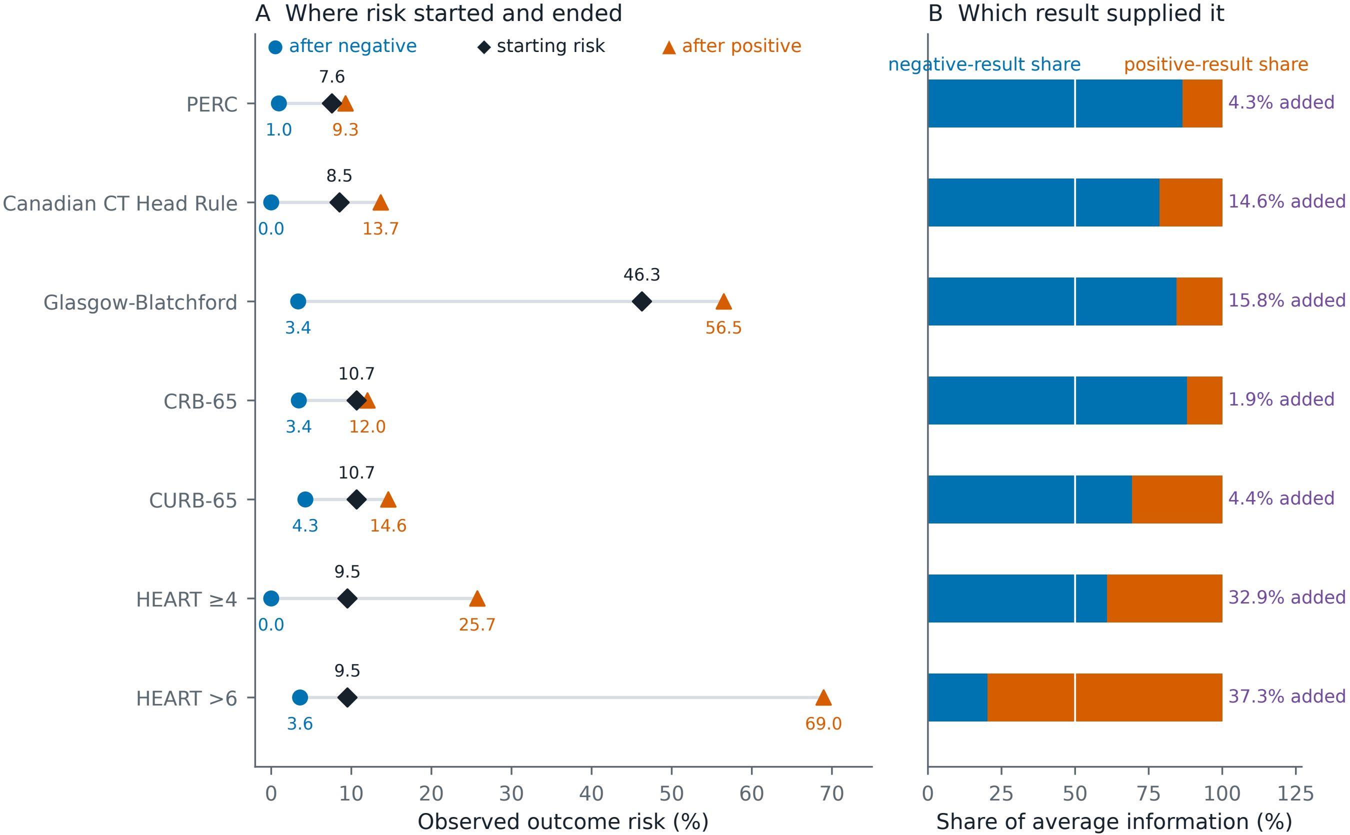

### figure1_clinical_examples.png

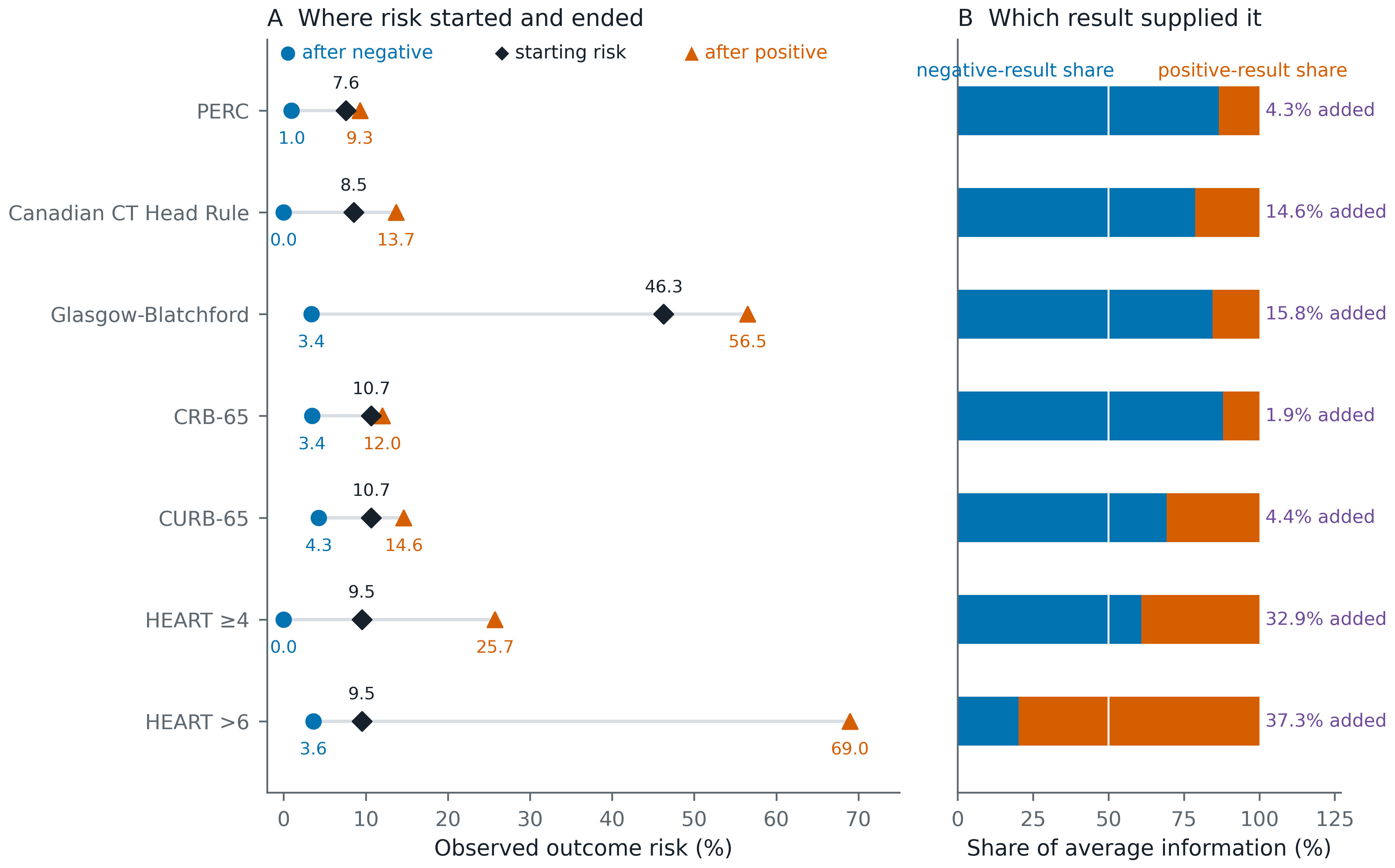

### figure1_clinical_examples_grayscale.pdf

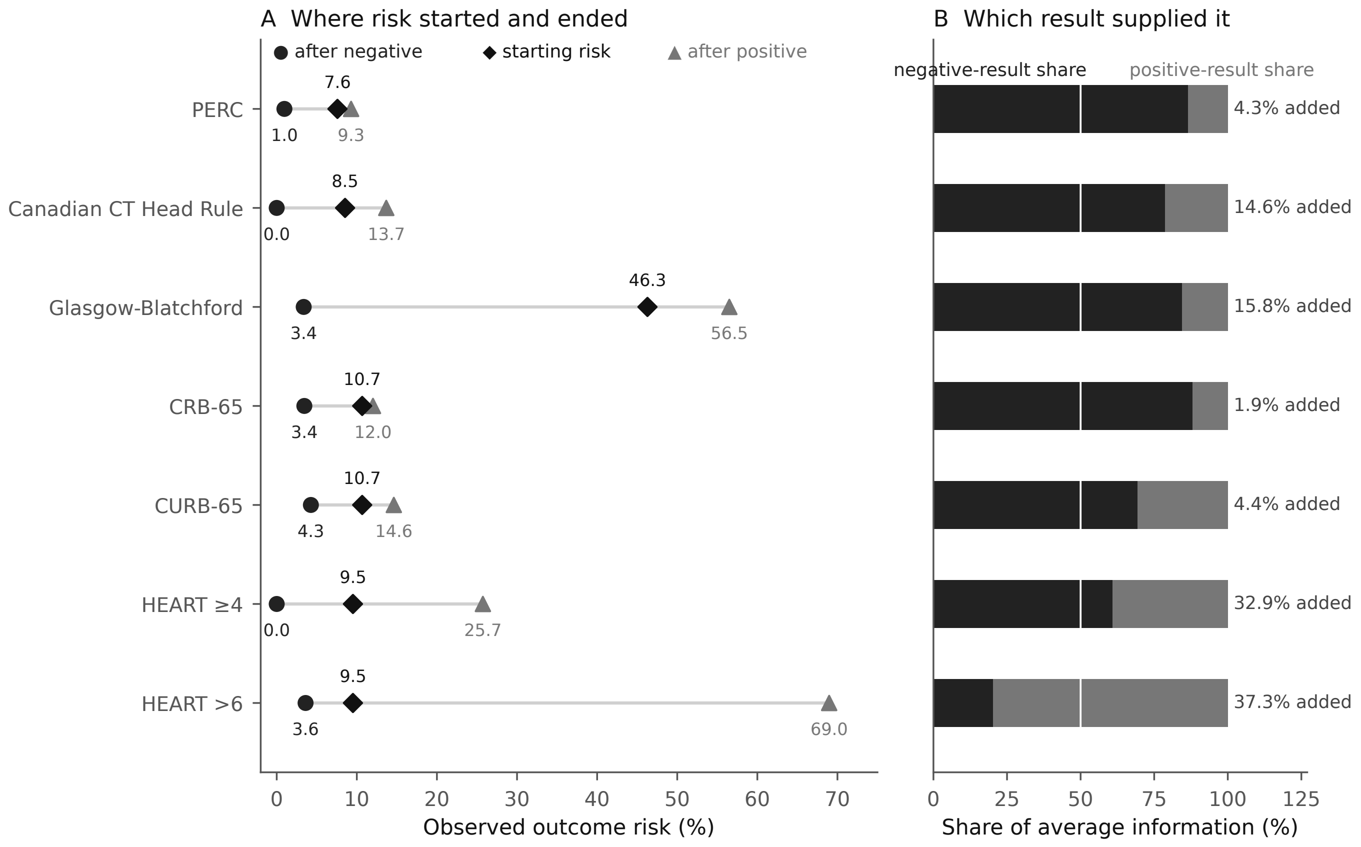

### figure1_clinical_examples_grayscale.png

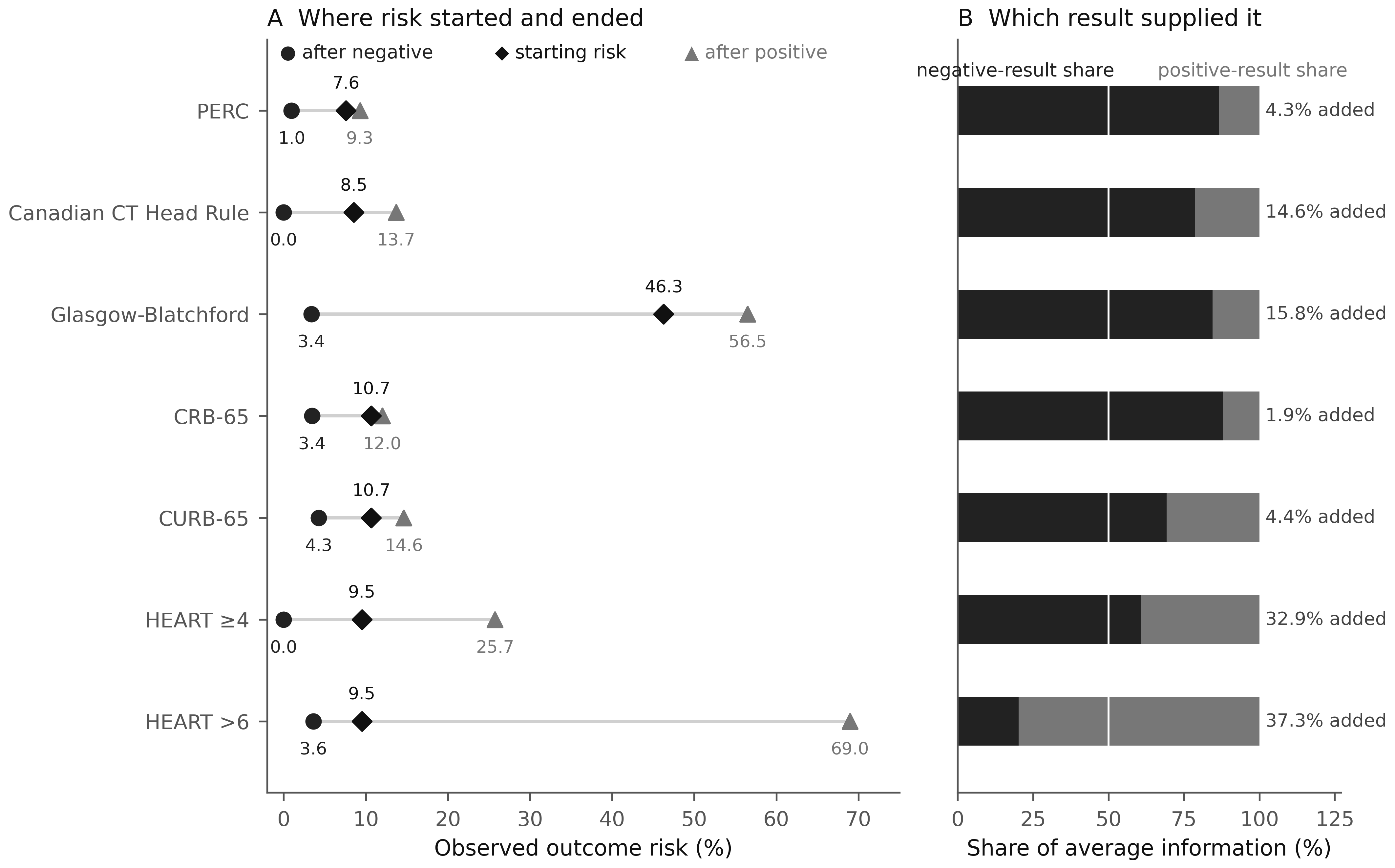

### figure1_selection_flow.pdf

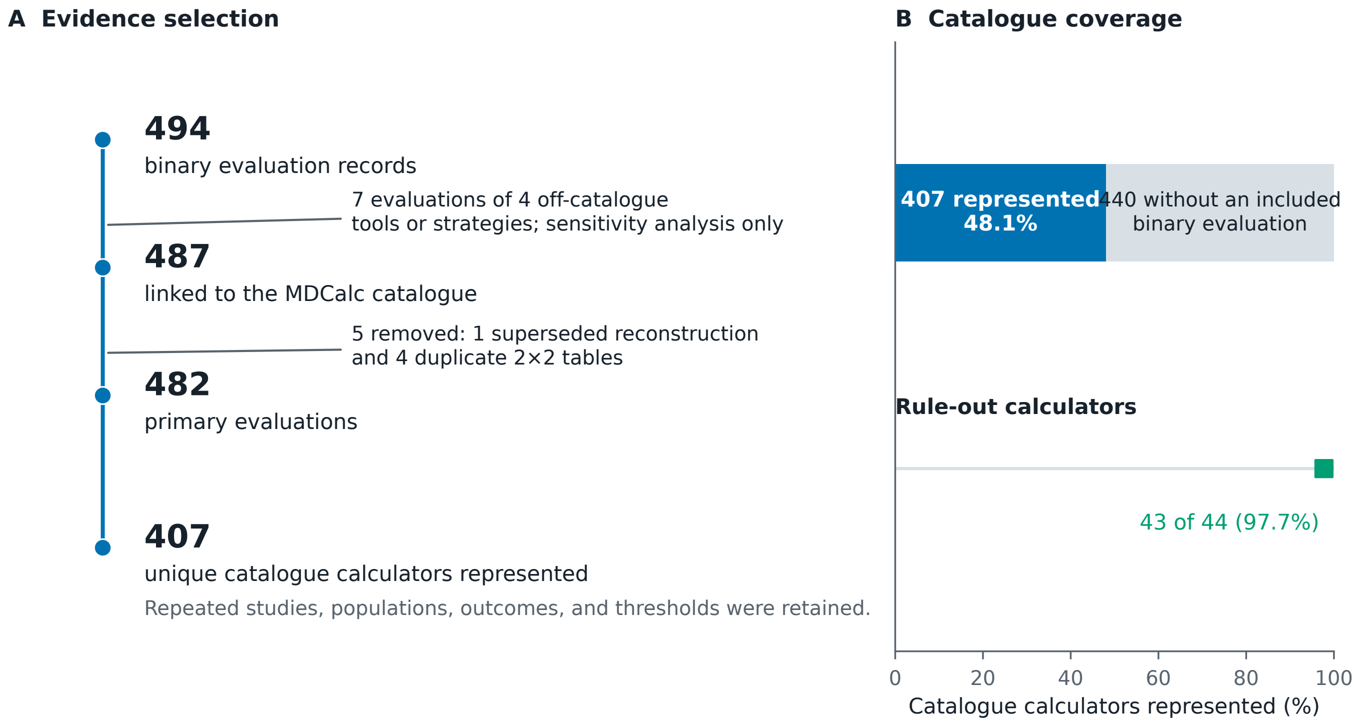

### figure1_selection_flow.png

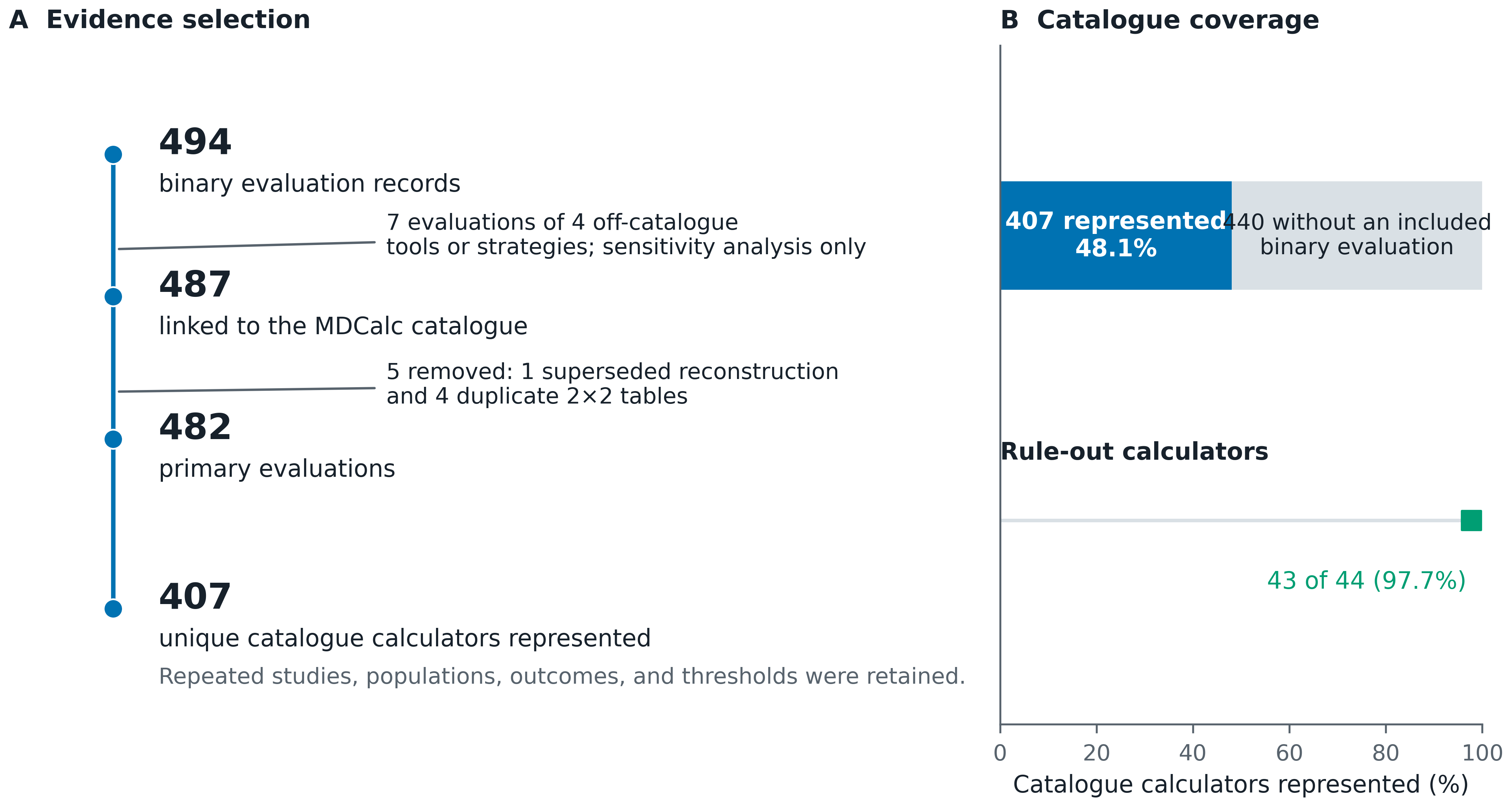

### figure1_selection_flow_grayscale.pdf

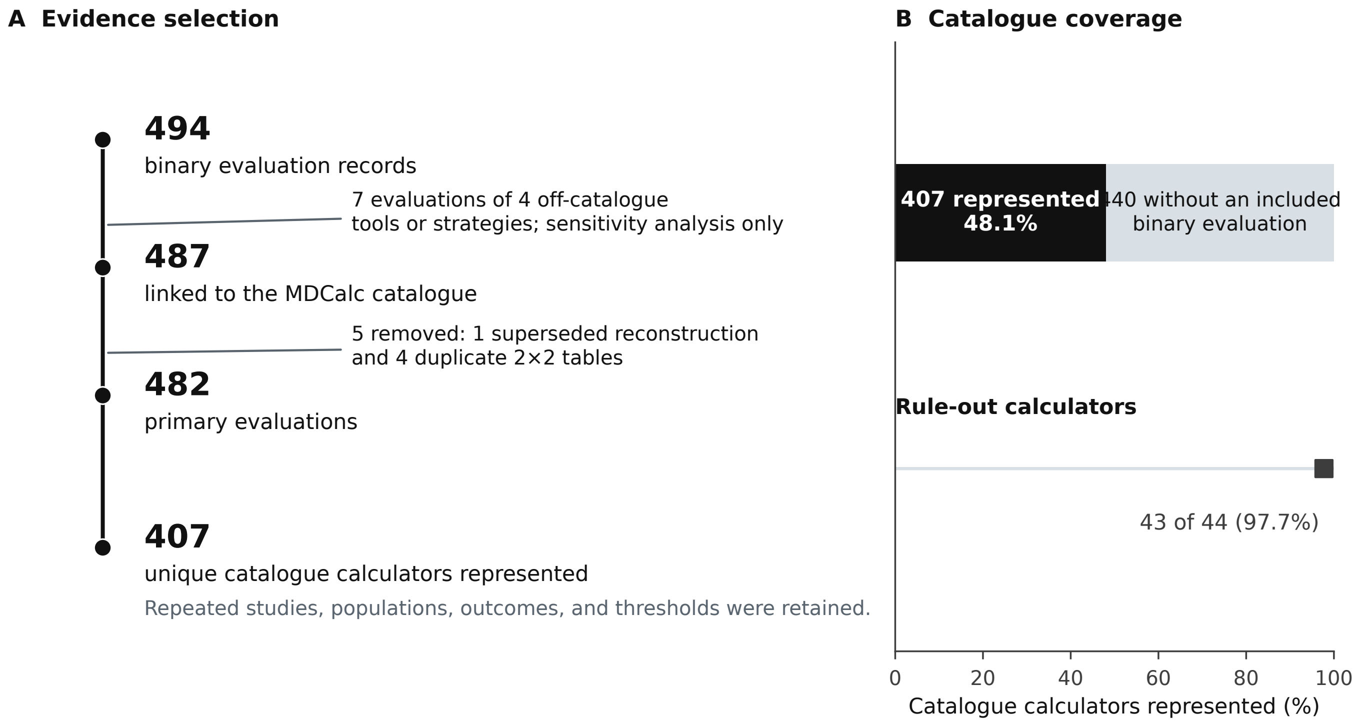

### figure1_selection_flow_grayscale.png

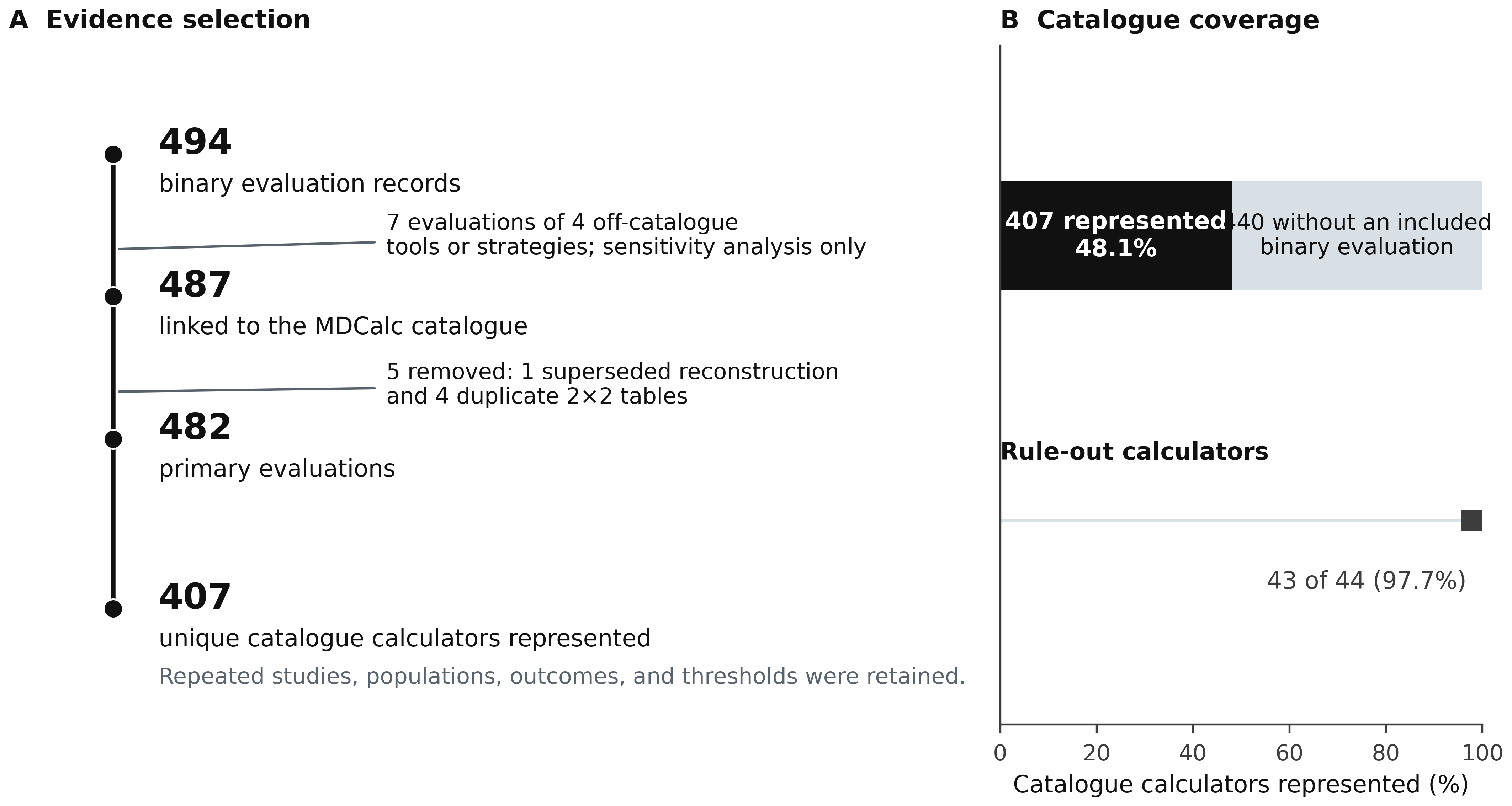

### figure2_catalogue_atlas.pdf

#### A calculator result has several quantitative meanings

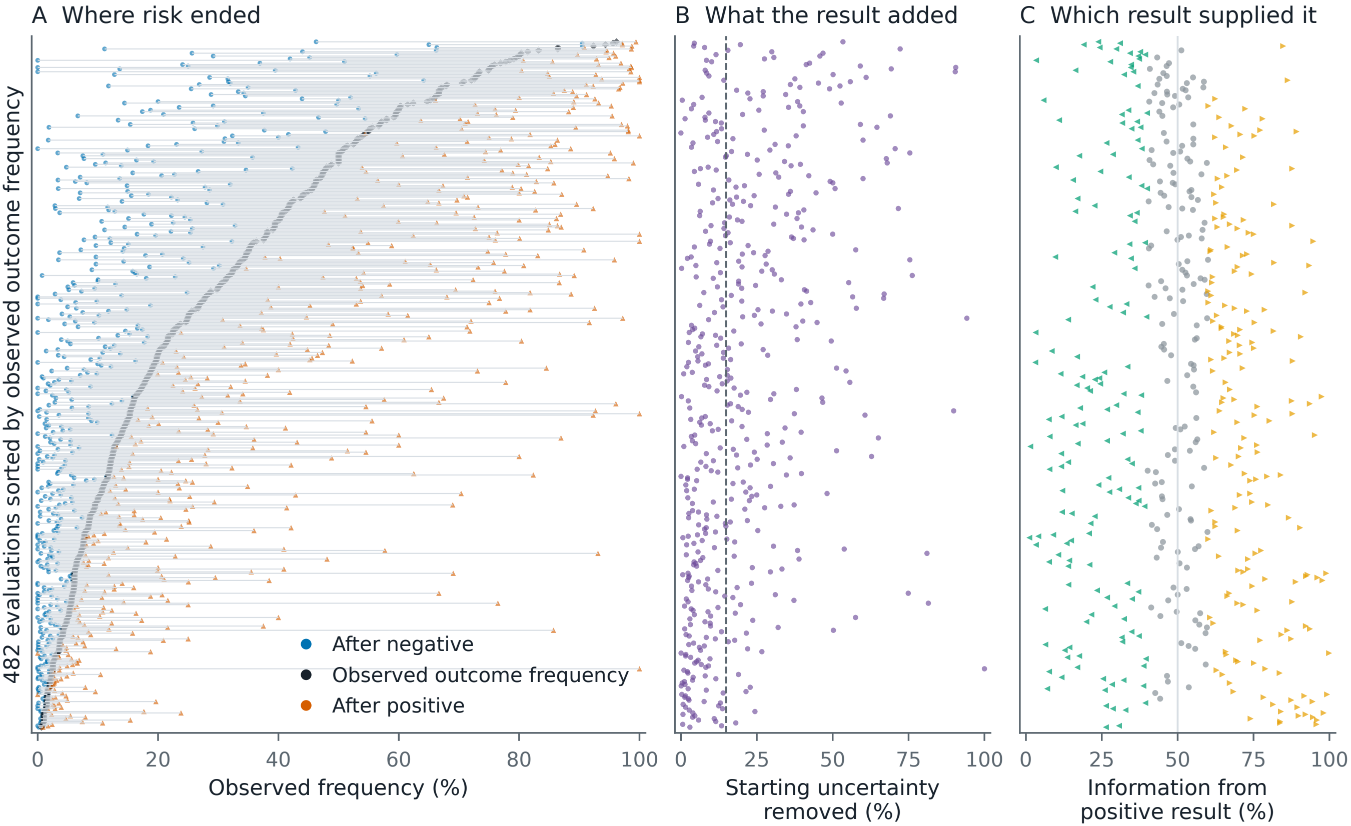

### figure2_catalogue_atlas.png

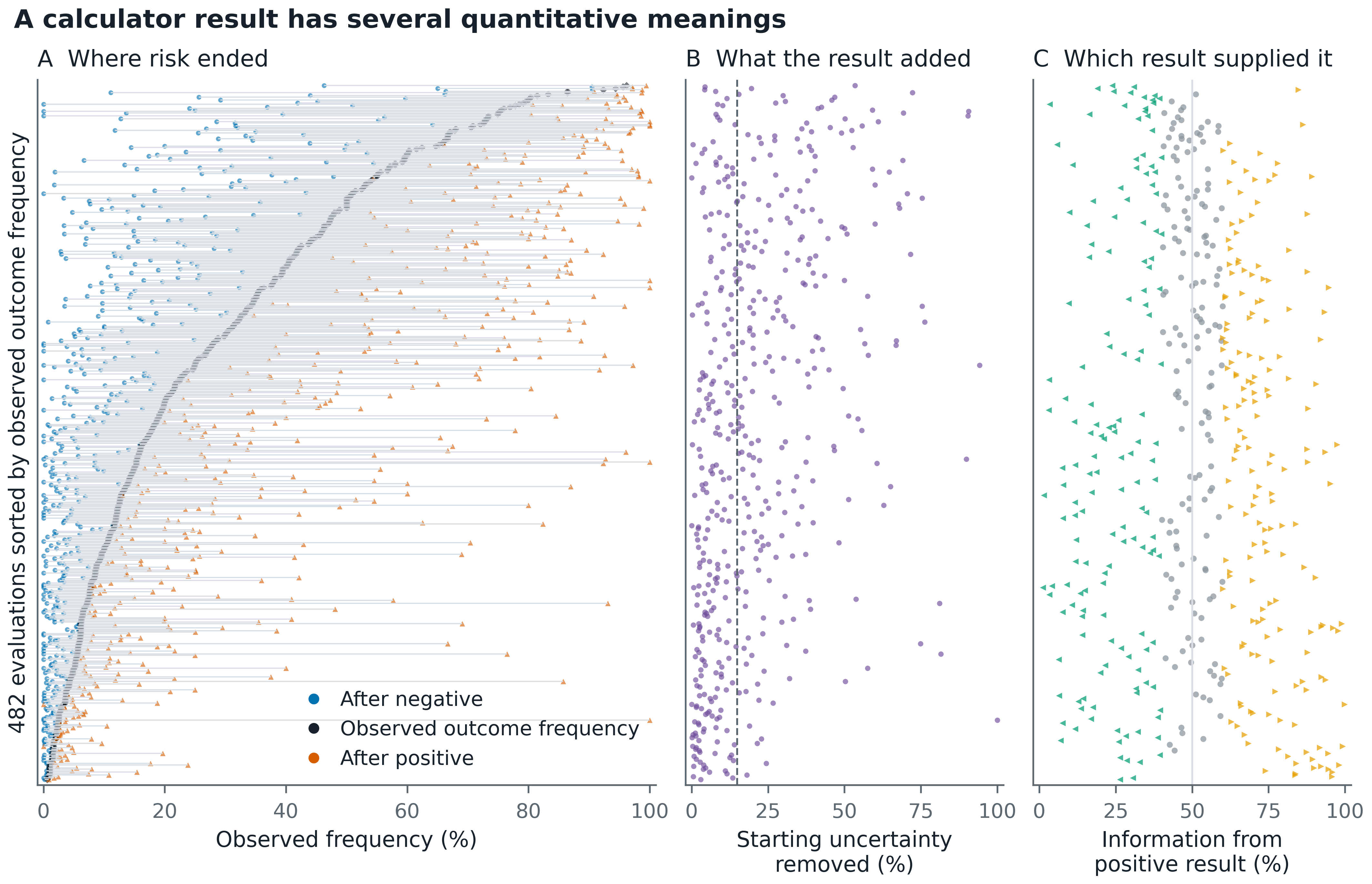

### figure2_catalogue_atlas_grayscale.pdf

#### A calculator result has several quantitative meanings

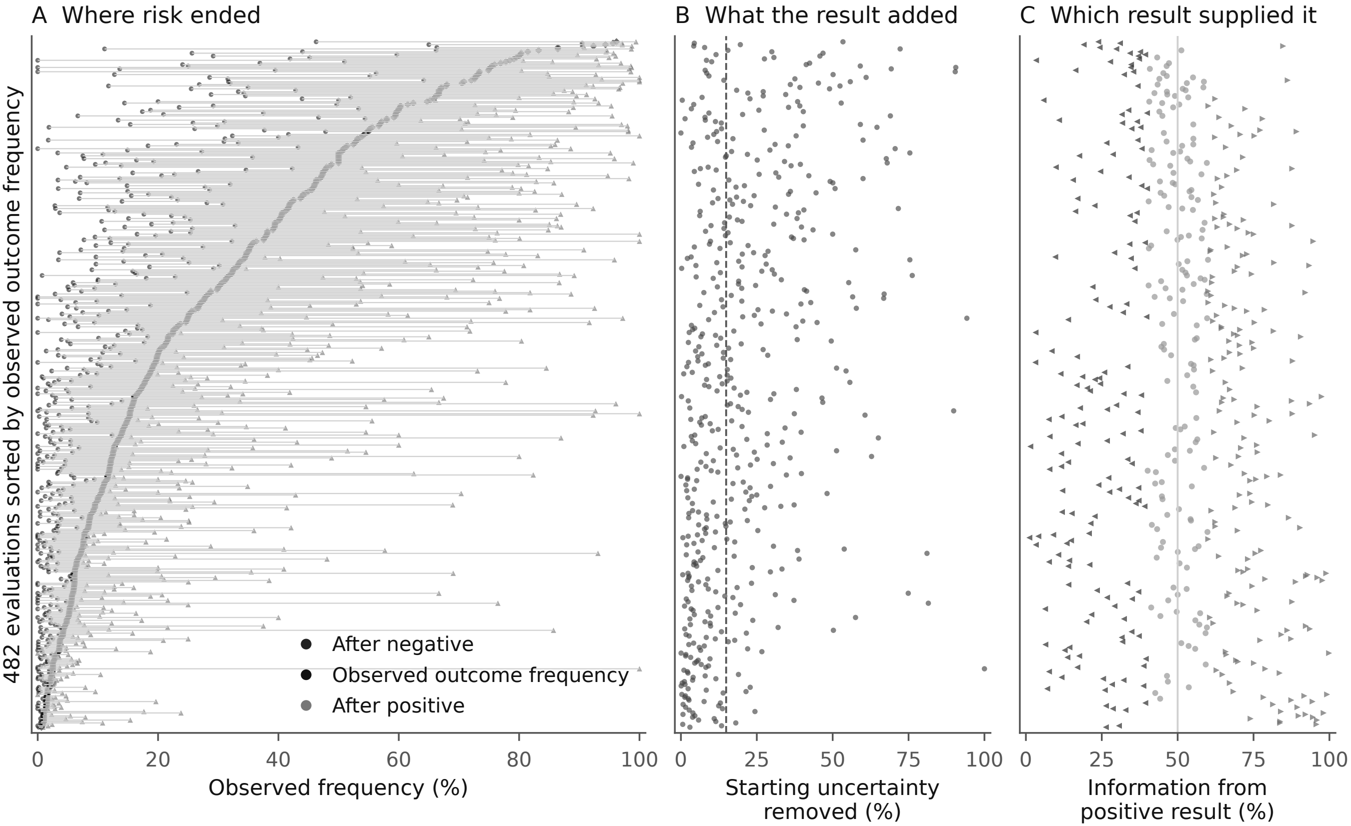

### figure2_catalogue_atlas_grayscale.png

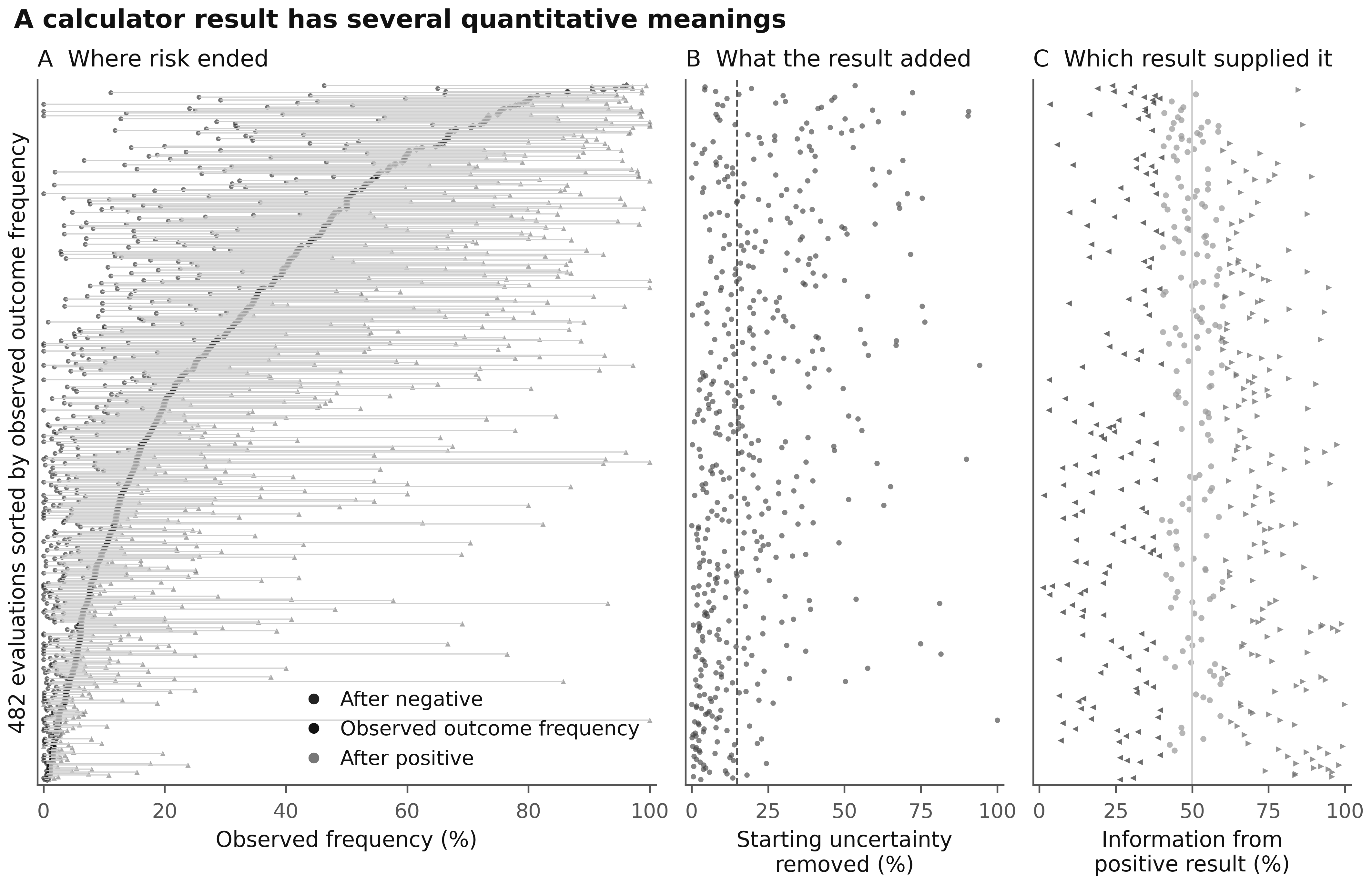

### figure2_information_yield_distribution.pdf

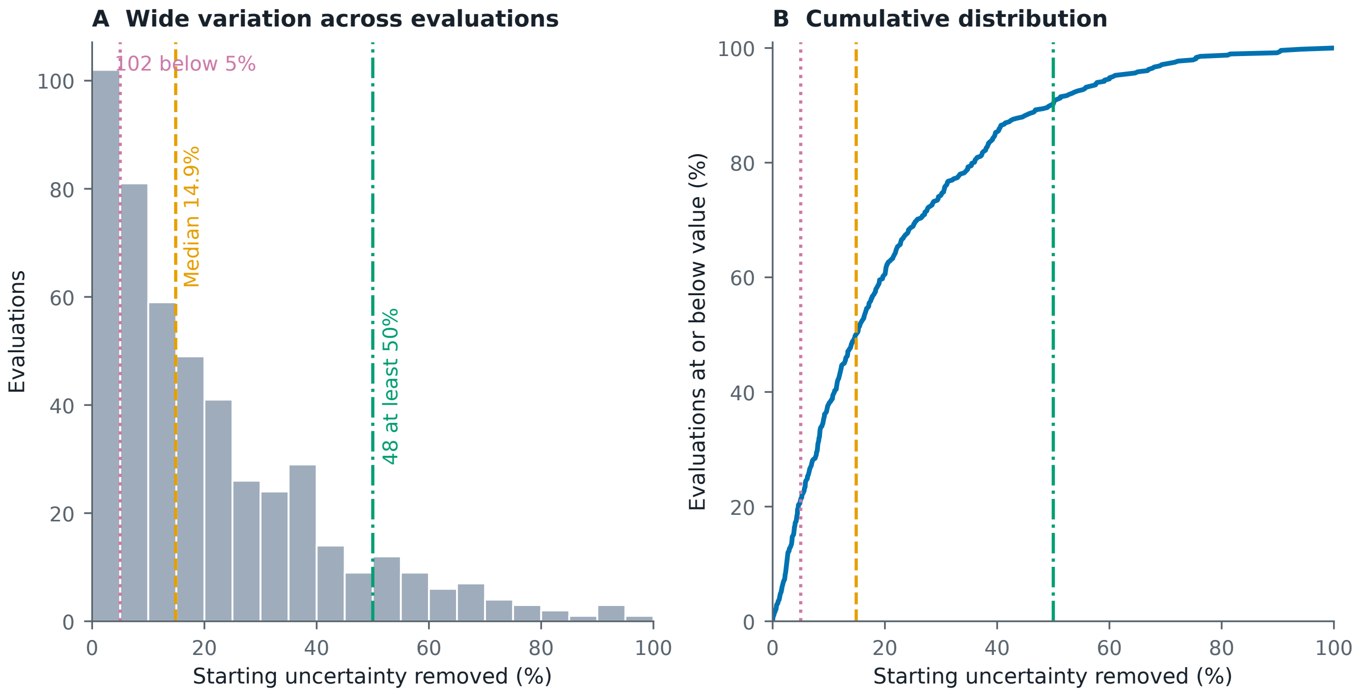

### figure2_information_yield_distribution.png

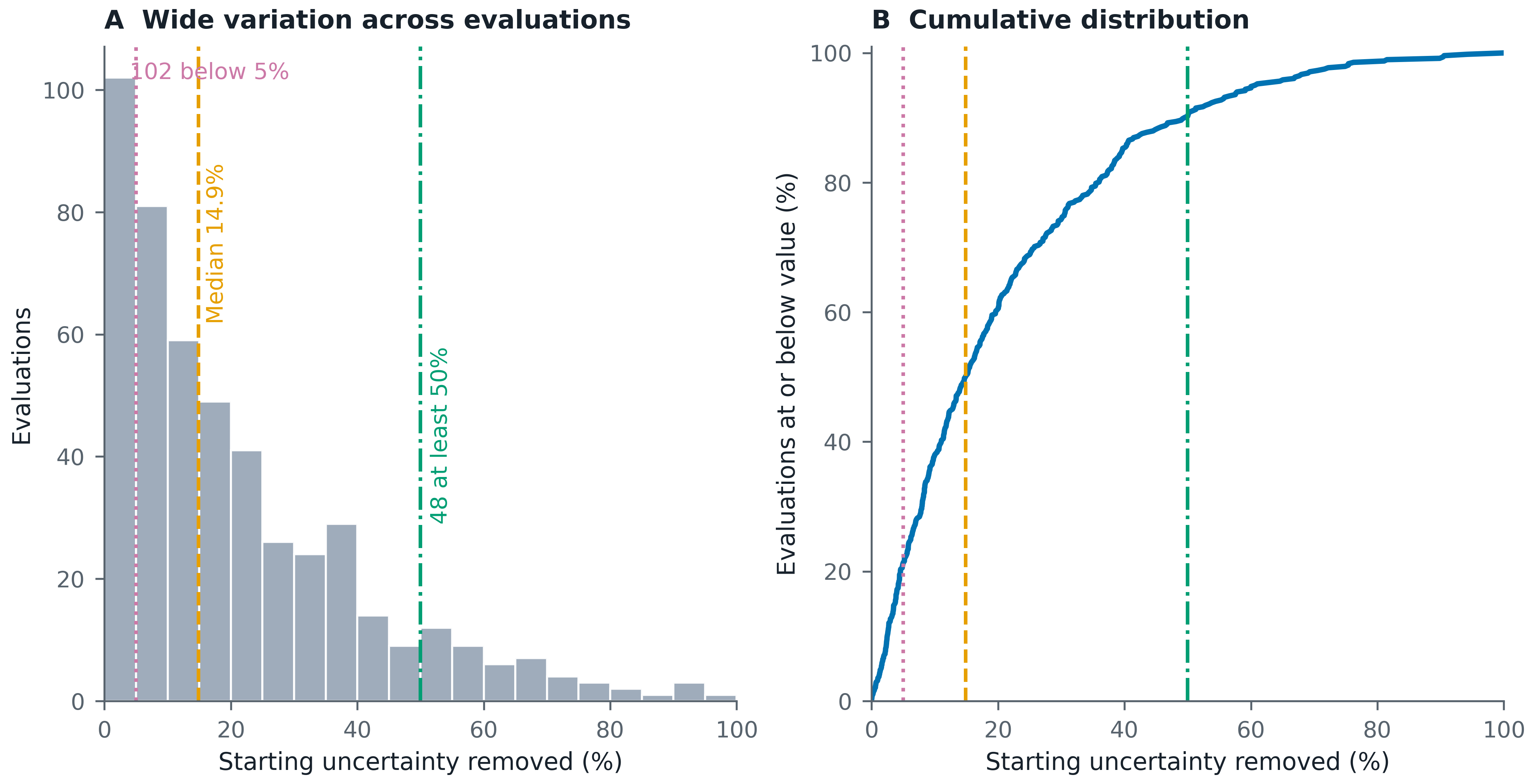

### figure2_information_yield_distribution_grayscale.pdf

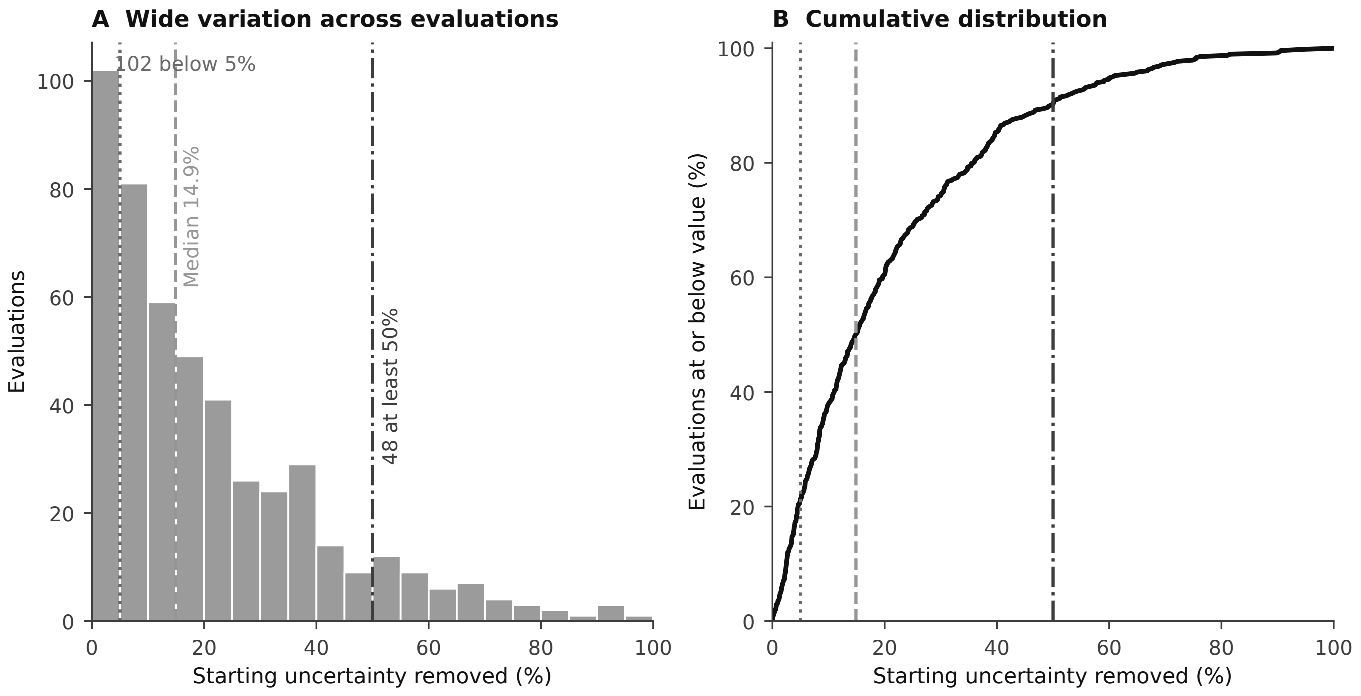

### figure2_information_yield_distribution_grayscale.png

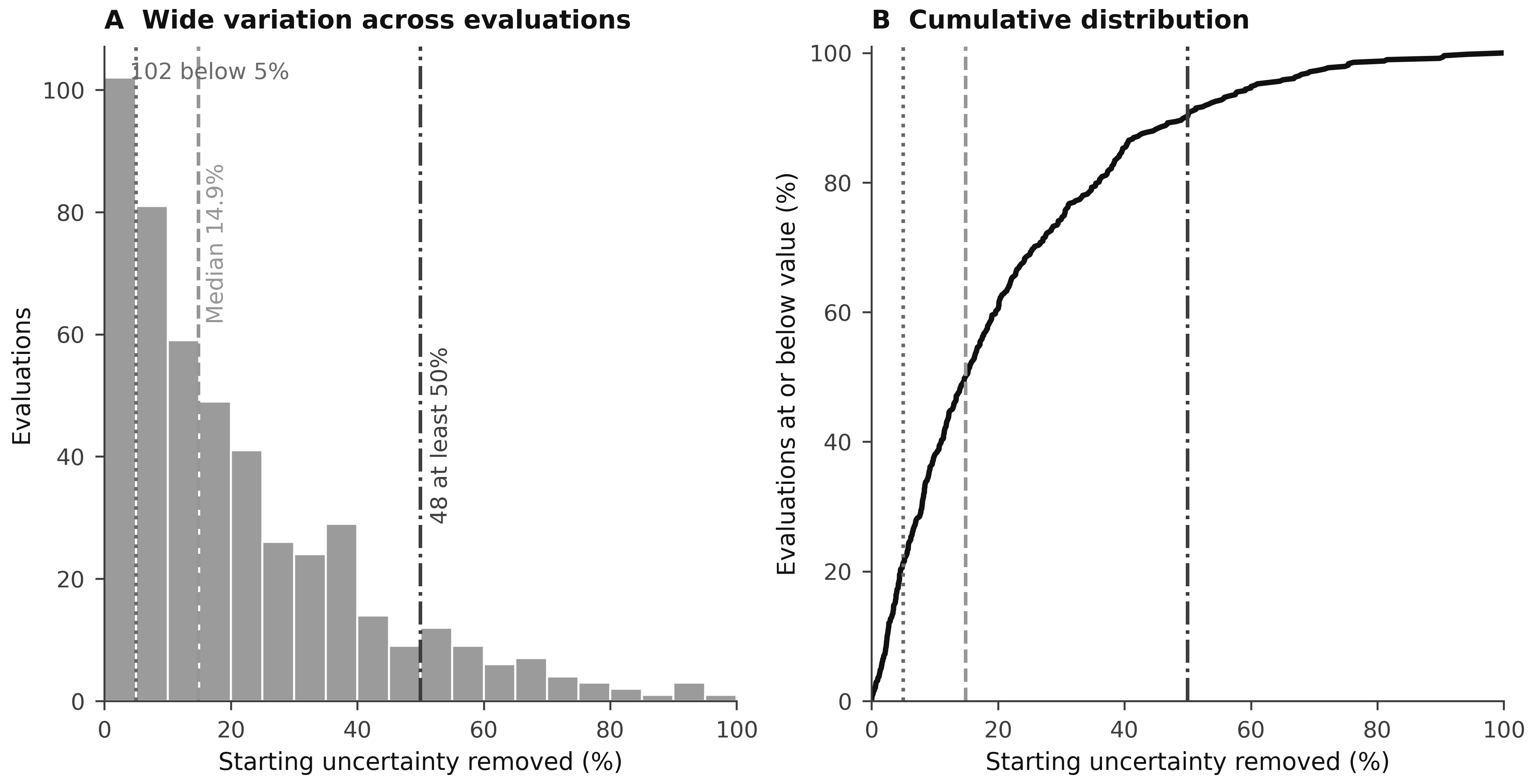

### figure3_result_information_contributions.pdf

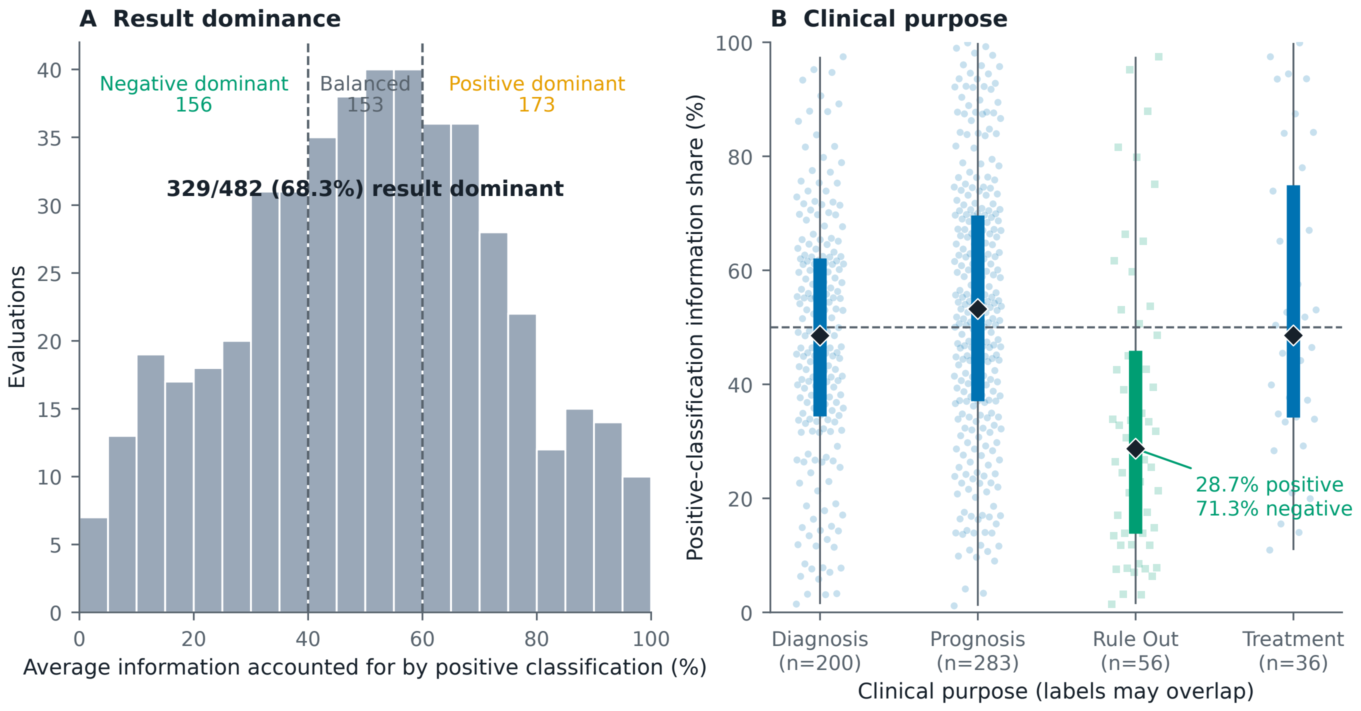

### figure3_result_information_contributions.png

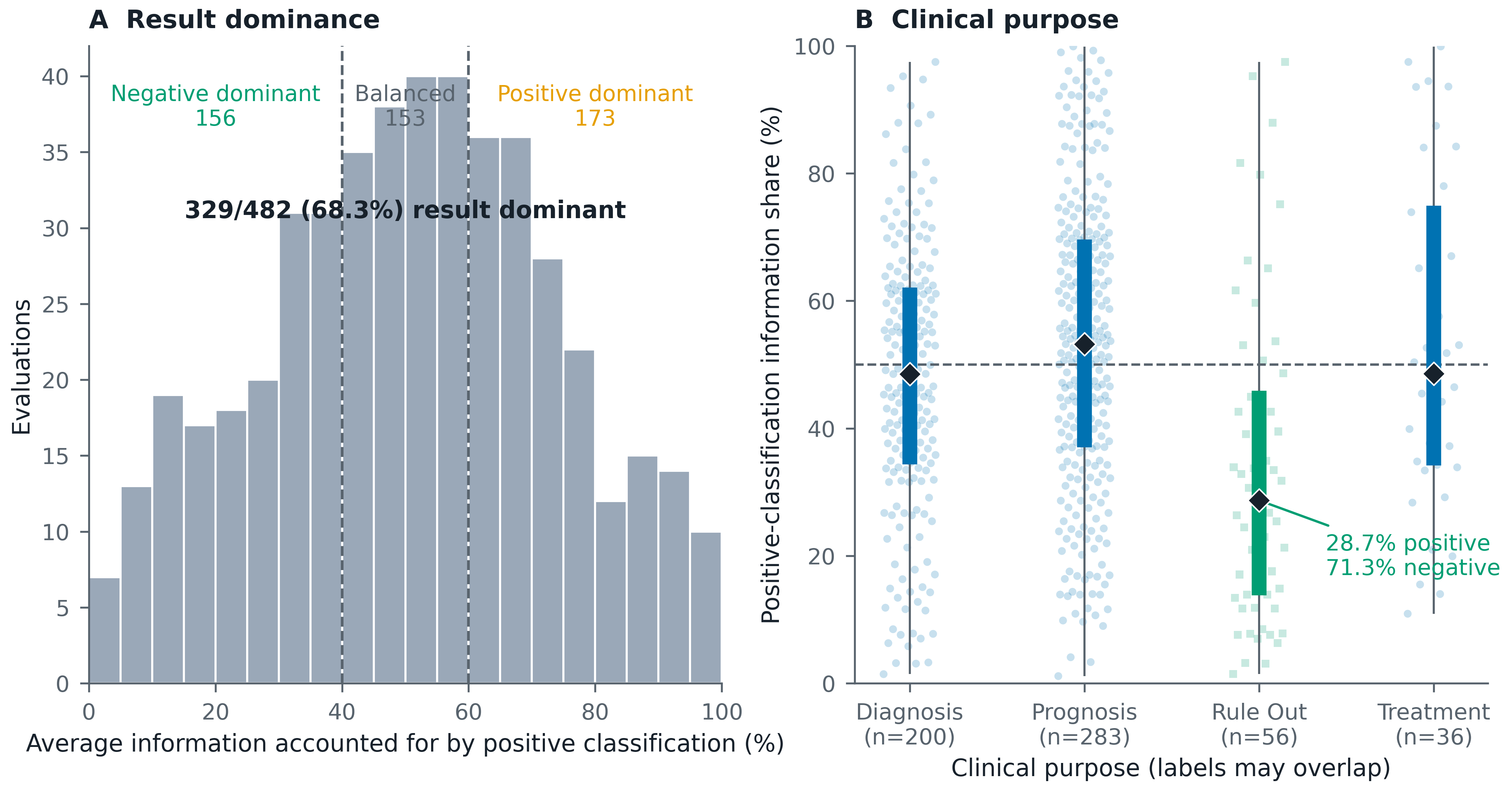

### figure3_result_information_contributions_grayscale.pdf

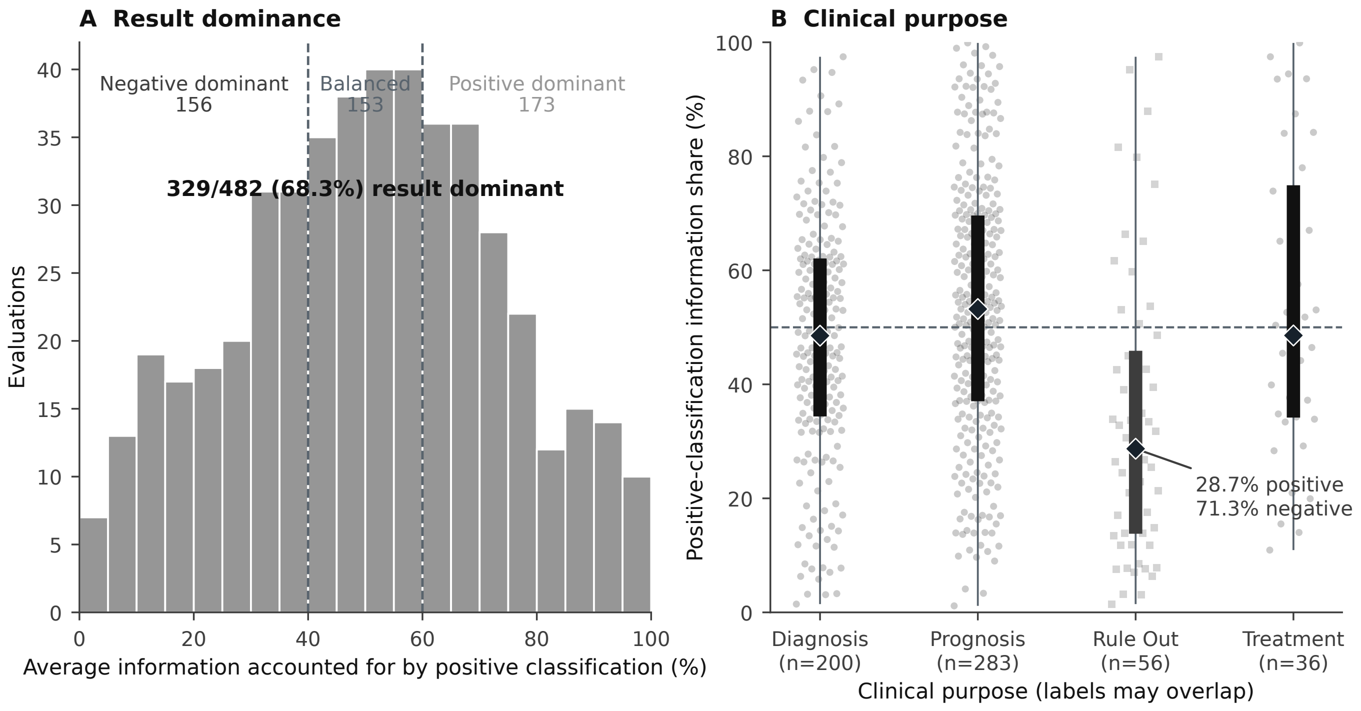

### figure3_result_information_contributions_grayscale.png

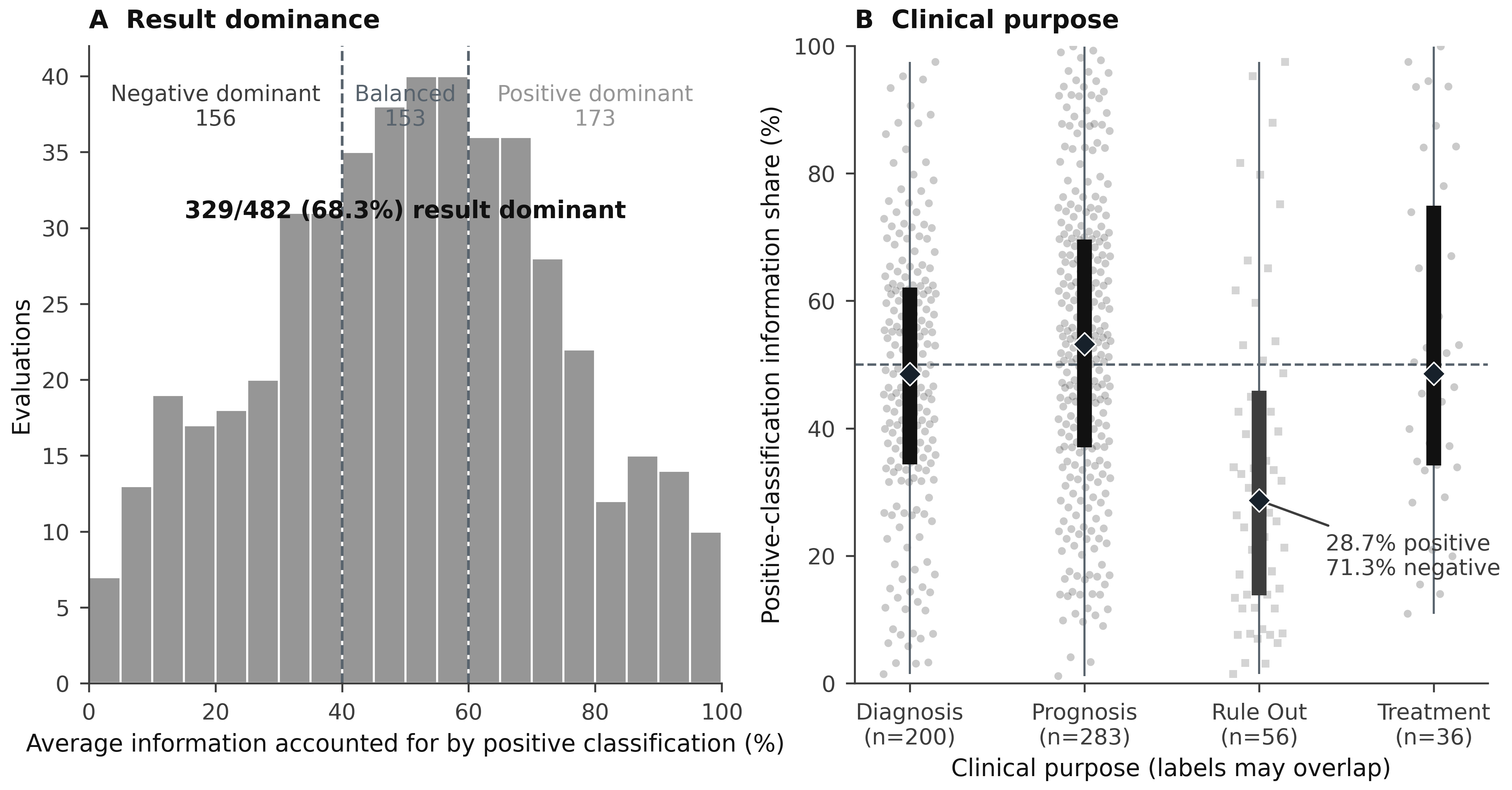

### figure3_ruleout_precision.pdf

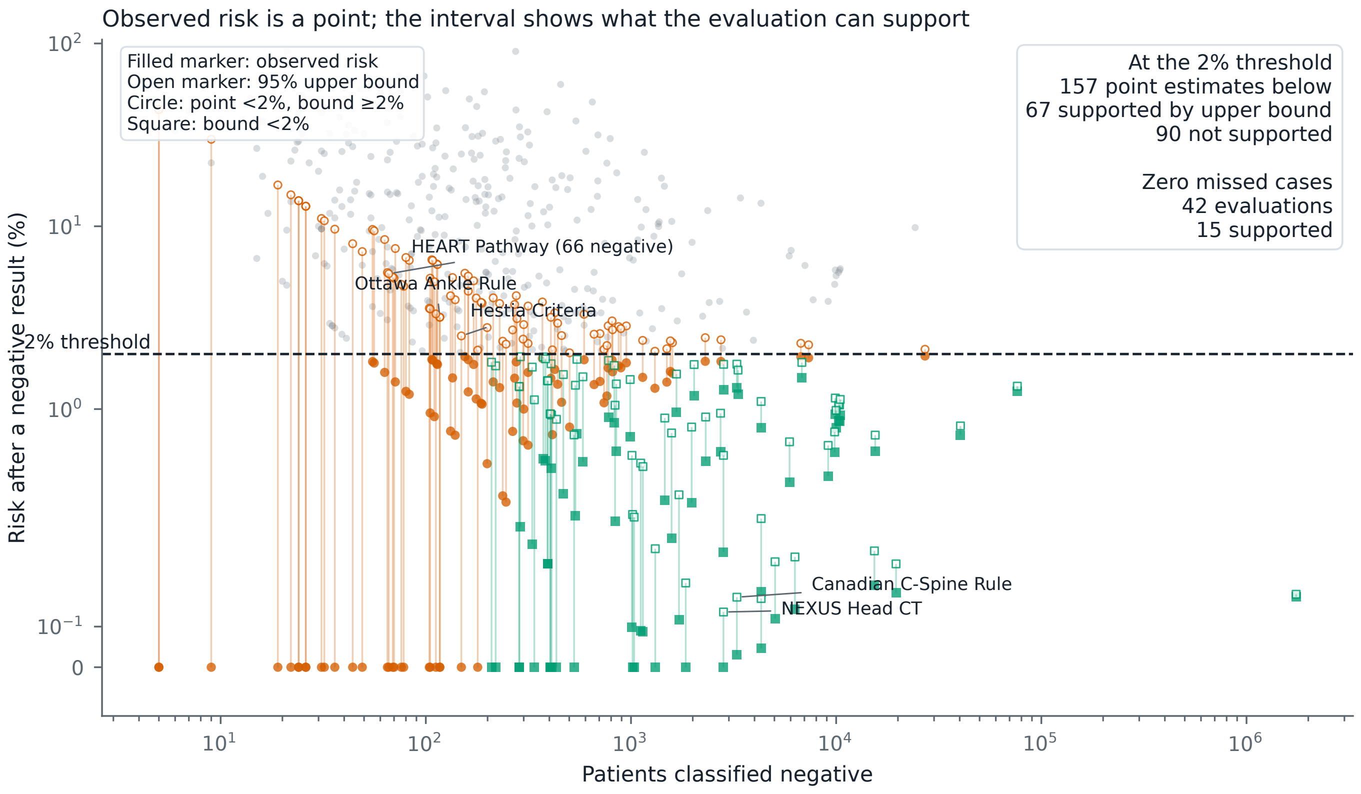

### figure3_ruleout_precision.png

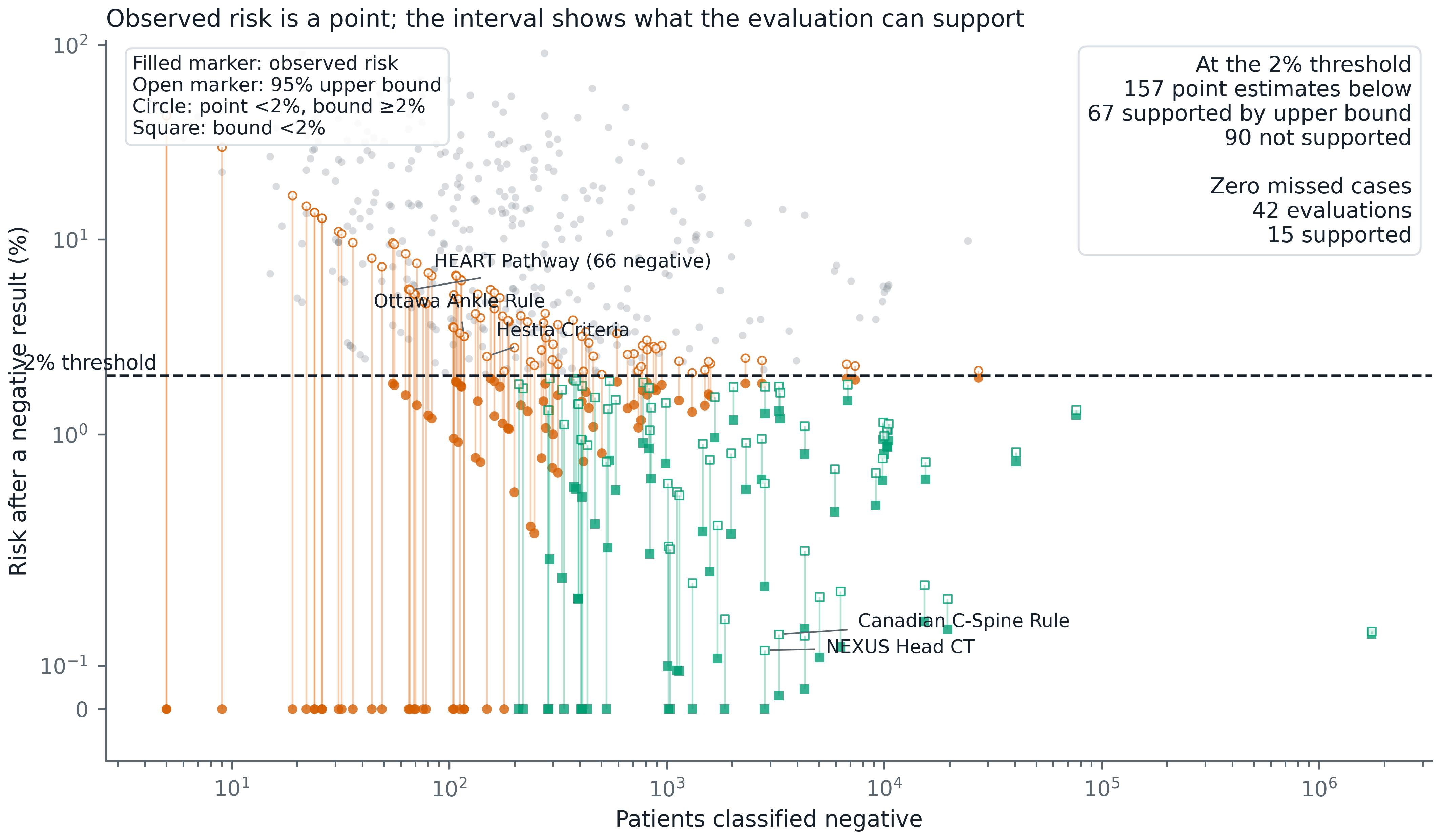

### figure3_ruleout_precision_grayscale.pdf

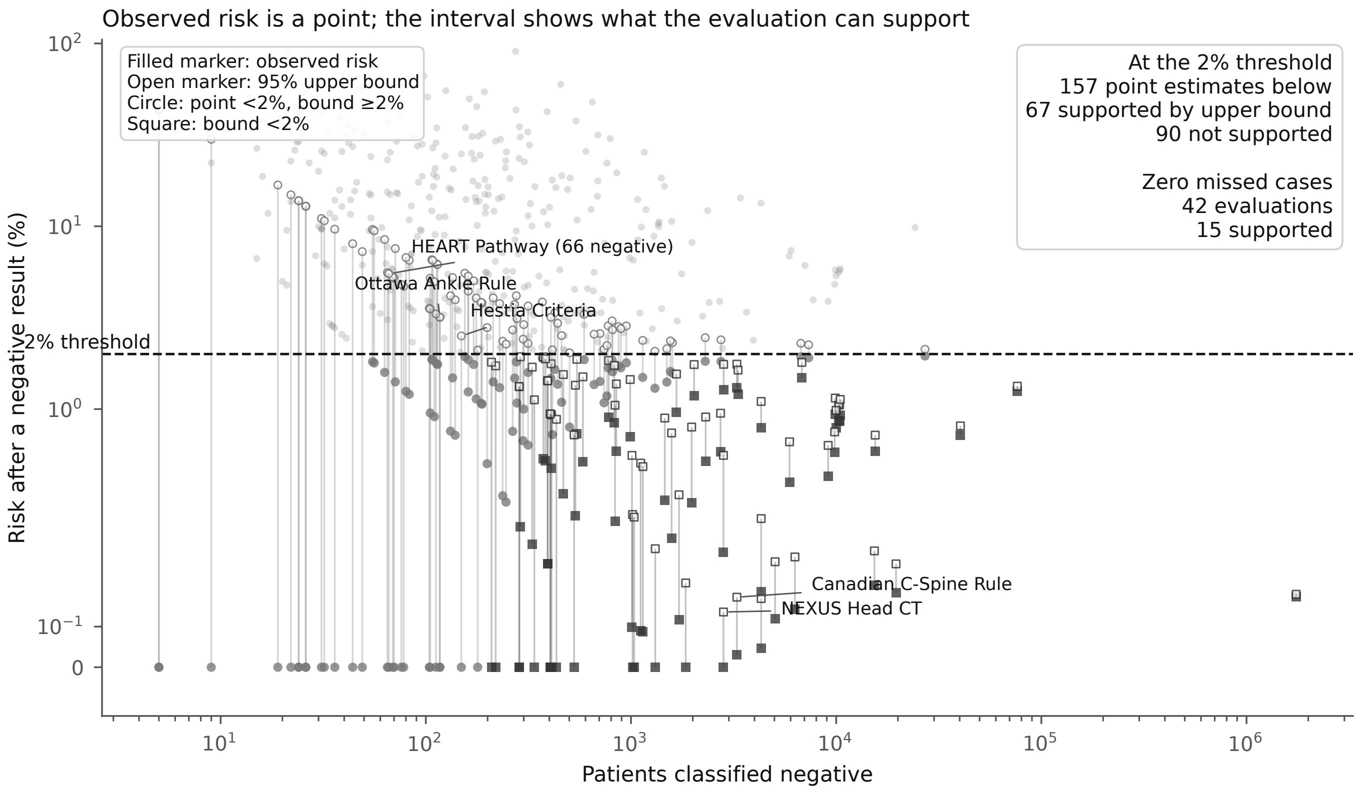

### figure3_ruleout_precision_grayscale.png

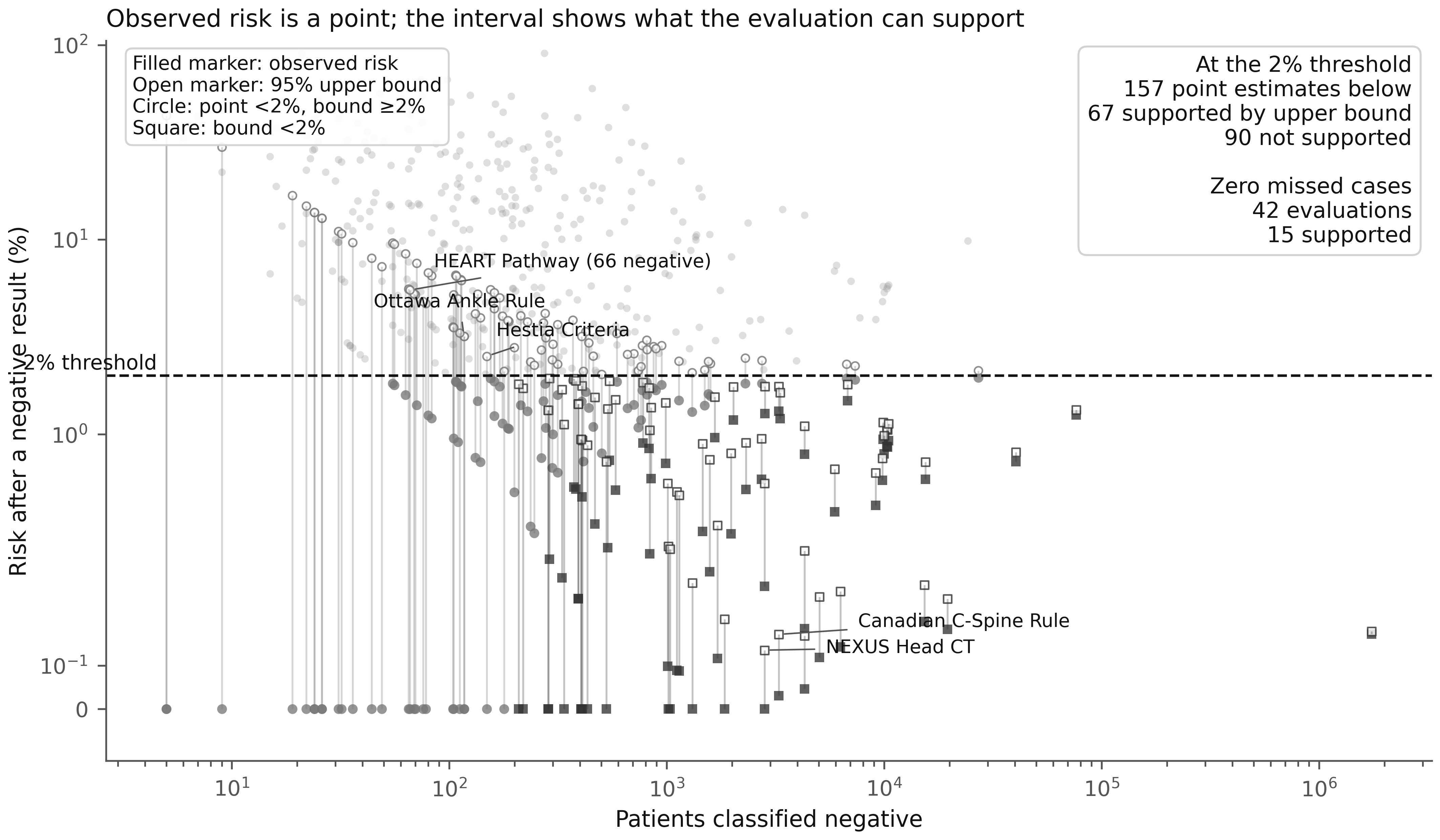

### figure4_prevalence_information_yield.pdf

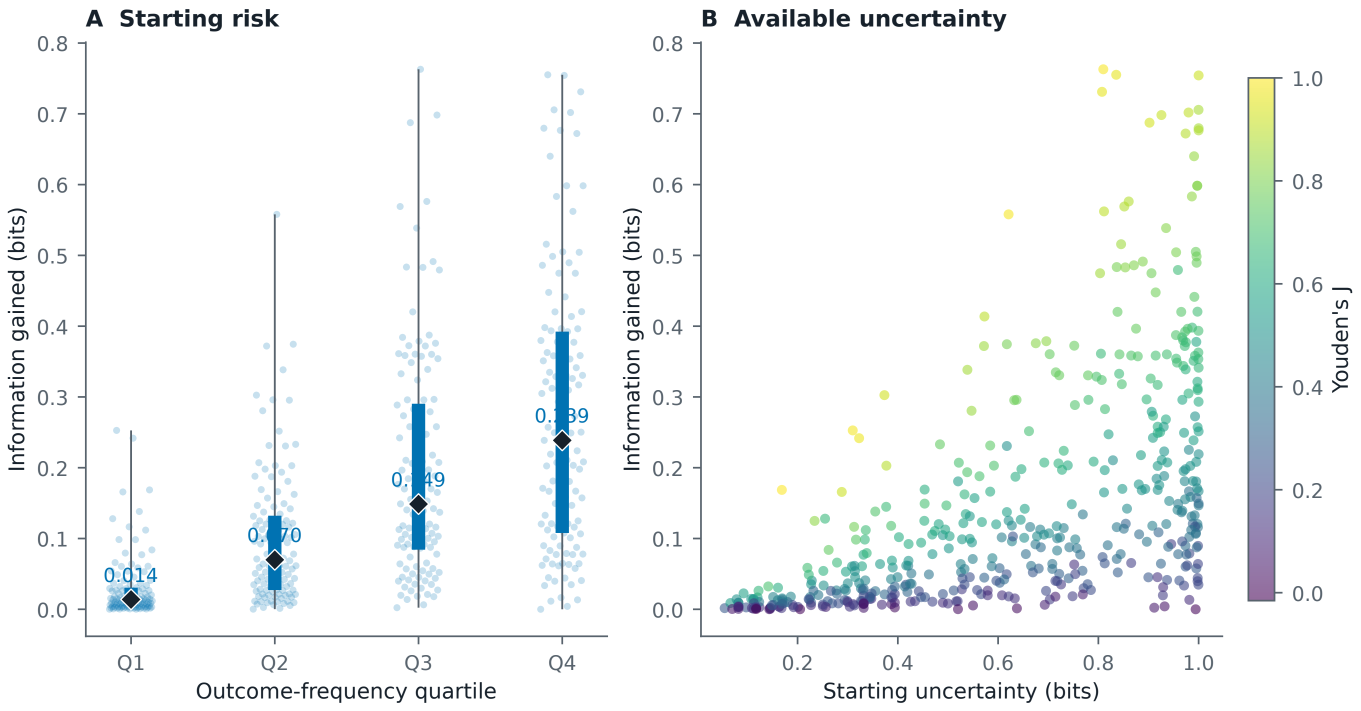

### figure4_prevalence_information_yield.png

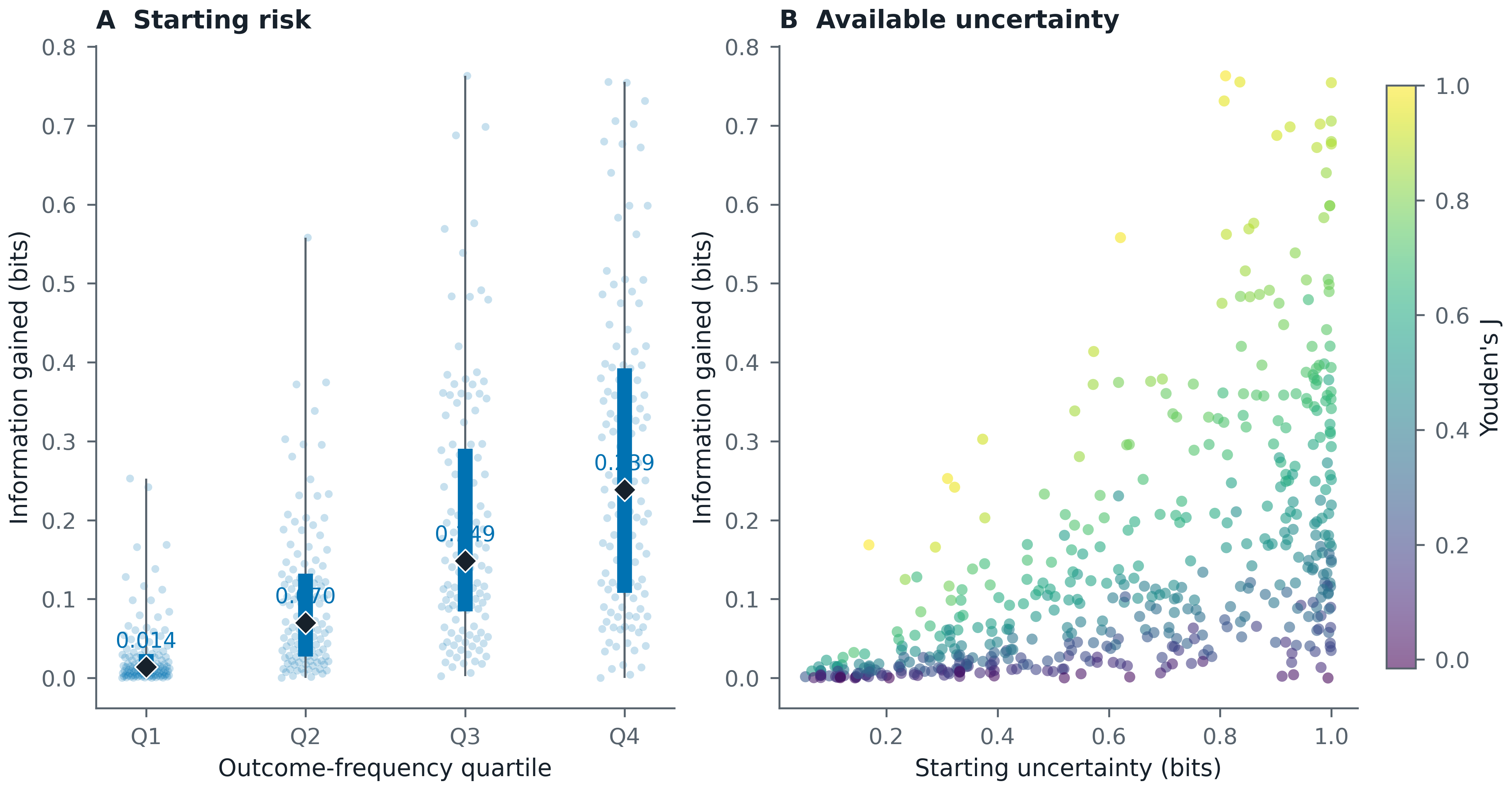

### figure4_prevalence_information_yield_grayscale.pdf

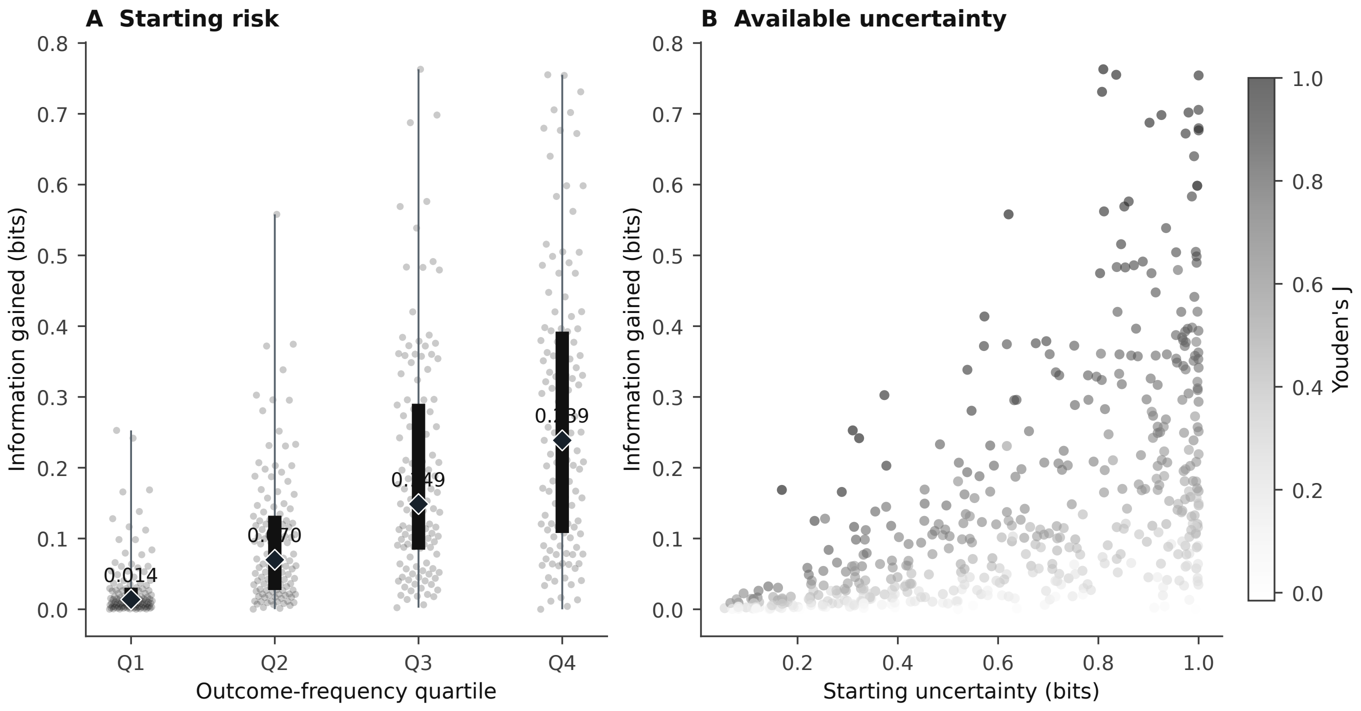

### figure4_prevalence_information_yield_grayscale.png

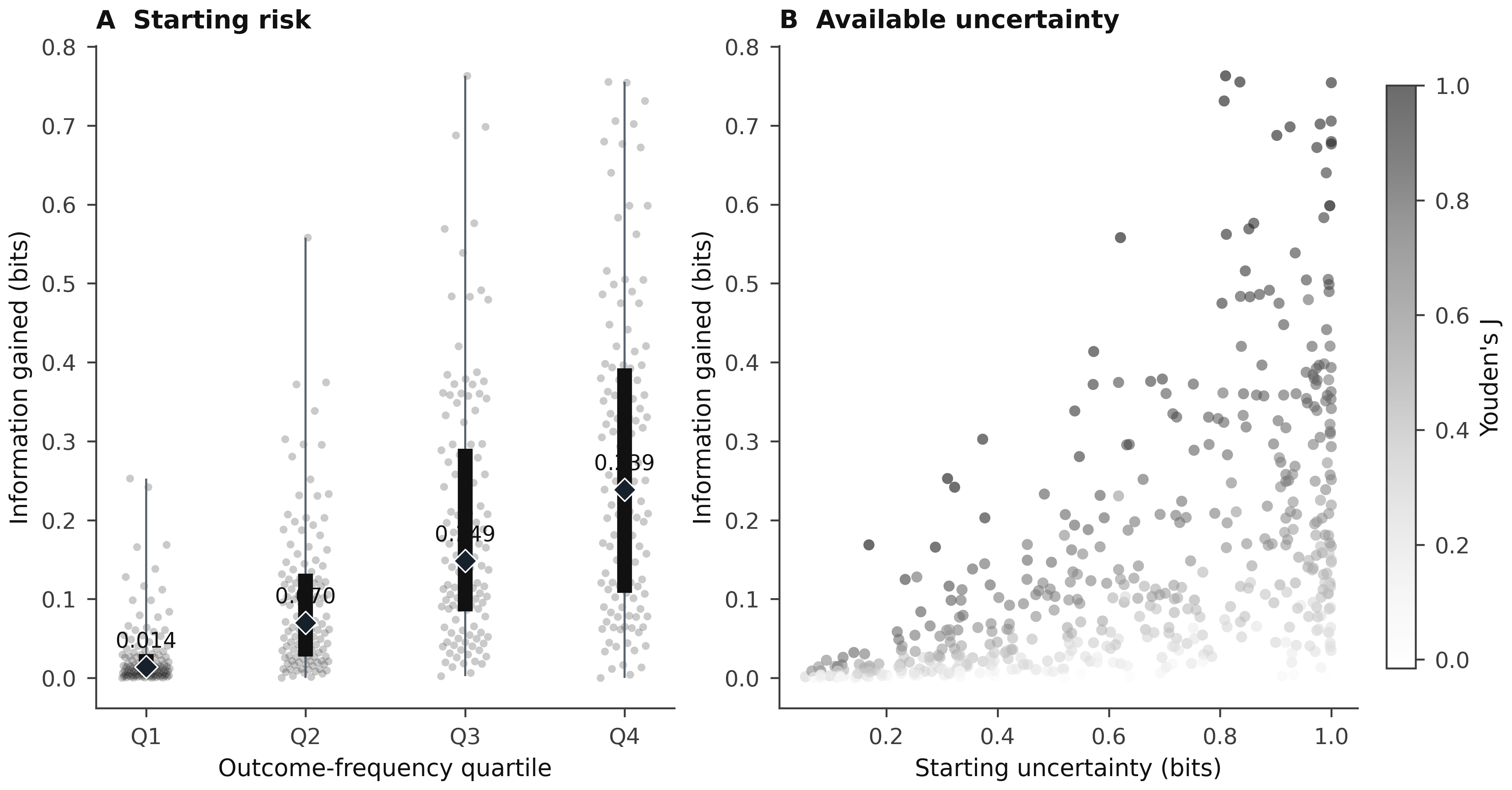

### figure5_ruleout_support.pdf

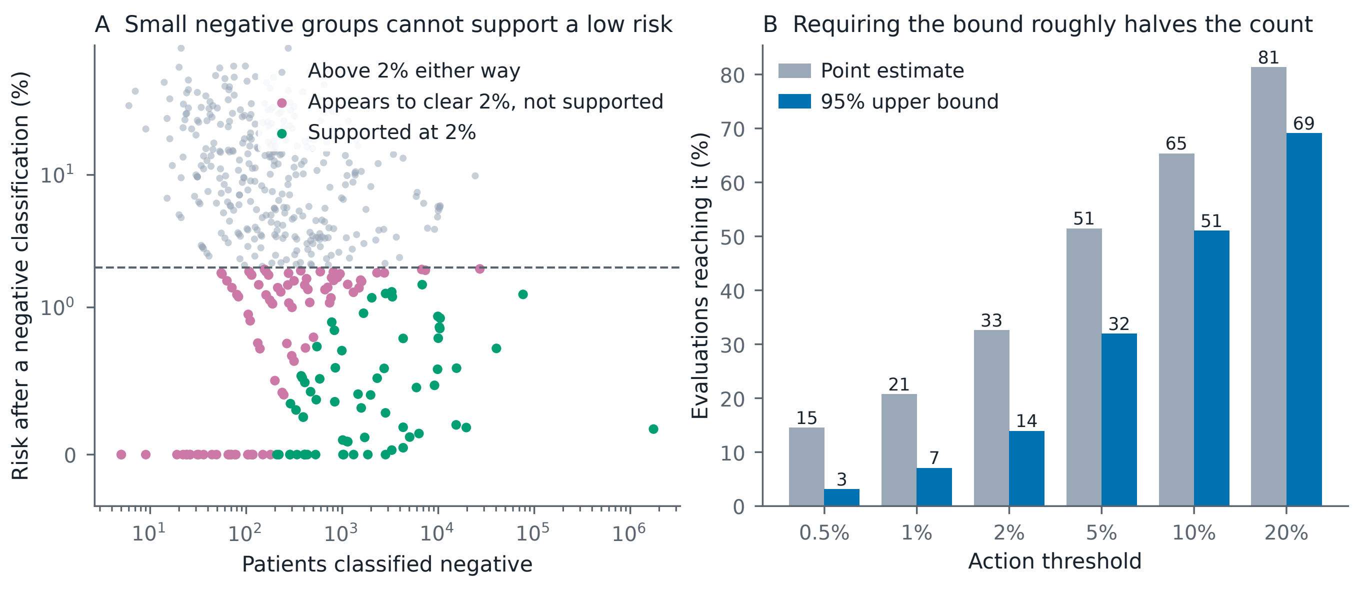

### figure5_ruleout_support.png

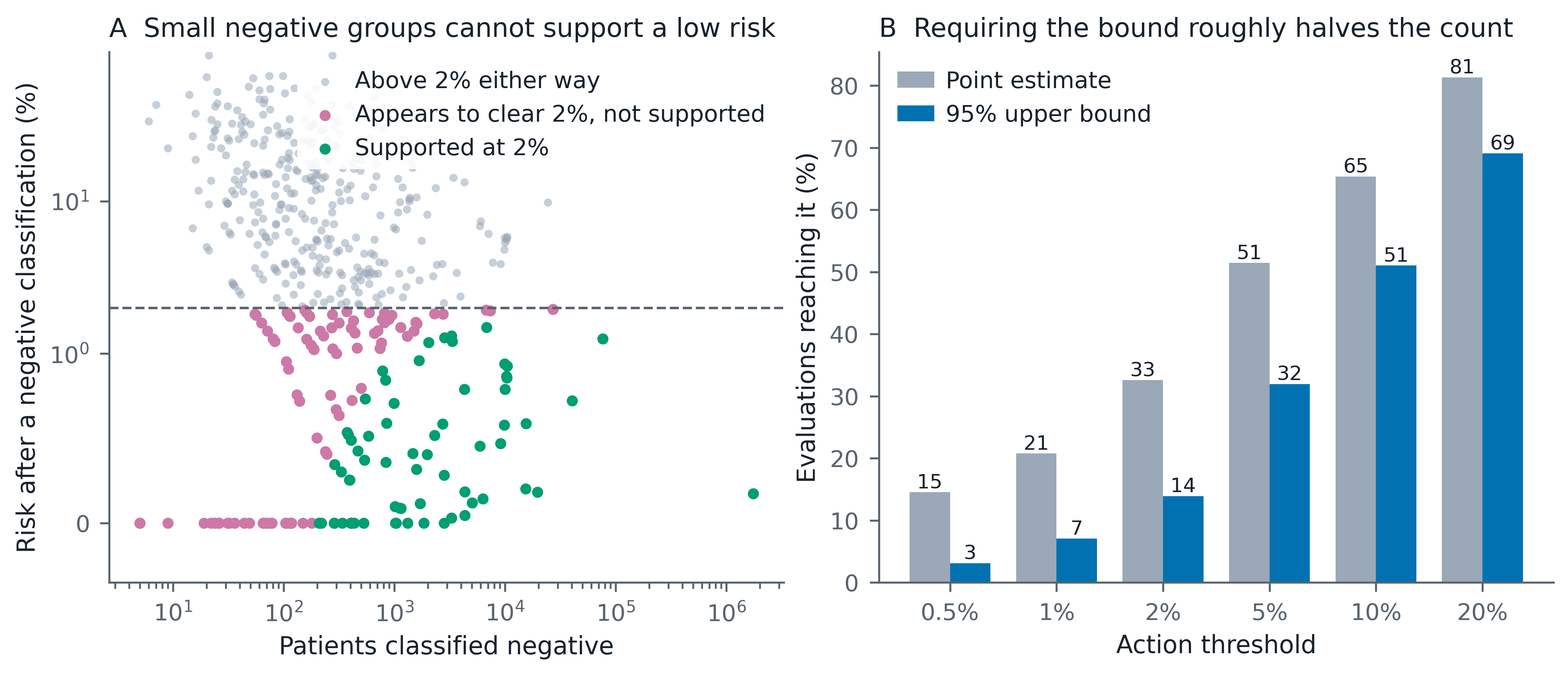

### figure5_ruleout_support_grayscale.pdf

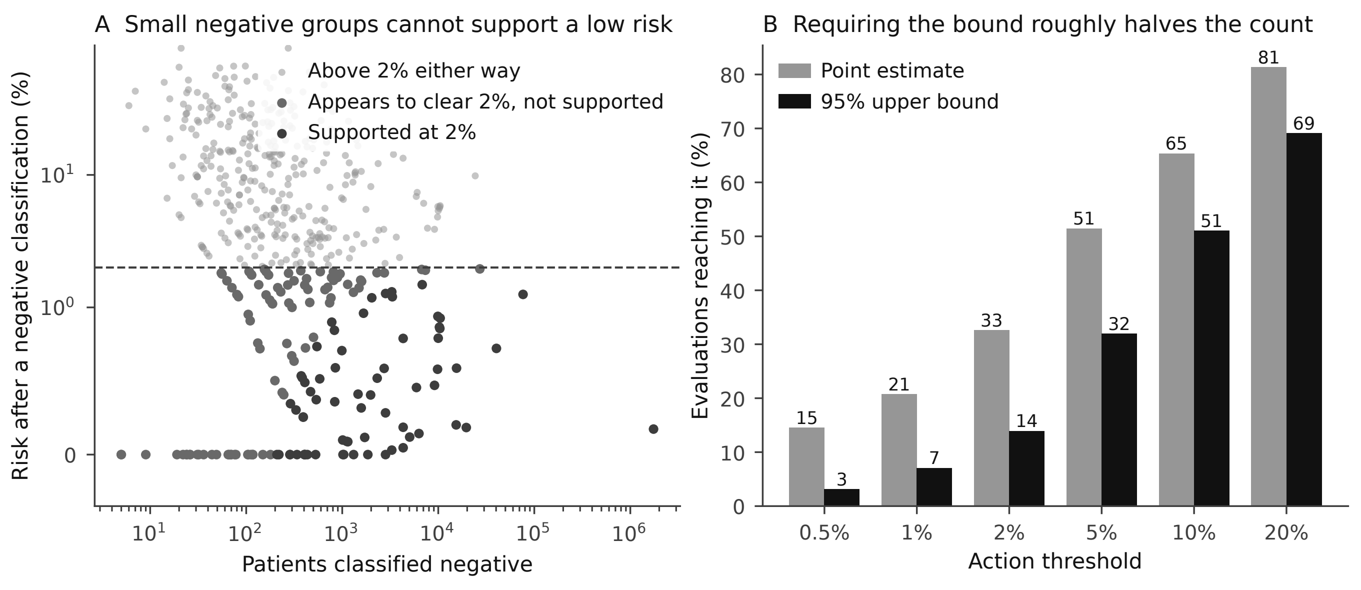

### figure5_ruleout_support_grayscale.png

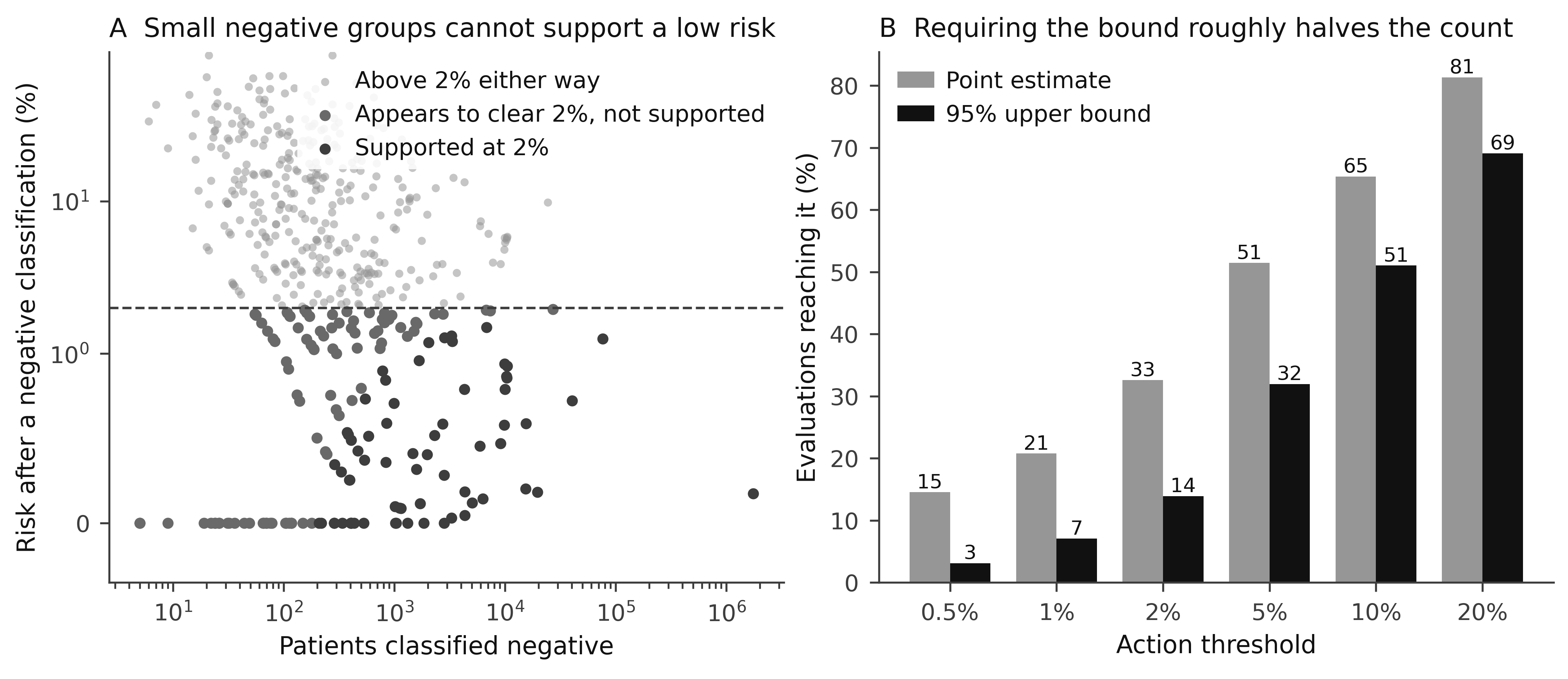

### figureS1_sample_size_information_yield.pdf

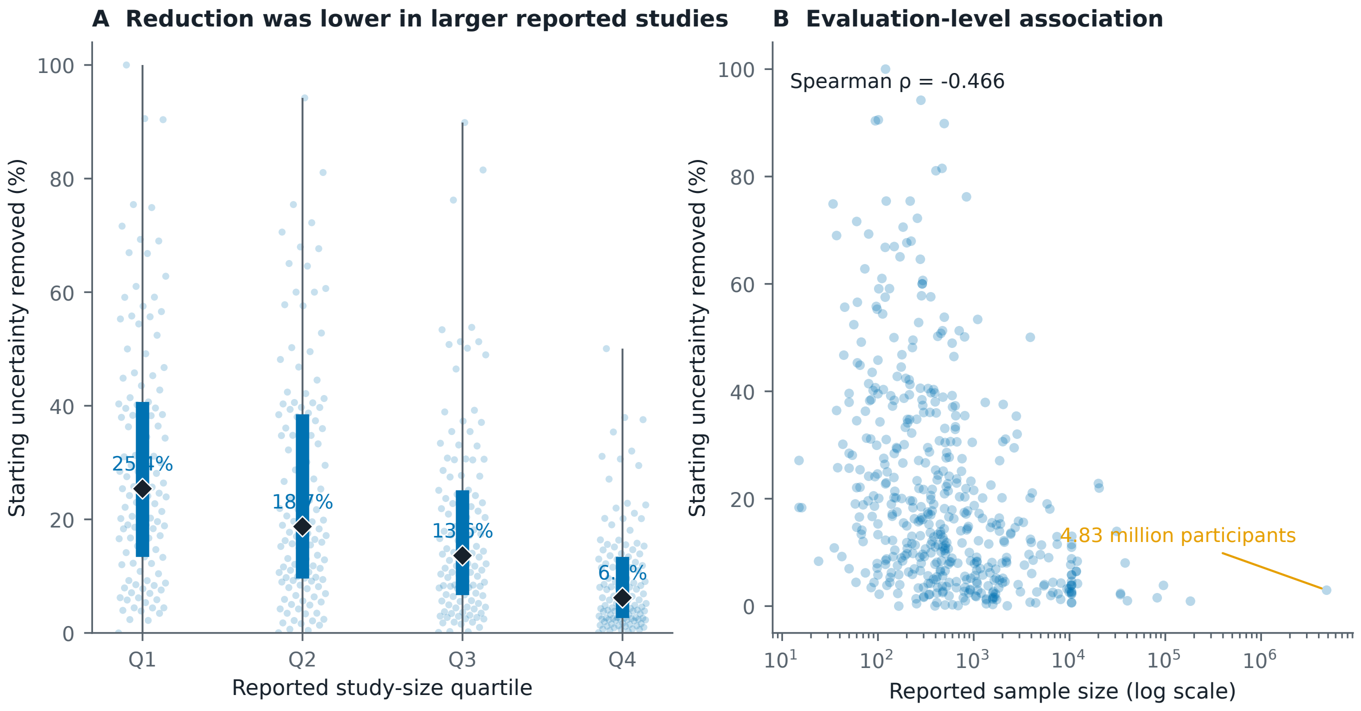

### figureS1_sample_size_information_yield.png

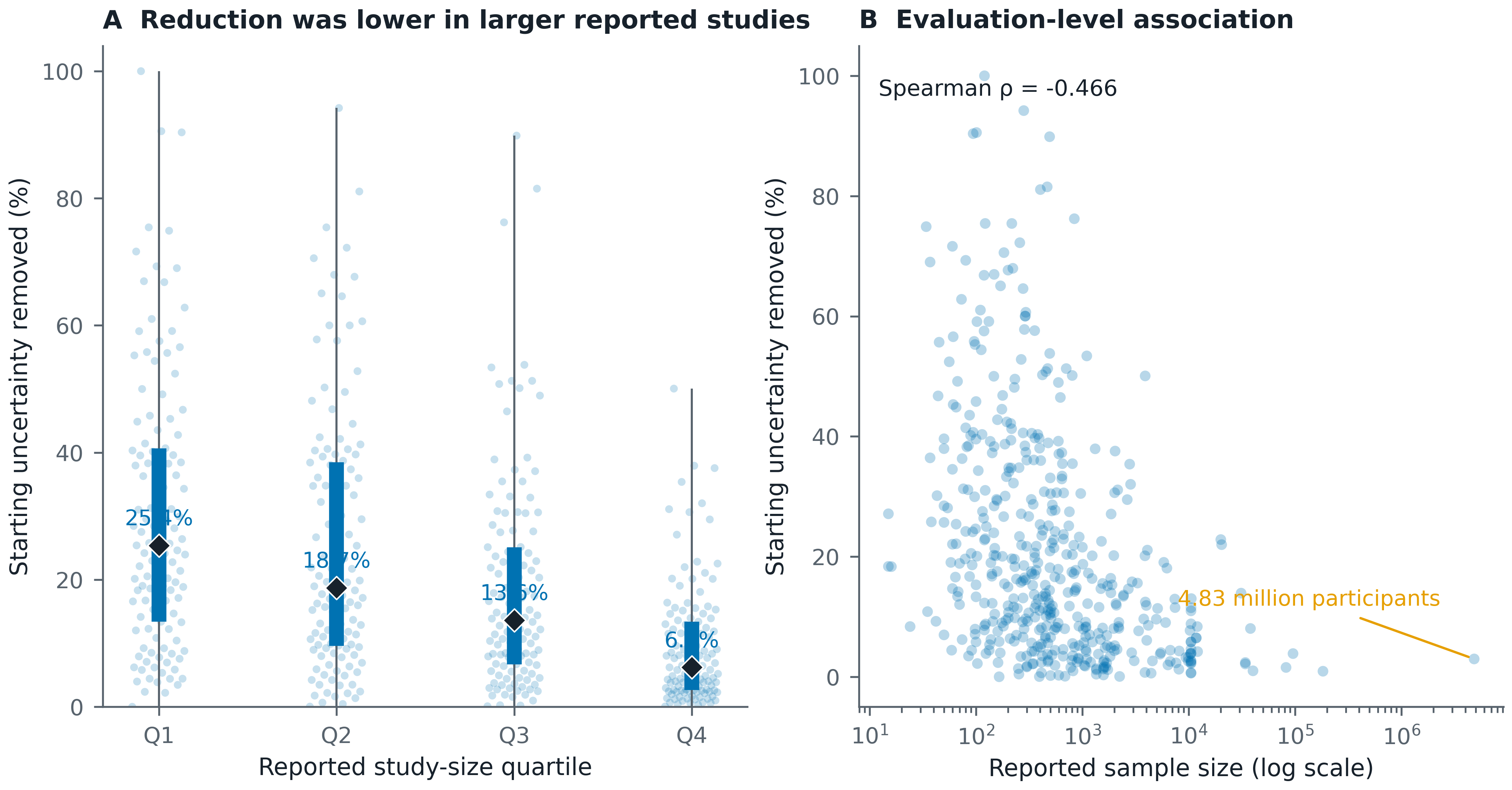

### figureS1_sample_size_information_yield_grayscale.pdf

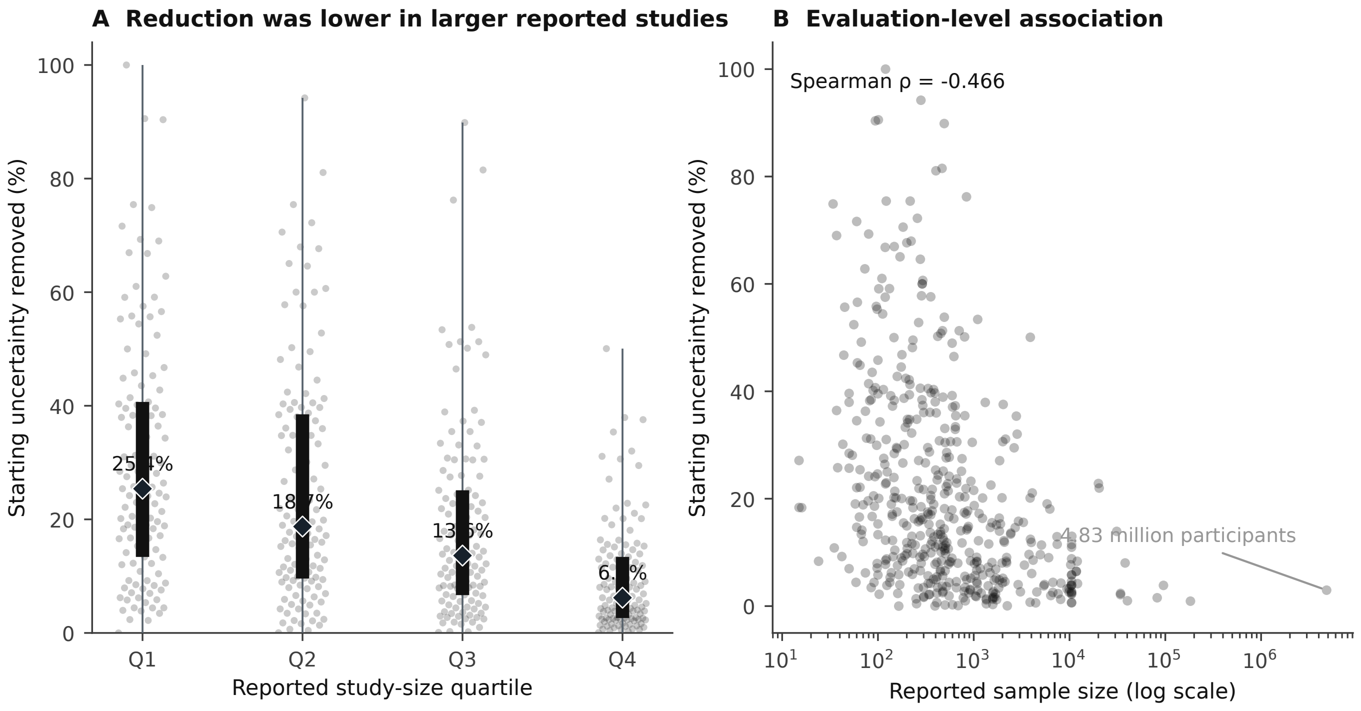

### figureS1_sample_size_information_yield_grayscale.png

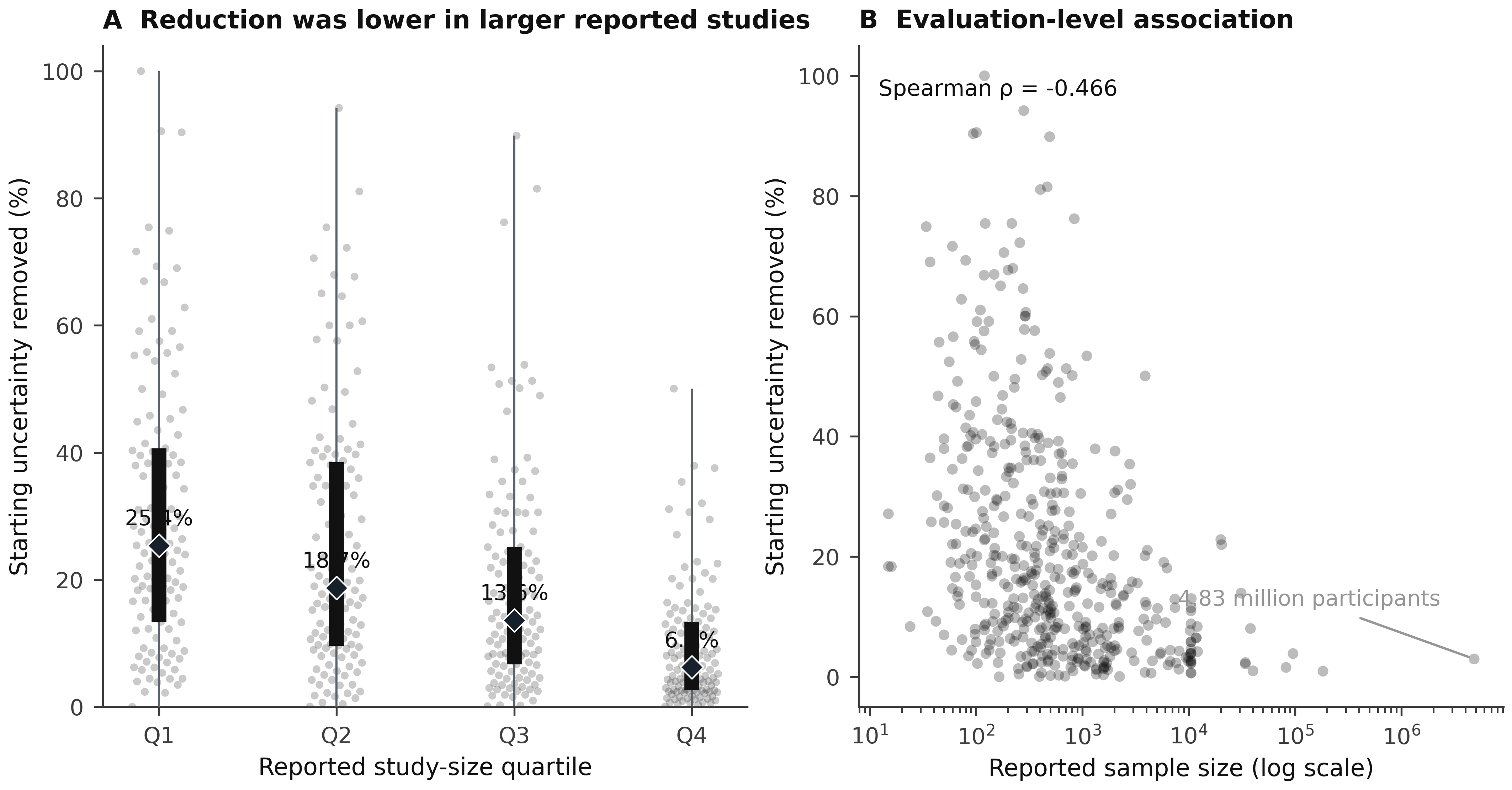

### figureS2_youden_information_relationship.pdf

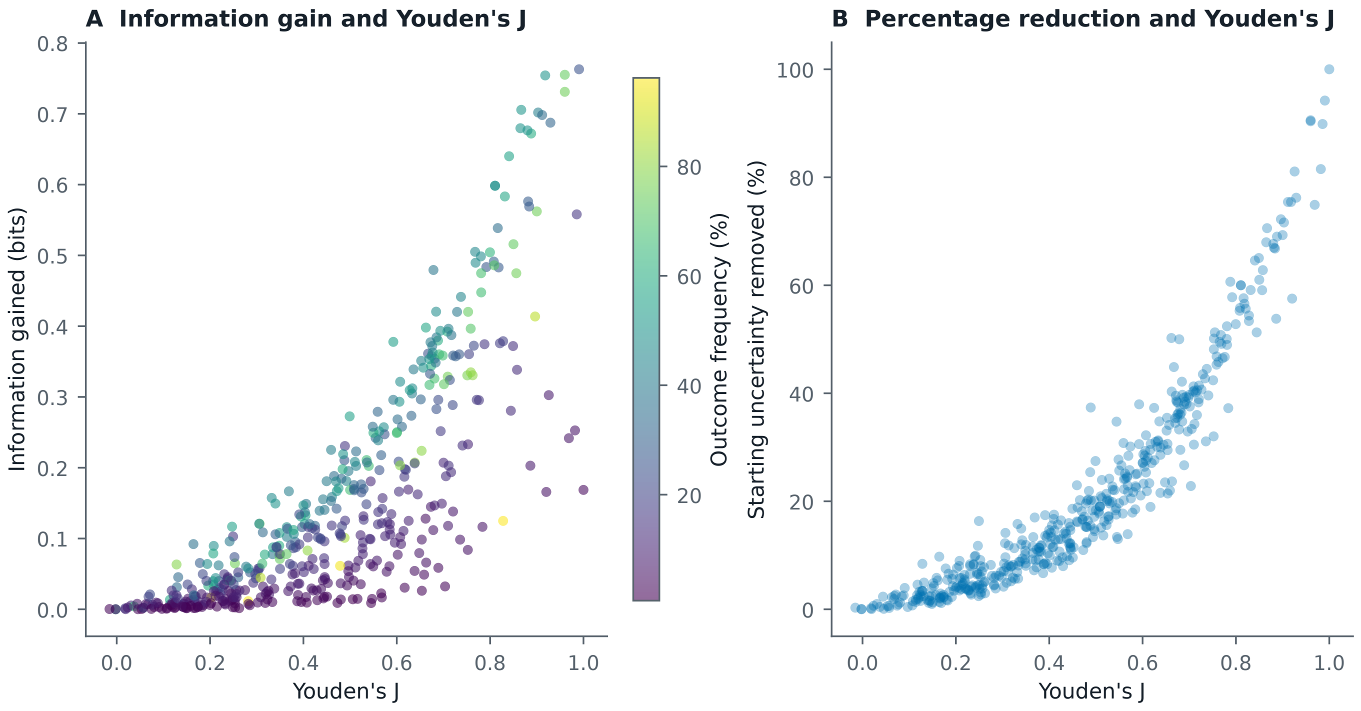

### figureS2_youden_information_relationship.png

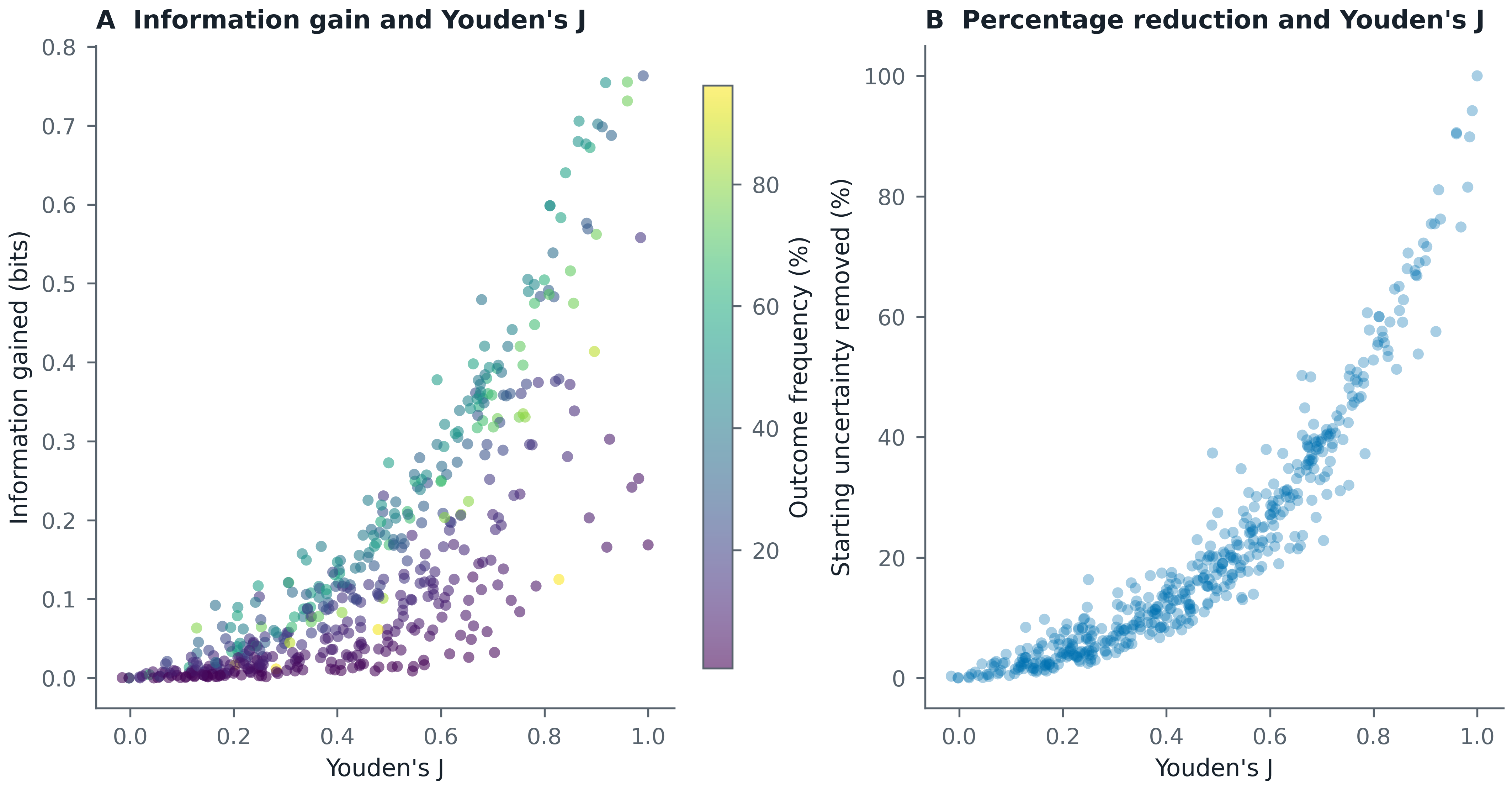

### figureS2_youden_information_relationship_grayscale.pdf

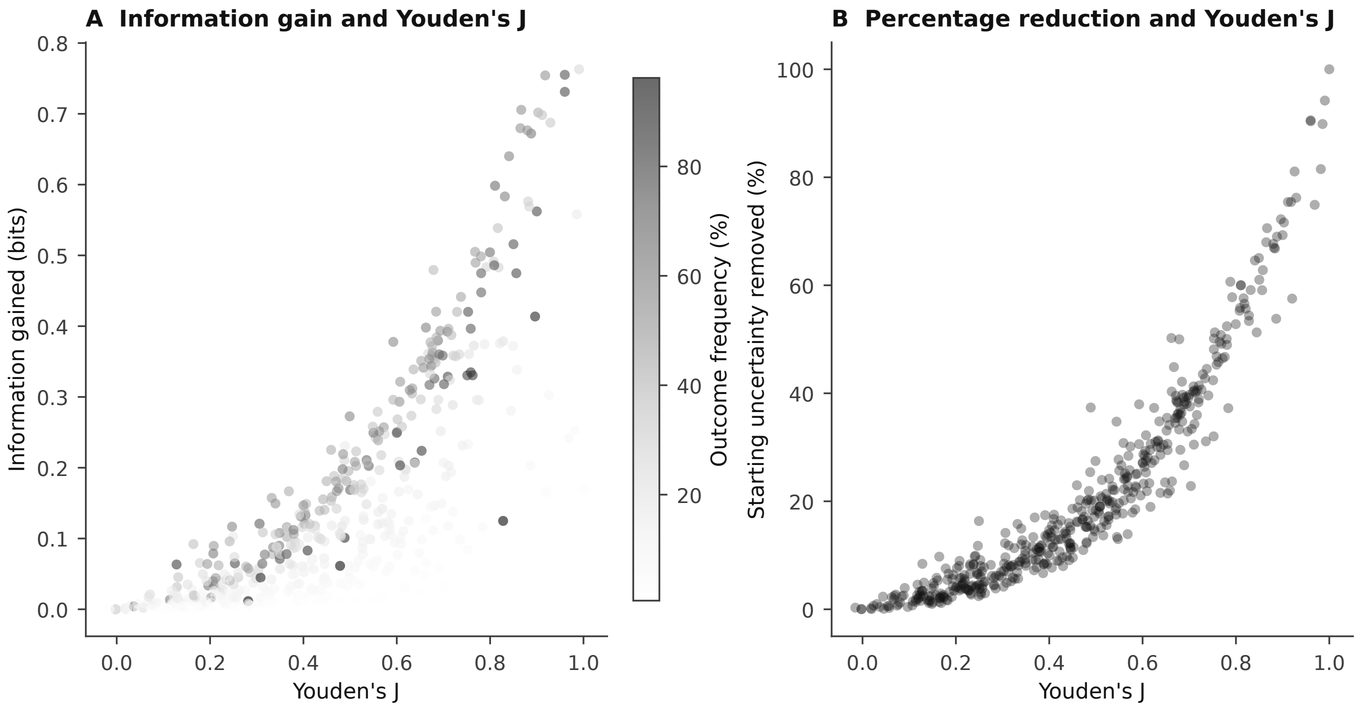

### figureS2_youden_information_relationship_grayscale.png

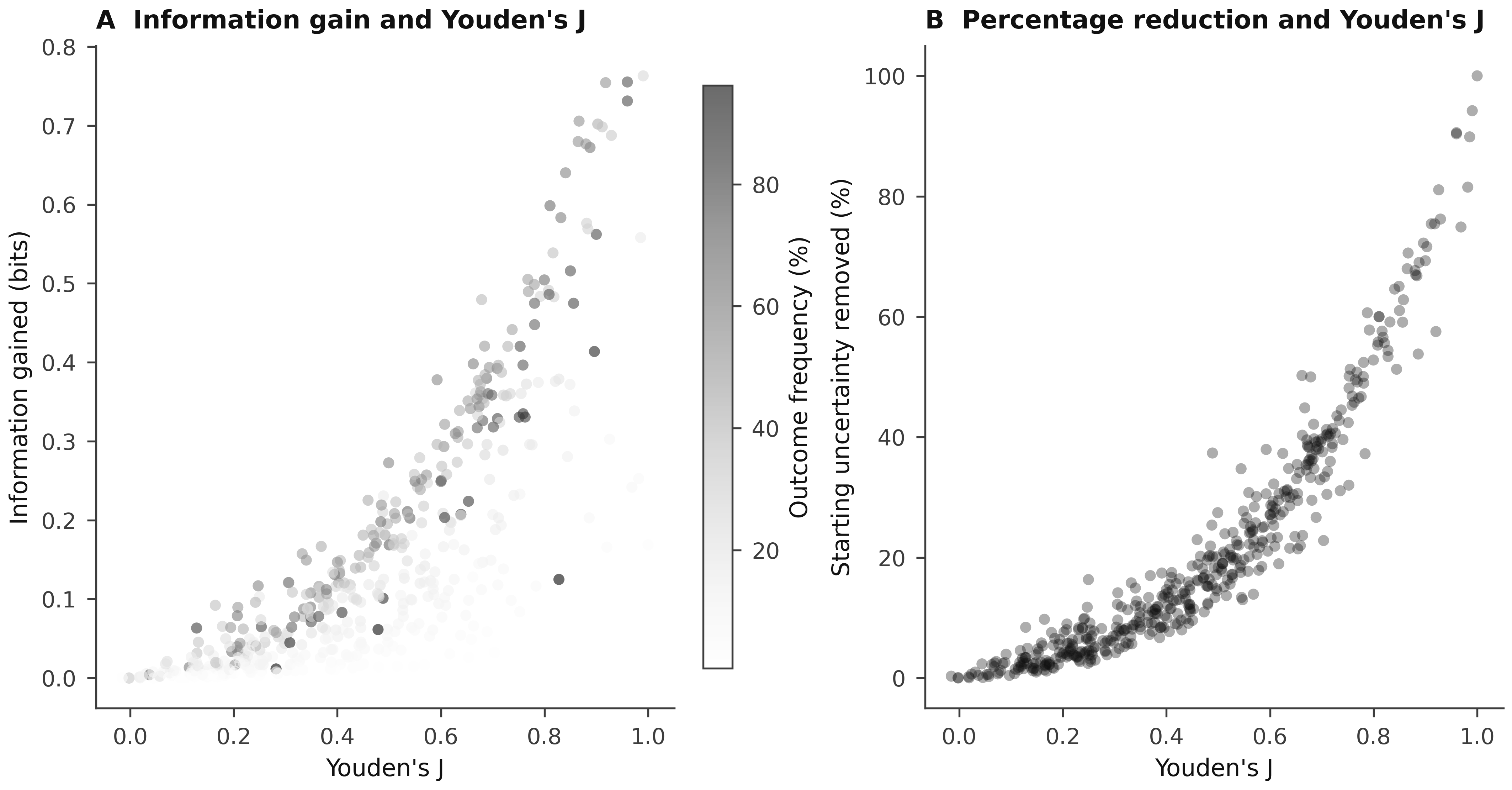

### figureS3_prevalence_standardization.pdf

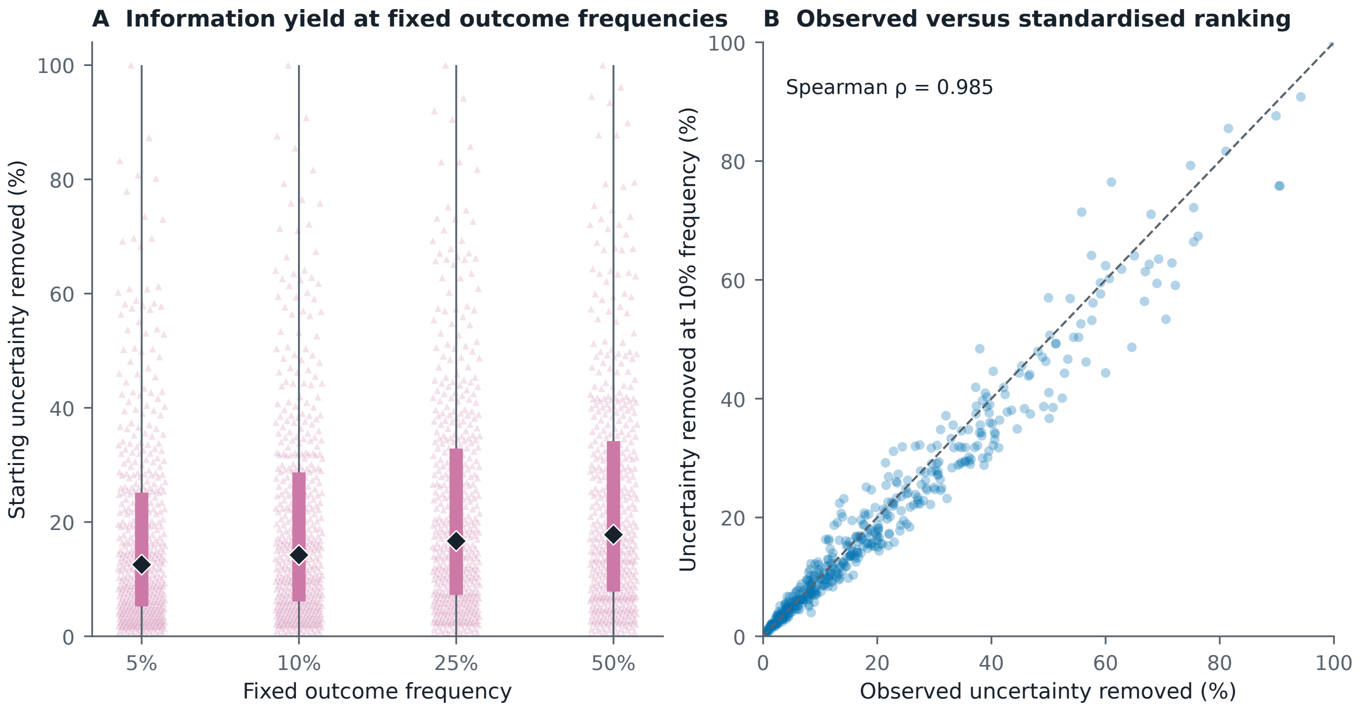

### figureS3_prevalence_standardization.png

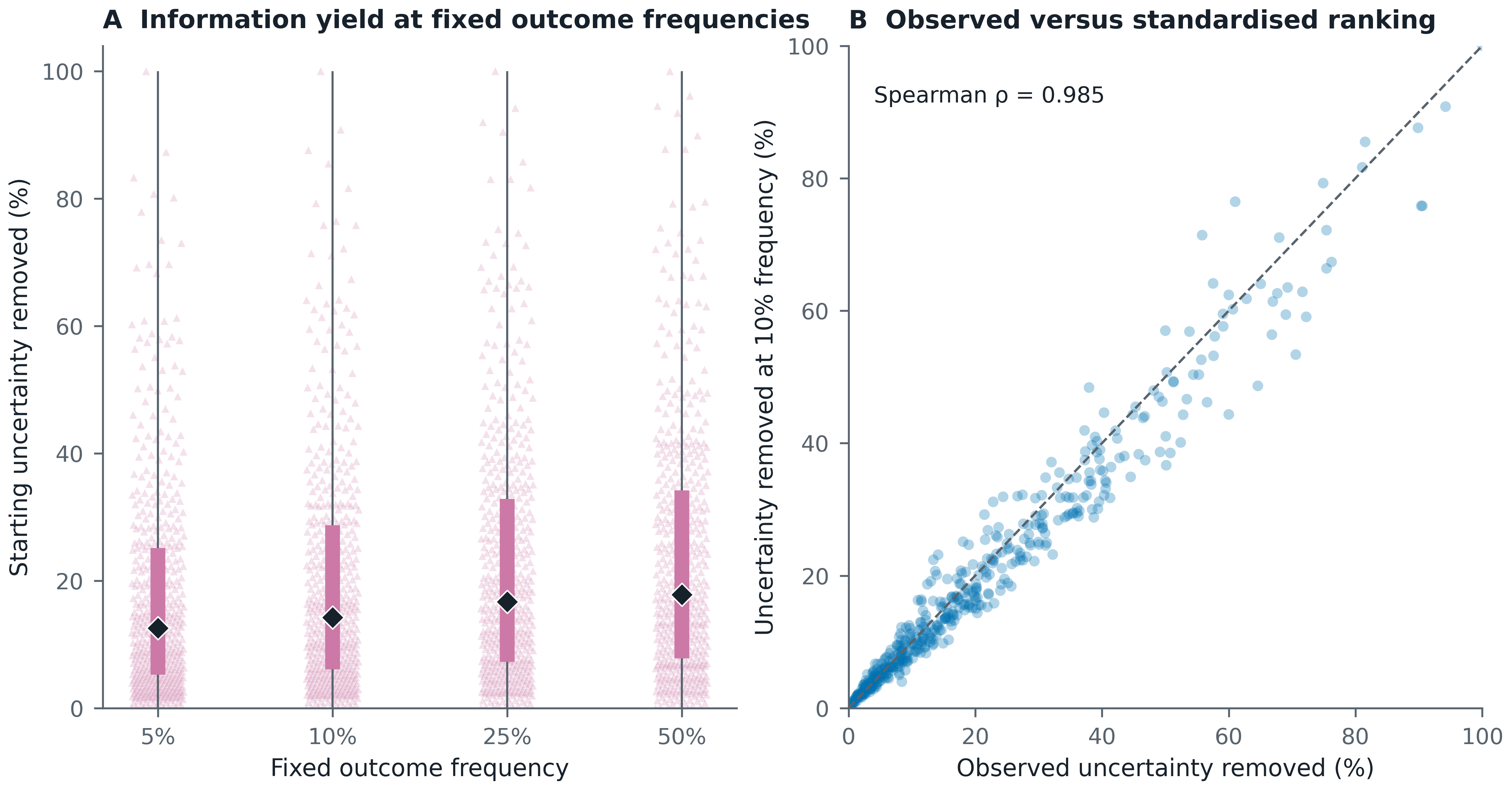

### figureS3_prevalence_standardization_grayscale.pdf

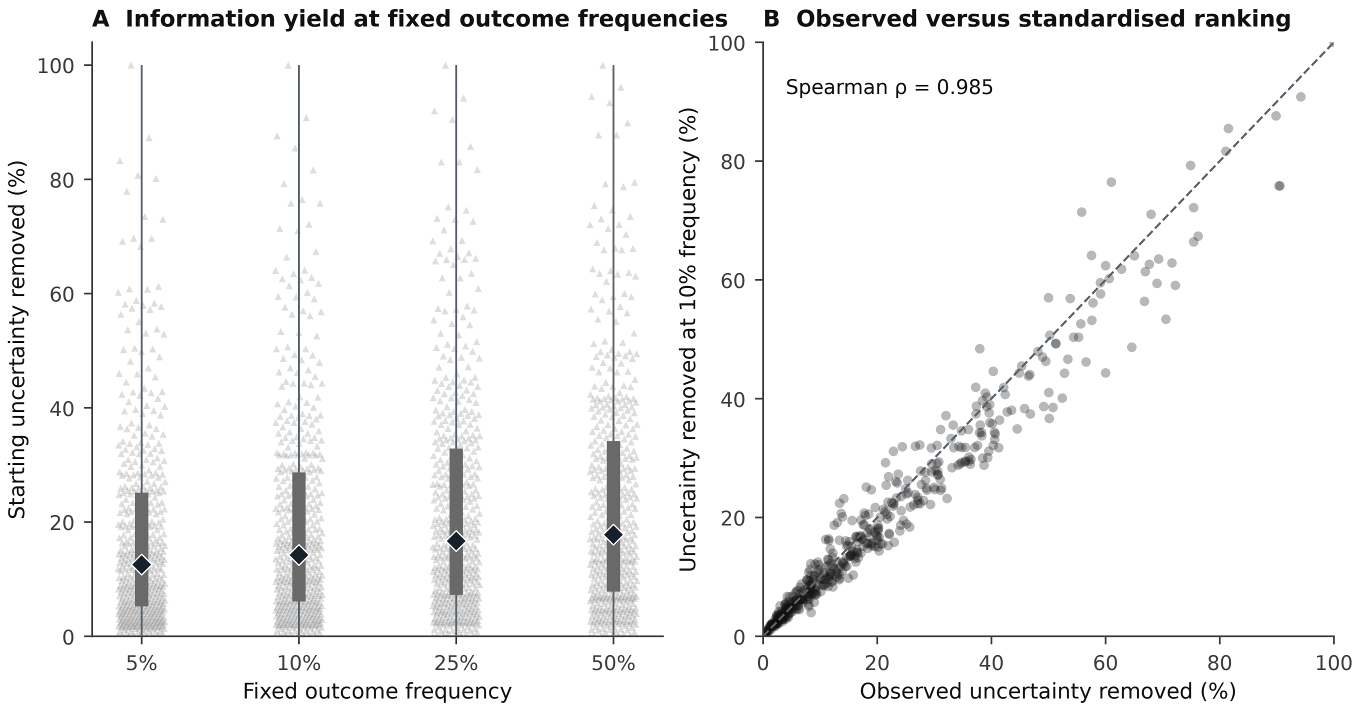

### figureS3_prevalence_standardization_grayscale.png

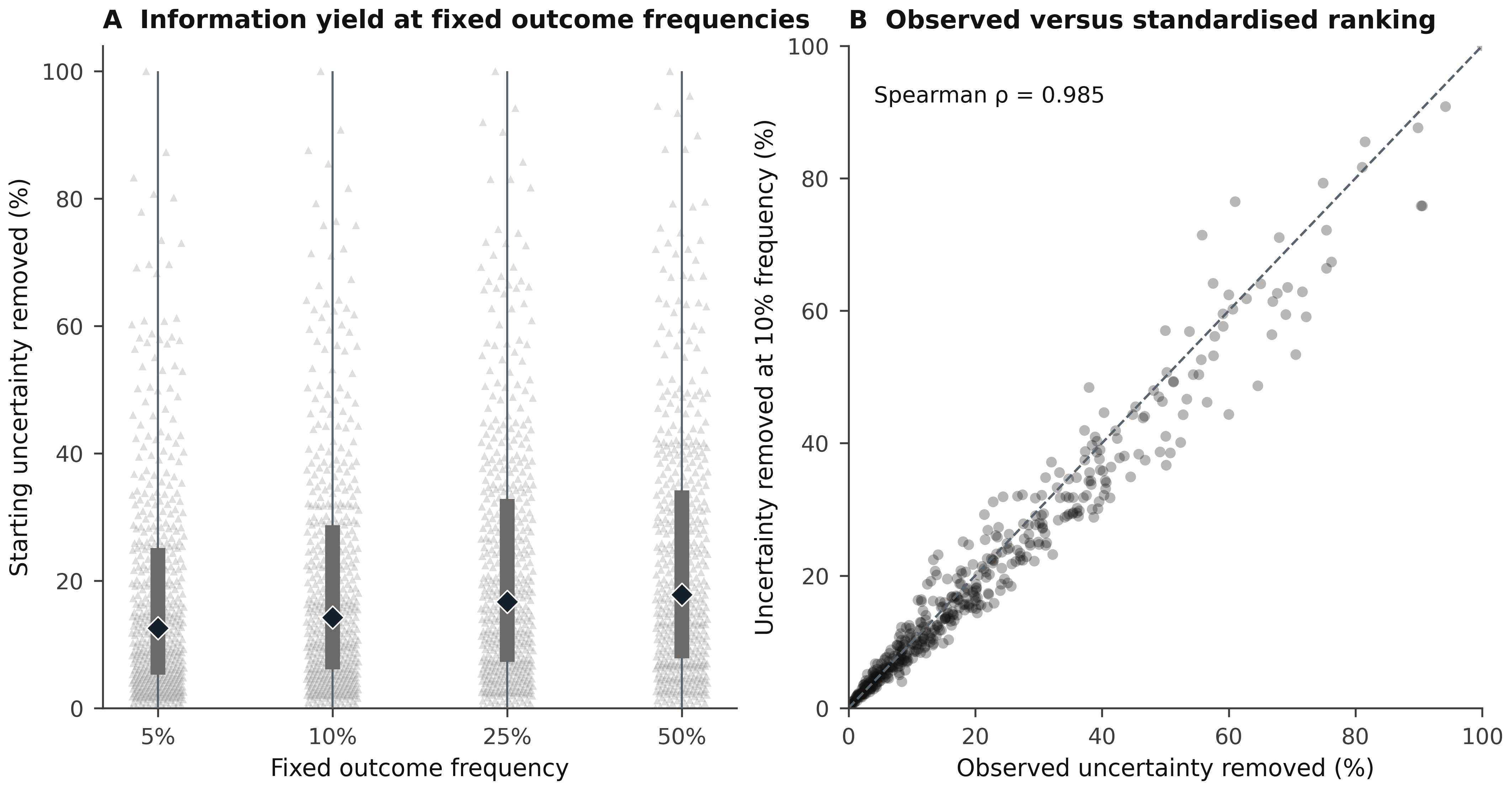

### figureS4_catalog_representation_by_purpose.pdf

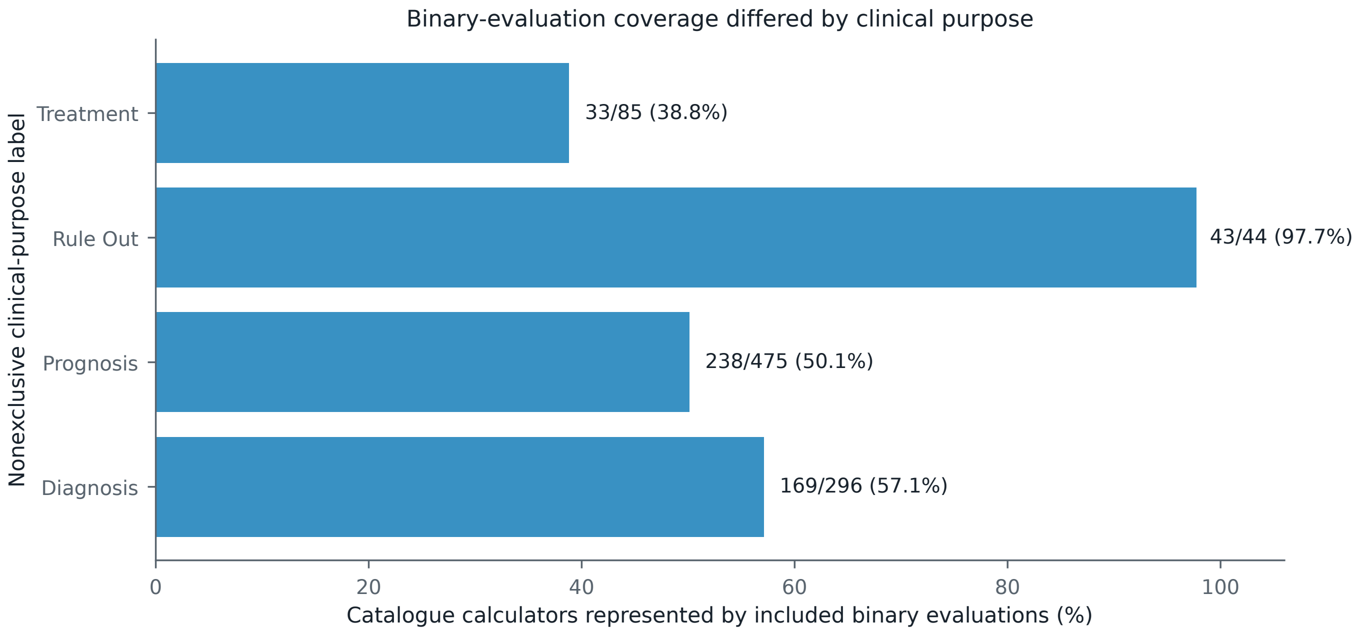

### figureS4_catalog_representation_by_purpose.png

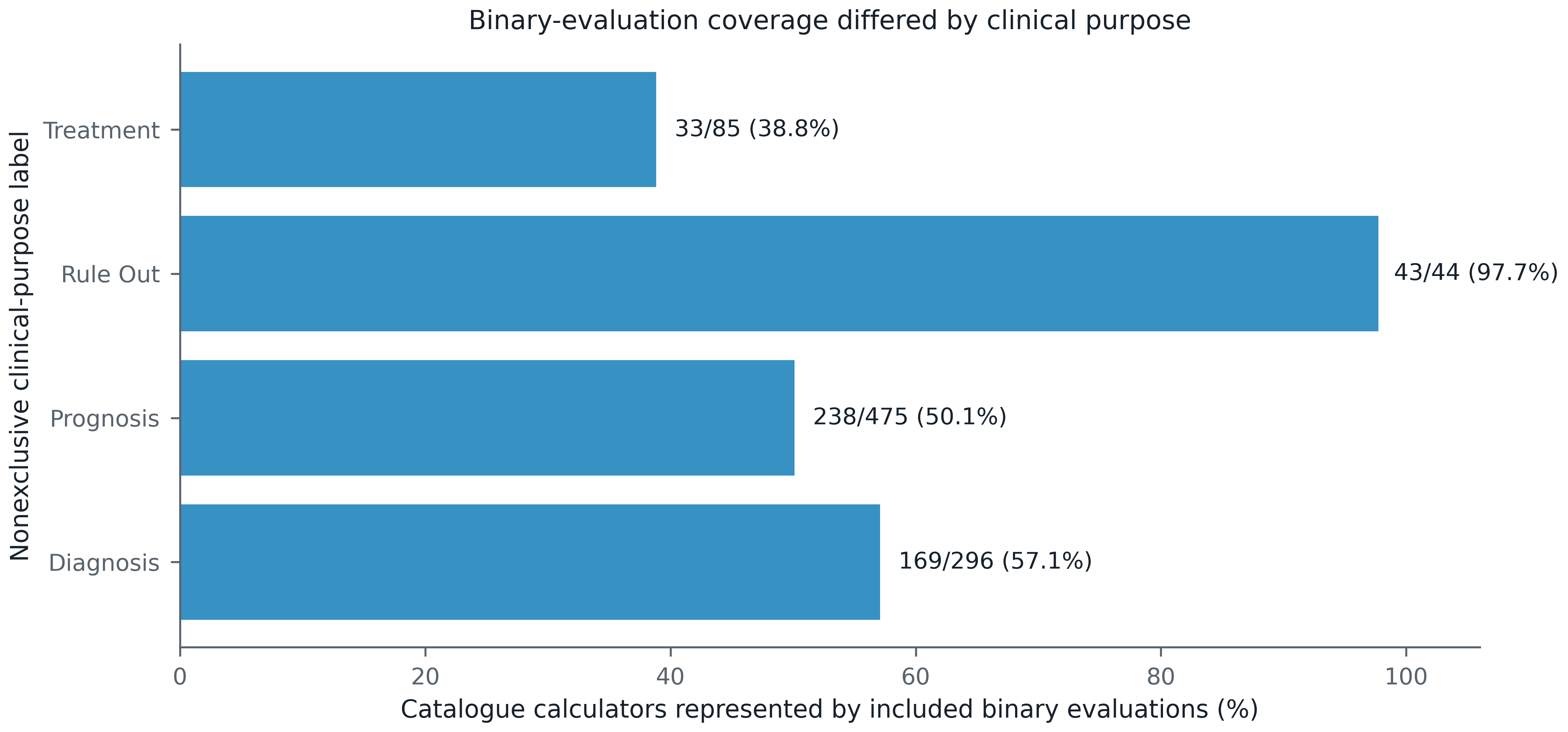

### figureS4_catalog_representation_by_purpose_grayscale.pdf

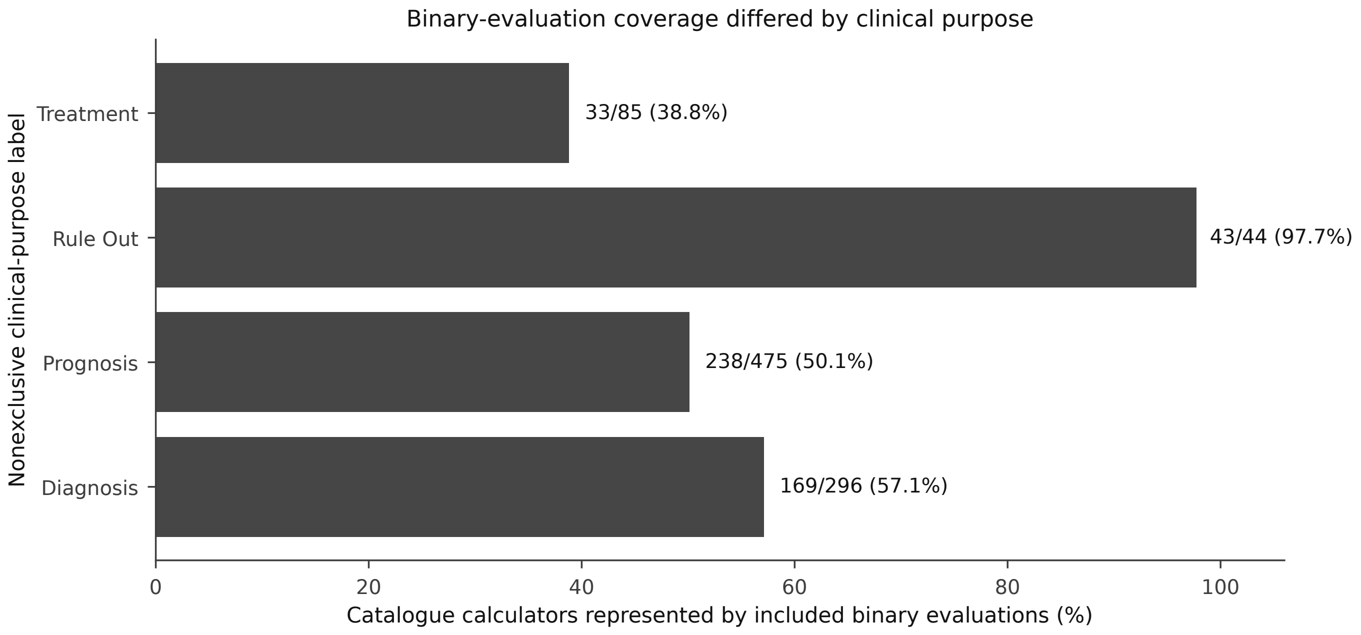

### figureS4_catalog_representation_by_purpose_grayscale.png

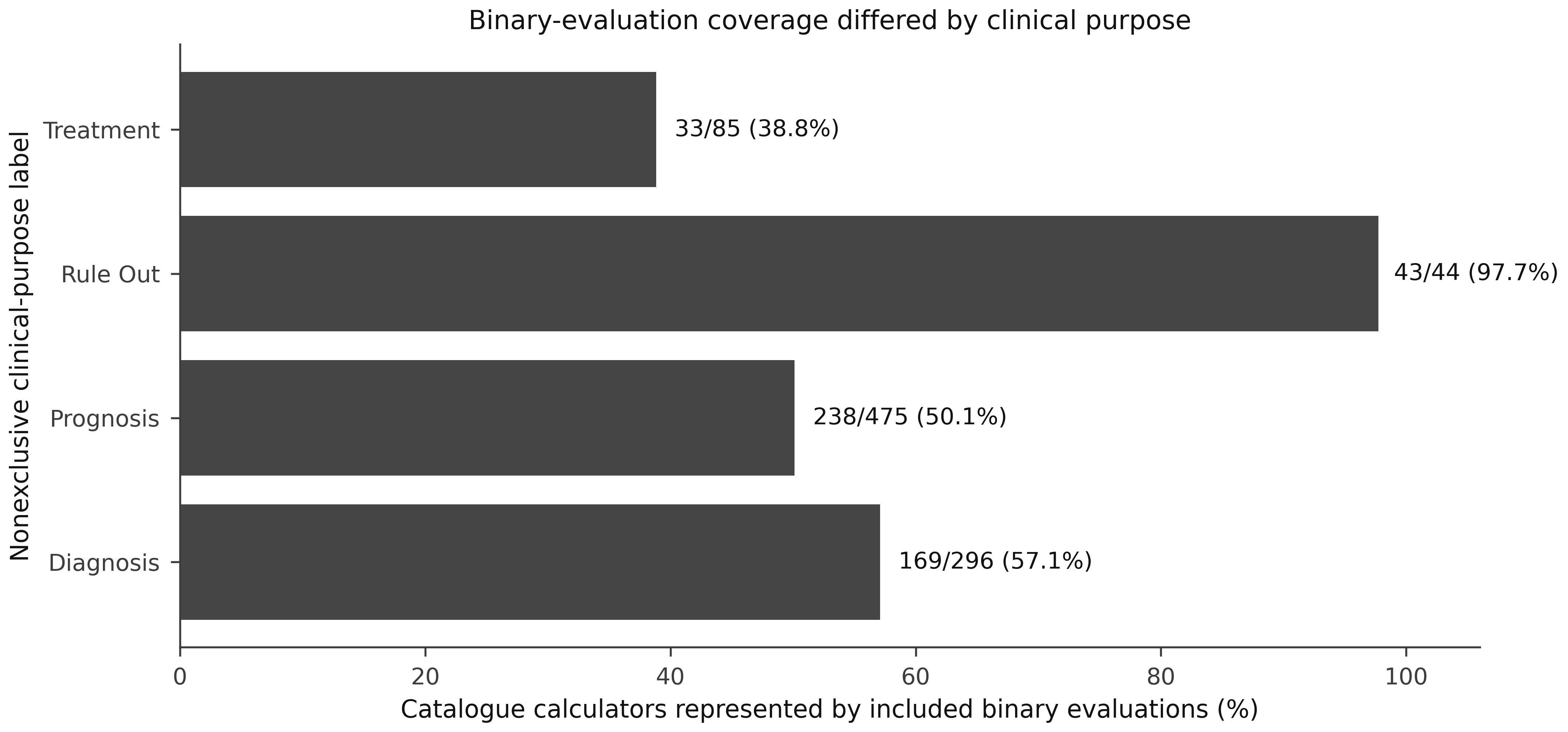

### figureS5_specialty_information_yield.pdf

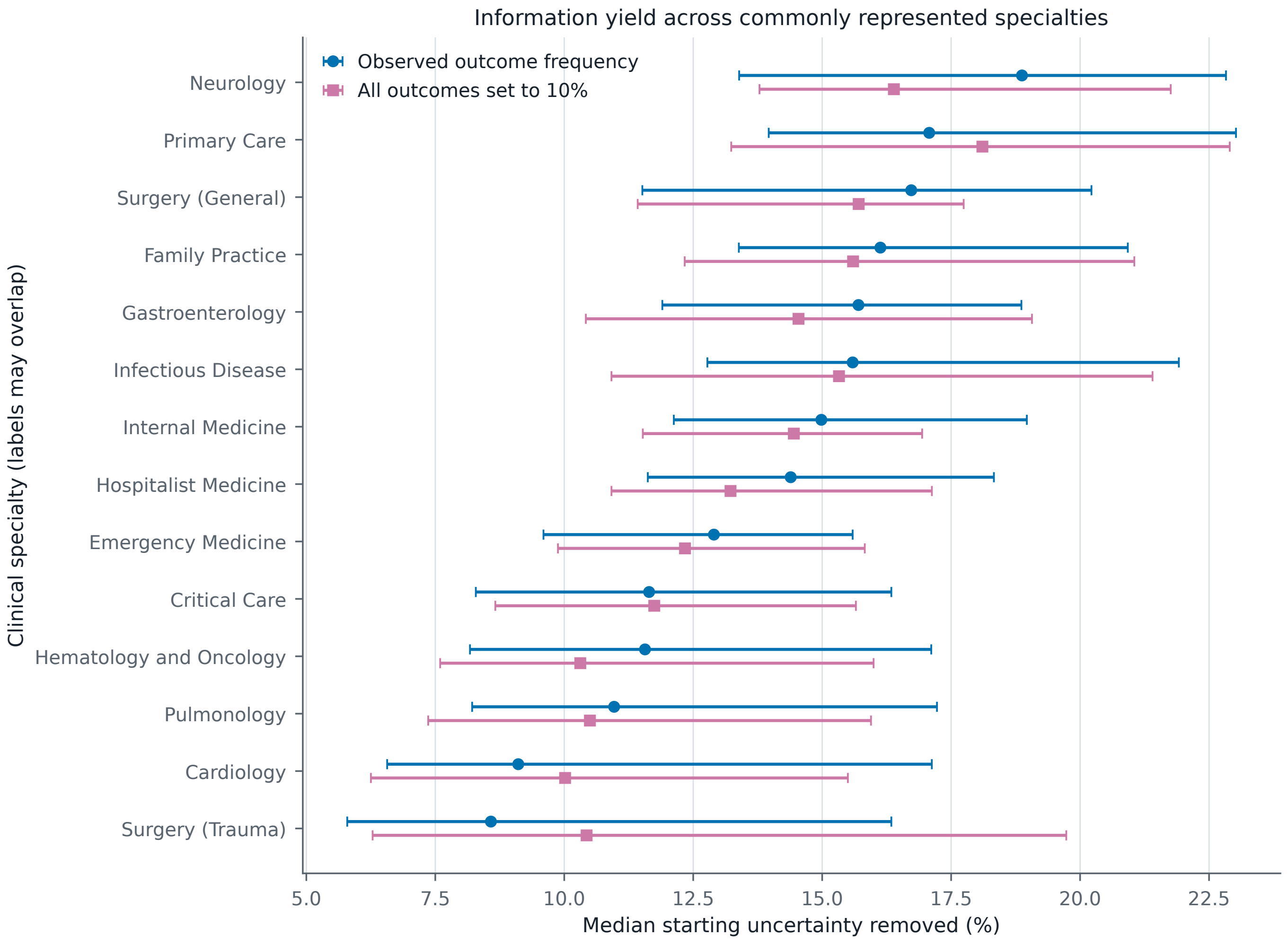

### figureS5_specialty_information_yield.png

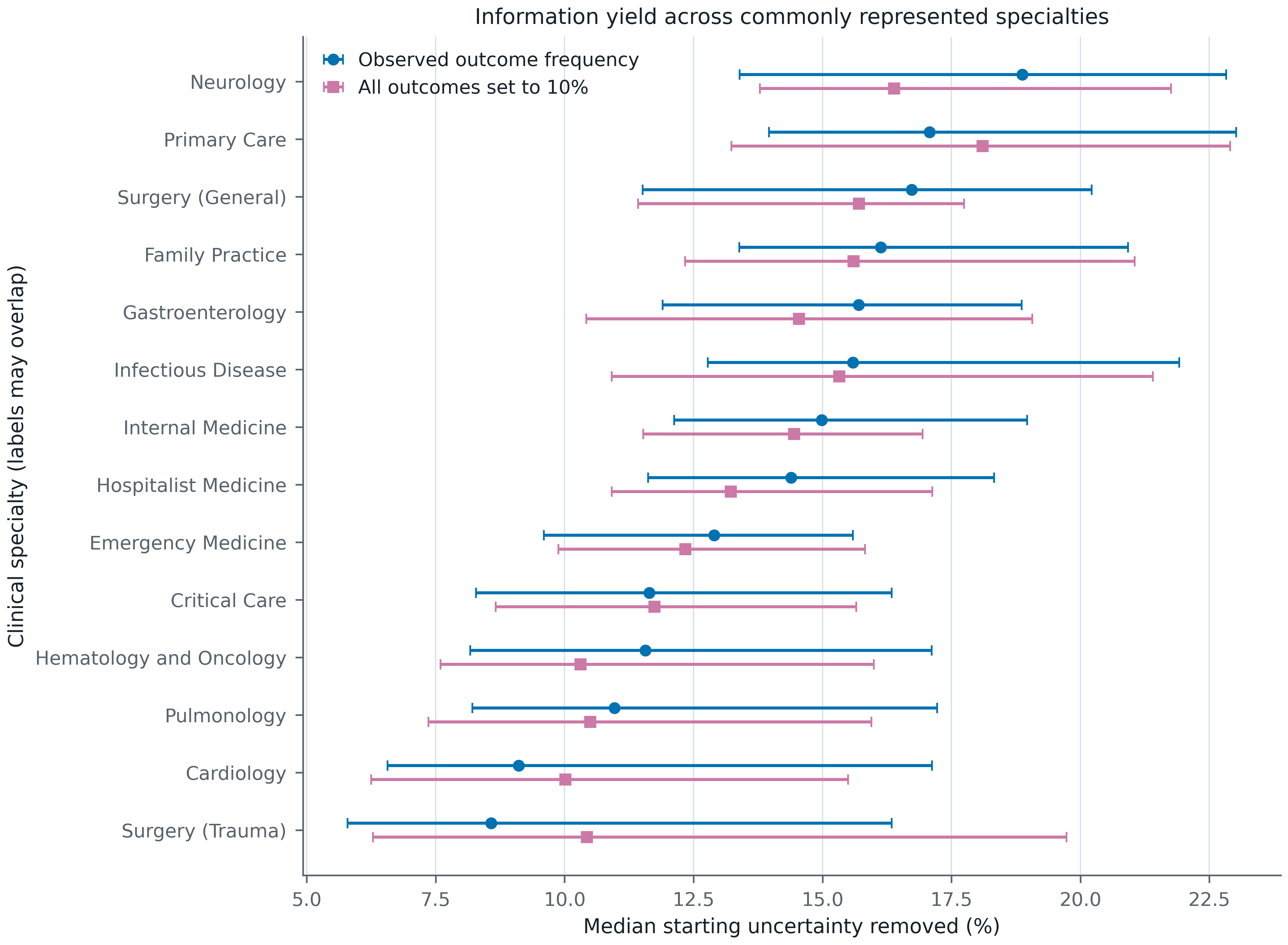

### figureS5_specialty_information_yield_grayscale.pdf

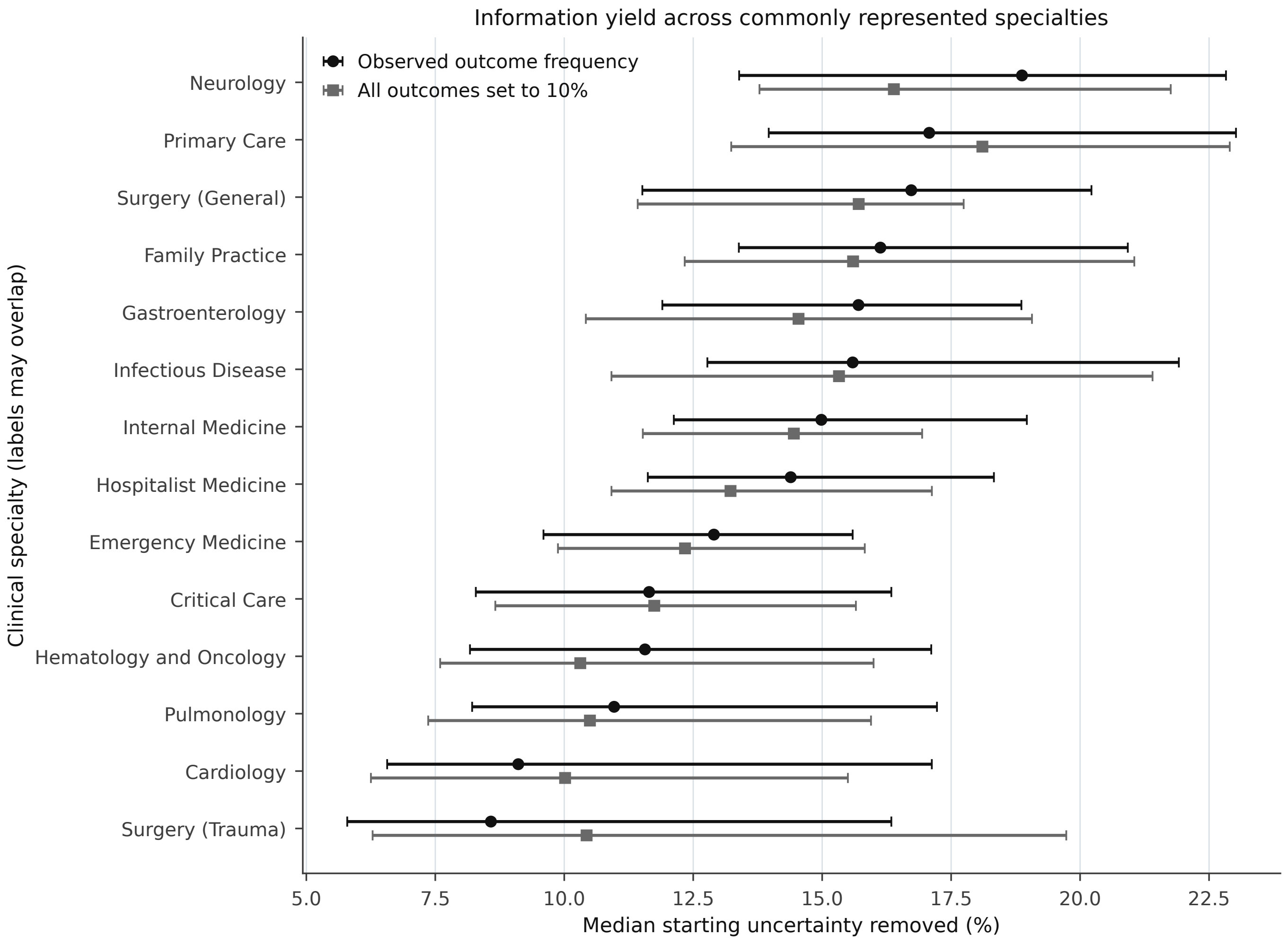

### figureS5_specialty_information_yield_grayscale.png

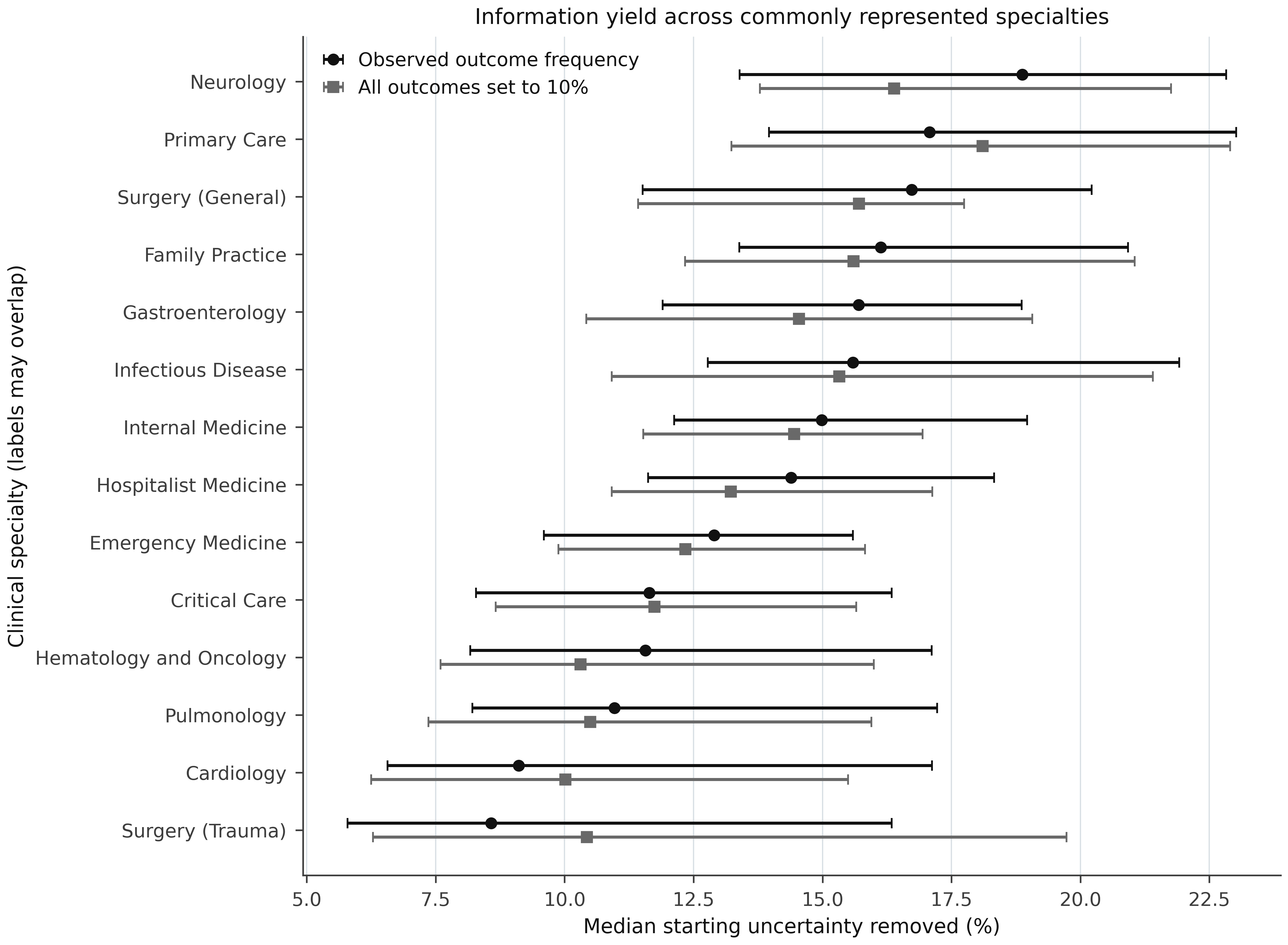

### graphical_abstract.png

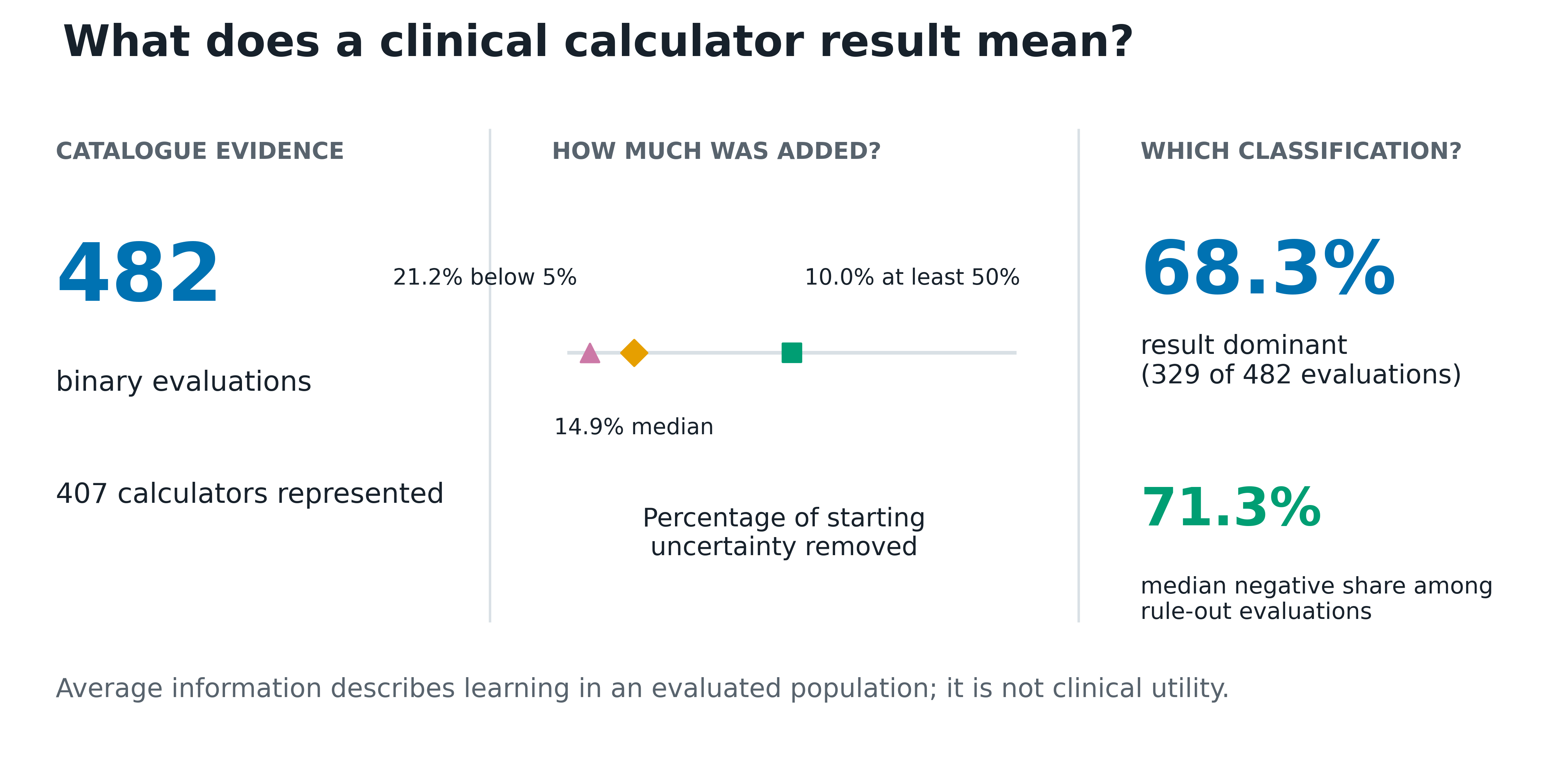

### graphical_abstract_grayscale.png

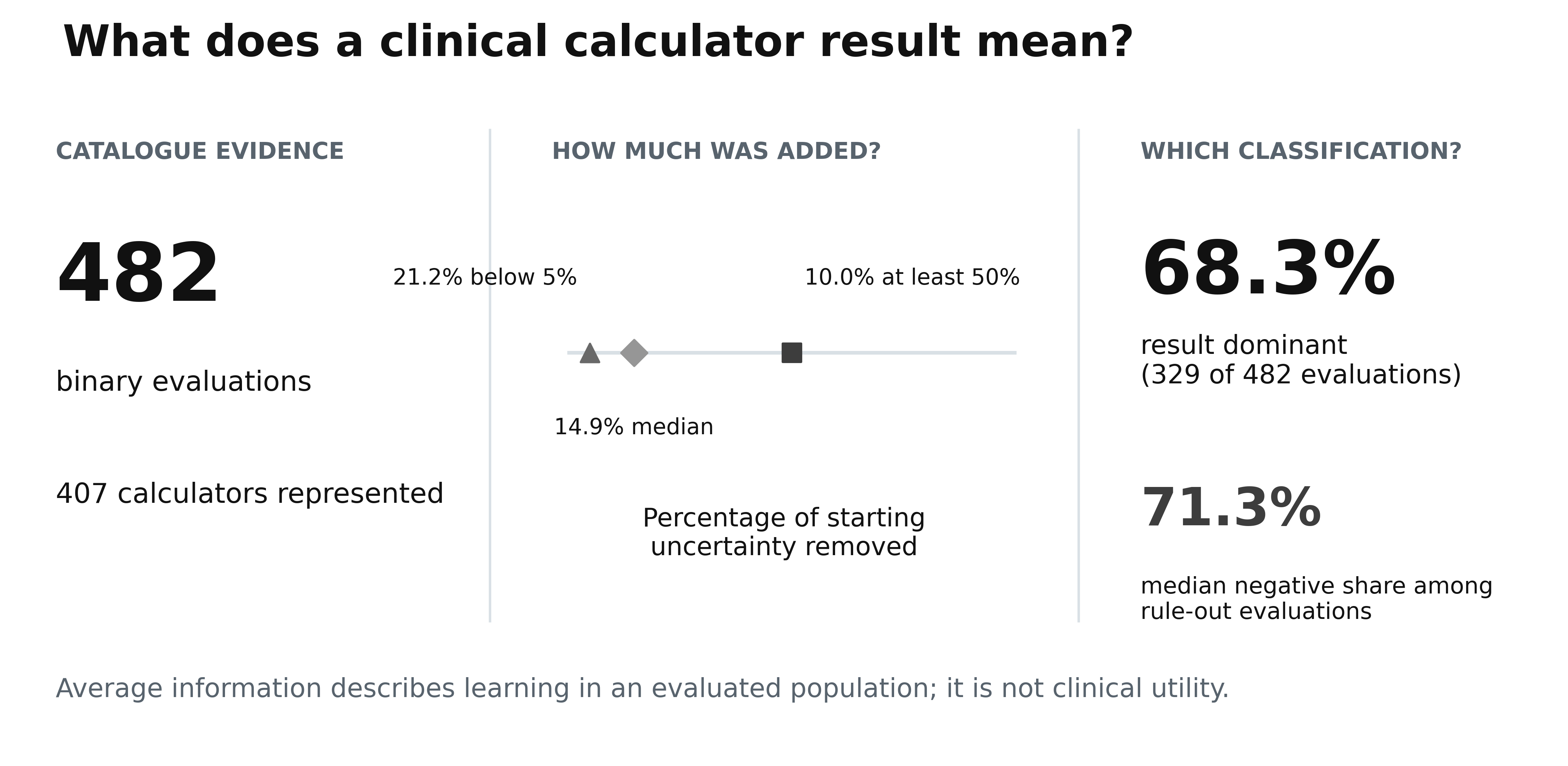
